# A framework to model global, regional, and national estimates of intimate partner violence

**DOI:** 10.1101/2020.11.19.20235101

**Authors:** Mathieu Maheu-Giroux, Lynnmarie Sardinha, Heidi Stöckl, Sarah Meyer, Arnaud Godin, Monica Alexander, Claudia García-Moreno

## Abstract

**Background:** Accurate and reliable estimates of violence against women statistics form the backbone of monitoring efforts to eliminate these human right violations and public health concerns. Estimating the prevalence of intimate partner violence (IPV) is challenging due to variations in case definition and recall period, surveyed populations, partner definition, level of age disaggregation, and survey representativeness, among others. In this paper, we aim to develop a sound and flexible statistical modeling framework for global, regional, and national IPV statistics.

**Methods:** We modeled IPV within a Bayesian multilevel modeling framework, accounting for heterogeneity of age groups using age-standardization, and age patterns and time trends using splines functions. Survey comparability is achieved using adjustment factors which are estimated using exact matching and their uncertainty accounted for. Both in-sample and out-of-sample comparisons are used for model validation, including posterior predictive checks. Post-processing of models’ outputs is performed to aggregate estimates at different geographic levels and age groups.

**Results:** A total of 307 unique studies conducted between 2000-2018, from 154 countries/territories/areas, and totaling nearly 1.8 million unique women responses informed lifetime IPV. Past year IPV had similar number of studies (n=332), countries represented (n=159), and individual responses (n=1.8 million). Roughly half of IPV observations required some adjustments. Posterior predictive checks suggest good model fit to data and out-of-sample comparisons provided reassuring results with small median prediction errors and appropriate coverage of predictions’ intervals.

**Conclusions:** The proposed modeling framework can pool both national and sub-national surveys, account for heterogeneous age groups and age trends, accommodate different surveyed population, adjust for differences in survey instruments, and efficiently propagate uncertainty to model outputs. By describing this model to reproducible levels of details, the accurate interpretation and responsible use of estimates for global monitoring of violence against women elimination efforts are supported, as part of the Sustainable Development Goals.

## Background

Violence against women (VAW) is a human rights violation, and a global health and development concern. To address this issue, countries agreed in 2015 to eliminate all forms of VAW as part of the *Sustainable Development Goals* (SDG). Accurate and reliable VAW statistics form the backbone of monitoring efforts, can help guide resources allocation, and, ultimately, enable the deployment of adequate and sustainable intersectoral responses [1-5]. For VAW elimination to be successful, indicators tracking VAW prevalence must be collected, analyzed, and reported.

Violence against women encompasses physical, sexual, and psychological violence perpetrated by an intimate partner, termed as intimate partner violence (IPV). It also includes sexual violence perpetrated by someone other than an intimate partner (e.g., a friend, family member, neighbor, stranger), termed non-partner sexual violence. Other types of –often overlapping– VAW include child sexual abuse, trafficking of women and girls, female genital mutilation, forced or early marriage, and killings in the name of honor [5]. Previous analyses pointed out that VAW has serious short- and long-term impacts on affected individuals, families, and wider societies and that further investments in research and data collection are required to better understand and address VAW epidemic trends [5-7].

Estimating prevalence of IPV is challenging for several reasons [8]. First, variations in case definition (definitions based on severity of acts) and recall periods (lifetime versus past year) are common. Second, lack of disaggregation between different forms of violence (physical, sexual, psychological) can pose comparability issues. Third, differences in surveyed population (all women, ever partnered, or currently partnered) and whether the perpetrator of violence is the current or most recent partner (versus any previous partners) further compound comparability of estimates. Fourth, reported survey estimates are often not age-disaggregated and, when available, heterogeneous age-group definitions are often encountered, with few observations for women aged 50 years and above. In addition, VAW data are sparse geographically (some countries do not have any estimate) and temporally (most countries with data have only one or two estimates). Considering these issues, comparing and longitudinally tracking IPV statistics requires overcoming several methodological hurdles.

In addition to these issues specific to IPV data, other more general issues need to be considered. First, VAW statistics, like other health indicators, are noisy [9, 10]. This means that the degree of observed heterogeneity can be large; larger than what would be expected from random sampling alone. This heterogeneity can be explained by differences in survey sampling schemes, geographical coverage, survey instruments and methods, and implementation issues, among others. Taken together, these considerations entail that statistical models are required to adjust, compare, and monitor VAW statistics within and across countries.

The objective of this article is to present a flexible statistical modeling framework for monitoring global, regional, and national IPV statistics which can inform the development of effective policies and programs to address VAW and that are in line with SDG monitoring. Specifically, we focus here on SDG indicator 5.2.1: the proportion of ever-partnered women and girls aged 15 years and older subjected to physical, sexual or psychological violence by a current or former intimate partner in the previous 12 months, by form of violence and by age. We first present a brief overview of the global VAW database, provide details on the chosen modeling framework, including adjustments and age modeling, present selected results and model validation, including posterior predictive checks, and discuss potential further steps to improve estimates of IPV statistics.

## Methods

### Global Violence Against Women database

The World Health Organization (WHO)’s global VAW database includes prevalence surveys/studies of physical, sexual and psychological IPV, sexual violence by any perpetrator, and non-partner sexual violence (NPSV). This database builds on the earlier database and systematic reviews that WHO curated [6, 7]. Briefly, all population-based studies conducted between 2000 and 2018, representative at either national or sub-national level, were eligible for inclusion. A few surveys that did not use questions referring to specific acts to measure violence were excluded as these are known to underestimate prevalence. For each eligible study, age-specific prevalence estimates and their denominator, preferably by 5-year age groups, were extracted for the different types of IPV (namely, physical IPV, sexual IPV, psychological IPV and physical and/or sexual IPV) and sexual violence (namely, sexual violence by any perpetrator, and non-partner sexual violence). If only prevalence estimates from a broad age group were available (e.g., 15-49 years old), it was extracted instead. Further, if reported, design-adjusted standard errors and lower and upper limit of the confidence intervals were recorded. For each observation, the following characteristics were extracted: country, author of publication/report, publication year, start and end years of data collection, the type of VAW, the surveyed population (all women, currently-partnered, ever partnered women), age-group for the estimate, the recall period for prevalence (lifetime, past year, past two years), and whether the study is nationally representative and if not, whether it was conducted in an urban, rural, or mixed urban/rural region. IPV estimates were further characterized according to whether the perpetrator included only the spouse (versus all types of intimate partners) and whether the experience of violence referred to only the current or most recent husband/partner (versus any husband/intimate partner).

### Pre-processing

A conceptual overview of methods used for data analysis is provided in Figure 1. This overview describes data inputs, data pre-processing, data analyses, and post-processing to obtain national, regional, and global estimates of IPV statistics.

**Figure 1.**
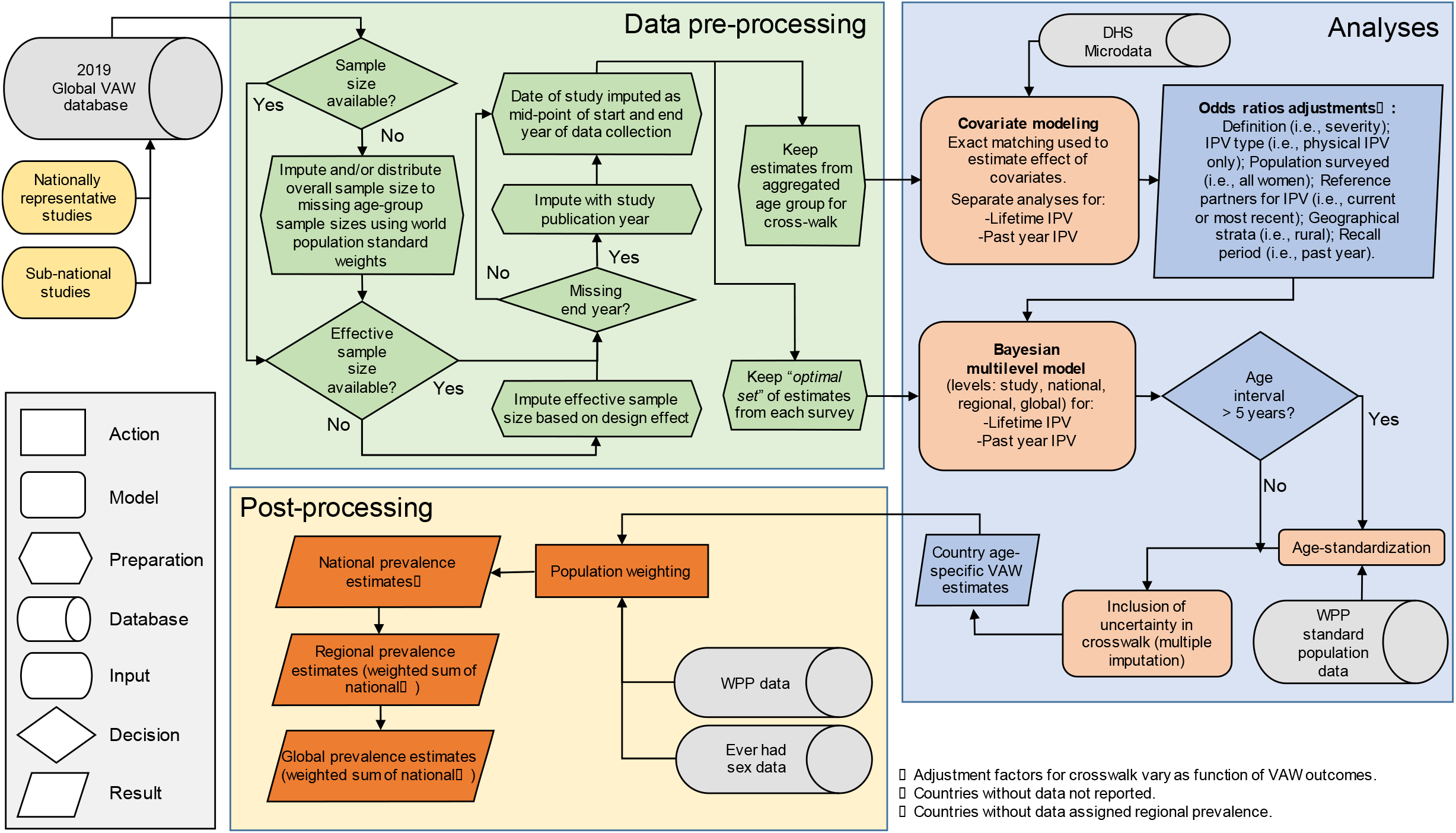
Conceptual overview of data inputs, data pre-processing, data analysis, and post-processing steps required to produce global, regional, and national violence against women statistics. (DHS: Demographic and Health Surveys; IPV: intimate partner violence; VAW: violence against women; WPP: World Population Prospect.)

The first step of the data pre-processing involves the imputation of some missing survey sample sizes. If the overall survey sample size of a specific study was available but the age-specific denominators of the prevalence estimates were missing, we imputed them by distributing this overall sample size proportionally to the age-specific size of the 2010 female population reported in the *United Nations World Population Prospect* (WPP) 2019 [11]. In rare instances where the study’s authors did not report information on the survey sample size, we conservatively assumed that the sample sizes would be of 3,000 and 1,000 for nationally representative and sub-national surveys, respectively. These sample sizes roughly correspond to the lowest tercile of the distribution of all sample sizes in the VAW database.

Population-based surveys included in the VAW database often use complex sampling schemes – for example using stratification and/or clustered sampling– that needs to be accounted for in the analyses. In all cases, the extracted estimates of point prevalence considered survey weighting (if applicable) but stratification and/or clustered sampling could impact the effective sample size of estimates and, hence, the precision of the survey. In cases where design-adjusted standard errors or confidence intervals were available, the effective sample size was derived from these quantities. If only a confidence interval was available, we used Wilson’s formula and applied it to the upper limit of the confidence interval to obtain standard errors [12]. For surveys where the effective sample size could not be numerically derived, we used a design effect of 2.5. This design effect corresponds to the median design effect obtained from standardized analyses of individual-level data from 89 *Demographic and Health Surveys* (DHS). Surveys for which the end date of data collection was not available were imputed using the date of publication as a proxy. Finally, where the upper age of the survey sample was missing or not reported, we assumed an open-ended age category.

The final pre-processing step was to create two datasets (Figure 1). The first one is to be used to calculate adjustment factors that will enable the combination of different types of estimates. In this case, we only kept the prevalence estimates from the broadest age-group (i.e., 15-49 years). The second dataset is the one used to model global, regional, and national estimates of IPV statistics. In this case, only the finest levels of age-stratification were retained. This was done to avoid double-counting women and artificially increasing the precision of the estimates. Similarly, if nationally representative prevalence estimates were available, observations from rural and urban areas of the same study were removed. This process was repeated and, if more than one prevalence estimates remained for each age group, we selected the ones from the “*optimal set*” of observations that used gold-standard methods and survey instruments. Specifically, we applied the following rules for each survey:

□ If a study has estimates from both “*severe physical and/or sexual violence only*” and “*severe and non-severe physical and/or sexual violence*”, we only keep the latter;
□ Keep those from “*physical and/or sexual IPV*” (if unavailable, “*physical IPV only*”; and if not recorded, “*sexual IPV only*”);
□ Retain observations when the surveyed population is composed of “*ever-partnered/married women*” (if not available, “*all women*”; otherwise, we retain “*currently-partnered women*”);
□ Preserve observations reflecting IPV perpetrated by “*any current/previous intimate partners/husbands*” (if unavailable, IPV experienced from the “*current or most recent intimate partner/husband*”).

### Multilevel modeling framework

Multilevel modeling is a useful statistical approach used to pool together observations from different sources. An advantage of the proposed multilevel approach relies on the use of random effects that enable the model to “*borrow strength*” across units. For example, if a country has only one sub-national survey with a small sample size, the accuracy and precision of that prevalence estimate can be improved by empirical observations from similar countries in the same region. Another appealing characteristic of such multilevel models is that the degree of pooling –in other words, how much information is shared between observations– is determined empirically by the data and not arbitrarily by the user [13]. A Bayesian implementation of these models is straightforward and uncertainty is efficiently propagated to model outputs using this approach [10].

The chosen model structure is based on similar meta-regressions of health indicators [6, 7, 9, 10, 14-19] and has five nested levels: 1) individual studies, 2) countries (including territories and areas), 3) regions, 4) super-regions, and 5) the world. Here, regions correspond to the classification used by the *Global Burden of Disease* (GBD) study that groups countries in 21 mutually exclusive regions (Figure 2), themselves grouped into seven “super-regions” (Figure 3), based on the similarities of their epidemiological profiles. The regression model uses a binomial likelihood where *y*_*it*_ is the survey-adjusted number of women reporting violence for observation *i* at calendar year *t* and *N*_*it*_ is the effective sample size for that observation.

**Figure 2.**
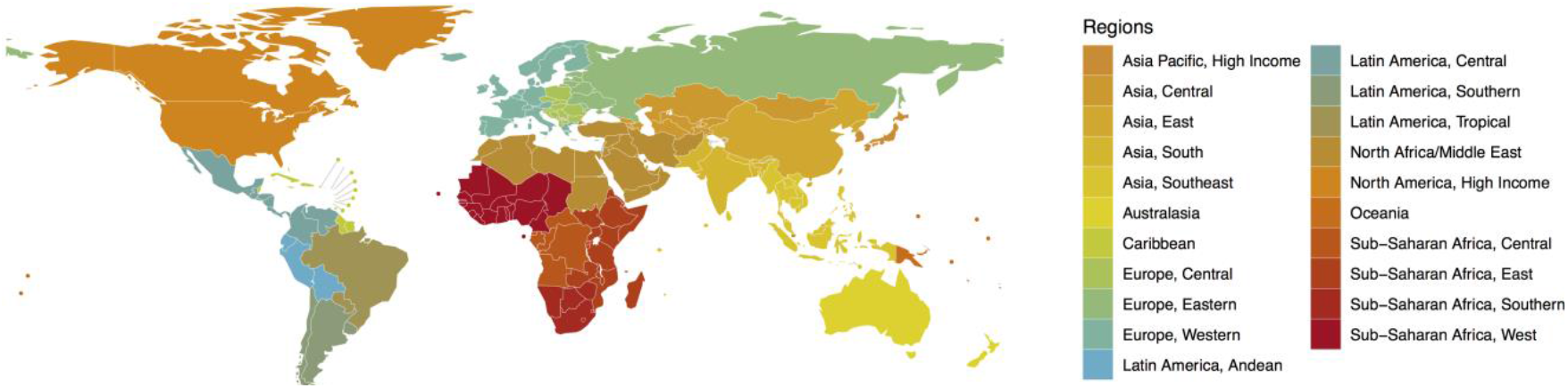
Classification of countries into twenty-one *Global Burden of Disease* “regions”. Disclaimer: The boundaries and names shown and the designations used on this map do not imply the expression of any opinion whatsoever on the part of the World Health Organization concerning the legal status of any country, territory, city or area or of its authorities, or concerning the delimitation of its frontiers or boundaries. Dotted and dashed lines on maps represent approximate border lines for which there may not yet be full agreement.

**Figure 3.**
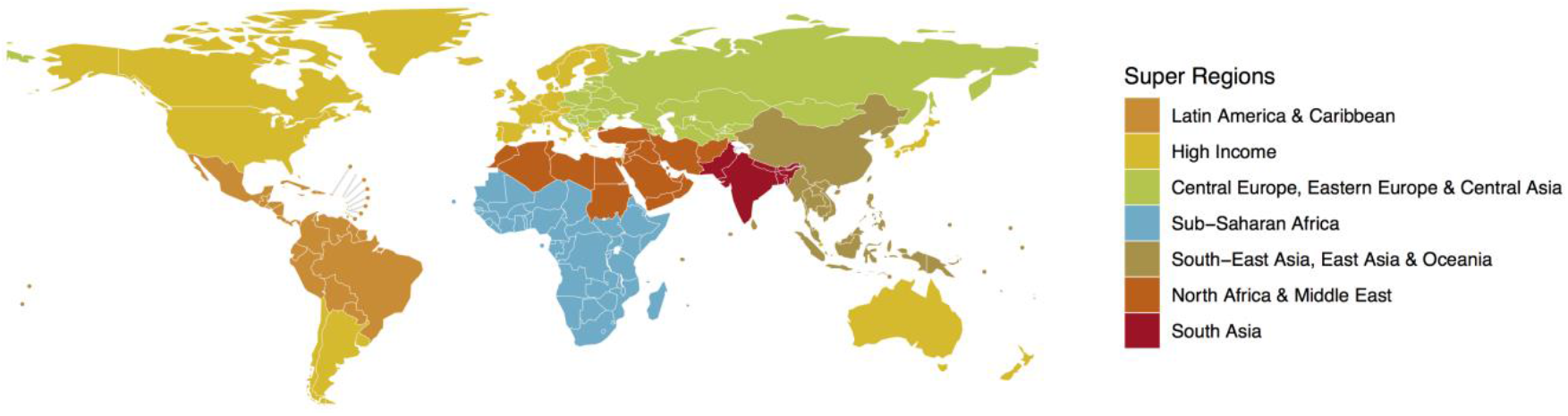
Classification of countries into seven *Global Burden of Disease* “super-regions”. Disclaimer: The boundaries and names shown and the designations used on this map do not imply the expression of any opinion whatsoever on the part of the World Health Organization concerning the legal status of any country, territory, city or area or of its authorities, or concerning the delimitation of its frontiers or boundaries. Dotted and dashed lines on maps represent approximate border lines for which there may not yet be full agreement.

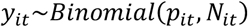

Further, the logit-transformed prevalence estimate *p*_*it*_ is equal to the sum of the study specific intercepts (i.e., the random effects; denoted *α*_*s[i]*_), the country-specific age adjustments (*γ*_*c[i]*_), the country-level time trend (*δ*_*c*_[_*i*_]_,*t*_), and the sum of the log-odds ratios of the adjustment factors (i.e., the crosswalk covariate modeling; *X*_*s[i]*_). In its simplest form, the model takes the following form:

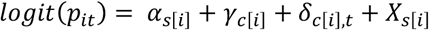

The four terms on the right-hand-side of this equation are detailed in the next sections.

### Random effects to account for study variability

Random effects are useful to account for unobserved heterogeneity and each study is assumed to have its own random intercept. We can further impose a hierarchy on these intercepts. This means that we can assume that each study, conducted within a selected country, should yield a prevalence estimate closer to the average prevalence of that country as opposed to that of other ones. We further posit that the average prevalence in a country should be closer to its regional prevalence than to that of other regions of the world. Nesting these effects within clear geographical units is statistically advantageous because it enables us to borrow strength from other geographical units to improve estimate of prevalence in data sparse settings. To model this hierarchy, we have the following equation for the intercept (*α*_*s[i]*_) of observation *i*:

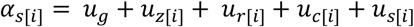

where *u*_*g*_ is the overall global intercept, *u*_*z*_ is the super-region effect, *u*_*r*_ is the regional effect, *u*_*c*_ is the country effect, and *u*_*s*_ is the study effect. It is assumed that these effects are normally distributed on the logit scale. The following non-informative priors were assigned to these parameters:

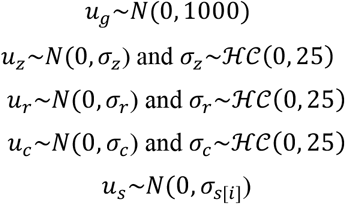

The degree of pooling between the different studies depends on the standard deviation of the random effects. A small standard deviation entails that the degree of pooling of prevalence estimates will be greater than if the standard deviation is large. The standard deviations for the super-region (*σ*_*z*_), region (*σ*_*r*_), and country (*σ*_*c*_) random effects are given weakly informative half-Cauchy (*ℋ𝒞*) priors with a scale parameter of 25, as suggested by Gelman [20]. We also consider that sub-national studies, such as the ones conducted in only one administrative region of a country, are inherently more variable than if they had been nationally representative. As such, they should potentially be given less weight than nationally representative surveys. To do so, we modeled the standard deviation of the study-level random effect depending on its representativeness at the national level [10]. This effectively means that sub-national studies have equal or more variability than those representative at the national level.

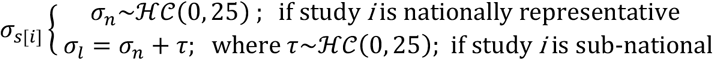

### Age modeling

Previous studies suggested that the relationship between age and IPV is not linear [21, 22]. Splines are a simple and effective way to model non-linear relationships using piecewise polynomials [23]. Based on our understanding of VAW epidemiology, we investigated several natural cubic splines with one knot (at 20, 25, 30, or 35 years) or two knots (at 20 and 35, 20 and 40, 25 and 35, and 25 and 40 years). The splines providing the best fit for each outcome was informed by the Deviance Information Criterion [24] and the Widely Applicable Information Criterion [25, 26]. In all cases, age is centered at 30 years old to improve model convergence. Because the data in the age groups above 65 years old are very sparse, we modified the splines so that prevalence among the ≥65 years age group remains constant. This was achieved by recoding all ages above 65 years to that value before calculating the splines and fitting the model. Our model assumes that each country has its own age pattern (*γ*_*c*_) but that this pattern is more similar across regions, and super-regions. In practice, this means that we have included country-specific coefficients (random slopes) for the natural cubic spline with *K* degrees of freedom, denoted *λ*_*c[i],k*_.

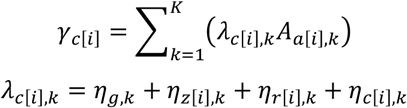

where *η*_*g,k*_ is a vector that contains the coefficients for the global age-prevalence pattern common to all studies, and *η*_*z[i],k*_, *η*_*r[i],k*_, and *η*_*c[i],k*_contains the super-region, region, and country-specific deviations from this overall pattern, respectively. In all cases, the midpoint of the 5-year age distribution is used to obtain the basis of the natural cubic spline (*A*_*a[i],k*_). The model specification is completed using non-informative normal prior distributions. Hyper-parameters for the standard deviations of the random coefficients (*ν*_*k*_) are given weakly informative half-Cauchy (*ℋ𝒞*) prior distributions.

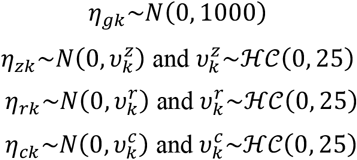

An additional complexity to consider is that of heterogeneous age groups. Some prevalence observations refer to 5-year age groups (at best), others to much wider ones (i.e., 15-49 years old). To enable inclusion of all observations and consider these age-heterogeneous categories, an age-standardizing approach was adopted [9]. The rationale for age-standardization is depicted in Figure 4 where it is showed that the prevalence in a wide age group is a function of both the age-specific prevalence and the underlying age distribution of the sampled population. The sample’s underlying age distribution is estimated from the *UN World Population Prospect* (2019 revision) [11] and we aggregated the 2010 country-level female age distributions for the 21 GBD regions. We used 2010 as this roughly corresponds to the median date of the survey data collection. Age-standardization is applied to all age groups for which the width of the age interval was larger than five years. In such instances, the effect of age on prevalence is modeled as a weighted average of the age-specific prevalence estimates for that country, over the lower (*l*_*i*_) and upper (*h*_*i*_) limits of the age interval to which observation *i* belongs, and where *w*_*a[r]*_ is the weight for the corresponding age group in region *r*:

**Figure 4.**
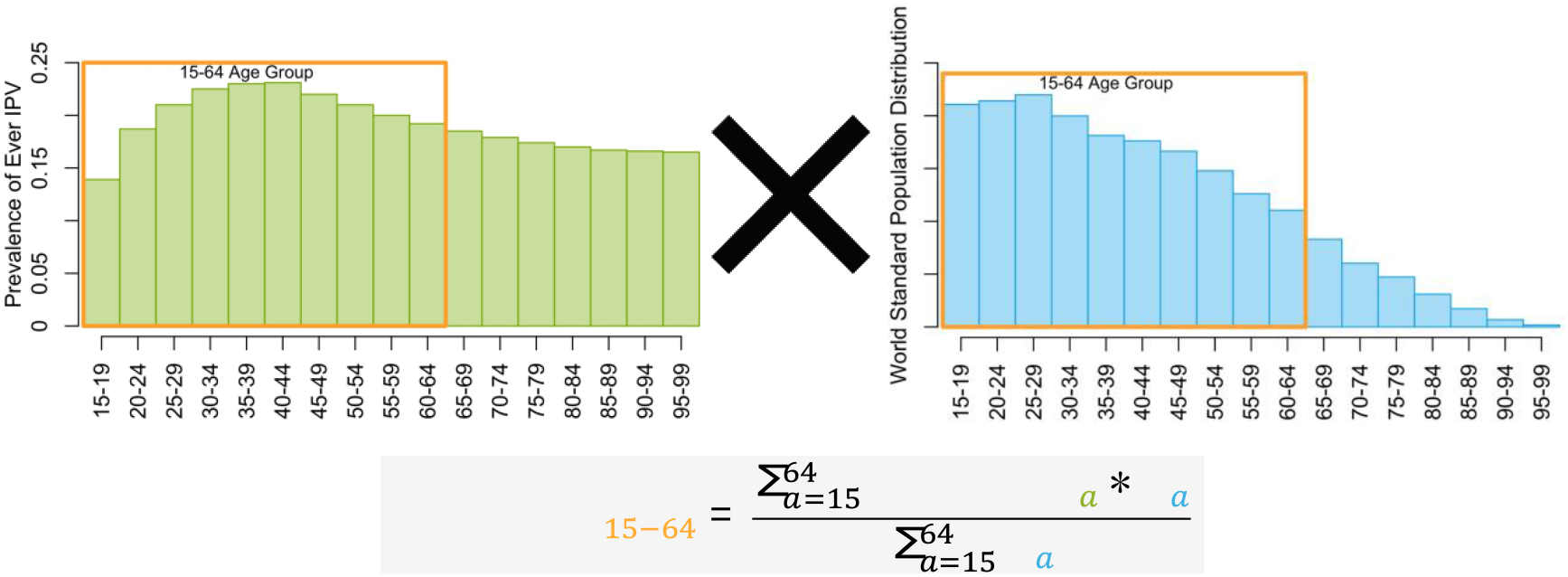
Visual representation of an age-standardizing model when age groups are heterogeneous. The left graph has fictional data for the age pattern for lifetime prevalence of intimate partner violence (IPV). The right graph shows that the underlying population distribution is not uniform. If IPV prevalence is estimated for the 15-64 age group, the resulting prevalence will be a function of the specific age pattern of IPV (left) and underlying age distribution (right); where the *a* subscript indexes the five-year age group. Hence, the equation below the graphs says that the prevalence in the 15-64 age group is a weighted average of the age-specific prevalence estimates where the weights correspond to the relative size of the age groups.

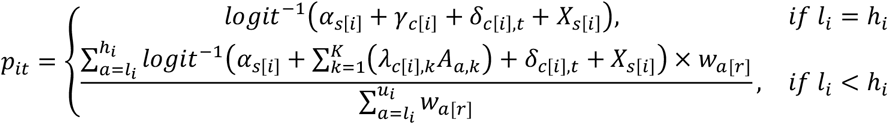

### Time trends

Prevalence of IPV could exhibit secular changes over the near 20-year study period. To allow for potential non-linear changes in prevalence, natural cubic splines with one knot placed at the median year of data collection were used (i.e., 2011). Here again, we modeled the country-specific time trend (*δ*_*c*[*i*],*t*_) hierarchically:

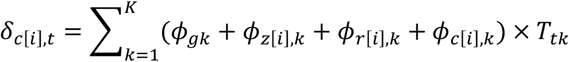

where *ϕ*_*gk*_, *ϕ*_*z*[*i*],*k*_, *ϕ*_*r*[*i*],*k*_, and *ϕ*_*c*[*i*],*k*_ contain the spline’s *K* coefficients for the global, super-region, region, and country-specific time trends. *T*_*tk*_ contains the basis matrix for the natural cubic splines for calendar year *t*. This specification is complemented with the following prior distributions.

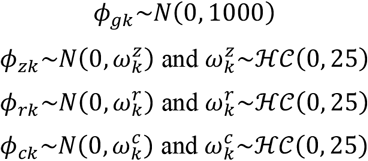

### Covariate modeling

To compare and combine prevalence estimates from different surveys, adjustments are required if those surveys used different outcome definitions and/or eligibility criteria. Covariate modeling, also termed cross-walk in the field of global descriptive epidemiology, is the process by which these adjustments values are estimated. A common way to conduct covariate modeling is to include indicator variables in the regression model, assuming that these fixed effects are constant across all studies and multiplicatively related [9]. Preliminary models using this approach suggested that the resulting adjustment factors could be affected by compositional bias. This type of bias could occur, for example, if studies that required a specific adjustment are more common in countries/regions with lower or higher IPV prevalence, potentially resulting in biased adjustment factors. To circumvent this issue, we chose a robust exact matching identification strategy [27] where the adjustment factors are calculated outside of the main meta-regression models.

Matching methods enable robust estimation by ensuring that observations, with and without the factor to be adjusted for, have the same distribution of other study characteristics (i.e., population surveyed, country, time of data collection, etc.). This is operationalized by matching on the survey’s identifier and this procedure provides us with the ideal comparison group to obtain unbiased adjustment factors. For all adjustment factors except geographical strata, we employed the following procedure:

□ First, we performed exact matching for each adjustment factor separately (Table 1). If more than one match is available from a specific survey, we only kept the one closest to the “*optimal set*” (as described in the preceding section). Studies that surveyed ever-partnered/married women do not always stratify and report their results by current partnership status. To increase the precision of our adjustment factors for this covariate, 89 DHS surveys with publicly available microdata were analyzed.

**Table 1.** List of covariates for which adjustments were estimated characteristics used for exact matching.
□ Second, we calculated the odds ratio comparing prevalence in the observation with the adjustment factor as compared to the reference group within each matched set.
□ Third, we pooled those odds ratios using meta-analytic approaches. Specifically, we used random-effect meta-analysis [28] and, to account for potential variability of the adjustment between regions, stratified results by the seven GBD super regions. This level was deemed appropriate since there were often too few matched observations to estimate adjustment factors for the 21 GBD regions separately. Region-specific adjustment factors were used if a region had more than three estimates; otherwise, the overall adjustment factor was chosen.

| Covariates to adjust | Exact matching on |
| --- | --- |
| IPV definition: “severe violence” (ref.: “all severity”) | Survey, population surveyed, violence type, age, geographical strata, reference partners. |
| IPV type: “physical only” (ref.: “physical and/or sexual”) | Survey, population surveyed, age, geographical strata, severity, reference partners. |
| IPV type: “sexual only” (ref.: “physical and/or sexual”) | Survey, population surveyed, age, geographical strata, severity, reference partners. |
| Population surveyed: “all women” (ref.: “ever-partnered/married”) | Survey, violence type, age, geographical strata, severity, reference partners. |
| Population surveyed: “currently partnered” (ref.: “ever-partnered/married”) | Survey, violence type, age, geographical strata, severity, reference partners. |
| Reference partners: “current/most recent” (ref.: “any current/previous partners”) | Survey, population surveyed, violence type, age, geographical strata, severity. |
| Geographical strata: “urban” or “rural” (ref.: “national”) | Survey, population surveyed, violence type, age, severity, reference partners. |

IPV=intimate partner violence.
\*Separate adjustments estimated for lifetime and past year IPV.

For geographical strata, we used a similar exact matching approach but, since the adjustment factor is not binary (i.e., “rural”, “urban”, “national”), we only used surveys that had information on all three categories. We then pooled the matched surveys using random effect logistic regressions with one random intercept per survey and random slopes that vary by the seven GBD super regions for the “rural” and “urban” areas (referent was “national”).

Once the adjustment factors are estimated, we create a vector *X*_*s[i]*_ summarizing adjustments required for each observation. If the observation pertains to a study in the “*optimal set*”, all indicators in *C*_*s[i]*_ are zeros, meaning that all covariates belong to the reference group. Otherwise, the adjustment is the sum of the log-odds ratio in vector *β*_*r*[*i*]_ multiplied by the binary covariates included in *C*_*s[i]*_, as outlined below.

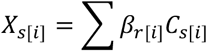

The approach outlined above does not consider the uncertainty in the meta-analyzed odds ratios. To address this, we independently sample values of those odds ratios from their distributions. To ensure appropriate coverage of the parameter space, Latin hypercube sampling is used and several *β*_*r*[*i*]_ vectors are created to represent this uncertainty. Whenever the odds ratio is structurally bound at the null, truncated distributions were used. This would be the case, for example, for estimates of severe IPV that cannot be higher than estimates of severe and non-severe IPV combined. The procedure by which we propagate the uncertainty of these adjustments to final results is described in the section titled *“Computations”*.

Covariates can also be used to improve out-of-sample predictions. For example, if country characteristics like per capita alcohol consumption or gross domestic product can explain between-country variation in IPV. If that is the case, including them in the model could improve estimation of IPV prevalence for countries without any data. However, previous studies on the topic did not find consistent relationships between these country-level covariates and IPV estimates [6, 7, 29]. For this reason, and because data is available for most countries, we do not consider inclusion of covariates to improve out-of-sample predictions.

### Constraints

Past year IPV should be lower or equal to lifetime IPV. Hence, these two IPV outcomes are jointly modelled to ensure that this constraint is respected. This is achieved by jointly performing the meta-regression described above and forcing model predictions for past year 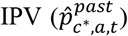 in a new country *c*^*^ (and country with data on only one type of estimate), for age group *a*, and calendar time *t* to be equal or lower to those of their corresponding prediction for lifetime 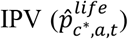, as outlined below:

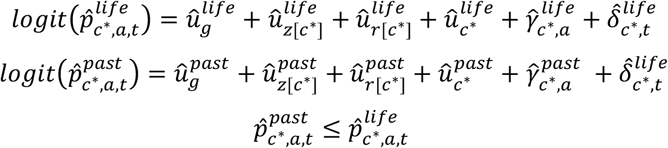

The difference between lifetime and past year IPV should also be relatively small for the youngest age-group of 15-19 years old as these girls and young women have been exposed to the risk of IPV for time periods that are more similar than those of older age groups. Preliminary analyses suggested that including a constraint such that the prevalence ratio of predicted lifetime versus past year IPV among this youngest age group is equal or smaller than 3 improved out-of-sample predictions. This conservative value was chosen based on the empirical observation that prevalence ratio of lifetime to past year IPV among 15-19 years (*a*^*^) old are always less than 3. This constraint was implemented as follow:

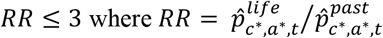

Models where run with and without these constraints to assess the impact of this specification. Adding constraints had only a minor impact on most country estimates but increase the precision of estimates in countries with data on only one type of IPV measure.

### Computations

The posterior distributions of the parameters of interests were obtained using Markov chain Monte Carlo simulations implemented through the JAGS software [30]. Inferences are based on 4 chains of 50,000 iterations (with an adaptation phase of 10,000 iterations and an additional 5,000 used as warm up), thinned at every 20^th^ iteration.

Uncertainty in the estimated log-odds ratios of the adjustment factors are considered by sampling a total of 10 vectors from their estimated distributions using Latin hypercube sampling. For each set, we fitted the Bayesian model using the procedure outlined above. We then mixed all draws from the posterior distributions of the sampled vectors and used these mixed draws to summarize the overall posterior distributions of parameter of interests [31, 32]. Convergence was examined using traceplots and we ensured that the potential scale reduction factor for all parameters and hyperparameters remained close to one [33]. Moreover, we verified that estimates were based on a minimum of roughly 1,000 independent samples from the posterior distributions [34].

### Model validation

The performance of our models is assessed using posterior predictive checks, and both in-sample and out-of-sample comparisons. Graphical posterior predictive checks enable one to visually assess how well simulations from the fitted model compare to the observed data [31]. This procedure is especially useful to understand the ways in which our multilevel model does not fit the observed IPV statistics. By systematically identifying where model predictions are not congruent with the observed data, we were able to improve estimates, through the iterative process of model building and refinement. In addition to this visual inspection, we computed selected summary statistics for in-sample comparisons, such as the median error, absolute error, and the proportion of empirical observations outside the lower and upper credible intervals. We also quantified model performance through out-of-sample comparisons by randomly excluding 20% of countries and 20% of studies from the datasets and comparing their model-predicted age-specific prevalence with the known-but-excluded empirical observations.

### Post-processing

The model described above provides us with estimated parameters for the global, regional, and country-level intercepts *(û*_*g*_, *û*_*z*[*c*]_, *û* _*r*[*c*]_, and *û*_*c*_) that, when combined with the spline’s coefficients 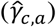 and the time trend 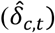, produces estimates of IPV by age and time 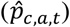 for all countries with available data using the equation below:

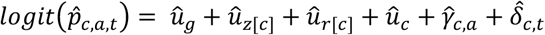

To estimate the prevalence for broader age groups (i.e., 15-49 years), at higher level of aggregation (i.e., regional and global), and for countries without data we first weighted the age-specific prevalence estimates by the age structure of their respective country (in 2018) considering the proportion of women who ever had sex. This is required because our denominator of interest for IPV is not all women but those ever-partnered/married. The definition of a partnership being variable around the world, the proportion of women who have entered the sexually active population is believed to be a better proxy of partnership formation than marriage.

We aggregated estimates of age-specific prevalence at the country level using the country’s own age distribution of the number of women who have ever had sex. If a country did not have any empirical observations informing IPV statistics, they are statistically imputed based on the regional average. The added uncertainty for that country’s estimate is further considered by sampling from the distribution of country-level intercepts (i.e., ∼*N*(0, *σ*_*c*_)). We then aggregate country-specific prevalence estimates at the regional level by summing the number of women having experienced IPV. The same approach was used to obtain global prevalence estimates.

All analyses are carried in the R statistical software [35] and selected packages [34, 36-38]. The code is available from a public repository (https://github.com/pop-health-mod/vawstats-release).

### Ethics

Ethics approval for secondary analyses of individual-level data to estimate covariate adjustments factors was obtained from McGill University’s Faculty of Medicine *Institutional Review Board*.

## Results

A total of 307 unique studies conducted between 2000-2018, from 154 countries, territories, or areas (Figure 5), covering all 21 regions of the world, and totaling 1,767,802 unique women responses, informed lifetime IPV (Table 2). Past year IPV had a slightly higher number of studies (n=332), countries, territories, or areas represented (n=159; Figure 6), totaling 1,763,989 individual responses. For lifetime and past year IPV surveys were conducted between 2000-2018, the median year of data collection were both 2011-2012.

**Table 2.**
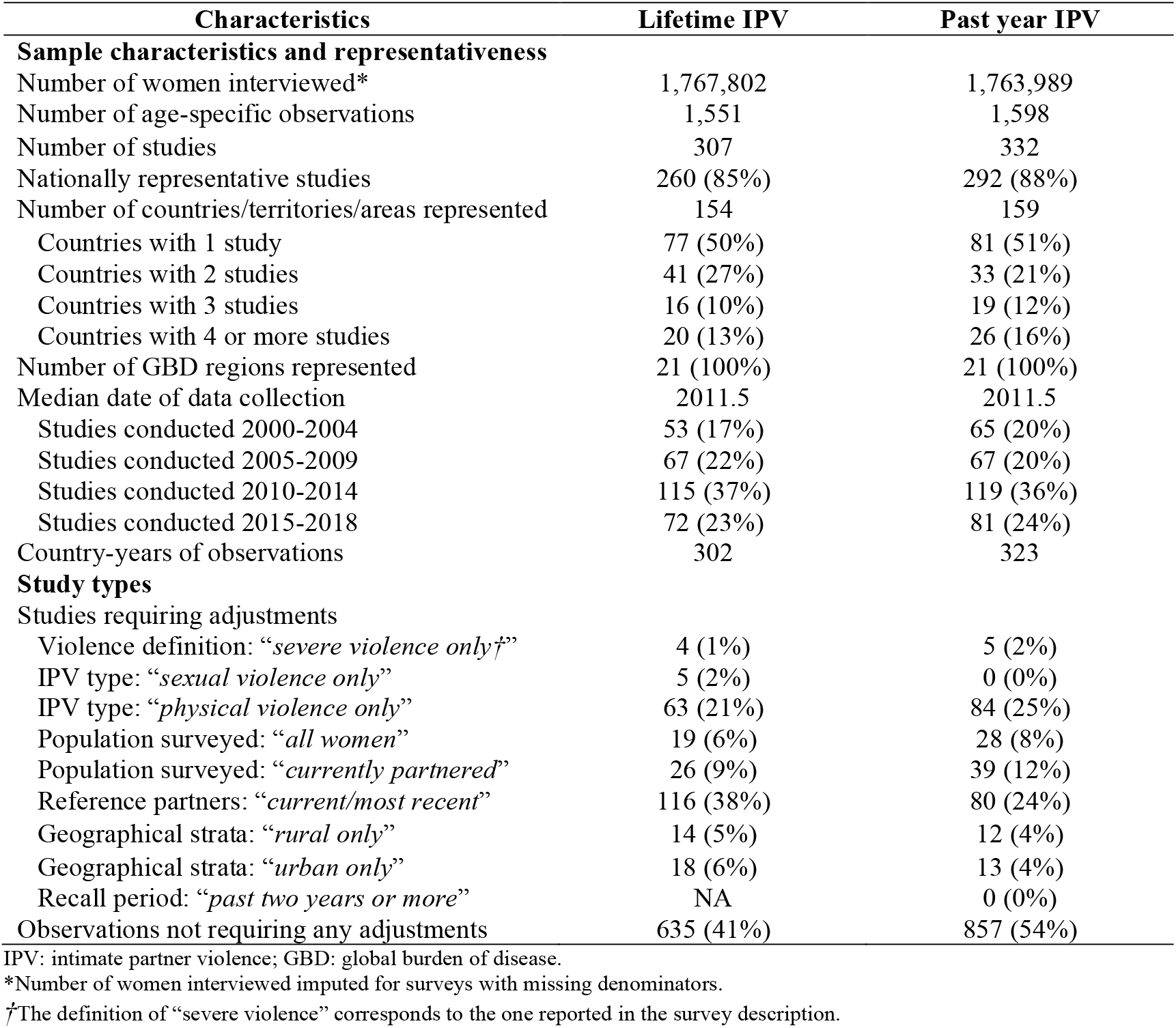
Characteristics of studies conducted between 2000 to 2018 measuring lifetime and past year intimate partner violence (IPV) informing estimates of global, regional, and national violence against women statistics.

**Figure 5.**
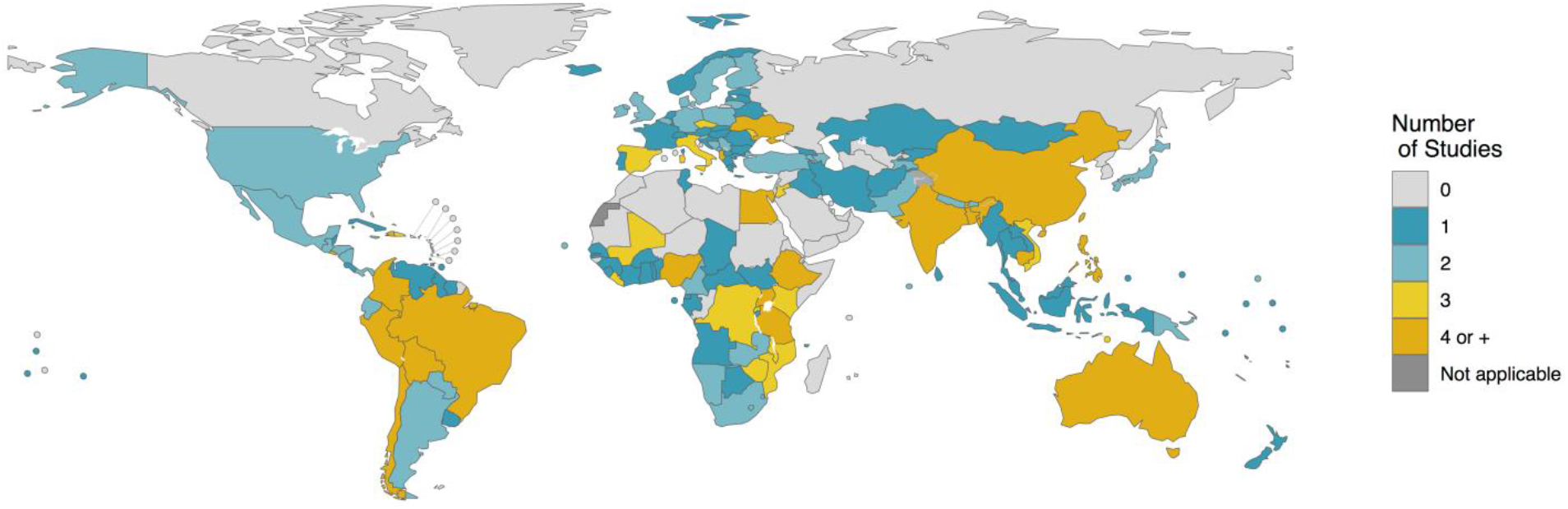
Map of data availability informing estimates of lifetime intimate partner violence (IPV) for the reference period 2000-2018. (Both nationally and sub-nationally representative studies are included.) Disclaimer: The boundaries and names shown and the designations used on this map do not imply the expression of any opinion whatsoever on the part of the World Health Organization concerning the legal status of any country, territory, city or area or of its authorities, or concerning the delimitation of its frontiers or boundaries. Dotted and dashed lines on maps represent approximate border lines for which there may not yet be full agreement.

**Figure 6.**
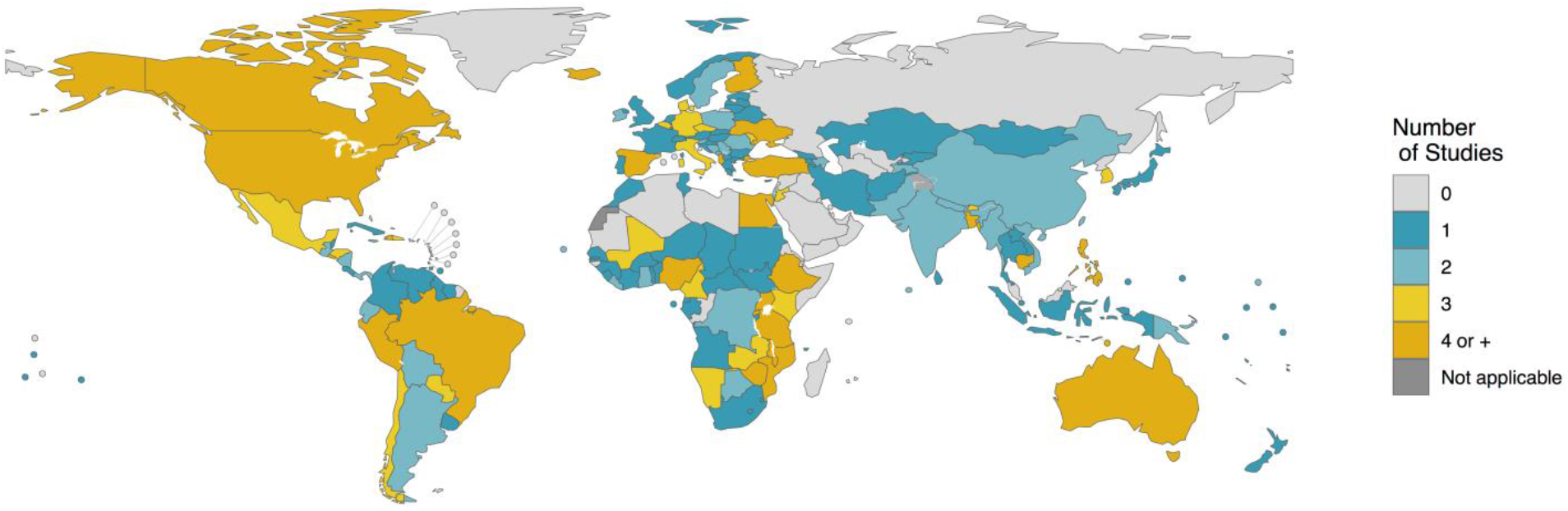
Map of data availability informing estimates of past year intimate partner violence (IPV) for the reference period 2000-2018. (Both nationally and sub-nationally representative studies are included.) Disclaimer: The boundaries and names shown and the designations used on this map do not imply the expression of any opinion whatsoever on the part of the World Health Organization concerning the legal status of any country, territory, city or area or of its authorities, or concerning the delimitation of its frontiers or boundaries. Dotted and dashed lines on maps represent approximate border lines for which there may not yet be full agreement.

### Adjustment factors

Roughly half of IPV observations pertain to the *optimal set* of surveys that collected IPV information using “*gold standard*” definitions and methods (Table 2). The most common adjustments required for lifetime (38%) and past year (24%) IPV studies was that the outcome definition measured violence from a *current and/or most recent partner only* as opposed to *any current/previous partner*. The second most common adjustment was for “*physical violence only*”, with 21% (lifetime) and 25% (past year) of surveys requiring adjustment. A slightly smaller proportion of studies surveyed “*currently partnered women*” only, whereas the “gold-standard” VAW questionnaire would have asked IPV questions to ever-partnered women. Table 3 presents these odds ratios for the two IPV outcomes. The detailed forest plots for all meta-analyses are presented as supplemental materials (Figure S1-15).

**Table 3.** Results of random effects meta-analysis for different adjustment factors, stratified by super region, for lifetime intimate partner violence (IPV) and past year IPV.

| **Adjustment factors by super regions**<br>**(and overall)** | **Lifetime IPV**<br>**OR (95%CI)** | **Past year IPV**<br>**OR (95%CI)** |
| --- | --- | --- |
| **Severe violence** (ref. all severity levels)§ |  |  |
| Central Europe, Eastern Europe & Central Asia | NA | 0.39 (0.34-0.45) |
| High Income | NA | NA |
| Latin America & Caribbean | 0.51 (0.43-0.60) | 0.62 (0.56-0.69) |
| North Africa & Middle East | NA | 0.47 (0.34-0.64) |
| South Asia | NA | 0.52 (0.44-0.61) |
| South-East Asia, East Asia & Oceania | 0.36 (0.25-0.50) | 0.57 (0.48-0.67) |
| Sub-Saharan Africa | 0.37 (0.33-0.42) | 0.61 (0.58-0.64) |
| *Overall* | 0.38 (0.34-0.42) | 0.57 (0.55-0.60) |
| **Physical IPV** (ref. physical and/or sexual IPV) |  |  |
| Central Europe, Eastern Europe & Central Asia | 0.93 (0.92-0.95) | 0.95 (0.93-0.97) |
| High Income | 0.86 (0.84-0.88) | 0.76 (0.70-0.83) |
| Latin America & Caribbean | 0.90 (0.87-0.93) | 0.84 (0.81-0.88) |
| North Africa & Middle East | 0.93 (0.90-0.96) | 0.83 (0.76-0.91) |
| South Asia | 0.82 (0.73-0.93) | 0.78 (0.67-0.90) |
| South-East Asia, East Asia & Oceania | 0.79 (0.73-0.84) | 0.78 (0.73-0.83) |
| Sub-Saharan Africa | 0.82 (0.78-0.86) | 0.79 (0.76-0.82) |
| *Overall* | 0.86 (0.84-0.87) | 0.81 (0.79-0.83) |
| **Sexual IPV** (ref. physical and/or sexual IPV) |  |  |
| Central Europe, Eastern Europe & Central Asia | 0.22 (0.20-0.25) | 0.22 (0.18-0.27) |
| High Income | 0.29 (0.24-0.34) | 0.27 (0.22-0.32) |
| Latin America & Caribbean | 0.26 (0.23-0.29) | 0.31 (0.28-0.35) |
| North Africa & Middle East | 0.19 (0.15-0.25) | 0.28 (0.18-0.43) |
| South Asia | 0.28 (0.21-0.37) | 0.36 (0.27-0.47) |
| South-East Asia, East Asia & Oceania | 0.31 (0.26-0.36) | 0.35 (0.28-0.44) |
| Sub-Saharan Africa | 0.27 (0.24-0.29) | 0.32 (0.29-0.35) |
| *Overall* | 0.26 (0.25-0.28) | 0.31 (0.29-0.33) |
| **All women surveyed** (ref. ever-partnered) |  |  |
| Central Europe, Eastern Europe & Central Asia | NA | NA |
| High Income | NA | NA |
| Latin America & Caribbean | NA | NA |
| North Africa & Middle East | NA | NA |
| South Asia | NA | NA |
| South-East Asia, East Asia & Oceania | NA | NA |
| Sub-Saharan Africa | NA | NA |
| *Overall* | 0.79 (0.74-0.84) | * |
| **Currently partnered women surveyed** (ref. ever-partnered) |  |  |
| Central Europe, Eastern Europe & Central Asia | 0.81 (0.71-0.92) | 0.88 (0.80-0.98) |
| High Income | NA | NA |
| Latin America & Caribbean | 0.85 (0.80-0.90) | 0.89 (0.82-0.96) |
| North Africa & Middle East | 0.94 (0.91-0.99) | 1.00 (0.99-1.02) |
| South Asia | 0.98 (0.97-0.98) | 1.06 (1.04-1.07) |
| South-East Asia, East Asia & Oceania | 0.92 (0.89-0.96) | 1.00 (0.98-1.02) |
| Sub-Saharan Africa | 0.93 (0.92-0.94) | 1.02 (1.00-1.04) |
| *Overall* | 0.91 (0.90-0.93) | 0.99 (0.97-1.01) |

**Table 3 (con't).** Results of random effects meta-analysis for different adjustment factors, stratified by super region, for lifetime intimate partner violence (IPV) and past year IPV.
| Adjustment factors by super regions<br>(and overall) | Lifetime IPV<br>OR (95%CI) | Past year IPV<br>OR (95%CI) |
| --- | --- | --- |
| **Partner perpetrating is current or most recent** (ref. any current or previous partners) |  |  |
| Central Europe, Eastern Europe & Central Asia | NA | NA |
| High Income | NA | 0.68 (0.58-0.81) |
| Latin America & Caribbean | 0.82 (0.71-0.95) | 0.99 (0.98-1.00) |
| North Africa & Middle East | NA | NA |
| South Asia | NA | NA |
| South-East Asia, East Asia & Oceania | 0.88 (0.82-0.95) | 0.99 (0.99-1.00) |
| Sub-Saharan Africa | 0.89 (0.86-0.92) | 0.98 (0.95-1.01) |
| Overall | 0.84 (0.77-0.93) | 0.97 (0.95-1.00) |
| **Geographical urban strata** (ref. nationally representative) |  |  |
| Central Europe, Eastern Europe & Central Asia | 0.98 (0.89-1.08) | 0.90 (0.82-0.98) |
| High Income | NA | NA |
| Latin America & Caribbean | 1.06 (0.97-1.16) | 1.04 (0.96-1.13) |
| North Africa & Middle East | 0.86 (0.79-0.95) | 0.88 (0.81-0.96) |
| South Asia | 0.76 (0.70-0.84) | 0.77 (0.71-0.83) |
| South-East Asia, East Asia & Oceania | 0.91 (0.83-1.00) | 0.93 (0.85-1.01) |
| Sub-Saharan Africa | 0.99 (0.91-1.08) | 0.98 (0.91-1.07) |
| Overall | 0.92 (0.85-1.01) | 0.91 (0.84-0.99) |
| **Geographical rural strata** (ref. nationally representative) |  |  |
| Central Europe, Eastern Europe & Central Asia | 1.00 (0.92-1.08) | 1.05 (0.99-1.12) |
| High Income | NA | NA |
| Latin America & Caribbean | 0.88 (0.82-0.95) | 0.94 (0.89-0.99) |
| North Africa & Middle East | 1.14 (1.06-1.23) | 1.08 (1.03-1.14) |
| South Asia | 1.14 (1.06-1.22) | 1.13 (1.07-1.19) |
| South-East Asia, East Asia & Oceania | 1.04 (0.96-1.12) | 1.03 (0.97-1.09) |
| Sub-Saharan Africa | 1.00 (0.93-1.08) | 1.01 (0.96-1.06) |
| Overall | 1.03 (0.96-1.11) | 1.04 (0.99-1.09) |

95%CI: 95% confidence interval; IPV: intimate partner violence; OR: odds ratio; VAW: violence against women.

Our meta-analyses indicate that, overall, the odds of having experienced IPV for women asked about their experience of only severe violence are 62% (lifetime) and 43% (past year) lower than if these women had reported on IPV for all severity levels (Table 3). Examining results by super-regions, we notice that contribution of severe violence to IPV is relatively more important in *Latin America & the Caribbean* for both lifetime and past year IPV, followed by *Sub-Saharan Africa* (especially past year IPV), as indicated by the highest odds ratios for these regions.

When surveys report “*physical IPV only*”, we find that the discrepancy with “*physical and/or sexual IPV*” estimates is greatest in *South East Asia, East Asia & Oceania*, as well as *South Asia* and *Sub-Saharan Africa* for lifetime IPV (Table 3). In addition to those super-regions, the *High Income* one also exhibited high discrepancy for past year IPV. For both lifetime and past year IPV, the odds of reporting IPV are reduced by 14-19% when women are asked about physical IPV only.

The surveys’ denominator (i.e. who is eligible to answer the IPV questions) affects IPV prevalence. Overall, the odds of reporting IPV –when only currently partnered women are surveyed– are reduced by 9% for lifetime IPV and 1% for past year IPV, respectively, as compared to observations where all ever-partnered women are included (Table 3). When the surveys’ denominator includes all women, and not only those that have ever been partnered, the odds of experiencing IPV is lower since a certain proportion of women, namely never-married/partnered women, will not have been exposed to the risk of IPV. Since there are no surveys of past year IPV that could be matched and compared, the odds ratios for lifetime IPV are used to adjust the surveys of past year IPV that recorded violence among all women.

Some surveys measured IPV perpetrated by the current and/or most recent partner only whereas others referred to IPV by all partners. Overall, the odds of reporting IPV is 16% lower for lifetime IPV if the question refers to the current or most recent partner compared to any current or previous partner. For past year IPV, the difference is much smaller: a 3% reduction in the odds of reporting IPV (Table 3).

Our results also show marked regional variations in IPV prevalence by urban and rural areas (Table 3). IPV is higher in urban areas of *Latin America & Caribbean* as compared to the national average for both lifetime and past year IPV, whereas the opposite pattern of higher prevalence in rural areas is observed for all other regions, especially for *North Africa & Middle East* and *South Asia*.

### Model validation

The full graphical posterior predictive checks for lifetime IPV (Figure S16) and past year IPV (Figure S17) are presented as supplemental materials for all regions. As a representative example, the graphical posterior checks for the *Western* region of *Sub-Saharan Africa* with both the observed age-specific prevalence estimates and the model predictions are presented in Figure 7. Overall, we find that the model fits the data well and that differences between data and model predictions, if any, are usually small and well within the uncertainty intervals of the prevalence estimates and model predictions. Further, in-sample comparisons suggest that prediction errors are very close to the expected null values indicating good fit with the empirical data and excellent coverage of uncertainty interval (Table 4). We also explored how those in-sample comparisons metrics varied by GBD regions and time periods (2000-04, 2005-09, 2010-14, 2015-18). These analyses revealed that median errors were smaller than 1% for all regions and time periods.

**Table 4.** In-sample comparisons of model fits with empirical data.

| VAW outcomes<br>(Nb. observations) | Median (in % point) |  | Outside 95% CrI |  |
| --- | --- | --- | --- | --- |
|  | Error | Absolute error | Below (%) | Above (%) |
| Lifetime IPV (1,551) | 0.0% | 1.5% | 1.3% | 1.3% |
| Past year IPV (1,598) | 0.0% | 1.0% | 2.2% | 1.9% |

95%CrI: 95% credible interval; IPV: intimate partner violence.
Comparisons defined as “*error = observed – predicted*”.

**Figure 7.**
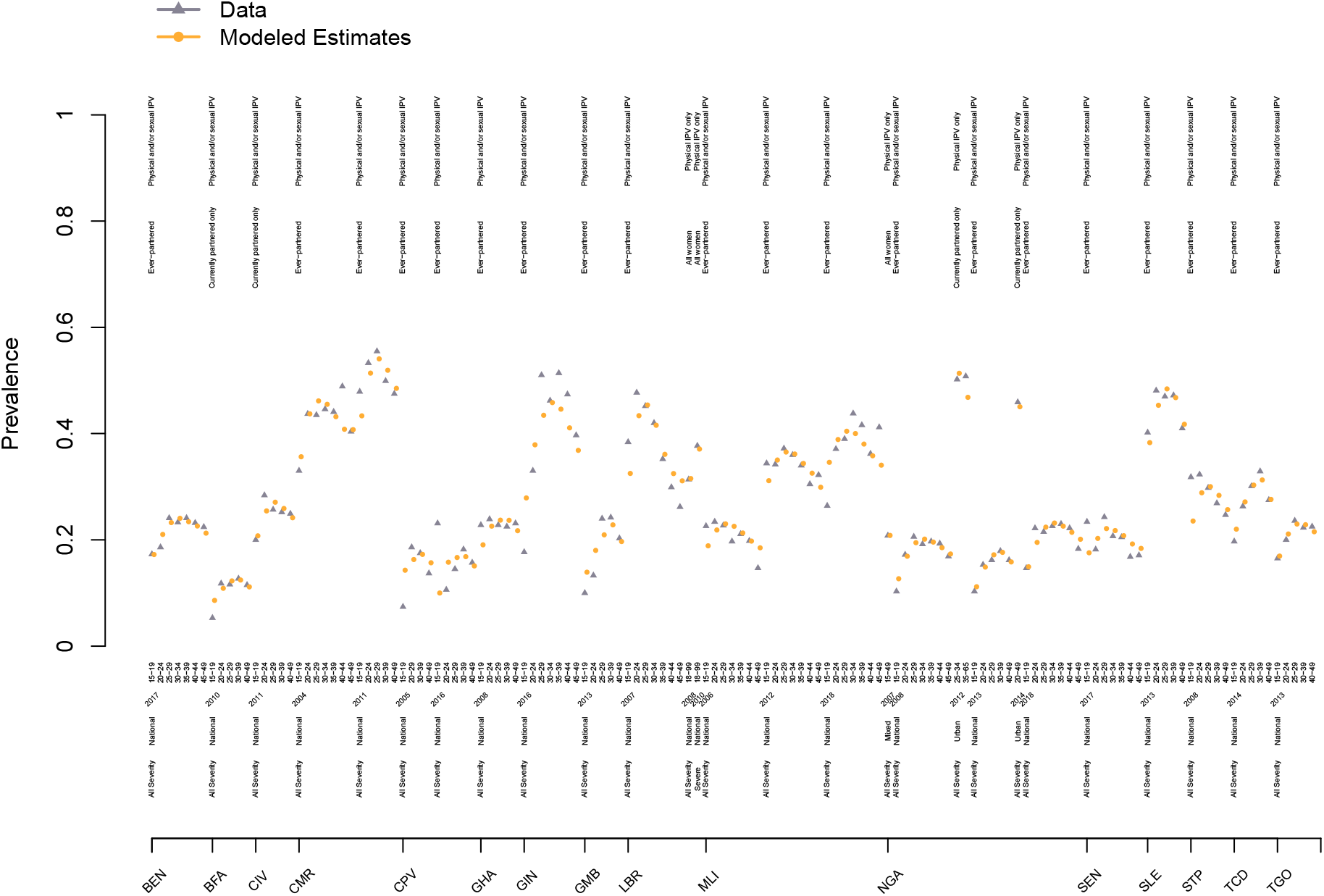
Graphical posterior predictive checks for 16 countries of the Western region of sub-Saharan Africa. Average prevalence for the observed data (triangle) are presented in grey while the model predictions are in yellow (round dots). The vertical lines correspond to the 95% confidence or uncertainty intervals of the data and prediction, respectively. The annotations above the country names described the type of prevalence estimates displayed, the year of data collection, the age group, the surveyed population, and the type of intimate partner violence recorded. (BEN: Benin; BFA: Burkina Faso; CIV: Côte d’Ivoire; CMR: Cameroon; CPV: Cabo Verde; GHA: Ghana; GIN: Guinea; GMB: The Gambia; LBR: Liberia; MLI: Mali; NGA: Nigeria; SEN: Senegal; SLE: Sierra Leone; STP: Sao Tome and Principe; TCD: Chad; TGO: Togo.)

Our out-of-sample comparisons where we excluded 20% of countries and compared the known-but-excluded country-level observations with model predictions are also in accordance with empirical estimates for both lifetime and past year IPV prevalence: median errors are reasonably close to zero. The median absolute error quantifies the typical magnitude of the predictions’ errors, regardless of their direction. It is less than 8% points for lifetime IPV and 4% points for past year IPV. The models’ predictions include the point estimates of the known-but-excluded observations close to 95% of the time (as expected) for the two IPV outcomes, suggesting that the model is appropriately propagating uncertainty.

Similarly, we can test the model’s ability to predict new surveys by excluding 20% of studies (instead of countries, as above). Here again, the model’s predictions are in accordance with the known-but-excluded survey estimates with small median and absolute errors and appropriate coverage of uncertainty intervals (Table 6).

**Table 5.** Out-of-sample comparisons of age-specific model-predicted prevalence in 20% of randomly excluded countries with the empirical observations from these countries (including territories and areas).

| VAW outcomes (Nb. countries excluded) | Median (in % point) |  | Outside 95% CrI |  |
| --- | --- | --- | --- | --- |
|  | Error | Absolute error | Below (%) | Above (%) |
| Lifetime IPV (30) | 1.3% | 7.6% | 1.4% | 1.0% |
| Past year IPV (31) | 0.6% | 3.8% | 1.9% | 1.6% |

95%CrI: 95% credible interval; IPV: intimate partner violence.
Comparisons defined as “*error = observed – predicted*”. To improve stability of the metrics used for out-of-sample comparison, the process was repeated 20 times and the median estimates are presented above.

**Table 6.**
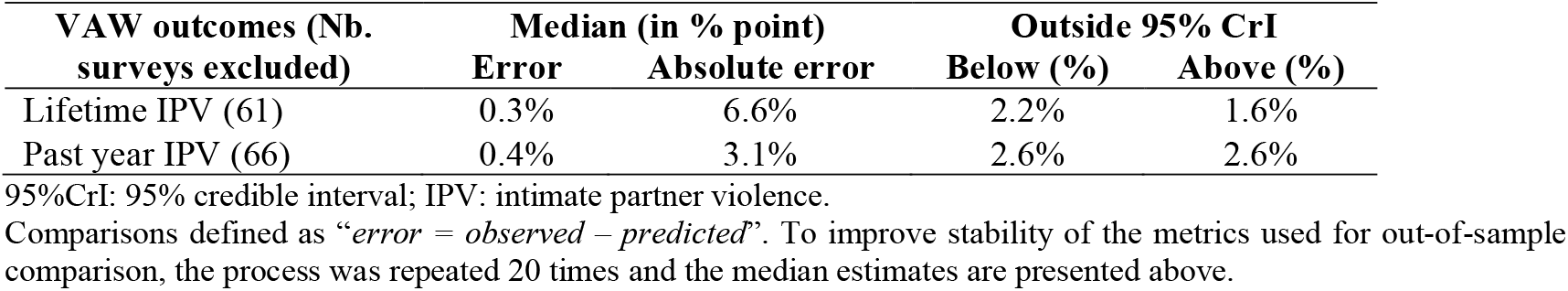
Out-of-sample comparisons of age-specific model-predicted prevalence in 20% of randomly excluded studies with the empirical observations from these studies.

## Discussion

Elimination of all forms of VAW remains a global priority. Reliable, accurate, and comparable VAW statistics are essential to monitor progress towards this goal. This paper describes a framework for modeling global, regional, and national estimates of IPV statistics that specifically addresses key data and measurement issues [2, 8]. The principal features of this model are its ability to pool both nationally and sub-nationally representative population surveys, account for heterogeneous age groups and age trends, accommodate different surveyed populations, adjust for differences in survey instruments, and efficiently propagate model uncertainty to model outputs.

The proposed multilevel framework is especially flexible in its parameterization. For example, both age trends and adjustment factors can vary by regions. For the latter, we used a robust identification strategy to estimate adjustments through exact matching of observations by survey identifier, whenever possible. We find that the relative contribution of physical IPV only to lifetime IPV is highest in the *Central Europe, Eastern Europe & Central Asia* and *North Africa & Middle East* super regions. As for the population surveyed, the inclusion of different groups of women according to partnership status influences prevalence estimates, especially for lifetime IPV, with ever-partnered women consistently providing higher prevalence than when surveying all women. Our estimation of adjustment factors offers interesting insights into the epidemiology of IPV by rural/urban areas. In all regions, except in *Latin America & Caribbean*, IPV is more common in rural areas as compared to the national average.

Some limitations need to be considered in interpreting our results. First, and most importantly, IPV statistics are based on self-reports. Violence is a sensitive topic and it is likely that some violence survivors under-report their experiences for a variety of reasons. However, we only included surveys that used act-specific questions; questions that are recognized for their ability to facilitate disclosure and elicit more accurate reports. Studies comparing men and women reports of past-year male-perpetrated IPV concluded that IPV indicators such as the ones included in this study are “reasonably reliable” [21]. As evidence accrues on the sensitivity and specificity of different methods for measuring IPV, prevalence estimates could be adjusted, if warranted, for the imperfect nature of those survey instruments.

A second limitation is that our analyses of IPV do not include psychological IPV, a widespread form of violence that can takes may different forms [39]. This diversity of forms, from engendering fears, to verbal abuse, humiliation, and enforcing social isolation, has impeded the adoption of a uniform definition of psychological IPV. This is an active research area and WHO has convened several expert meetings on this specific topic over the last few years and is currently doing empirical analyses of existing data. The current lack of consensus nevertheless hampers the inclusion of psychological IPV in our estimates for the time being.

A third limitation is that some populous countries, such as Russia and others in North Africa and the Middle East, have yet to conduct recent VAW surveys. The absence of national level data on the burden of violence is a significant challenge for establishing the magnitude and patterns of IPV. This evidence is crucial to advocating for prioritization of investments in national and local policies and programs aimed at eliminating IPV and other forms of VAW. Having said this, most of the world’s most populous countries have conducted such surveys in the last 18 years (Bangladesh, Brazil, India, Indonesia, Mexico, Nigeria, Pakistan, and the United States) such that more than 90% of the world’s women and girls reside in a country with at least one survey data point for both lifetime and past year IPV.

In conclusion, we have described to reproducible levels of details a flexible modeling framework to estimate global, regional, and national prevalence of IPV, with reassuring results from both in-sample and out-of-sample comparisons. Following proposed best reporting practice [40], the information provided in this study will support the accurate interpretation and responsible use of IPV statistics for informing national and international VAW prevention interventions and policies, and the monitoring efforts towards the elimination of VAW as part of the SDG agenda.

## Data Availability

The database has yet to be publicly released.

## Abbreviations

DHS: Demographic and Health Survey
GBD: Global burden of disease
IPV: intimate partner violence
SDG: Sustainable Development Goals
VAW: violence against women

## Acknowledgments

We are indebted to participants of the “*Expert Group Meeting on Methodological and Measurement Issues for VAW Statistics”*, those of the *“Technical Advisory Group on VAW Estimation and Data”*, and those of the *“Interagency Working Group on VAW Estimation and Data”* for helpful comments and discussion on earlier versions of the models. We also acknowledge Kody Crowell for contributions to data management and Gretchen Stevens and Max Petzold for their insightful statistical advice. MMG’s research program is funded by a *Tier II Canada Research Chair* in *Population Health Modeling*. This study was supported by funding provided by the *Department of International Development* (DFID) to the *Joint WHO Joint Programme on Strengthening VAW Data Collection, Reporting and Use*.

