## Supplemental Materials for "A framework to model global, regional, and national estimates of intimate partner violence"

### Appendix - A framework for modeling global, regional, and national estimates of violence against women statistics

#### Table of Contents

**Figure S1.** Forest plot of the random effect meta-analysis for severe lifetime intimate partner violence (IPV), as compared to violence from all severity levels. ... 2
**Figure S2.** Forest plot of the random effect meta-analysis for severe past year intimate partner violence (IPV), as compared to violence from all severity levels. ... 3
**Figure S3.** Forest plot of the random effect meta-analysis for lifetime physical intimate partner violence (IPV) only, as compared to physical and/or sexual violence. ... 4
**Figure S4.** Forest plot of the random effect meta-analysis for past year physical intimate partner violence (IPV) only, as compared to physical and/or sexual violence. ... 5
**Figure S5.** Forest plot of the random effect meta-analysis for lifetime sexual intimate partner violence (IPV) only, as compared to physical and/or sexual violence. ... 6
**Figure S6.** Forest plot of the random effect meta-analysis for past year for sexual intimate partner violence (IPV) only, as compared to physical and/or sexual violence. ... 7
**Figure S7.** Forest plot of the random effect meta-analysis for lifetime intimate partner violence (IPV) when all women are surveyed, as compared to ever-partnered women. ... 8
**Figure S8.** Forest plot of the random effect meta-analysis for lifetime intimate partner violence (IPV) when currently-partnered women are surveyed, as compared to ever-partnered women. ... 9
**Figure S9.** Forest plot of the random effect meta-analysis for past year intimate partner violence (IPV) when currently partnered women are surveyed, as compared to ever-partnered women. ... 10
**Figure S10.** Forest plot of the random effect meta-analysis for lifetime intimate partner violence (IPV) when the reference partner is the current or most recent one, as compared to any current or previous ones. ... 11
**Figure S11.** Forest plot of the random effect meta-analysis for past year intimate partner violence (IPV) when the reference partner is the current or most recent one, as compared to any current or previous ones. ... 12
**Figure S12.** Plot of the data used to estimate, through random-effect logistic regression, the adjustment factor for lifetime intimate partner violence (IPV) in urban regions, as compared to a nationally representative sample. ... 13
**Figure S13.** Plot of the data used to estimate, through random-effect logistic regression, the adjustment factor for past year intimate partner violence (IPV) in urban regions, as compared to a nationally representative sample. ... 14
**Figure S14.** Plot of the data used to estimate, through random-effect logistic regression, the adjustment factor for lifetime intimate partner violence (IPV) in rural regions, as compared to a nationally representative sample. ... 15
**Figure S15.** Plot of the data used to estimate, through random-effect logistic regression, the adjustment factor for past year intimate partner violence (IPV) in rural regions, as compared to a nationally representative sample. ... 16
**Figure S16.** Posterior predictive checks for the lifetime intimate partner violence (IPV) model. ... 17
**Figure S17.** Posterior predictive checks for the past year intimate partner violence (IPV) model. ... 18

*(Note that the effective sample sizes are used. This can explain the slight inequalities between the denominators.)*

**Figure S1.** Forest plot of the random effect meta-analysis for severe lifetime intimate partner violence (IPV), as compared to violence from all severity levels.

| Source | OR (95% CI) |
|---|---|
| **SuperRegion = High Income** | |
| Japan – (Yoshihama et al., 2005) | 0.24 [0.16; 0.35] |
| Total | 0.24 [0.16; 0.35] |
| Heterogeneity: not applicable | |
| **SuperRegion = Central Europe, Eastern Europe & Central Asia** | |
| Moldova – (NCPM/Moldova, and ORC Macro, 2006) | 0.39 [0.34; 0.46] |
| Serbia – (Otasevic et al., 2005) | 0.30 [0.22; 0.40] |
| Total | 0.35 [0.27; 0.46] |
| Heterogeneity: $\chi^2_1 = 2.57$ ($P = .11$), $I^2 = 61\%$ | |
| **SuperRegion = South–East Asia, East Asia & Oceania** | |
| Cambodia – (NIS/Cambodia, DGH/Cambodia, and ORC Macro, 2001) | 0.53 [0.42; 0.69] |
| Cambodia – (NIPH/Cambodia, NIS/Cambodia, and ORC Macro, 2006) | 0.49 [0.36; 0.66] |
| Thailand – (Kanchanachitra et al., 2005) | 0.22 [0.17; 0.29] |
| Thailand – (Kanchanachitra et al., 2005) | 0.25 [0.19; 0.32] |
| Samoa – ( Posiulai et al., 2005) | 0.40 [0.32; 0.50] |
| Total | 0.36 [0.25; 0.50] |
| Heterogeneity: $\chi^2_4 = 35.39$ ($P < .001$), $I^2 = 89\%$ | |
| **SuperRegion = South Asia** | |
| Bangladesh – (Garcia–Moreno et al., 2005) | 0.22 [0.17; 0.27] |
| Bangladesh – (Garcia–Moreno et al., 2005) | 0.17 [0.14; 0.21] |
| India – (IIPS/India, and Macro International, 2007) | 0.35 [0.34; 0.37] |
| Total | 0.24 [0.14; 0.39] |
| Heterogeneity: $\chi^2_2 = 57.49$ ($P < .001$), $I^2 = 97\%$ | |
| **SuperRegion = Latin America & Caribbean** | |
| Bolivia – (Gutiérrez, 2004) | 0.66 [0.61; 0.70] |
| Brazil – (Schraiber et al., 2005) | 0.49 [0.37; 0.65] |
| Brazil – (Schraiber et al., 2005) | 0.48 [0.38; 0.60] |
| Colombia – (Profamilia/Colombia, 2000) | 0.41 [0.37; 0.45] |
| Dominican Republic – (Achécar et al., 2003) | 0.61 [0.53; 0.71] |
| Dominican Republic – (CESDEM/Republica Dominicana and Macro International, 2008) | 0.55 [0.47; 0.65] |
| Haïti – (Cayemittes et al., 2001) | 0.51 [0.39; 0.66] |
| Haïti – (Cayemittes et al., 2007) | 0.39 [0.31; 0.49] |
| Total | 0.51 [0.43; 0.60] |
| Heterogeneity: $\chi^2_7 = 79.06$ ($P < .001$), $I^2 = 91\%$ | |
| **SuperRegion = North Africa & Middle East** | |
| Egypt – (MHP/Egypt, and NNPC/Egypt, 2006) | 0.33 [0.29; 0.37] |
| Jordan – (DoS/Jordan, and Macro International, 2008) | 0.39 [0.31; 0.49] |
| Total | 0.35 [0.29; 0.42] |
| Heterogeneity: $\chi^2_1 = 2.01$ ($P = .16$), $I^2 = 50\%$ | |
| **SuperRegion = Sub–Saharan Africa** | |
| Cameroon – (INS/Cameroun, and ORC Macro, 2005) | 0.37 [0.31; 0.44] |
| Ethiopia – (Berhane et al., 2005) | 0.23 [0.20; 0.27] |
| Liberia – (LISGIS/Liberia, MOHSW/Liberia, NACP/Liberia, and Macro International, 2008) | 0.41 [0.34; 0.50] |
| Malawi – (NSO/Malawi, and ORC Macro, 2005) | 0.42 [0.37; 0.47] |
| Namibia – (January et al., 2005) | 0.48 [0.38; 0.59] |
| Rwanda – (INSR/Rwanda, and ORC Macro, 2006) | 0.43 [0.36; 0.51] |
| Tanzania – (MoHCDGEC/Tanzania Mainland, MoH/Zanzibar, NBS/Tanzania, OCGS/Zanzibar, and ICF, 2016) | 0.30 [0.24; 0.37] |
| Tanzania – (MoHCDGEC/Tanzania Mainland, MoH/Zanzibar, NBS/Tanzania, OCGS/Zanzibar, and ICF, 2016) | 0.31 [0.25; 0.39] |
| Uganda – (UBOS/Uganda, and Macro International, 2007) | 0.45 [0.37; 0.55] |
| Zambia – (CSO/Zambia, MOH/Zambia, TDRC/Zambia, and University of Zambia, 2009) | 0.32 [0.28; 0.37] |
| Zimbabwe – (CSO/Zimbabwe, and Macro International, 2007) | 0.45 [0.39; 0.51] |
| Total | 0.37 [0.33; 0.42] |
| Heterogeneity: $\chi^2_{10} = 64.99$ ($P < .001$), $I^2 = 85\%$ | |
| Total | 0.38 [0.34; 0.42] |

Heterogeneity: $\chi^2_{31} = 511.19$ ($P < .001$), $I^2 = 94\%$

Residual heterogeneity: $\chi^2_{25} = 241.52$ ($P < .001$), $I^2 = 90\%$

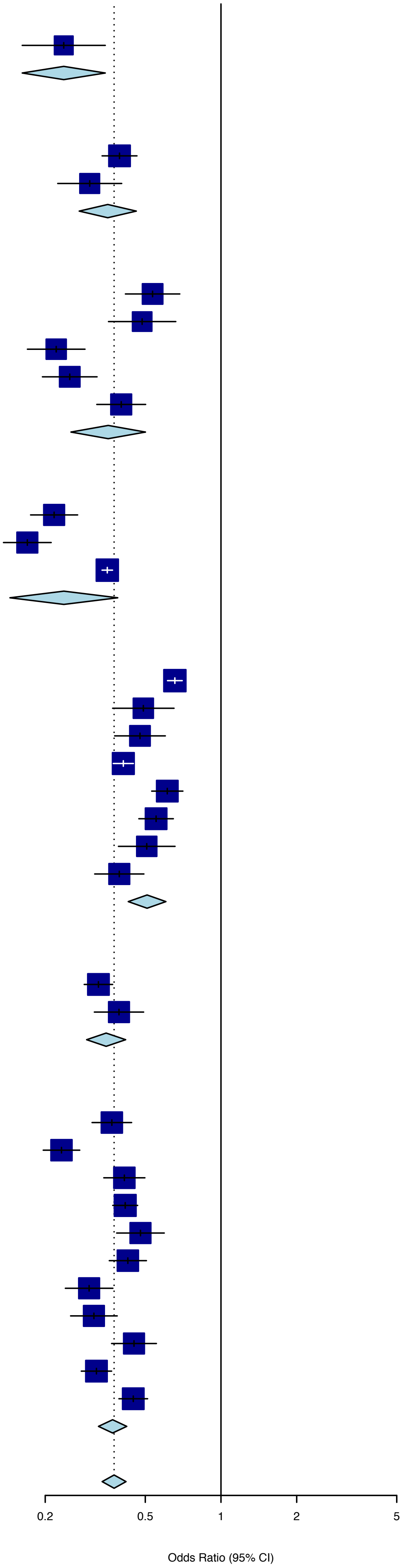

**Figure S2.** Forest plot of the random effect meta-analysis for severe past year intimate partner violence (IPV), as compared to violence from all severity levels.

| Source | OR (95% CI) |
|---|---|
| **SuperRegion = North Africa & Middle East** | |
| Afghanistan – (CSO/Afghanistan, MPH/Afghanistan, and ICF, 2017) | 0.32 [0.31; 0.34] |
| Egypt – (MHP/Egypt and ICF International, 2015) | 0.37 [0.33; 0.42] |
| Jordan – (DoS/Jordan, and Macro International, 2008) | 0.62 [0.53; 0.72] |
| Jordan – (DoS/Jordan, and ICF International, 2013) | 0.64 [0.58; 0.71] |
| Jordan – (DoS/Jordan, and ICF, 2019) | 0.48 [0.43; 0.54] |
| Total | 0.47 [0.34; 0.64] |
| Heterogeneity: $\chi^2_4 = 214.06$ ($P < .001$), $I^2 = 98\%$ | |
| **SuperRegion = Sub–Saharan Africa** | |
| Angola – (INE/Angola, MINSA/Angola, and ICF, 2017) | 0.52 [0.48; 0.56] |
| Burundi – (DHS, 2016) | 0.75 [0.69; 0.81] |
| Benin – (INSAE/Bénin, and ICF, 2019) | 0.61 [0.54; 0.71] |
| Burkina Faso – (DHS, 2010) | 0.53 [0.47; 0.59] |
| Cote d'Ivoire – (DHS, 2012) | 0.54 [0.49; 0.61] |
| Cameroon – (INS/Cameroun, and ORC Macro, 2005) | 0.56 [0.50; 0.63] |
| Cameroon – (INS/Cameroun, and ICF International, 2012) | 0.51 [0.46; 0.56] |
| Democratic Republic of the Congo – (MP/Congo, and Macro International, 2008) | 0.60 [0.55; 0.67] |
| Democratic Republic of the Congo – (MPSMRM/Congo, MSP/Congo, and ICF International, 2014) | 0.57 [0.52; 0.62] |
| Comoros – (DGSP/Comores, and ICF International, 2014) | 0.44 [0.31; 0.62] |
| Ethiopia – (CSA/Ethiopia, and ICF, 2016) | 0.61 [0.54; 0.68] |
| Gabon – (DGS/Gabon, and ICF International, 2013) | 0.59 [0.53; 0.65] |
| Ghana – (GSS, GHS, and Macro International, 2009) | 0.58 [0.48; 0.70] |
| Gambia – (GBOS, and ICF International, 2014) | 0.60 [0.49; 0.74] |
| Kenya – (CBS/Kenya, MOH/Kenya, and ORC Macro, 2004) | 0.60 [0.55; 0.65] |
| Kenya – (KNBS/Kenya, NACC/Kenya, NASCP/Kenya, MPHS/Kenya, and KMRI, 2010) | 0.62 [0.57; 0.68] |
| Kenya – (KNBS/Kenya, MOH/Kenya, NACC/Kenya, KMRI/Kenya, NCPD/Kenya, 2015) | 0.62 [0.56; 0.69] |
| Liberia – (LISGIS/Liberia, MOHSW/Liberia, NACP/Liberia, and Macro International, 2008) | 0.56 [0.51; 0.62] |
| Mali – (Samaké et al., 2007) | 0.59 [0.54; 0.64] |
| Mali – (CPS/SSDSPF/Mali, INSTAT/Mali, INFO–STAT/Mali, and ICF International, 2014) | 0.54 [0.47; 0.61] |
| Mali – (INSTAT/Mali, CPSSSD, and ICF, 2019) | 0.55 [0.48; 0.63] |
| Moçambique – (MISAU/Moçambique, INE/Moçambique, and ICF International, 2013) | 0.50 [0.45; 0.55] |
| Moçambique – (INS, INE, PNCM/Moçambique and ICF, 2019) | 0.53 [0.45; 0.62] |
| Malawi – (NSO/Malawi, and ORC Macro, 2005) | 0.69 [0.64; 0.74] |
| Malawi – (NSO/Malawi, and ICF Macro, 2011) | 0.77 [0.70; 0.85] |
| Malawi – (NSO/Malawi, and ICF, 2017) | 0.74 [0.68; 0.82] |
| Namibia – (MoHSS/Namibia, and ICF International, 2014) | 0.71 [0.58; 0.88] |
| Nigeria – (NPC/Nigeria, and ICF Macro, 2009) | 0.48 [0.45; 0.51] |
| Nigeria – (NPC/Nigeria, and ICF International, 2014) | 0.58 [0.54; 0.63] |
| Nigeria – (NPC/Nigeria, and ICF, 2019) | 0.60 [0.54; 0.66] |
| Rwanda – (INSR/Rwanda, and ORC Macro, 2006) | 0.58 [0.51; 0.65] |
| Rwanda – (NISR/Rwanda, MOH/Rwanda, and ICF International, 2012) | 0.95 [0.87; 1.04] |
| Rwanda – (DHS, 2015) | 0.65 [0.55; 0.78] |
| Senegal – (ANSD/Sénégal, and ICF, 2018) | 0.66 [0.54; 0.79] |
| Sierra Leone – (SSL/Sierra Leone, and ICF International, 2014) | 0.50 [0.45; 0.56] |
| Sao Tome and Principe – (INE/São Tomé, and ICF Macro, 2010) | 0.54 [0.45; 0.64] |
| Chad – (INSEED/Tchad, MSP/Tchad, and ICF International, 2016) | 0.64 [0.56; 0.73] |
| Togo – (MPDAT/Togo, MS/Togo, and ICF International, 2015) | 0.67 [0.59; 0.76] |
| Tanzania – (NBS/Tanzania, and ICF Macro, 2011) | 0.59 [0.55; 0.64] |
| Tanzania – (MoHCDGEC/Tanzania, MoH/Zanzibar, NBS/Tanzania, OCGS/Zanzibar, and ICF, 2016) | 0.65 [0.60; 0.70] |
| Uganda – (UBOS/Uganda, and Macro International, 2007) | 0.75 [0.66; 0.86] |
| Uganda – (DHS, 2011) | 0.76 [0.65; 0.88] |
| Uganda – (UBOS/Uganda, and ICF, 2018) | 0.74 [0.69; 0.80] |
| South Africa – (NDH/South Africa, and ICF, 2019) | 0.63 [0.54; 0.75] |
| Zambia – (CSO/Zambia, MOH/Zambia, TDRC/Zambia, and University of Zambia, 2009) | 0.49 [0.45; 0.54] |
| Zambia – (CSO/Zambia, MOH/Zambia, University of Zambia THVL, DPS, TDRC/Zambia, and ICF International, 2015) | 0.65 [0.61; 0.70] |
| Zimbabwe – (CSO/Zimbabwe, and Macro International, 2007) | 0.65 [0.59; 0.71] |
| Zimbabwe – (ZIMSTAT/Zimbabwe, and ICF International, 2012) | 0.65 [0.60; 0.72] |
| Zimbabwe – (ZNSA/Zimbabwe, and ICF International, 2016) | 0.64 [0.57; 0.70] |
| Total | 0.61 [0.58; 0.64] |
| Heterogeneity: $\chi^2_{48} = 399.83$ ($P < .001$), $I^2 = 88\%$ | |
| **SuperRegion = Central Europe, Eastern Europe & Central Asia** | |
| Armenia – (DHS, 2016) | 0.27 [0.18; 0.40] |
| Azerbaijan – (SSC/Azerbaijan, and Macro International, 2008) | 0.44 [0.37; 0.53] |
| Kyrgyz Republic – (NSC, MOH/Kyrgyz Republic NSC, and ICF International, 2013) | 0.41 [0.36; 0.47] |
| Moldova – (NCPM/Moldova, and ORC Macro, 2006) | 0.47 [0.42; 0.52] |
| Tajikistan – (SA/Tajikistan, MOH/Tajikistan, and ICF International, 2013) | 0.39 [0.33; 0.45] |
| Tajikistan – (SA, MOH/Tajikistan, and ICF, 2018) | 0.31 [0.27; 0.35] |
| Ukraine – (UCSR, SSC/Ukraine, MOH/Ukraine, and Macro International, 2008) | 0.46 [0.37; 0.58] |
| Total | 0.39 [0.34; 0.45] |
| Heterogeneity: $\chi^2_6 = 29.56$ ($P < .001$), $I^2 = 80\%$ | |
| **SuperRegion = South Asia** | |
| Bangladesh – (NIPORT/Bangladesh, Mitra, Associates/Bangladesh, and Macro International, 2009) | 0.60 [0.54; 0.67] |
| India – (DHS, 2006) | 0.50 [0.49; 0.52] |
| Nepal – (MOHP/Nepal, New ERA/Nepal, and ICF International, 2012) | 0.72 [0.62; 0.83] |
| Nepal – (MOH/Nepal, New ERA/Nepal, and ICF, 2017) | 0.59 [0.50; 0.69] |
| Pakistan – (NIPS/Pakistan, and ICF International, 2013) | 0.32 [0.28; 0.38] |
| Pakistan – (NIPS/Pakistan, and ICF, 2019) | 0.45 [0.38; 0.53] |
| Total | 0.52 [0.44; 0.61] |
| Heterogeneity: $\chi^2_5 = 69.72$ ($P < .001$), $I^2 = 93\%$ | |
| **SuperRegion = Latin America & Caribbean** | |
| Colombia – (DHS, 2005) | 0.50 [0.48; 0.52] |
| Colombia – (Profamilia/Colombia, 2005) | 0.46 [0.45; 0.48] |
| Colombia – (MSPSP, 2015) | 0.46 [0.44; 0.48] |
| Dominican Republic – (Achécar et al., 2003) | 0.68 [0.62; 0.74] |
| Dominican Republic – (CESDEM/Republica Dominicana and Macro International, 2008) | 0.64 [0.58; 0.72] |
| Dominican Republic – (CESDEM/Republica Dominicana, and ICF International, 2014) | 0.56 [0.50; 0.63] |
| Guatemala – (DHS, 2015) | 0.57 [0.49; 0.66] |
| Honduras – (SS/Honduras, INE/Honduras, and ICF International, 2013) | 0.61 [0.56; 0.67] |
| Haïti – (Cayemittes et al., 2001) | 0.86 [0.76; 0.97] |
| Haiti – (DHS, 2006) | 0.77 [0.66; 0.89] |
| Haïti – (Cayemittes et al., 2013) | 0.80 [0.72; 0.89] |
| Haïti – (IHE/Haïti, and ICF, 2018) | 0.77 [0.67; 0.88] |
| Total | 0.62 [0.56; 0.69] |
| Heterogeneity: $\chi^2_{11} = 332.1$ ($P < .001$), $I^2 = 97\%$ | |
| **SuperRegion = South–East Asia, East Asia & Oceania** | |
| Cambodia – (NIS/Cambodia, DGH/Cambodia, and ORC Macro, 2001) | 0.60 [0.51; 0.71] |
| Cambodia – (NIPH/Cambodia, NIS/Cambodia, and ORC Macro, 2006) | 0.64 [0.50; 0.81] |
| Cambodia – (NIS/Cambodia, DGH/Cambodia, and ICF International, 2015) | 0.67 [0.56; 0.79] |
| Maldives – (DHS, 2016) | 0.52 [0.40; 0.67] |
| Myanmar – (DHS, 2016) | 0.54 [0.45; 0.65] |
| Philippines – (NSO/Philippines, and ICF Macro, 2009) | 0.66 [0.58; 0.74] |
| Philippines – (PSA/Philippines, and ICF International, 2014) | 0.71 [0.62; 0.82] |
| Philippines – (PSA/Philippines, and ICF, 2018) | 0.64 [0.56; 0.73] |
| Papua New Guinea – (DHS, 2017) | 0.74 [0.68; 0.80] |
| Timor–leste – (NSD/Timor–Leste, MF/Timor–Leste, and ICF Macro, 2010) | 0.30 [0.26; 0.36] |
| Timor–leste – (GDS, MF/Timor Leste, and ICF, 2018) | 0.41 [0.36; 0.45] |
| Total | 0.57 [0.48; 0.67] |
| Heterogeneity: $\chi^2_{10} = 147.68$ ($P < .001$), $I^2 = 93\%$ | |
| Total | 0.57 [0.55; 0.60] |
| Heterogeneity: $\chi^2_{89} = 2056.92$ ($P < .001$), $I^2 = 96\%$ | |
| Residual heterogeneity: $\chi^2_{84} = 1192.96$ ($P < .001$), $I^2 = 93\%$ | |

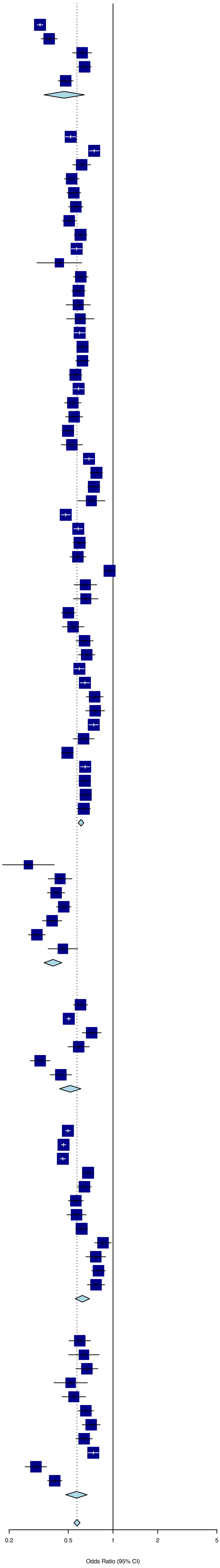

**Figure S3.** Forest plot of the random effect meta-analysis for lifetime physical intimate partner violence (IPV) only, as compared to physical and/or sexual violence.

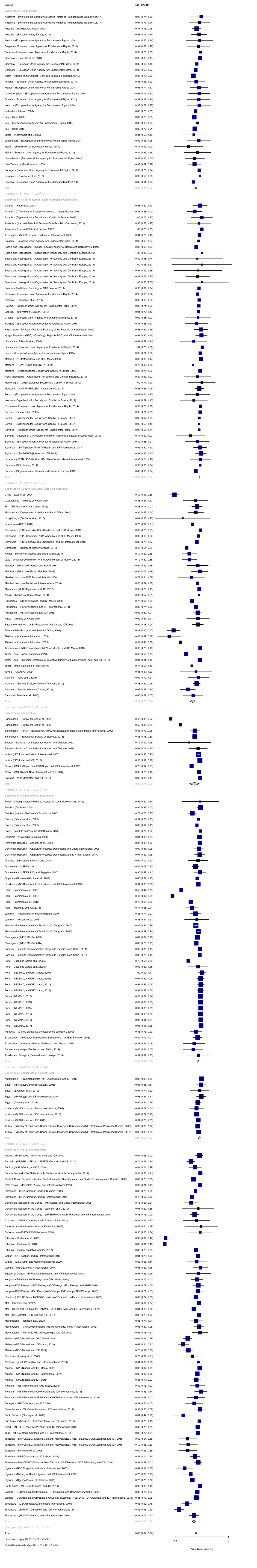
Source
OR (95% CI)
Odds Ratio (95% CI)

**Figure S4.** Forest plot of the random effect meta-analysis for past year physical intimate partner violence (IPV) only, as compared to physical and/or sexual violence.

| Source | OR (95% CI) |
|---|---|
| **SuperRegion = High Income** | |
| Argentina – (Ministerio de Justicia y Derechos Humanos Presidencia de la Nacion, 2017) | 1.00 [0.46; 2.18] |
| Argentina – (Ministerio de Justicia y Derechos Humanos Presidencia de la Nacion, 2017) | 1.00 [0.50; 1.98] |
| Australia – (Mouzos and Makai, 2004) | 0.74 [0.52; 1.06] |
| Austria – (European Union Agency for Fundamental Rights, 2014) | 0.64 [0.30; 1.38] |
| Belgium – (European Union Agency for Fundamental Rights, 2014) | 0.82 [0.49; 1.36] |
| Chile – (Ministerio del Interior y Seguridad Pública, 2013) | 0.32 [0.25; 0.40] |
| Cyprus – (European Union Agency for Fundamental Rights, 2014) | 1.00 [0.51; 1.98] |
| Germany – (European Union Agency for Fundamental Rights, 2014) | 1.00 [0.52; 1.94] |
| Denmark – (European Union Agency for Fundamental Rights, 2014) | 0.78 [0.41; 1.45] |
| Denmark – (National Institute of Public Health, 2017) | 0.79 [0.52; 1.19] |
| Spain – (Ministerio de Sanidad, Servicios Sociales e Igualdad, 2015) | 0.68 [0.50; 0.91] |
| Finland – (European Union Agency for Fundamental Rights, 2014) | 0.79 [0.46; 1.37] |
| Finland – (Institute of Criminology and Legal Policy – University of Helsinki, 2013) | 0.81 [0.59; 1.10] |
| Finland – (Institute of Criminology and Legal Policy – University of Helsinki, 2014) | 0.79 [0.56; 1.11] |
| Finland – (Institute of Criminology and Legal Policy – University of Helsinki, 2015) | 0.80 [0.58; 1.10] |
| Finland – (Institute of Criminology and Legal Policy – University of Helsinki, 2016) | 0.81 [0.57; 1.14] |
| Finland – (Institute of Criminology and Legal Policy – University of Helsinki, 2017) | 0.92 [0.69; 1.23] |
| France – (European Union Agency for Fundamental Rights, 2014) | 1.00 [0.58; 1.71] |
| United Kingdom – (European Union Agency for Fundamental Rights, 2014) | 0.79 [0.46; 1.37] |
| Greece – (European Union Agency for Fundamental Rights, 2014) | 0.82 [0.49; 1.36] |
| Ireland – (European Union Agency for Fundamental Rights, 2014) | 1.00 [0.51; 1.98] |
| Iceland – (Gíslason, 2009) | 0.63 [0.32; 1.24] |
| Italy – (Istat, 2006) | 0.70 [0.58; 0.86] |
| Italy – (European Union Agency for Fundamental Rights, 2014) | 0.82 [0.50; 1.36] |
| Italy – (Istat, 2015) | 0.80 [0.64; 0.99] |
| Japan – (Yoshihama et al., 2005) | 0.84 [0.43; 1.65] |
| Luxembourg – (European Union Agency for Fundamental Rights, 2014) | 1.00 [0.43; 2.34] |
| Malta – (European Union Agency for Fundamental Rights, 2014) | 0.72 [0.37; 1.38] |
| Netherlands – (European Union Agency for Fundamental Rights, 2014) | 0.78 [0.45; 1.37] |
| Portugal – (European Union Agency for Fundamental Rights, 2014) | 0.78 [0.45; 1.37] |
| Singapore – (Bouhours et al., 2013) | 1.00 [0.32; 3.12] |
| Sweden – (European Union Agency for Fundamental Rights, 2014) | 0.59 [0.32; 1.07] |
| Total | 0.76 [0.69; 0.83] |
| Heterogeneity: $\chi^2_{31} = 58.92$ ($P = .002$), $I^2 = 47\%$ | |
| **SuperRegion = Central Europe, Eastern Europe & Central Asia** | |
| Albania – (Haarr et al., 2013) | 0.89 [0.73; 1.09] |
| Albania – ( The Institut of Statistics of Albania ... Instat/Albania, 2018) | 0.86 [0.65; 1.15] |
| Armenia – (National Statistical Service, 2017) | 1.00 [0.66; 1.52] |
| Azerbaijan – (SSC/Azerbaijan, and Macro International, 2008) | 0.94 [0.74; 1.20] |
| Bulgaria – (European Union Agency for Fundamental Rights, 2014) | 1.00 [0.62; 1.62] |
| Belarus – (Institute of Sociology of NAS Belarus, 2019) | 0.94 [0.56; 1.55] |
| Czechia – (European Union Agency for Fundamental Rights, 2014) | 1.00 [0.57; 1.76] |
| Czechia – (...Buriánek et al., 2013) | 1.00 [0.39; 2.54] |
| Estonia – (European Union Agency for Fundamental Rights, 2014) | 1.00 [0.45; 2.24] |
| Georgia – (UN Women/GEOSTAT, 2018) | 0.89 [0.43; 1.85] |
| Croatia – (European Union Agency for Fundamental Rights, 2014) | 1.00 [0.51; 1.94] |
| Hungary – (European Union Agency for Fundamental Rights, 2014) | 0.82 [0.50; 1.36] |
| Kazakhstan – (Ministry of National Economy of the Republic of Kazakhstan, 2017) | 0.95 [0.70; 1.29] |
| Kyrgyz Republic – (NSC, MOH/Kyrgyz Republic NSC, and ICF International, 2013) | 0.99 [0.83; 1.18] |
| Lithuania – (European Union Agency for Fundamental Rights, 2014) | 1.00 [0.55; 1.80] |
| Latvia – (European Union Agency for Fundamental Rights, 2014) | 1.00 [0.59; 1.70] |
| Moldova – (NCPM/Moldova, and ORC Macro, 2006) | 0.96 [0.78; 1.18] |
| Moldova – (UNW, UNDP, and UNFPA, 2011) | 0.82 [0.23; 2.86] |
| Mongolia – (NSO, UNFPA, SDC, Australian Aid, 2018) | 0.93 [0.79; 1.09] |
| Poland – (European Union Agency for Fundamental Rights, 2014) | 1.00 [0.45; 2.24] |
| Romania – (European Union Agency for Fundamental Rights, 2014) | 1.00 [0.62; 1.61] |
| Serbia – (Otasevic et al., 2005) | 0.83 [0.41; 1.66] |
| Slovakia – (European Union Agency for Fundamental Rights, 2014) | 1.00 [0.61; 1.63] |
| Slovenia – (Institute of Criminology, Ministry of Interior and Faculty of Social Work, 2010) | 0.59 [0.21; 1.65] |
| Slovenia – (European Union Agency for Fundamental Rights, 2014) | 1.00 [0.43; 2.33] |
| Tajikistan – (SA/Tajikistan, MOH/Tajikistan, and ICF International, 2013) | 0.94 [0.78; 1.14] |
| Tajikistan – (SA, MOH/Tajikistan, and ICF, 2018) | 0.98 [0.84; 1.15] |
| Ukraine – (UCSR, SSC/Ukraine, MOH/Ukraine, and Macro International, 2008) | 0.99 [0.73; 1.34] |
| Ukraine – (GfK Ukraine, 2014) | 0.94 [0.63; 1.40] |
| Total | 0.95 [0.93; 0.97] |
| Heterogeneity: $\chi^2_{28} = 3.08$ ($P > .99$), $I^2 = 0\%$ | |
| **SuperRegion = South–East Asia, East Asia & Oceania** | |
| Cook Islands – (Ministry of Health, 2014) | 0.68 [0.38; 1.21] |
| Fiji – (Fiji Women's Crisis Centre, 2013) | 0.78 [0.64; 0.94] |
| Micronesia – (Department of Health and Social Affairs, 2014) | 0.78 [0.58; 1.06] |
| Hong Kong – (Bouhours et al., 2015) | 0.50 [0.12; 2.00] |
| Indonesia – (UNDP, 2016) | 0.90 [0.66; 1.23] |
| Cambodia – (NIS/Cambodia, DGH/Cambodia, and ORC Macro, 2001) | 0.94 [0.75; 1.19] |
| Cambodia – (NIPH/Cambodia, NIS/Cambodia, and ORC Macro, 2006) | 0.93 [0.65; 1.31] |
| Cambodia – (NIS/Cambodia, DGH/Cambodia, and ICF International, 2015) | 0.84 [0.65; 1.09] |
| Cambodia – (Ministry of Women's Affairs, 2015) | 0.59 [0.42; 0.82] |
| Kiribati – (Ministry of Internal and Social Affairs, 2010) | 0.85 [0.68; 1.06] |
| Laos – (National Commission for the Advancement of Women, 2015) | 0.64 [0.44; 0.92] |
| Maldives – (Ministry of Gender and Family, 2011) | 0.88 [0.56; 1.37] |
| Maldives – (Ministry of Health Maldives, 2018) | 0.98 [0.70; 1.39] |
| Marshall Islands – (DHS/Marshall Islands, 2008) | 0.76 [0.53; 1.09] |
| Marshall Islands – (Ministry of Internal Affairs, 2014) | 0.88 [0.60; 1.28] |
| Myanmar – (MoHS/Myanmar, and ICF, 2017) | 0.92 [0.71; 1.19] |
| Nauru – (Ministry of Home Affairs, 2014) | 0.93 [0.51; 1.71] |
| Philippines – (NSO/Philippines, and ICF Macro, 2009) | 0.71 [0.58; 0.85] |
| Philippines – (PSA/Philippines, and ICF International, 2014) | 0.75 [0.60; 0.93] |
| Philippines – (PSA/Philippines, and ICF, 2018) | 0.79 [0.65; 0.95] |
| Palau – (Ministry of Health, 2014) | 0.85 [0.48; 1.49] |
| Papua New Guinea – (NSO/Papua New Guinea, and ICF, 2019) | 0.88 [0.76; 1.02] |
| Thailand – (Kanchanachitra et al., 2005) | 0.32 [0.21; 0.49] |
| Thailand – (Kanchanachitra et al., 2005) | 0.52 [0.36; 0.75] |
| Timor–leste – (NSD/Timor–Leste, MF/Timor–Leste, and ICF Macro, 2010) | 0.96 [0.78; 1.20] |
| Timor–Leste – (Asia Foundation, 2016) | 0.64 [0.49; 0.83] |
| Timor–Leste – (General Directorate of Statistics, Ministry of Finance/Timor Leste, and ICF, 2018) | 0.94 [0.80; 1.10] |
| Tonga – (Ma'a Fafine mo e Famili, 2012) | 0.63 [0.35; 1.12] |
| Tuvalu – (CSDSPC, 2009) | 0.95 [0.60; 1.49] |
| Vietnam – (Vung et al., 2008) | 0.87 [0.55; 1.37] |
| Vietnam – (General Statistics Office of Vietnam, 2010) | 0.69 [0.59; 0.82] |
| Vanuatu – (Vanuatu Women's Centre, 2011) | 0.63 [0.51; 0.76] |
| Samoa – ( Posiulai et al., 2005) | 0.75 [0.55; 1.03] |
| Total | 0.78 [0.72; 0.84] |
| Heterogeneity: $\chi^2_{32} = 58.6$ ($P = .003$), $I^2 = 45\%$ | |
| **SuperRegion = South Asia** | |
| Bangladesh – (Garcia–Moreno et al., 2005) | 0.40 [0.30; 0.54] |
| Bangladesh – (Garcia–Moreno et al., 2005) | 0.54 [0.41; 0.71] |
| Bangladesh – (NIPORT/Bangladesh, Mitra, Associates/Bangladesh, and Macro International, 2009) | 0.72 [0.61; 0.85] |
| Bangladesh – (Bangladesh Bureau of Statistics, 2016) | 0.71 [0.66; 0.77] |
| Bhutan – (National Commission for Women and Children, 2019) | 0.83 [0.62; 1.10] |
| India – (IIPS/India, and Macro International, 2007) | 0.86 [0.83; 0.90] |
| India – (IIPS/India, and ICF, 2017) | 0.94 [0.90; 0.98] |
| Nepal – (MOHP/Nepal, New ERA/Nepal, and ICF International, 2012) | 0.72 [0.57; 0.91] |
| Nepal – (MOH/Nepal, New ERA/Nepal, and ICF, 2017) | 0.88 [0.69; 1.11] |
| Pakistan – (NIPS/Pakistan, and ICF International, 2013) | 1.00 [0.82; 1.21] |
| Pakistan – (NIPS/Pakistan, and ICF, 2019) | 0.93 [0.74; 1.16] |
| Total | 0.78 [0.66; 0.91] |
| Heterogeneity: $\chi^2_{10} = 88.43$ ($P < .001$), $I^2 = 89\%$ | |
| **SuperRegion = Latin America & Caribbean** | |
| Belize – (Young Macfarlane Belize Institute for Local Development, 2015) | 0.76 [0.32; 1.77] |
| Bolivia – (Instituto Nacional de Estadística, 2017) | 0.73 [0.64; 0.84] |
| Brazil – (Schraiber et al., 2005) | 0.88 [0.53; 1.45] |
| Brazil – (Schraiber et al., 2005) | 0.85 [0.59; 1.23] |
| Brazil – (Instituto de Pesquisa DataSenado, 2017) | 0.89 [0.49; 1.61] |
| Colombia – (Bott et al., 2015) | 0.95 [0.88; 1.02] |
| Dominican Republic – (CESDEM/Republica Dominicana and Macro International, 2008) | 0.93 [0.79; 1.08] |
| Dominican Republic – (CESDEM/Republica Dominicana, and ICF International, 2014) | 0.93 [0.79; 1.10] |
| Grenada – (Nicholson and Deshong., 2018) | 0.79 [0.48; 1.29] |
| Guatemala – (MSPAS, 2011) | 0.77 [0.67; 0.88] |
| Guatemala – (MSPAS, INE, and Segeplán, 2017) | 0.92 [0.79; 1.08] |
| Guyana – (Contreras–Urbina et al., 2019) | 0.80 [0.54; 1.19] |
| Honduras – (SS/Honduras, INE/Honduras, and Macro International, 2006) | 0.71 [0.62; 0.82] |
| Honduras – (SS/Honduras, INE/Honduras, and ICF International, 2013) | 0.90 [0.79; 1.03] |
| Haïti – (Cayemittes et al., 2001) | 0.56 [0.43; 0.72] |
| Haïti – (Cayemittes et al., 2007) | 0.64 [0.50; 0.83] |
| Haïti – (Cayemittes et al., 2013) | 0.66 [0.56; 0.79] |
| Haïti – (IHE/Haïti, and ICF, 2018) | 0.70 [0.56; 0.87] |
| Jamaica – (National Family Planning Board, 2010) | 0.84 [0.68; 1.02] |
| Jamaica – (Williams et al., 2018) | 0.84 [0.47; 1.50] |
| Mexico – (Instituto National de Estadística Y Geografía, 2004) | 0.63 [0.59; 0.68] |
| Mexico – (Instituto National de Estadística Y Geografía, 2007) | 0.74 [0.71; 0.78] |
| Mexico – (Instituto National de Estadística Y Geografía, 2016) | 0.88 [0.85; 0.92] |
| Nicaragua – (INIDE MINSA, 2008) | 0.85 [0.73; 0.98] |
| Nicaragua – (INIDE MINSA, 2014) | 0.80 [0.68; 0.94] |
| Panama – (Instituto Conmemorativo Gorgas de Estudios de la Salud, 2011) | 0.90 [0.74; 1.10] |
| Peru – (Guezmes Garcia et al., 2005) | 0.86 [0.60; 1.21] |
| Peru – (Guezmes Garcia et al., 2005) | 0.63 [0.49; 0.81] |
| Peru – (INEI/Perú, and ORC Macro, 2007) | 1.00 [0.87; 1.16] |
| Peru – (INEI/Perú, and ORC Macro, 2009) | 0.92 [0.82; 1.04] |
| Peru – (INEI/Perú, and ORC Macro, 2010) | 0.94 [0.83; 1.05] |
| Peru – (INEI/Perú, and ORC Macro, 2011) | 0.92 [0.81; 1.04] |
| Peru – (INEI/Perú, 2012) | 0.92 [0.82; 1.03] |
| Peru – (INEI/Perú, 2013) | 0.93 [0.83; 1.04] |
| Peru – (INEI/Perú, 2014) | 0.94 [0.83; 1.07] |
| Peru – (INEI/Perú, 2015) | 0.92 [0.82; 1.03] |
| Peru – (INEI/Perú, 2016) | 0.92 [0.84; 1.02] |
| Peru – (INEI/Perú, 2017) | 0.94 [0.84; 1.04] |
| Peru – (INEI/Perú, 2018) | 0.94 [0.85; 1.03] |
| El salvador – (Asociación Demográfica Salvadoreña – ADS/El Salvador, 2008) | 0.88 [0.72; 1.07] |
| El salvador – (Navarros–Mantas, Velásquez, and Megías, 2015) | 0.71 [0.43; 1.19] |
| Suriname – (Joseph, Pemberton and Phillip, 2019) | 0.71 [0.40; 1.27] |
| Trinidad and Tobago – (Pemberton and Joseph, 2018) | 0.91 [0.49; 1.67] |
| Total | 0.84 [0.80; 0.88] |
| Heterogeneity: $\chi^2_{42} = 180.22$ ($P < .001$), $I^2 = 77\%$ | |
| **SuperRegion = North Africa & Middle East** | |
| Afghanistan – (CSO/Afghanistan, MPH/Afghanistan, and ICF, 2017) | 0.99 [0.93; 1.05] |
| Egypt – (MHP/Egypt and ICF International, 2015) | 0.96 [0.82; 1.12] |
| Egypt – (Duvuury et al., 2015) | 0.82 [0.75; 0.90] |
| Jordan – (DoS/Jordan, and Macro International, 2008) | 0.81 [0.65; 1.01] |
| Jordan – (DoS/Jordan, and ICF International, 2013) | 0.77 [0.66; 0.90] |
| Jordan – (DoS/Jordan, and ICF, 2019) | 0.91 [0.78; 1.06] |
| Turkey – (Ministry of Family and Social Policies, Hacettepe University and NEE Institute of Population Studies, 2009) | 0.69 [0.61; 0.79] |
| Turkey – (Ministry of Family and Social Policies, Hacettepe University and NEE Institute of Population Studies, 2015) | 0.72 [0.60; 0.87] |
| Total | 0.83 [0.75; 0.93] |
| Heterogeneity: $\chi^2_{7} = 37.58$ ($P < .001$), $I^2 = 81\%$ | |
| **SuperRegion = Sub–Saharan Africa** | |
| Angola – (INE/Angola, MINSA/Angola, and ICF, 2017) | 0.92 [0.83; 1.02] |
| Burundi – (MPBGP, MSPLS,– ISTEEBU/Burundi, and ICF, 2017) | 0.56 [0.49; 0.64] |
| Benin – (INSAE/Bénin, and ICF, 2019) | 0.78 [0.64; 0.96] |
| Burkina faso – (Institut National de la Statistique et de la Démographie, 2012) | 0.67 [0.58; 0.78] |
| Central African Republic – (Institut Centrafricain des Statistiques, et des Etudes Economiques et Sociales, 2009) | 0.82 [0.74; 0.91] |
| Côte d'Ivoire – (INS/Côte d'Ivoire, and ICF International, 2013) | 0.96 [0.82; 1.13] |
| Cameroon – (INS/Cameroun, and ORC Macro, 2005) | 0.79 [0.64; 0.98] |
| Cameroon – (INS/Cameroun, and ICF International, 2012) | 0.84 [0.72; 0.99] |
| Cameroon – (MPH/Cameroon, NIS/Cameroon, UNICEF, 2014) | 0.92 [0.81; 1.04] |
| Democratic Republic of the Congo – (MP/Congo, and Macro International, 2008) | 0.77 [0.64; 0.91] |
| Democratic Republic of the Congo – (MPSMRM/Congo, MSP/Congo, and ICF International, 2014) | 0.75 [0.66; 0.85] |
| Comoros – (DGSP/Comores, and ICF International, 2014) | 0.88 [0.56; 1.37] |
| Cabo verde – (ICIEG, INE/Cabo Verde, 2016) | 0.94 [0.66; 1.33] |
| Ethiopia – (Berhane et al., 2005) | 0.35 [0.29; 0.43] |
| Ethiopia – (Deribe et al., 2012) | 0.57 [0.41; 0.78] |
| Ethiopia – (Central Statistical Agency, 2017) | 0.81 [0.68; 0.97] |
| Gabon – (DGS/Gabon, and ICF International, 2013) | 0.87 [0.74; 1.02] |
| Ghana – (GSS, GHS, and Macro International, 2009) | 0.88 [0.65; 1.19] |
| Gambia – (GBOS, and ICF International, 2014) | 0.93 [0.69; 1.26] |
| Equatorial Guinea – (PIP/Guinea Ecuatorial, and ICF International, 2012) | 0.88 [0.64; 1.21] |
| Kenya – (CBS/Kenya, MOH/Kenya, and ORC Macro, 2004) | 0.82 [0.70; 0.96] |
| Kenya – (KNBS/Kenya, NACC/Kenya, NASCP/Kenya, MPHS/Kenya, and KMRI, 2010) | 0.88 [0.77; 1.02] |
| Kenya – (KNBS/Kenya, MOH/Kenya, NACC/Kenya, KMRI/Kenya, NCPD/Kenya, 2015) | 0.86 [0.73; 1.01] |
| Liberia – (LISGIS/Liberia, MOHSW/Liberia, NACP/Liberia, and Macro International, 2008) | 0.87 [0.75; 1.01] |
| Mali – (Samaké et al., 2007) | 0.92 [0.82; 1.03] |
| Mali – (CPS/SSDSPF/Mali, INSTAT/Mali, INFO–STAT/Mali, and ICF International, 2014) | 0.72 [0.60; 0.88] |
| Mali – (INSTAT/Mali, CPSSSD, and ICF, 2019) | 0.84 [0.69; 1.02] |
| Moçambique – (MISAU/Moçambique, INE/Moçambique, and ICF International, 2013) | 0.91 [0.80; 1.04] |
| Moçambique – (INS, INE, PNCM/Moçambique and ICF, 2019) | 0.94 [0.74; 1.19] |
| Malawi – (NSO/Malawi, and ORC Macro, 2005) | 0.62 [0.54; 0.71] |
| Malawi – (NSO/Malawi, and ICF Macro, 2011) | 0.56 [0.31; 1.02] |
| Malawi – (NSO/Malawi, and ICF Macro, 2011) | 0.66 [0.37; 1.17] |
| Malawi – (NSO/Malawi, and ICF Macro, 2011) | 0.65 [0.47; 0.88] |
| Malawi – (NSO/Malawi, and ICF Macro, 2011) | 0.60 [0.38; 0.96] |
| Malawi – (NSO/Malawi, and ICF Macro, 2011) | 0.57 [0.38; 0.86] |
| Malawi – (NSO/Malawi, and ICF Macro, 2011) | 0.67 [0.48; 0.94] |
| Malawi – (NSO/Malawi, and ICF Macro, 2011) | 0.61 [0.33; 1.10] |
| Malawi – (NSO/Malawi, and ICF, 2017) | 0.61 [0.52; 0.71] |
| Namibia – (January et al., 2005) | 0.78 [0.57; 1.06] |
| Namibia – (MoHSS/Namibia, and ICF International, 2014) | 0.91 [0.63; 1.29] |
| Nigeria – (NPC/Nigeria, and ICF Macro, 2009) | 0.95 [0.75; 1.22] |
| Nigeria – (NPC/Nigeria, and ICF Macro, 2009) | 0.94 [0.71; 1.25] |
| Nigeria – (NPC/Nigeria, and ICF Macro, 2009) | 0.93 [0.68; 1.29] |
| Nigeria – (NPC/Nigeria, and ICF Macro, 2009) | 0.92 [0.74; 1.16] |
| Nigeria – (NPC/Nigeria, and ICF Macro, 2009) | 0.89 [0.58; 1.36] |
| Nigeria – (NPC/Nigeria, and ICF Macro, 2009) | 0.92 [0.72; 1.17] |
| Nigeria – (NPC/Nigeria, and ICF Macro, 2009) | 0.93 [0.77; 1.13] |
| Nigeria – (NPC/Nigeria, and ICF International, 2014) | 0.83 [0.75; 0.92] |
| Nigeria – (NPC/Nigeria, and ICF, 2019) | 0.84 [0.73; 0.97] |
| Rwanda – (INSR/Rwanda, and ORC Macro, 2006) | 0.82 [0.66; 1.01] |
| Rwanda – (NISR/Rwanda, MOH/Rwanda, and ICF International, 2012) | 0.97 [0.82; 1.14] |
| Rwanda – (NISR/Rwanda, MFEP/Rwanda, MOH/Rwanda, and ICF International, 2016) | 0.82 [0.62; 1.07] |
| Senegal – (ANSD/Sénégal, and ICF, 2018) | 0.70 [0.52; 0.95] |
| Sierra Leone – (SSL/Sierra Leone, and ICF International, 2014) | 0.93 [0.80; 1.09] |
| South Sudan – (Ellsberg et al., 2018) | 0.64 [0.45; 0.91] |
| Sao Tome and Principe – (INE/São Tomé, and ICF Macro, 2010) | 0.96 [0.75; 1.23] |
| Chad – (INSEED/Tchad, MSP/Tchad, and ICF International, 2016) | 0.87 [0.71; 1.06] |
| Togo – (MPDAT/Togo, MS/Togo, and ICF International, 2015) | 0.83 [0.69; 1.01] |
| Tanzania – (MoHCDGEC/Tanzania Mainland, MoH/Zanzibar, NBS/Tanzania, OCGS/Zanzibar, and ICF, 2016) | 0.56 [0.42; 0.75] |
| Tanzania – (MoHCDGEC/Tanzania Mainland, MoH/Zanzibar, NBS/Tanzania, OCGS/Zanzibar, and ICF, 2016) | 0.63 [0.47; 0.86] |
| Tanzania – (Mccloskey et al., 2005) | 0.72 [0.57; 0.91] |
| Tanzania – (NBS/Tanzania, and ICF Macro, 2011) | 0.87 [0.77; 0.99] |
| Tanzania – (MoHCDGEC/Tanzania, MoH/Zanzibar, NBS/Tanzania, OCGS/Zanzibar, and ICF, 2016) | 0.88 [0.79; 0.99] |
| Uganda – (UBOS/Uganda, and Macro International, 2007) | 0.67 [0.53; 0.84] |
| Uganda – (Ministry of Health/Uganda, and ICF International, 2012) | 0.64 [0.50; 0.82] |
| Uganda – (Uganda Bureau of Statistics, 2018) | 0.68 [0.60; 0.77] |
| South Africa – (NDH/South Africa, and ICF, 2019) | 0.89 [0.72; 1.09] |
| Zambia – (CSO/Zambia, MOH/Zambia, TDRC/Zambia, and University of Zambia, 2009) | 0.87 [0.75; 1.00] |
| Zambia – (CSO/Zambia, MOH/Zambia, University of Zambia THVL, DPS, TDRC/Zambia, and ICF International, 2015) | 0.75 [0.67; 0.84] |
| Zimbabwe – (CSO/Zimbabwe, and Macro International, 2007) | 0.78 [0.68; 0.91] |
| Zimbabwe – (ZIMSTAT/Zimbabwe, and ICF International, 2012) | 0.66 [0.40; 1.08] |
| Zimbabwe – (ZIMSTAT/Zimbabwe, and ICF International, 2012) | 0.72 [0.54; 0.98] |
| Zimbabwe – (ZIMSTAT/Zimbabwe, and ICF International, 2012) | 0.68 [0.47; 0.99] |
| Zimbabwe – (ZIMSTAT/Zimbabwe, and ICF International, 2012) | 0.74 [0.39; 1.38] |
| Zimbabwe – (ZIMSTAT/Zimbabwe, and ICF International, 2012) | 0.70 [0.41; 1.19] |
| Zimbabwe – (ZIMSTAT/Zimbabwe, and ICF International, 2012) | 0.70 [0.46; 1.08] |
| Zimbabwe – (ZIMSTAT/Zimbabwe, and ICF International, 2012) | 0.68 [0.50; 0.91] |
| Zimbabwe – (ZNSA/Zimbabwe, and ICF International, 2016) | 0.73 [0.62; 0.85] |
| Total | 0.79 [0.76; 0.82] |
| Heterogeneity: $\chi^2_{77} = 236.41$ ($P < .001$), $I^2 = 67\%$ | |
| Total | 0.81 [0.79; 0.83] |
| Heterogeneity: $\chi^2_{233} = 732.30$ ($P < .001$), $I^2 = 68\%$ | |
| Residual heterogeneity: $\chi^2_{227} = 663.26$ ($P < .001$), $I^2 = 66\%$ | |

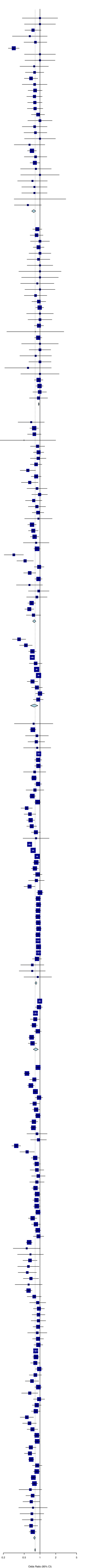

**Figure S5.** Forest plot of the random effect meta-analysis for lifetime sexual intimate partner violence (IPV) only, as compared to physical and/or sexual violence.

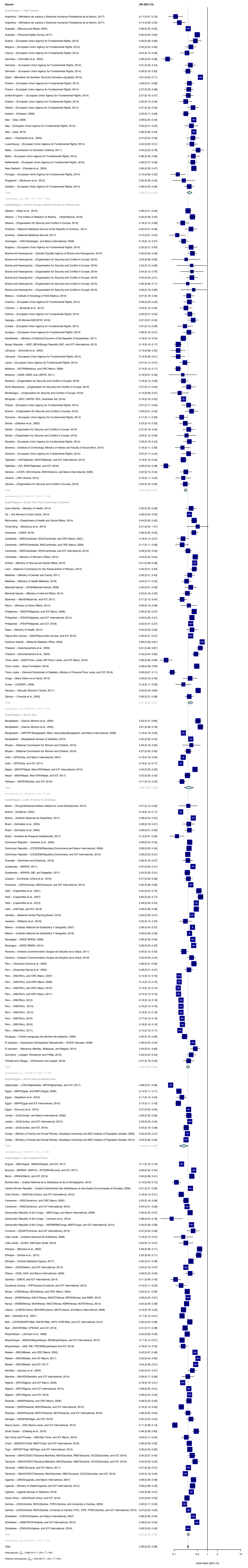
Source
OR (95% CI)
SuperRegion = High Income
Argentina – (Ministerio de Justicia y Derechos Humanos Presidencia de la Nacion, 2017) 0.11 [0.07; 0.18]
Argentina – (Ministerio de Justicia y Derechos Humanos Presidencia de la Nacion, 2017) 0.14 [0.09; 0.22]
Australia – (Mouzos and Makai, 2004) 0.26 [0.23; 0.30]
Australia – (Personal Safety Survey, 2017) 0.50 [0.40; 0.62]
Austria – (European Union Agency for Fundamental Rights, 2014) 0.43 [0.28; 0.66]
Belgium – (European Union Agency for Fundamental Rights, 2014) 0.32 [0.22; 0.46]
Cyprus – (European Union Agency for Fundamental Rights, 2014) 0.24 [0.15; 0.39]
Germany – (Schrottle et al., 2004) 0.06 [0.04; 0.09]
Germany – (European Union Agency for Fundamental Rights, 2014) 0.31 [0.22; 0.44]
Denmark – (European Union Agency for Fundamental Rights, 2014) 0.26 [0.19; 0.35]
Spain – (Ministerio de Sanidad, Servicios Sociales e Igualdad, 2015) 0.61 [0.53; 0.71]
Finland – (European Union Agency for Fundamental Rights, 2014) 0.29 [0.21; 0.39]
France – (European Union Agency for Fundamental Rights, 2014) 0.27 [0.20; 0.38]
United Kingdom – (European Union Agency for Fundamental Rights, 2014) 0.27 [0.19; 0.37]
Greece – (European Union Agency for Fundamental Rights, 2014) 0.22 [0.15; 0.34]
Ireland – (European Union Agency for Fundamental Rights, 2014) 0.37 [0.25; 0.55]
Iceland – (Gíslason, 2009) 0.22 [0.17; 0.29]
Italy – (Istat, 2006) 0.39 [0.35; 0.43]
Italy – (European Union Agency for Fundamental Rights, 2014) 0.33 [0.21; 0.52]
Italy – (Istat, 2015) 0.39 [0.35; 0.43]
Japan – (Yoshihama et al., 2005) 0.37 [0.24; 0.56]
Luxembourg – (European Union Agency for Fundamental Rights, 2014) 0.34 [0.23; 0.51]
Malta – (Commission on Domestic Violence, 2011) 0.52 [0.35; 0.78]
Malta – (European Union Agency for Fundamental Rights, 2014) 0.38 [0.25; 0.58]
Netherlands – (European Union Agency for Fundamental Rights, 2014) 0.36 [0.27; 0.49]
New Zealand – (Fanslow et al., 2004) 0.39 [0.33; 0.47]
Portugal – (European Union Agency for Fundamental Rights, 2014) 0.13 [0.08; 0.22]
Singapore – (Bouhours et al., 2013) 0.20 [0.09; 0.42]
Sweden – (European Union Agency for Fundamental Rights, 2014) 0.28 [0.20; 0.39]
Total 0.29 [0.24; 0.35]
Heterogeneity: $\chi^2_{28} = 266.11$ ($P < .001$), $I^2 = 89\%$
SuperRegion = Central Europe, Eastern Europe & Central Asia
Albania – (Haarr et al., 2013) 0.26 [0.21; 0.33]
Albania – ( The Institut of Statistics of Albania ... Instat/Albania, 2018) 0.35 [0.28; 0.45]
Albania – (Organisation for Security and Conflict in Europe, 2019) 0.18 [0.12; 0.28]
Armenia – (National Statistical Service of the Republic of Armenia , 2011) 0.32 [0.21; 0.49]
Armenia – (National Statistical Service, 2017) 0.12 [0.07; 0.22]
Azerbaijan – (SSC/Azerbaijan, and Macro International, 2008) 0.19 [0.14; 0.27]
Bulgaria – (European Union Agency for Fundamental Rights, 2014) 0.32 [0.21; 0.50]
Bosnia and Hezergovina – (Gender Equality Agency of Bosnia and Herzegovina, 2013) 0.35 [0.26; 0.48]
Bosnia and Hezergovina – (Organisation for Security and Conflict in Europe, 2019) 0.23 [0.08; 0.62]
Bosnia and Hezergovina – (Organisation for Security and Conflict in Europe, 2019) 0.34 [0.12; 0.96]
Bosnia and Hezergovina – (Organisation for Security and Conflict in Europe, 2019) 0.34 [0.15; 0.76]
Bosnia and Hezergovina – (Organisation for Security and Conflict in Europe, 2019) 0.34 [0.23; 0.51]
Bosnia and Hezergovina – (Organisation for Security and Conflict in Europe, 2019) 0.26 [0.09; 0.71]
Bosnia and Hezergovina – (Organisation for Security and Conflict in Europe, 2019) 0.39 [0.16; 0.96]
Belarus – (Institute of Sociology of NAS Belarus, 2019) 0.27 [0.19; 0.40]
Czechia – (European Union Agency for Fundamental Rights, 2014) 0.29 [0.20; 0.42]
Czechia – (...Buriánek et al., 2013) 0.23 [0.15; 0.35]
Estonia – (European Union Agency for Fundamental Rights, 2014) 0.30 [0.21; 0.42]
Georgia – (UN Women/GEOSTAT, 2018) 0.37 [0.27; 0.53]
Croatia – (European Union Agency for Fundamental Rights, 2014) 0.21 [0.13; 0.36]
Hungary – (European Union Agency for Fundamental Rights, 2014) 0.28 [0.19; 0.41]
Kazakhstan – (Ministry of National Economy of the Republic of Kazakhstan, 2017) 0.19 [0.15; 0.24]
Kyrgyz Republic – (NSC, MOH/Kyrgyz Republic NSC, and ICF International, 2013) 0.13 [0.10; 0.17]
Lithuania – (Schrottle et al., 2003) 0.13 [0.08; 0.22]
Lithuania – (European Union Agency for Fundamental Rights, 2014) 0.13 [0.08; 0.21]
Latvia – (European Union Agency for Fundamental Rights, 2014) 0.21 [0.14; 0.31]
Moldova – (NCPM/Moldova, and ORC Macro, 2006) 0.13 [0.10; 0.17]
Moldova – (UNW, UNDP, and UNFPA, 2011) 0.18 [0.07; 0.48]
Moldova – (Organisation for Security and Conflict in Europe, 2019) 0.19 [0.14; 0.26]
North Macedonia – (Organisation for Security and Conflict in Europe, 2019) 0.27 [0.17; 0.45]
Montenegro – (Organisation for Security and Conflict in Europe, 2019) 0.15 [0.08; 0.27]
Mongolia – (NSO, UNFPA, SDC, Australian Aid, 2018) 0.19 [0.16; 0.22]
Poland – (European Union Agency for Fundamental Rights, 2014) 0.27 [0.17; 0.43]
Kosovo – (Organisation for Security and Conflict in Europe, 2019) 0.33 [0.21; 0.53]
Romania – (European Union Agency for Fundamental Rights, 2014) 0.17 [0.11; 0.26]
Serbia – (Otasevic et al., 2005) 0.22 [0.14; 0.33]
Serbia – (Organisation for Security and Conflict in Europe, 2019) 0.27 [0.18; 0.43]
Serbia – (Organisation for Security and Conflict in Europe, 2019) 0.24 [0.12; 0.46]
Slovakia – (European Union Agency for Fundamental Rights, 2014) 0.29 [0.19; 0.43]
Slovenia – (Institute of Criminology, Ministry of Interior and Faculty of Social Work, 2010) 0.18 [0.11; 0.30]
Slovenia – (European Union Agency for Fundamental Rights, 2014) 0.27 [0.17; 0.43]
Tajikistan – (SA/Tajikistan, MOH/Tajikistan, and ICF International, 2013) 0.19 [0.15; 0.24]
Tajikistan – (SA, MOH/Tajikistan, and ICF, 2018) 0.06 [0.04; 0.08]
Ukraine – (UCSR, SSC/Ukraine, MOH/Ukraine, and Macro International, 2008) 0.22 [0.15; 0.34]
Ukraine – (GfK Ukraine, 2014) 0.19 [0.11; 0.32]
Ukraine – (Organisation for Security and Conflict in Europe, 2019) 0.22 [0.16; 0.30]
Total 0.22 [0.20; 0.25]
Heterogeneity: $\chi^2_{45} = 189.92$ ($P < .001$), $I^2 = 76\%$
SuperRegion = South–East Asia, East Asia & Oceania
Cook Islands – (Ministry of Health, 2014) 0.32 [0.22; 0.48]
Fiji – (Fiji Women's Crisis Centre, 2013) 0.29 [0.25; 0.33]
Micronesia – (Department of Health and Social Affairs, 2014) 0.40 [0.32; 0.50]
Hong Kong – (Bouhours et al., 2015) 0.51 [0.24; 1.07]
Indonesia – (UNDP, 2016) 0.39 [0.30; 0.52]
Cambodia – (NIS/Cambodia, DGH/Cambodia, and ORC Macro, 2001) 0.18 [0.12; 0.27]
Cambodia – (NIPH/Cambodia, NIS/Cambodia, and ORC Macro, 2006) 0.17 [0.11; 0.28]
Cambodia – (NIS/Cambodia, DGH/Cambodia, and ICF International, 2015) 0.26 [0.20; 0.34]
Cambodia – (Ministry of Women's Affairs, 2015) 0.43 [0.34; 0.54]
Kiribati – (Ministry of Internal and Social Affairs, 2010) 0.41 [0.36; 0.48]
Laos – (National Commission for the Advancement of Women, 2015) 0.40 [0.31; 0.53]
Maldives – (Ministry of Gender and Family, 2011) 0.29 [0.21; 0.42]
Maldives – (Ministry of Health Maldives, 2018) 0.24 [0.17; 0.32]
Marshall Islands – (DHS/Marshall Islands, 2008) 0.32 [0.21; 0.48]
Marshall Islands – (Ministry of Internal Affairs, 2014) 0.25 [0.18; 0.35]
Myanmar – (MoHS/Myanmar, and ICF, 2017) 0.17 [0.12; 0.24]
Nauru – (Ministry of Home Affairs, 2014) 0.28 [0.16; 0.48]
Philippines – (NSO/Philippines, and ICF Macro, 2009) 0.40 [0.34; 0.47]
Philippines – (PSA/Philippines, and ICF International, 2014) 0.34 [0.29; 0.41]
Philippines – (PSA/Philippines, and ICF, 2018) 0.32 [0.27; 0.37]
Palau – (Ministry of Health, 2014) 0.34 [0.23; 0.50]
Papua New Guinea – (NSO/Papua New Guinea, and ICF, 2019) 0.32 [0.27; 0.37]
Solomon Islands – (National Statistics Office, 2009) 0.69 [0.59; 0.81]
Thailand – (Kanchanachitra et al., 2005) 0.61 [0.46; 0.81]
Thailand – (Kanchanachitra et al., 2005) 0.45 [0.34; 0.60]
Timor–leste – (NSD/Timor–Leste, MF/Timor–Leste, and ICF Macro, 2010) 0.06 [0.04; 0.09]
Timor–Leste – (Asia Foundation, 2016) 0.48 [0.38; 0.60]
Timor–Leste – (General Directorate of Statistics, Ministry of Finance/Timor Leste, and ICF, 2018) 0.09 [0.07; 0.11]
Tonga – (Ma'a Fafine mo e Famili, 2012) 0.30 [0.18; 0.49]
Tuvalu – (CSDSPC, 2009) 0.19 [0.11; 0.33]
Vanuatu – (Vanuatu Women's Centre, 2011) 0.53 [0.43; 0.64]
Samoa – ( Posiulai et al., 2005) 0.28 [0.21; 0.38]
Total 0.31 [0.25; 0.37]
Heterogeneity: $\chi^2_{31} = 368.98$ ($P < .001$), $I^2 = 92\%$
SuperRegion = South Asia
Bangladesh – (Garcia–Moreno et al., 2005) 0.52 [0.41; 0.66]
Bangladesh – (Garcia–Moreno et al., 2005) 0.61 [0.48; 0.78]
Bangladesh – (NIPORT/Bangladesh, Mitra, Associates/Bangladesh, and Macro International, 2009) 0.19 [0.16; 0.22]
Bangladesh – (Bangladesh Bureau of Statistics, 2016) 0.32 [0.30; 0.34]
Bhutan – (National Commission for Women and Children, 2012) 0.33 [0.18; 0.62]
Bhutan – (National Commission for Women and Children, 2019) 0.27 [0.20; 0.35]
India – (IIPS/India, and Macro International, 2007) 0.19 [0.18; 0.20]
India – (IIPS/India, and ICF, 2017) 0.16 [0.15; 0.17]
Nepal – (MOHP/Nepal, New ERA/Nepal, and ICF International, 2012) 0.42 [0.35; 0.52]
Nepal – (MOH/Nepal, New ERA/Nepal, and ICF, 2017) 0.25 [0.20; 0.32]
Pakistan – (NIPS/Pakistan, and ICF, 2019) 0.17 [0.13; 0.23]
Total 0.28 [0.20; 0.38]
Heterogeneity: $\chi^2_{10} = 453.83$ ($P < .001$), $I^2 = 98\%$
SuperRegion = Latin America & Caribbean
Belize – (Young Macfarlane Belize Institute for Local Development, 2015) 0.27 [0.14; 0.50]
Bolivia – (Gutiérrez, 2004) 0.16 [0.14; 0.17]
Bolivia – (Instituto Nacional de Estadística, 2017) 0.38 [0.34; 0.42]
Brazil – (Schraiber et al., 2005) 0.28 [0.18; 0.41]
Brazil – (Schraiber et al., 2005) 0.29 [0.21; 0.39]
Brazil – (Instituto de Pesquisa DataSenado, 2017) 0.12 [0.07; 0.20]
Dominican Republic – (Achécar et al., 2003) 0.29 [0.24; 0.35]
Dominican Republic – (CESDEM/Republica Dominicana and Macro International, 2008) 0.28 [0.23; 0.34]
Dominican Republic – (CESDEM/Republica Dominicana, and ICF International, 2014) 0.26 [0.22; 0.31]
Grenada – (Nicholson and Deshong., 2018) 0.26 [0.18; 0.37]
Guatemala – (MSPAS, 2011) 0.37 [0.33; 0.41]
Guatemala – (MSPAS, INE, and Segeplán, 2017) 0.25 [0.20; 0.31]
Guyana – (Contreras–Urbina et al., 2019) 0.27 [0.20; 0.36]
Honduras – (SS/Honduras, INE/Honduras, and ICF International, 2013) 0.32 [0.28; 0.36]
Haïti – (Cayemittes et al., 2001) 0.55 [0.44; 0.70]
Haïti – (Cayemittes et al., 2007) 0.62 [0.50; 0.77]
Haïti – (Cayemittes et al., 2013) 0.46 [0.40; 0.54]
Haïti – (IHE/Haïti, and ICF, 2018) 0.46 [0.39; 0.56]
Jamaica – (National Family Planning Board, 2010) 0.34 [0.29; 0.41]
Jamaica – (Williams et al., 2018) 0.22 [0.14; 0.34]
Mexico – (Instituto National de Estadística Y Geografía, 2007) 0.36 [0.34; 0.37]
Mexico – (Instituto National de Estadística Y Geografía, 2016) 0.29 [0.28; 0.30]
Nicaragua – (INIDE MINSA, 2008) 0.36 [0.33; 0.40]
Nicaragua – (INIDE MINSA, 2014) 0.39 [0.34; 0.43]
Panama – (Instituto Conmemorativo Gorgas de Estudios de la Salud, 2011) 0.20 [0.15; 0.25]
Panama – (Instituto Conmemorativo Gorgas de Estudios de la Salud, 2018) 0.33 [0.25; 0.44]
Peru – (Guezmes Garcia et al., 2005) 0.39 [0.31; 0.50]
Peru – (Guezmes Garcia et al., 2005) 0.28 [0.21; 0.37]
Peru – (INEI/Perú, and ORC Macro, 2007) 0.14 [0.12; 0.16]
Peru – (INEI/Perú, and ORC Macro, 2009) 0.14 [0.12; 0.16]
Peru – (INEI/Perú, and ORC Macro, 2010) 0.13 [0.12; 0.15]
Peru – (INEI/Perú, and ORC Macro, 2011) 0.14 [0.12; 0.16]
Peru – (INEI/Perú, 2012) 0.16 [0.14; 0.18]
Peru – (INEI/Perú, 2013) 0.16 [0.14; 0.18]
Peru – (INEI/Perú, 2014) 0.16 [0.14; 0.18]
Peru – (INEI/Perú, 2015) 0.17 [0.15; 0.19]
Peru – (INEI/Perú, 2016) 0.18 [0.16; 0.19]
Peru – (INEI/Perú, 2017) 0.15 [0.13; 0.17]
Paraguay – (Centro paraguayo de estudios de población, 2009) 0.20 [0.16; 0.26]
El salvador – (Asociación Demográfica Salvadoreña – ADS/El Salvador, 2008) 0.36 [0.32; 0.42]
El salvador – (Navarros–Mantas, Velásquez, and Megías, 2015) 0.43 [0.31; 0.60]
Suriname – (Joseph, Pemberton and Phillip, 2019) 0.33 [0.24; 0.44]
Trinidad and Tobago – (Pemberton and Joseph, 2018) 0.27 [0.19; 0.40]
Total 0.26 [0.23; 0.30]
Heterogeneity: $\chi^2_{42} = 1553.85$ ($P < .001$), $I^2 = 97\%$
SuperRegion = North Africa & Middle East
Afghanistan – (CSO/Afghanistan, MPH/Afghanistan, and ICF, 2017) 0.08 [0.07; 0.09]
Egypt – (MHP/Egypt, and NNPC/Egypt, 2006) 0.14 [0.11; 0.17]
Egypt – (Seedhom et al., 2010) 0.17 [0.12; 0.24]
Egypt – (MHP/Egypt and ICF International, 2015) 0.13 [0.11; 0.16]
Egypt – (Duvuury et al., 2015) 0.27 [0.25; 0.30]
Jordan – (DoS/Jordan, and Macro International, 2008) 0.28 [0.22; 0.35]
Jordan – (DoS/Jordan, and ICF International, 2013) 0.29 [0.25; 0.34]
Jordan – (DoS/Jordan, and ICF, 2019) 0.23 [0.19; 0.28]
Turkey – (Ministry of Family and Social Policies, Hacettepe University and NEE Institute of Population Studies, 2009) 0.25 [0.23; 0.27]
Turkey – (Ministry of Family and Social Policies, Hacettepe University and NEE Institute of Population Studies, 2015) 0.23 [0.20; 0.26]
Total 0.19 [0.14; 0.26]
Heterogeneity: $\chi^2_{9} = 539.03$ ($P < .001$), $I^2 = 98\%$
SuperRegion = Sub–Saharan Africa
Angola – (INE/Angola, MINSA/Angola, and ICF, 2017) 0.17 [0.15; 0.19]
Burundi – (MPBGP, MSPLS,– ISTEEBU/Burundi, and ICF, 2017) 0.39 [0.35; 0.44]
Benin – (INSAE/Bénin, and ICF, 2019) 0.34 [0.28; 0.41]
Burkina faso – (Institut National de la Statistique et de la Démographie, 2012) 0.12 [0.09; 0.15]
Central African Republic – (Institut Centrafricain des Statistiques, et des Etudes Economiques et Sociales, 2009) 0.31 [0.27; 0.35]
Côte d'Ivoire – (INS/Côte d'Ivoire, and ICF International, 2013) 0.16 [0.13; 0.21]
Cameroon – (INS/Cameroun, and ORC Macro, 2005) 0.23 [0.18; 0.28]
Cameroon – (INS/Cameroun, and ICF International, 2012) 0.24 [0.21; 0.29]
Democratic Republic of the Congo – (MP/Congo, and Macro International, 2008) 0.30 [0.25; 0.37]
Democratic Republic of the Congo – (Johnson et al., 2010) 0.08 [0.04; 0.18]
Democratic Republic of the Congo – (MPSMRM/Congo, MSP/Congo, and ICF International, 2014) 0.33 [0.29; 0.38]
Comoros – (DGSP/Comores, and ICF International, 2014) 0.37 [0.24; 0.58]
Cabo verde – (Instituto Nacional de Estatística, 2008) 0.19 [0.10; 0.37]
Cabo verde – (ICIEG, INE/Cabo Verde, 2016) 0.22 [0.14; 0.37]
Ethiopia – (Berhane et al., 2005) 0.58 [0.48; 0.71]
Ethiopia – (Deribe et al., 2012) 0.55 [0.42; 0.71]
Ethiopia – (Central Statistical Agency, 2017) 0.32 [0.27; 0.38]
Gabon – (DGS/Gabon, and ICF International, 2013) 0.22 [0.19; 0.27]
Ghana – (GSS, GHS, and Macro International, 2009) 0.30 [0.22; 0.42]
Gambia – (GBOS, and ICF International, 2014) 0.11 [0.08; 0.16]
Equatorial Guinea – (PIP/Guinea Ecuatorial, and ICF International, 2012) 0.16 [0.11; 0.23]
Kenya – (CBS/Kenya, MOH/Kenya, and ORC Macro, 2004) 0.25 [0.21; 0.29]
Kenya – (KNBS/Kenya, NACC/Kenya, NASCP/Kenya, MPHS/Kenya, and KMRI, 2010) 0.26 [0.22; 0.31]
Kenya – (KNBS/Kenya, MOH/Kenya, NACC/Kenya, KMRI/Kenya, NCPD/Kenya, 2015) 0.24 [0.20; 0.28]
Liberia – (LISGIS/Liberia, MOHSW/Liberia, NACP/Liberia, and Macro International, 2008) 0.19 [0.16; 0.23]
Mali – (Samaké et al., 2007) 0.17 [0.14; 0.21]
Mali – (CPS/SSDSPF/Mali, INSTAT/Mali, INFO–STAT/Mali, and ICF International, 2014) 0.30 [0.24; 0.36]
Mali – (INSTAT/Mali, CPSSSD, and ICF, 2019) 0.21 [0.17; 0.26]
Moçambique – (Johnson et al., 2008) 0.25 [0.20; 0.30]
Moçambique – (MISAU/Moçambique, INE/Moçambique, and ICF International, 2013) 0.17 [0.14; 0.21]
Moçambique – (INS, INE, PNCM/Moçambique and ICF, 2019) 0.19 [0.14; 0.25]
Malawi – (NSO/Malawi, and ORC Macro, 2005) 0.42 [0.37; 0.48]
Malawi – (NSO/Malawi, and ICF Macro, 2011) 0.52 [0.45; 0.60]
Malawi – (NSO/Malawi, and ICF, 2017) 0.44 [0.38; 0.51]
Namibia – (January et al., 2005) 0.35 [0.27; 0.47]
Namibia – (MoHSS/Namibia, and ICF International, 2014) 0.26 [0.17; 0.39]
Nigeria – (NPC/Nigeria, and ICF Macro, 2009) 0.18 [0.16; 0.21]
Nigeria – (NPC/Nigeria, and ICF International, 2014) 0.28 [0.25; 0.31]
Nigeria – (NPC/Nigeria, and ICF, 2019) 0.29 [0.25; 0.34]
Rwanda – (INSR/Rwanda, and ORC Macro, 2006) 0.28 [0.22; 0.35]
Rwanda – (NISR/Rwanda, MOH/Rwanda, and ICF International, 2012) 0.16 [0.14; 0.20]
Rwanda – (NISR/Rwanda, MFEP/Rwanda, MOH/Rwanda, and ICF International, 2016) 0.26 [0.20; 0.34]
Senegal – (ANSD/Sénégal, and ICF, 2018) 0.32 [0.24; 0.42]
Sierra Leone – (SSL/Sierra Leone, and ICF International, 2014) 0.11 [0.09; 0.13]
South Sudan – (Ellsberg et al., 2018) 0.46 [0.36; 0.60]
Sao Tome and Principe – (INE/São Tomé, and ICF Macro, 2010) 0.23 [0.17; 0.32]
Chad – (INSEED/Tchad, MSP/Tchad, and ICF International, 2016) 0.28 [0.23; 0.35]
Togo – (MPDAT/Togo, MS/Togo, and ICF International, 2015) 0.30 [0.25; 0.36]
Tanzania – (MoHCDGEC/Tanzania Mainland, MoH/Zanzibar, NBS/Tanzania, OCGS/Zanzibar, and ICF, 2016) 0.35 [0.27; 0.45]
Tanzania – (MoHCDGEC/Tanzania Mainland, MoH/Zanzibar, NBS/Tanzania, OCGS/Zanzibar, and ICF, 2016) 0.43 [0.33; 0.55]
Tanzania – (NBS/Tanzania, and ICF Macro, 2011) 0.27 [0.23; 0.31]
Tanzania – (MoHCDGEC/Tanzania, MoH/Zanzibar, NBS/Tanzania, OCGS/Zanzibar, and ICF, 2016) 0.22 [0.19; 0.25]
Uganda – (UBOS/Uganda, and Macro International, 2007) 0.38 [0.30; 0.48]
Uganda – (Ministry of Health/Uganda, and ICF International, 2012) 0.36 [0.28; 0.45]
Uganda – (Uganda Bureau of Statistics, 2018) 0.33 [0.29; 0.37]
South Africa – (NDH/South Africa, and ICF, 2019) 0.24 [0.20; 0.30]
Zambia – (CSO/Zambia, MOH/Zambia, TDRC/Zambia, and University of Zambia, 2009) 0.20 [0.17; 0.24]
Zambia – (CSO/Zambia, MOH/Zambia, University of Zambia THVL, DPS, TDRC/Zambia, and ICF International, 2015) 0.27 [0.24; 0.30]
Zimbabwe – (CSO/Zimbabwe, and Macro International, 2007) 0.38 [0.32; 0.44]
Zimbabwe – (ZIMSTAT/Zimbabwe, and ICF International, 2012) 0.48 [0.42; 0.55]
Zimbabwe – (ZNSA/Zimbabwe, and ICF International, 2016) 0.26 [0.23; 0.30]
Total 0.27 [0.24; 0.29]
Heterogeneity: $\chi^2_{60} = 872.24$ ($P < .001$), $I^2 = 93\%$
Total 0.26 [0.25; 0.28]
Heterogeneity: $\chi^2_{231} = 5084.25$ ($P < .001$), $I^2 = 95\%$
Residual heterogeneity: $\chi^2_{225} = 4243.96$ ($P < .001$), $I^2 = 95\%$
0.1
0.5
1
2
10
Odds Ratio (95% CI)

**Figure S6.** Forest plot of the random effect meta-analysis for past year for sexual intimate partner violence (IPV) only, as compared to physical and/or sexual violence.

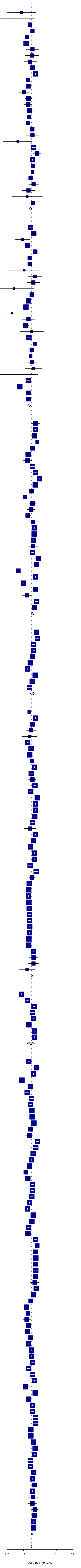


| Source | OR (95% CI) |
|---|---|
| SuperRegion = High Income | |
| Argentina – (Ministerio de Justicia y Derechos Humanos Presidencia de la Nacion, 2017) | 0.08 [0.01; 0.58] |
| Argentina – (Ministerio de Justicia y Derechos Humanos Presidencia de la Nacion, 2017) | 0.03 [0.00; 0.46] |
| Australia – (Mouzos and Makai, 2004) | 0.24 [0.16; 0.38] |
| Austria – (European Union Agency for Fundamental Rights, 2014) | 0.35 [0.14; 0.88] |
| Belgium – (European Union Agency for Fundamental Rights, 2014) | 0.16 [0.07; 0.39] |
| Chile – (Ministerio del Interior y Seguridad Pública, 2013) | 0.14 [0.10; 0.20] |
| Cyprus – (European Union Agency for Fundamental Rights, 2014) | 0.35 [0.14; 0.88] |
| Germany – (European Union Agency for Fundamental Rights, 2014) | 0.33 [0.13; 0.83] |
| Denmark – (European Union Agency for Fundamental Rights, 2014) | 0.25 [0.10; 0.63] |
| Denmark – (National Institute of Public Health, 2017) | 0.35 [0.21; 0.60] |
| Spain – (Ministerio de Sanidad, Servicios Sociales e Igualdad, 2015) | 0.53 [0.38; 0.73] |
| Finland – (European Union Agency for Fundamental Rights, 2014) | 0.19 [0.08; 0.46] |
| Finland – (Institute of Criminology and Legal Policy – University of Helsinki, 2013) | 0.19 [0.12; 0.32] |
| Finland – (Institute of Criminology and Legal Policy – University of Helsinki, 2014) | 0.11 [0.05; 0.22] |
| Finland – (Institute of Criminology and Legal Policy – University of Helsinki, 2015) | 0.20 [0.12; 0.33] |
| Finland – (Institute of Criminology and Legal Policy – University of Helsinki, 2016) | 0.19 [0.11; 0.33] |
| Finland – (Institute of Criminology and Legal Policy – University of Helsinki, 2017) | 0.23 [0.14; 0.36] |
| France – (European Union Agency for Fundamental Rights, 2014) | 0.21 [0.08; 0.50] |
| United Kingdom – (European Union Agency for Fundamental Rights, 2014) | 0.19 [0.08; 0.46] |
| Greece – (European Union Agency for Fundamental Rights, 2014) | 0.33 [0.17; 0.64] |
| Ireland – (European Union Agency for Fundamental Rights, 2014) | 0.35 [0.14; 0.88] |
| Iceland – (Gíslason, 2009) | 0.04 [0.01; 0.33] |
| Italy – (Istat, 2006) | 0.41 [0.32; 0.52] |
| Italy – (European Union Agency for Fundamental Rights, 2014) | 0.65 [0.38; 1.11] |
| Italy – (Istat, 2015) | 0.34 [0.26; 0.46] |
| Japan – (Yoshihama et al., 2005) | 0.36 [0.15; 0.86] |
| Luxembourg – (European Union Agency for Fundamental Rights, 2014) | 0.36 [0.11; 1.13] |
| Malta – (European Union Agency for Fundamental Rights, 2014) | 0.26 [0.11; 0.66] |
| Netherlands – (European Union Agency for Fundamental Rights, 2014) | 0.40 [0.20; 0.79] |
| Portugal – (European Union Agency for Fundamental Rights, 2014) | 0.20 [0.08; 0.48] |
| Singapore – (Bouhours et al., 2013) | 0.17 [0.02; 1.38] |
| Sweden – (European Union Agency for Fundamental Rights, 2014) | 0.39 [0.20; 0.76] |
| Total | 0.27 [0.22; 0.32] |
| Heterogeneity: $\chi^2_{31} = 84.2$ ($P < .001$), $I^2 = 63\%$ | |
| SuperRegion = Central Europe, Eastern Europe & Central Asia | |
| Albania – (Haarr et al., 2013) | 0.27 [0.21; 0.36] |
| Albania – ( The Institut of Statistics of Albania ... Instat/Albania, 2018) | 0.42 [0.30; 0.59] |
| Armenia – (National Statistical Service, 2017) | 0.09 [0.03; 0.24] |
| Azerbaijan – (SSC/Azerbaijan, and Macro International, 2008) | 0.18 [0.12; 0.26] |
| Bulgaria – (European Union Agency for Fundamental Rights, 2014) | 0.50 [0.28; 0.89] |
| Belarus – (Institute of Sociology of NAS Belarus, 2019) | 0.23 [0.11; 0.51] |
| Czechia – (European Union Agency for Fundamental Rights, 2014) | 0.23 [0.09; 0.57] |
| Czechia – (...Buriánek et al., 2013) | 0.11 [0.01; 0.87] |
| Estonia – (European Union Agency for Fundamental Rights, 2014) | 0.49 [0.18; 1.33] |
| Georgia – (UN Women/GEOSTAT, 2018) | 0.39 [0.16; 0.96] |
| Croatia – (European Union Agency for Fundamental Rights, 2014) | 0.03 [0.00; 0.44] |
| Hungary – (European Union Agency for Fundamental Rights, 2014) | 0.32 [0.16; 0.62] |
| Kazakhstan – (Ministry of National Economy of the Republic of Kazakhstan, 2017) | 0.20 [0.13; 0.30] |
| Kyrgyz Republic – (NSC, MOH/Kyrgyz Republic NSC, and ICF International, 2013) | 0.14 [0.10; 0.19] |
| Lithuania – (European Union Agency for Fundamental Rights, 2014) | 0.02 [0.00; 0.34] |
| Latvia – (European Union Agency for Fundamental Rights, 2014) | 0.20 [0.08; 0.48] |
| Moldova – (NCPM/Moldova, and ORC Macro, 2006) | 0.14 [0.09; 0.20] |
| Moldova – (UNW, UNDP, and UNFPA, 2011) | 0.31 [0.06; 1.60] |
| Mongolia – (NSO, UNFPA, SDC, Australian Aid, 2018) | 0.21 [0.17; 0.27] |
| Poland – (European Union Agency for Fundamental Rights, 2014) | 0.49 [0.18; 1.33] |
| Romania – (European Union Agency for Fundamental Rights, 2014) | 0.32 [0.16; 0.62] |
| Serbia – (Otasevic et al., 2005) | 0.27 [0.10; 0.73] |
| Slovakia – (European Union Agency for Fundamental Rights, 2014) | 0.31 [0.16; 0.62] |
| Slovenia – (Institute of Criminology, Ministry of Interior and Faculty of Social Work, 2010) | 0.39 [0.12; 1.26] |
| Slovenia – (European Union Agency for Fundamental Rights, 2014) | 0.04 [0.00; 0.72] |
| Tajikistan – (SA/Tajikistan, MOH/Tajikistan, and ICF International, 2013) | 0.19 [0.14; 0.26] |
| Tajikistan – (SA, MOH/Tajikistan, and ICF, 2018) | 0.06 [0.04; 0.09] |
| Ukraine – (UCSR, SSC/Ukraine, MOH/Ukraine, and Macro International, 2008) | 0.19 [0.12; 0.31] |
| Ukraine – (GfK Ukraine, 2014) | 0.20 [0.11; 0.38] |
| Total | 0.22 [0.17; 0.27] |
| Heterogeneity: $\chi^2_{28} = 105.06$ ($P < .001$), $I^2 = 73\%$ | |
| SuperRegion = South–East Asia, East Asia & Oceania | |
| Cook Islands – (Ministry of Health, 2014) | 0.54 [0.29; 1.01] |
| Fiji – (Fiji Women's Crisis Centre, 2013) | 0.53 [0.43; 0.65] |
| Micronesia – (Department of Health and Social Affairs, 2014) | 0.46 [0.31; 0.69] |
| Hong Kong – (Bouhours et al., 2015) | 0.66 [0.19; 2.37] |
| Indonesia – (UNDP, 2016) | 0.36 [0.20; 0.62] |
| Cambodia – (NIS/Cambodia, DGH/Cambodia, and ORC Macro, 2001) | 0.19 [0.13; 0.28] |
| Cambodia – (NIPH/Cambodia, NIS/Cambodia, and ORC Macro, 2006) | 0.18 [0.10; 0.32] |
| Cambodia – (NIS/Cambodia, DGH/Cambodia, and ICF International, 2015) | 0.34 [0.24; 0.47] |
| Cambodia – (Ministry of Women's Affairs, 2015) | 0.51 [0.39; 0.68] |
| Kiribati – (Ministry of Internal and Social Affairs, 2010) | 0.90 [0.72; 1.12] |
| Laos – (National Commission for the Advancement of Women, 2015) | 0.50 [0.34; 0.75] |
| Maldives – (Ministry of Gender and Family, 2011) | 0.30 [0.17; 0.56] |
| Maldives – (Ministry of Health Maldives, 2018) | 0.12 [0.06; 0.25] |
| Marshall Islands – (DHS/Marshall Islands, 2008) | 0.35 [0.23; 0.54] |
| Marshall Islands – (Ministry of Internal Affairs, 2014) | 0.29 [0.17; 0.47] |
| Myanmar – (MoHS/Myanmar, and ICF, 2017) | 0.18 [0.12; 0.28] |
| Nauru – (Ministry of Home Affairs, 2014) | 0.38 [0.18; 0.79] |
| Philippines – (NSO/Philippines, and ICF Macro, 2009) | 0.45 [0.36; 0.56] |
| Philippines – (PSA/Philippines, and ICF International, 2014) | 0.43 [0.34; 0.56] |
| Philippines – (PSA/Philippines, and ICF, 2018) | 0.39 [0.31; 0.49] |
| Palau – (Ministry of Health, 2014) | 0.37 [0.18; 0.76] |
| Papua New Guinea – (NSO/Papua New Guinea, and ICF, 2019) | 0.35 [0.30; 0.41] |
| Thailand – (Kanchanachitra et al., 2005) | 0.77 [0.54; 1.09] |
| Thailand – (Kanchanachitra et al., 2005) | 0.62 [0.44; 0.88] |
| Timor–leste – (NSD/Timor–Leste, MF/Timor–Leste, and ICF Macro, 2010) | 0.05 [0.03; 0.08] |
| Timor–Leste – (Asia Foundation, 2016) | 0.52 [0.42; 0.66] |
| Timor–Leste – (General Directorate of Statistics, Ministry of Finance/Timor Leste, and ICF, 2018) | 0.10 [0.07; 0.13] |
| Tonga – (Ma'a Fafine mo e Famili, 2012) | 0.54 [0.30; 0.97] |
| Tuvalu – (CSDSPC, 2009) | 0.16 [0.08; 0.32] |
| Vanuatu – (Vanuatu Women's Centre, 2011) | 0.63 [0.51; 0.76] |
| Samoa – ( Posiulai et al., 2005) | 0.45 [0.31; 0.63] |
| Total | 0.35 [0.28; 0.45] |
| Heterogeneity: $\chi^2_{30} = 344.17$ ($P < .001$), $I^2 = 91\%$ | |
| SuperRegion = South Asia | |
| Bangladesh – (Garcia–Moreno et al., 2005) | 0.58 [0.44; 0.77] |
| Bangladesh – (Garcia–Moreno et al., 2005) | 0.68 [0.52; 0.89] |
| Bangladesh – (NIPORT/Bangladesh, Mitra, Associates/Bangladesh, and Macro International, 2009) | 0.39 [0.33; 0.48] |
| Bangladesh – (Bangladesh Bureau of Statistics, 2016) | 0.42 [0.38; 0.45] |
| Bhutan – (National Commission for Women and Children, 2019) | 0.36 [0.25; 0.52] |
| India – (IIPS/India, and Macro International, 2007) | 0.25 [0.24; 0.27] |
| India – (IIPS/India, and ICF, 2017) | 0.18 [0.17; 0.19] |
| Nepal – (MOHP/Nepal, New ERA/Nepal, and ICF International, 2012) | 0.50 [0.39; 0.65] |
| Nepal – (MOH/Nepal, New ERA/Nepal, and ICF, 2017) | 0.33 [0.24; 0.45] |
| Pakistan – (NIPS/Pakistan, and ICF, 2019) | 0.22 [0.16; 0.31] |
| Total | 0.36 [0.26; 0.49] |
| Heterogeneity: $\chi^2_{9} = 405.58$ ($P < .001$), $I^2 = 98\%$ | |
| SuperRegion = Latin America & Caribbean | |
| Belize – (Young Macfarlane Belize Institute for Local Development, 2015) | 0.22 [0.06; 0.78] |
| Bolivia – (Instituto Nacional de Estadística, 2017) | 0.52 [0.46; 0.60] |
| Brazil – (Schraiber et al., 2005) | 0.35 [0.22; 0.55] |
| Brazil – (Schraiber et al., 2005) | 0.29 [0.15; 0.59] |
| Brazil – (Instituto de Pesquisa DataSenado, 2017) | 0.23 [0.08; 0.66] |
| Colombia – (Bott et al., 2015) | 0.18 [0.16; 0.20] |
| Dominican Republic – (CESDEM/Republica Dominicana and Macro International, 2008) | 0.28 [0.23; 0.35] |
| Dominican Republic – (CESDEM/Republica Dominicana, and ICF International, 2014) | 0.24 [0.19; 0.30] |
| Grenada – (Nicholson and Deshong., 2018) | 0.33 [0.15; 0.72] |
| Guatemala – (MSPAS, 2011) | 0.46 [0.39; 0.54] |
| Guatemala – (MSPAS, INE, and Segeplán, 2017) | 0.29 [0.23; 0.36] |
| Guyana – (Contreras–Urbina et al., 2019) | 0.33 [0.20; 0.55] |
| Honduras – (SS/Honduras, INE/Honduras, and Macro International, 2006) | 0.49 [0.42; 0.57] |
| Honduras – (SS/Honduras, INE/Honduras, and ICF International, 2013) | 0.28 [0.23; 0.33] |
| Haïti – (Cayemittes et al., 2001) | 0.66 [0.52; 0.85] |
| Haïti – (Cayemittes et al., 2007) | 0.53 [0.40; 0.69] |
| Haïti – (Cayemittes et al., 2013) | 0.54 [0.45; 0.64] |
| Haïti – (IHE/Haïti, and ICF, 2018) | 0.48 [0.38; 0.61] |
| Jamaica – (National Family Planning Board, 2010) | 0.35 [0.27; 0.45] |
| Jamaica – (Williams et al., 2018) | 0.25 [0.11; 0.57] |
| Mexico – (Instituto National de Estadística Y Geografía, 2004) | 0.53 [0.49; 0.57] |
| Mexico – (Instituto National de Estadística Y Geografía, 2007) | 0.41 [0.39; 0.44] |
| Mexico – (Instituto National de Estadística Y Geografía, 2016) | 0.27 [0.24; 0.30] |
| Nicaragua – (INIDE MINSA, 2008) | 0.45 [0.38; 0.53] |
| Nicaragua – (INIDE MINSA, 2014) | 0.45 [0.37; 0.54] |
| Panama – (Instituto Conmemorativo Gorgas de Estudios de la Salud, 2011) | 0.25 [0.19; 0.33] |
| Peru – (Guezmes Garcia et al., 2005) | 0.57 [0.45; 0.74] |
| Peru – (Guezmes Garcia et al., 2005) | 0.33 [0.21; 0.50] |
| Peru – (INEI/Perú, and ORC Macro, 2007) | 0.22 [0.17; 0.27] |
| Peru – (INEI/Perú, and ORC Macro, 2009) | 0.21 [0.17; 0.25] |
| Peru – (INEI/Perú, and ORC Macro, 2010) | 0.20 [0.17; 0.24] |
| Peru – (INEI/Perú, and ORC Macro, 2011) | 0.21 [0.18; 0.26] |
| Peru – (INEI/Perú, 2012) | 0.22 [0.18; 0.26] |
| Peru – (INEI/Perú, 2013) | 0.22 [0.19; 0.27] |
| Peru – (INEI/Perú, 2014) | 0.22 [0.19; 0.27] |
| Peru – (INEI/Perú, 2015) | 0.24 [0.20; 0.28] |
| Peru – (INEI/Perú, 2016) | 0.22 [0.19; 0.26] |
| Peru – (INEI/Perú, 2017) | 0.21 [0.18; 0.25] |
| Peru – (INEI/Perú, 2018) | 0.21 [0.18; 0.24] |
| El salvador – (Asociación Demográfica Salvadoreña – ADS/El Salvador, 2008) | 0.41 [0.32; 0.52] |
| El salvador – (Navarros–Mantas, Velásquez, and Megías, 2015) | 0.41 [0.23; 0.75] |
| Suriname – (Joseph, Pemberton and Phillip, 2019) | 0.40 [0.20; 0.79] |
| Trinidad and Tobago – (Pemberton and Joseph, 2018) | 0.17 [0.06; 0.48] |
| Total | 0.32 [0.28; 0.36] |
| Heterogeneity: $\chi^2_{42} = 819.03$ ($P < .001$), $I^2 = 95\%$ | |
| SuperRegion = North Africa & Middle East | |
| Afghanistan – (CSO/Afghanistan, MPH/Afghanistan, and ICF, 2017) | 0.08 [0.07; 0.08] |
| Egypt – (MHP/Egypt and ICF International, 2015) | 0.17 [0.13; 0.22] |
| Egypt – (Duvuury et al., 2015) | 0.43 [0.38; 0.48] |
| Jordan – (DoS/Jordan, and Macro International, 2008) | 0.35 [0.26; 0.46] |
| Jordan – (DoS/Jordan, and ICF International, 2013) | 0.39 [0.32; 0.47] |
| Jordan – (DoS/Jordan, and ICF, 2019) | 0.21 [0.17; 0.27] |
| Turkey – (Ministry of Family and Social Policies, Hacettepe University and NEE Institute of Population Studies, 2009) | 0.47 [0.41; 0.55] |
| Turkey – (Ministry of Family and Social Policies, Hacettepe University and NEE Institute of Population Studies, 2015) | 0.45 [0.36; 0.56] |
| Total | 0.28 [0.16; 0.47] |
| Heterogeneity: $\chi^2_{7} = 785.5$ ($P < .001$), $I^2 = 99\%$ | |
| SuperRegion = Sub–Saharan Africa | |
| Angola – (INE/Angola, MINSA/Angola, and ICF, 2017) | 0.21 [0.18; 0.24] |
| Burundi – (MPBGP, MSPLS,– ISTEEBU/Burundi, and ICF, 2017) | 0.59 [0.51; 0.67] |
| Benin – (INSAE/Bénin, and ICF, 2019) | 0.40 [0.31; 0.51] |
| Burkina faso – (Institut National de la Statistique et de la Démographie, 2012) | 0.08 [0.06; 0.11] |
| Central African Republic – (Institut Centrafricain des Statistiques, et des Etudes Economiques et Sociales, 2009) | 0.25 [0.22; 0.29] |
| Côte d'Ivoire – (INS/Côte d'Ivoire, and ICF International, 2013) | 0.16 [0.13; 0.21] |
| Cameroon – (INS/Cameroun, and ORC Macro, 2005) | 0.30 [0.23; 0.39] |
| Cameroon – (INS/Cameroun, and ICF International, 2012) | 0.26 [0.21; 0.32] |
| Cameroon – (MPH/Cameroon, NIS/Cameroon, UNICEF, 2014) | 0.32 [0.28; 0.37] |
| Democratic Republic of the Congo – (MP/Congo, and Macro International, 2008) | 0.33 [0.27; 0.39] |
| Democratic Republic of the Congo – (MPSMRM/Congo, MSP/Congo, and ICF International, 2014) | 0.43 [0.37; 0.49] |
| Comoros – (DGSP/Comores, and ICF International, 2014) | 0.27 [0.14; 0.51] |
| Cabo verde – (ICIEG, INE/Cabo Verde, 2016) | 0.22 [0.13; 0.38] |
| Ethiopia – (Berhane et al., 2005) | 0.69 [0.57; 0.83] |
| Ethiopia – (Deribe et al., 2012) | 0.55 [0.40; 0.76] |
| Ethiopia – (Central Statistical Agency, 2017) | 0.38 [0.30; 0.47] |
| Gabon – (DGS/Gabon, and ICF International, 2013) | 0.30 [0.24; 0.36] |
| Ghana – (GSS, GHS, and Macro International, 2009) | 0.22 [0.15; 0.33] |
| Gambia – (GBOS, and ICF International, 2014) | 0.14 [0.08; 0.24] |
| Equatorial Guinea – (PIP/Guinea Ecuatorial, and ICF International, 2012) | 0.18 [0.12; 0.27] |
| Kenya – (CBS/Kenya, MOH/Kenya, and ORC Macro, 2004) | 0.34 [0.29; 0.42] |
| Kenya – (KNBS/Kenya, NACC/Kenya, NASCP/Kenya, MPHS/Kenya, and KMRI, 2010) | 0.35 [0.30; 0.42] |
| Kenya – (KNBS/Kenya, MOH/Kenya, NACC/Kenya, KMRI/Kenya, NCPD/Kenya, 2015) | 0.32 [0.26; 0.39] |
| Liberia – (LISGIS/Liberia, MOHSW/Liberia, NACP/Liberia, and Macro International, 2008) | 0.23 [0.19; 0.28] |
| Mali – (Samaké et al., 2007) | 0.17 [0.14; 0.21] |
| Mali – (CPS/SSDSPF/Mali, INSTAT/Mali, INFO–STAT/Mali, and ICF International, 2014) | 0.38 [0.30; 0.47] |
| Mali – (INSTAT/Mali, CPSSSD, and ICF, 2019) | 0.32 [0.25; 0.41] |
| Moçambique – (MISAU/Moçambique, INE/Moçambique, and ICF International, 2013) | 0.19 [0.16; 0.23] |
| Moçambique – (INS, INE, PNCM/Moçambique and ICF, 2019) | 0.18 [0.13; 0.27] |
| Malawi – (NSO/Malawi, and ORC Macro, 2005) | 0.53 [0.46; 0.61] |
| Malawi – (NSO/Malawi, and ICF Macro, 2011) | 0.66 [0.42; 1.05] |
| Malawi – (NSO/Malawi, and ICF Macro, 2011) | 0.53 [0.29; 0.97] |
| Malawi – (NSO/Malawi, and ICF Macro, 2011) | 0.56 [0.37; 0.84] |
| Malawi – (NSO/Malawi, and ICF Macro, 2011) | 0.54 [0.29; 1.00] |
| Malawi – (NSO/Malawi, and ICF Macro, 2011) | 0.55 [0.40; 0.77] |
| Malawi – (NSO/Malawi, and ICF Macro, 2011) | 0.51 [0.36; 0.73] |
| Malawi – (NSO/Malawi, and ICF Macro, 2011) | 0.48 [0.26; 0.88] |
| Malawi – (NSO/Malawi, and ICF, 2017) | 0.57 [0.49; 0.67] |
| Namibia – (January et al., 2005) | 0.41 [0.29; 0.59] |
| Namibia – (MoHSS/Namibia, and ICF International, 2014) | 0.27 [0.17; 0.44] |
| Nigeria – (NPC/Nigeria, and ICF Macro, 2009) | 0.15 [0.10; 0.23] |
| Nigeria – (NPC/Nigeria, and ICF Macro, 2009) | 0.19 [0.12; 0.29] |
| Nigeria – (NPC/Nigeria, and ICF Macro, 2009) | 0.16 [0.09; 0.27] |
| Nigeria – (NPC/Nigeria, and ICF Macro, 2009) | 0.20 [0.14; 0.29] |
| Nigeria – (NPC/Nigeria, and ICF Macro, 2009) | 0.17 [0.12; 0.24] |
| Nigeria – (NPC/Nigeria, and ICF Macro, 2009) | 0.27 [0.15; 0.49] |
| Nigeria – (NPC/Nigeria, and ICF Macro, 2009) | 0.19 [0.14; 0.26] |
| Nigeria – (NPC/Nigeria, and ICF International, 2014) | 0.31 [0.27; 0.35] |
| Nigeria – (NPC/Nigeria, and ICF, 2019) | 0.31 [0.26; 0.37] |
| Rwanda – (INSR/Rwanda, and ORC Macro, 2006) | 0.36 [0.28; 0.46] |
| Rwanda – (NISR/Rwanda, MOH/Rwanda, and ICF International, 2012) | 0.19 [0.16; 0.24] |
| Rwanda – (NISR/Rwanda, MFEP/Rwanda, MOH/Rwanda, and ICF International, 2016) | 0.35 [0.25; 0.49] |
| Senegal – (ANSD/Sénégal, and ICF, 2018) | 0.46 [0.33; 0.65] |
| Sierra Leone – (SSL/Sierra Leone, and ICF International, 2014) | 0.14 [0.11; 0.17] |
| South Sudan – (Ellsberg et al., 2018) | 0.50 [0.35; 0.72] |
| Sao Tome and Principe – (INE/São Tomé, and ICF Macro, 2010) | 0.20 [0.14; 0.28] |
| Chad – (INSEED/Tchad, MSP/Tchad, and ICF International, 2016) | 0.35 [0.27; 0.44] |
| Togo – (MPDAT/Togo, MS/Togo, and ICF International, 2015) | 0.35 [0.27; 0.44] |
| Tanzania – (MoHCDGEC/Tanzania Mainland, MoH/Zanzibar, NBS/Tanzania, OCGS/Zanzibar, and ICF, 2016) | 0.54 [0.39; 0.74] |
| Tanzania – (MoHCDGEC/Tanzania Mainland, MoH/Zanzibar, NBS/Tanzania, OCGS/Zanzibar, and ICF, 2016) | 0.55 [0.41; 0.74] |
| Tanzania – (NBS/Tanzania, and ICF Macro, 2011) | 0.28 [0.24; 0.32] |
| Tanzania – (MoHCDGEC/Tanzania, MoH/Zanzibar, NBS/Tanzania, OCGS/Zanzibar, and ICF, 2016) | 0.28 [0.24; 0.32] |
| Uganda – (UBOS/Uganda, and Macro International, 2007) | 0.41 [0.32; 0.52] |
| Uganda – (Ministry of Health/Uganda, and ICF International, 2012) | 0.50 [0.39; 0.64] |
| Uganda – (Uganda Bureau of Statistics, 2018) | 0.47 [0.41; 0.53] |
| South Africa – (NDH/South Africa, and ICF, 2019) | 0.25 [0.19; 0.35] |
| Zambia – (CSO/Zambia, MOH/Zambia, TDRC/Zambia, and University of Zambia, 2009) | 0.27 [0.23; 0.32] |
| Zambia – (CSO/Zambia, MOH/Zambia, University of Zambia THVL, DPS, TDRC/Zambia, and ICF International, 2015) | 0.41 [0.37; 0.47] |
| Zimbabwe – (CSO/Zimbabwe, and Macro International, 2007) | 0.34 [0.29; 0.41] |
| Zimbabwe – (ZIMSTAT/Zimbabwe, and ICF International, 2012) | 0.46 [0.27; 0.79] |
| Zimbabwe – (ZIMSTAT/Zimbabwe, and ICF International, 2012) | 0.34 [0.25; 0.48] |
| Zimbabwe – (ZIMSTAT/Zimbabwe, and ICF International, 2012) | 0.38 [0.18; 0.80] |
| Zimbabwe – (ZIMSTAT/Zimbabwe, and ICF International, 2012) | 0.34 [0.18; 0.63] |
| Zimbabwe – (ZIMSTAT/Zimbabwe, and ICF International, 2012) | 0.49 [0.31; 0.78] |
| Zimbabwe – (ZIMSTAT/Zimbabwe, and ICF International, 2012) | 0.46 [0.31; 0.68] |
| Zimbabwe – (ZIMSTAT/Zimbabwe, and ICF International, 2012) | 0.41 [0.30; 0.56] |
| Zimbabwe – (ZNSA/Zimbabwe, and ICF International, 2016) | 0.41 [0.35; 0.49] |
| Total | 0.32 [0.29; 0.35] |
| Heterogeneity: $\chi^2_{76} = 851.86$ ($P < .001$), $I^2 = 91\%$ | |
| Total | 0.31 [0.29; 0.33] |
| Heterogeneity: $\chi^2_{229} = 3830.92$ ($P < .001$), $I^2 = 94\%$ | |
| Residual heterogeneity: $\chi^2_{223} = 3395.41$ ($P < .001$), $I^2 = 93\%$ | |

**Figure S7.** Forest plot of the random effect meta-analysis for lifetime intimate partner violence (IPV) when all women are surveyed, as compared to ever-partnered women.

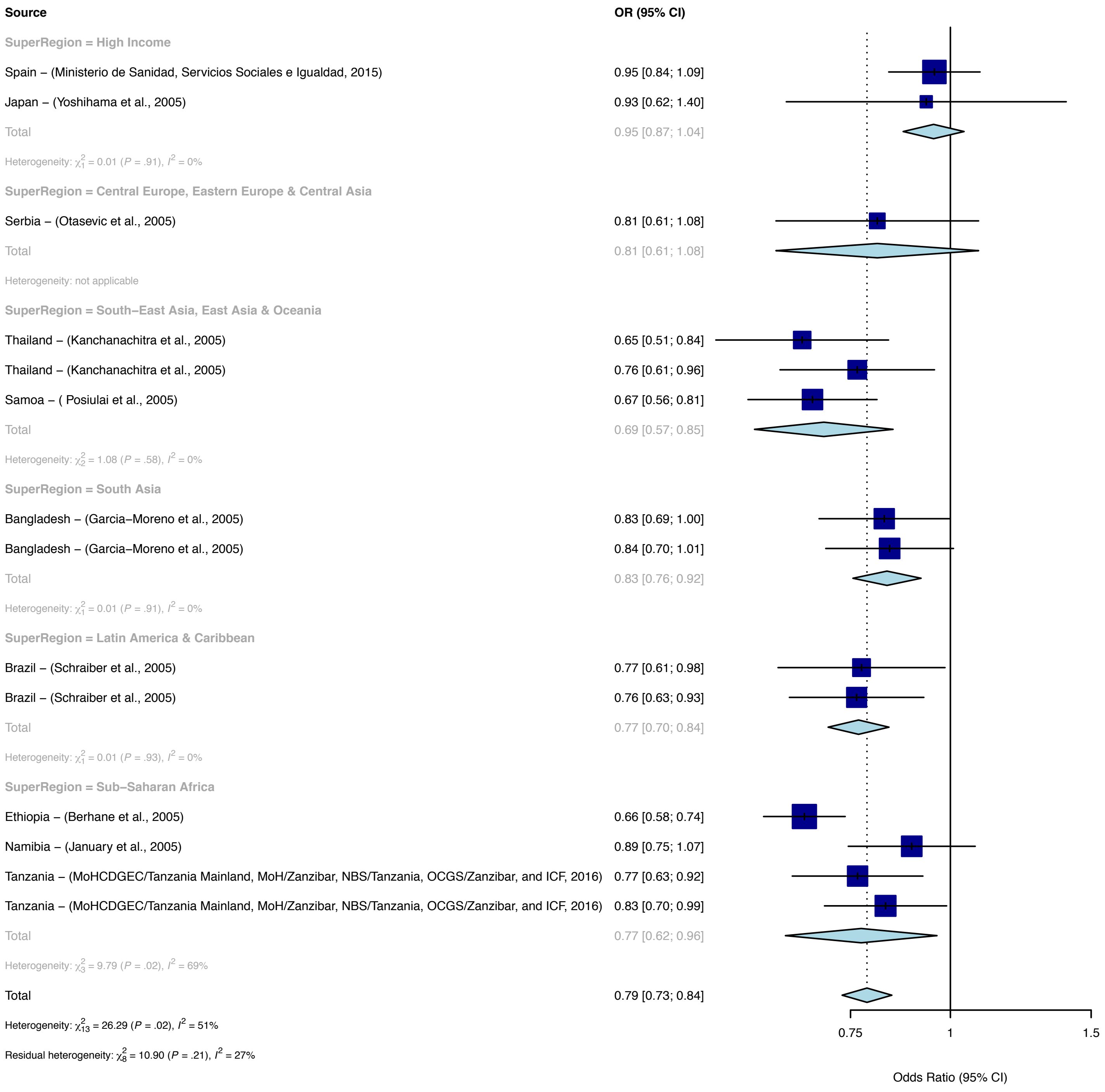
Source
OR (95% CI)
SuperRegion = High Income
Spain – (Ministerio de Sanidad, Servicios Sociales e Igualdad, 2015) 0.95 [0.84; 1.09]
Japan – (Yoshihama et al., 2005) 0.93 [0.62; 1.40]
Total 0.95 [0.87; 1.04]
Heterogeneity: $\chi^2_1 = 0.01$ ($P$ = .91), $I^2 = 0\%$
SuperRegion = Central Europe, Eastern Europe & Central Asia
Serbia – (Otasevic et al., 2005) 0.81 [0.61; 1.08]
Total 0.81 [0.61; 1.08]
Heterogeneity: not applicable
SuperRegion = South–East Asia, East Asia & Oceania
Thailand – (Kanchanachitra et al., 2005) 0.65 [0.51; 0.84]
Thailand – (Kanchanachitra et al., 2005) 0.76 [0.61; 0.96]
Samoa – ( Posiulai et al., 2005) 0.67 [0.56; 0.81]
Total 0.69 [0.57; 0.85]
Heterogeneity: $\chi^2_2 = 1.08$ ($P$ = .58), $I^2 = 0\%$
SuperRegion = South Asia
Bangladesh – (Garcia–Moreno et al., 2005) 0.83 [0.69; 1.00]
Bangladesh – (Garcia–Moreno et al., 2005) 0.84 [0.70; 1.01]
Total 0.83 [0.76; 0.92]
Heterogeneity: $\chi^2_1 = 0.01$ ($P$ = .91), $I^2 = 0\%$
SuperRegion = Latin America & Caribbean
Brazil – (Schraiber et al., 2005) 0.77 [0.61; 0.98]
Brazil – (Schraiber et al., 2005) 0.76 [0.63; 0.93]
Total 0.77 [0.70; 0.84]
Heterogeneity: $\chi^2_1 = 0.01$ ($P$ = .93), $I^2 = 0\%$
SuperRegion = Sub–Saharan Africa
Ethiopia – (Berhane et al., 2005) 0.66 [0.58; 0.74]
Namibia – (January et al., 2005) 0.89 [0.75; 1.07]
Tanzania – (MoHCDGEC/Tanzania Mainland, MoH/Zanzibar, NBS/Tanzania, OCGS/Zanzibar, and ICF, 2016) 0.77 [0.63; 0.92]
Tanzania – (MoHCDGEC/Tanzania Mainland, MoH/Zanzibar, NBS/Tanzania, OCGS/Zanzibar, and ICF, 2016) 0.83 [0.70; 0.99]
Total 0.77 [0.62; 0.96]
Heterogeneity: $\chi^2_3 = 9.79$ ($P$ = .02), $I^2 = 69\%$
Total 0.79 [0.73; 0.84]
Heterogeneity: $\chi^2_{13} = 26.29$ ($P$ = .02), $I^2 = 51\%$
Residual heterogeneity: $\chi^2_8 = 10.90$ ($P$ = .21), $I^2 = 27\%$
0.75
1
1.5
Odds Ratio (95% CI)

**Figure S8.** Forest plot of the random effect meta-analysis for lifetime intimate partner violence (IPV) when currently-partnered women are surveyed, as compared to ever-partnered women.

| Source | OR (95% CI) |
|---|---|
| **SuperRegion = North Africa & Middle East** | |
| Afghanistan – (CSO/Afghanistan, MPH/Afghanistan, and ICF, 2017) | 1.00 [0.96; 1.04] |
| Egypt – (MHP/Egypt and ICF International, 2015) | 0.94 [0.87; 1.02] |
| Jordan – (DoS/Jordan, and Macro International, 2008) | 0.91 [0.81; 1.02] |
| Jordan – (DoS/Jordan, and ICF International, 2013) | 0.94 [0.87; 1.02] |
| Jordan – (DoS/Jordan, and ICF, 2019) | 0.88 [0.81; 0.96] |
| Total | 0.94 [0.89; 1.00] |
| Heterogeneity: $\chi^2_4 = 8.33$ ($P = .08$), $I^2 = 52\%$ | |
| **SuperRegion = Sub–Saharan Africa** | |
| Angola – (INE/Angola, MINSA/Angola, and ICF, 2017) | 0.97 [0.90; 1.04] |
| Burundi – (DHS, 2016) | 0.93 [0.87; 0.99] |
| Benin – (INSAE/Bénin, and ICF, 2019) | 0.97 [0.88; 1.07] |
| Burkina Faso – (DHS, 2010) | 0.93 [0.85; 1.02] |
| Cote d'Ivoire – (DHS, 2012) | 0.98 [0.89; 1.07] |
| Cameroon – (INS/Cameroun, and ORC Macro, 2005) | 0.98 [0.88; 1.10] |
| Cameroon – (INS/Cameroun, and ICF International, 2012) | 0.95 [0.86; 1.03] |
| Democratic Republic of the Congo – (MP/Congo, and Macro International, 2008) | 0.99 [0.89; 1.11] |
| Democratic Republic of the Congo – (MPSMRM/Congo, MSP/Congo, and ICF International, 2014) | 0.94 [0.88; 1.02] |
| Comoros – (DGSP/Comores, and ICF International, 2014) | 0.95 [0.77; 1.17] |
| Ethiopia – (CSA/Ethiopia, and ICF, 2016) | 0.93 [0.85; 1.02] |
| Gabon – (DGS/Gabon, and ICF International, 2013) | 0.92 [0.84; 1.01] |
| Ghana – (GSS, GHS, and Macro International, 2009) | 0.88 [0.75; 1.03] |
| Gambia – (GBOS, and ICF International, 2014) | 0.94 [0.83; 1.05] |
| Kenya – (CBS/Kenya, MOH/Kenya, and ORC Macro, 2004) | 0.94 [0.86; 1.02] |
| Kenya – (KNBS/Kenya, NACC/Kenya, NASCP/Kenya, MPHS/Kenya, and KMRI, 2010) | 0.89 [0.82; 0.97] |
| Kenya – (KNBS/Kenya, MOH/Kenya, NACC/Kenya, KMRI/Kenya, NCPD/Kenya, 2015) | 0.88 [0.80; 0.96] |
| Liberia – (LISGIS/Liberia, MOHSW/Liberia, NACP/Liberia, and Macro International, 2008) | 0.93 [0.84; 1.02] |
| Mali – (Samaké et al., 2007) | 0.98 [0.92; 1.06] |
| Mali – (CPS/SSDSPF/Mali, INSTAT/Mali, INFO–STAT/Mali, and ICF International, 2014) | 1.00 [0.90; 1.11] |
| Mali – (INSTAT/Mali, CPSSSD, and ICF, 2019) | 1.01 [0.91; 1.11] |
| Moçambique – (MISAU/Moçambique, INE/Moçambique, and ICF International, 2013) | 0.93 [0.86; 1.01] |
| Moçambique – (INS, INE, PNCM/Moçambique and ICF, 2019) | 0.95 [0.83; 1.09] |
| Malawi – (NSO/Malawi, and ORC Macro, 2005) | 0.93 [0.86; 0.99] |
| Malawi – (NSO/Malawi, and ICF Macro, 2011) | 0.88 [0.80; 0.96] |
| Malawi – (NSO/Malawi, and ICF, 2017) | 0.91 [0.84; 0.99] |
| Namibia – (MoHSS/Namibia, and ICF International, 2014) | 0.93 [0.78; 1.11] |
| Nigeria – (NPC/Nigeria, and ICF Macro, 2009) | 0.94 [0.89; 0.99] |
| Nigeria – (NPC/Nigeria, and ICF International, 2014) | 0.93 [0.89; 0.98] |
| Nigeria – (NPC/Nigeria, and ICF, 2019) | 0.94 [0.87; 1.01] |
| Rwanda – (INSR/Rwanda, and ORC Macro, 2006) | 0.89 [0.79; 1.00] |
| Rwanda – (NISR/Rwanda, MOH/Rwanda, and ICF International, 2012) | 0.91 [0.82; 1.00] |
| Rwanda – (DHS, 2015) | 0.90 [0.79; 1.04] |
| Senegal – (ANSD/Sénégal, and ICF, 2018) | 0.95 [0.83; 1.08] |
| Sierra Leone – (SSL/Sierra Leone, and ICF International, 2014) | 0.99 [0.91; 1.08] |
| Sao Tome and Principe – (INE/São Tomé, and ICF Macro, 2010) | 0.99 [0.84; 1.15] |
| Chad – (INSEED/Tchad, MSP/Tchad, and ICF International, 2016) | 0.92 [0.83; 1.02] |
| Togo – (MPDAT/Togo, MS/Togo, and ICF International, 2015) | 0.94 [0.86; 1.03] |
| Tanzania – (NBS/Tanzania, and ICF Macro, 2011) | 0.87 [0.81; 0.95] |
| Tanzania – (MoHCDGEC/Tanzania, MoH/Zanzibar, NBS/Tanzania, OCGS/Zanzibar, and ICF, 2016) | 0.87 [0.81; 0.93] |
| Uganda – (UBOS/Uganda, and Macro International, 2007) | 0.96 [0.83; 1.10] |
| Uganda – (DHS, 2011) | 0.95 [0.83; 1.10] |
| Uganda – (UBOS/Uganda, and ICF, 2018) | 0.91 [0.85; 0.97] |
| South Africa – (NDH/South Africa, and ICF, 2019) | 0.87 [0.75; 1.00] |
| Zambia – (CSO/Zambia, MOH/Zambia, TDRC/Zambia, and University of Zambia, 2009) | 0.92 [0.84; 1.01] |
| Zambia – (CSO/Zambia, MOH/Zambia, University of Zambia THVL, DPS, TDRC/Zambia, and ICF International, 2015) | 0.93 [0.88; 0.99] |
| Zimbabwe – (CSO/Zimbabwe, and Macro International, 2007) | 0.93 [0.85; 1.01] |
| Zimbabwe – (ZIMSTAT/Zimbabwe, and ICF International, 2012) | 0.93 [0.86; 1.01] |
| Zimbabwe – (ZNSA/Zimbabwe, and ICF International, 2016) | 0.94 [0.87; 1.02] |
| Total | 0.93 [0.92; 0.94] |
| Heterogeneity: $\chi^2_{48} = 29.47$ ($P = .98$), $I^2 = 0\%$ | |
| **SuperRegion = Central Europe, Eastern Europe & Central Asia** | |
| Armenia – (DHS, 2016) | 0.67 [0.55; 0.81] |
| Azerbaijan – (SSC/Azerbaijan, and Macro International, 2008) | 0.80 [0.70; 0.91] |
| Kyrgyz Republic – (NSC, MOH/Kyrgyz Republic NSC, and ICF International, 2013) | 0.86 [0.78; 0.95] |
| Moldova – (NCPM/Moldova, and ORC Macro, 2006) | 0.82 [0.74; 0.90] |
| Tajikistan – (SA/Tajikistan, MOH/Tajikistan, and ICF International, 2013) | 0.95 [0.85; 1.05] |
| Tajikistan – (SA, MOH/Tajikistan, and ICF, 2018) | 0.94 [0.86; 1.03] |
| Ukraine – (UCSR, SSC/Ukraine, MOH/Ukraine, and Macro International, 2008) | 0.57 [0.47; 0.70] |
| Total | 0.81 [0.69; 0.95] |
| Heterogeneity: $\chi^2_6 = 31.55$ ($P < .001$), $I^2 = 81\%$ | |
| **SuperRegion = Latin America & Caribbean** | |
| Colombia – (DHS, 2005) | 0.79 [0.76; 0.83] |
| Colombia – (Profamilia/Colombia, 2005) | 0.77 [0.74; 0.80] |
| Colombia – (MSPSP, 2015) | 0.74 [0.71; 0.77] |
| Dominican Republic – (Achécar et al., 2003) | 0.78 [0.71; 0.85] |
| Dominican Republic – (CESDEM/Republica Dominicana and Macro International, 2008) | 0.82 [0.75; 0.90] |
| Dominican Republic – (CESDEM/Republica Dominicana, and ICF International, 2014) | 0.82 [0.75; 0.90] |
| Guatemala – (DHS, 2015) | 0.79 [0.73; 0.87] |
| Honduras – (SS/Honduras, INE/Honduras, and Macro International, 2006) | 1.08 [1.01; 1.16] |
| Honduras – (SS/Honduras, INE/Honduras, and ICF International, 2013) | 0.84 [0.79; 0.89] |
| Haïti – (Cayemittes et al., 2001) | 0.84 [0.74; 0.96] |
| Haiti – (DHS, 2006) | 0.96 [0.83; 1.10] |
| Haïti – (Cayemittes et al., 2013) | 0.98 [0.90; 1.06] |
| Haïti – (IHE/Haïti, and ICF, 2018) | 0.93 [0.84; 1.02] |
| Total | 0.85 [0.79; 0.91] |
| Heterogeneity: $\chi^2_{12} = 136.79$ ($P < .001$), $I^2 = 91\%$ | |
| **SuperRegion = South Asia** | |
| India – (DHS, 2006) | 0.98 [0.96; 1.00] |
| Nepal – (MOHP/Nepal, New ERA/Nepal, and ICF International, 2012) | 0.99 [0.89; 1.10] |
| Nepal – (MOH/Nepal, New ERA/Nepal, and ICF, 2017) | 0.96 [0.87; 1.07] |
| Pakistan – (NIPS/Pakistan, and ICF, 2019) | 0.97 [0.87; 1.09] |
| Total | 0.98 [0.97; 0.99] |
| Heterogeneity: $\chi^2_3 = 0.17$ ($P = .98$), $I^2 = 0\%$ | |
| **SuperRegion = South–East Asia, East Asia & Oceania** | |
| Cambodia – (NIS/Cambodia, DGH/Cambodia, and ORC Macro, 2001) | 0.93 [0.80; 1.09] |
| Cambodia – (NIPH/Cambodia, NIS/Cambodia, and ORC Macro, 2006) | 0.93 [0.78; 1.11] |
| Cambodia – (NIS/Cambodia, DGH/Cambodia, and ICF International, 2015) | 0.92 [0.82; 1.04] |
| Maldives – (DHS, 2016) | 0.78 [0.68; 0.89] |
| Myanmar – (DHS, 2016) | 0.88 [0.77; 1.01] |
| Philippines – (NSO/Philippines, and ICF Macro, 2009) | 0.93 [0.85; 1.02] |
| Philippines – (PSA/Philippines, and ICF International, 2014) | 0.89 [0.82; 0.97] |
| Philippines – (PSA/Philippines, and ICF, 2018) | 0.91 [0.85; 0.98] |
| Papua New Guinea – (DHS, 2017) | 0.97 [0.88; 1.06] |
| Timor–leste – (NSD/Timor–Leste, MF/Timor–Leste, and ICF Macro, 2010) | 0.98 [0.86; 1.11] |
| Timor–leste – (GDS, MF/Timor Leste, and ICF, 2018) | 1.00 [0.91; 1.10] |
| Total | 0.92 [0.89; 0.96] |
| Heterogeneity: $\chi^2_{10} = 12.07$ ($P = .28$), $I^2 = 17\%$ | |
| Total | 0.91 [0.90; 0.93] |
| Heterogeneity: $\chi^2_{88} = 460.83$ ($P < .001$), $I^2 = 81\%$ | |
| Residual heterogeneity: $\chi^2_{83} = 218.38$ ($P < .001$), $I^2 = 62\%$ | |

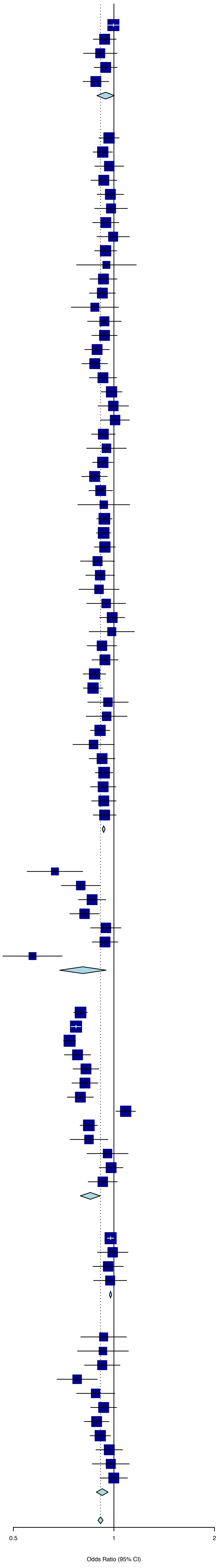

**Figure S9.** Forest plot of the random effect meta-analysis for past year intimate partner violence (IPV) when currently partnered women are surveyed, as compared to ever-partnered women.

| Source | OR (95% CI) |
|---|---|
| **SuperRegion = North Africa & Middle East** | |
| Afghanistan – (CSO/Afghanistan, MPH/Afghanistan, and ICF, 2017) | 1.01 [0.97; 1.04] |
| Egypt – (MHP/Egypt and ICF International, 2015) | 0.99 [0.90; 1.09] |
| Jordan – (DoS/Jordan, and Macro International, 2008) | 0.98 [0.86; 1.12] |
| Jordan – (DoS/Jordan, and ICF International, 2013) | 1.04 [0.94; 1.14] |
| Jordan – (DoS/Jordan, and ICF, 2019) | 0.97 [0.88; 1.07] |
| Total | 1.00 [0.98; 1.02] |
| Heterogeneity: $\chi^2_4 = 1.04$ ($P = .90$), $I^2 = 0\%$ | |
| **SuperRegion = Sub–Saharan Africa** | |
| Angola – (INE/Angola, MINSA/Angola, and ICF, 2017) | 0.99 [0.92; 1.07] |
| Burundi – (DHS, 2016) | 1.14 [1.06; 1.23] |
| Benin – (INSAE/Bénin, and ICF, 2019) | 0.98 [0.87; 1.11] |
| Burkina Faso – (DHS, 2010) | 0.99 [0.90; 1.09] |
| Cote d'Ivoire – (DHS, 2012) | 1.05 [0.96; 1.16] |
| Cameroon – (INS/Cameroun, and ORC Macro, 2005) | 1.06 [0.95; 1.18] |
| Cameroon – (INS/Cameroun, and ICF International, 2012) | 1.05 [0.96; 1.16] |
| Democratic Republic of the Congo – (MP/Congo, and Macro International, 2008) | 1.17 [1.05; 1.31] |
| Democratic Republic of the Congo – (MPSMRM/Congo, MSP/Congo, and ICF International, 2014) | 0.99 [0.92; 1.07] |
| Comoros – (DGSP/Comores, and ICF International, 2014) | 0.92 [0.70; 1.21] |
| Ethiopia – (CSA/Ethiopia, and ICF, 2016) | 1.01 [0.91; 1.12] |
| Gabon – (DGS/Gabon, and ICF International, 2013) | 1.02 [0.93; 1.12] |
| Ghana – (GSS, GHS, and Macro International, 2009) | 0.97 [0.81; 1.15] |
| Gambia – (GBOS, and ICF International, 2014) | 0.97 [0.81; 1.16] |
| Kenya – (CBS/Kenya, MOH/Kenya, and ORC Macro, 2004) | 1.06 [0.97; 1.15] |
| Kenya – (KNBS/Kenya, NACC/Kenya, NASCP/Kenya, MPHS/Kenya, and KMRI, 2010) | 0.93 [0.85; 1.02] |
| Kenya – (KNBS/Kenya, MOH/Kenya, NACC/Kenya, KMRI/Kenya, NCPD/Kenya, 2015) | 1.01 [0.91; 1.11] |
| Liberia – (LISGIS/Liberia, MOHSW/Liberia, NACP/Liberia, and Macro International, 2008) | 0.97 [0.88; 1.07] |
| Mali – (Samaké et al., 2007) | 1.01 [0.94; 1.08] |
| Mali – (CPS/SSDSPF/Mali, INSTAT/Mali, INFO–STAT/Mali, and ICF International, 2014) | 1.00 [0.89; 1.12] |
| Mali – (INSTAT/Mali, CPSSSD, and ICF, 2019) | 1.01 [0.90; 1.14] |
| Moçambique – (MISAU/Moçambique, INE/Moçambique, and ICF International, 2013) | 0.98 [0.90; 1.06] |
| Moçambique – (INS, INE, PNCM/Moçambique and ICF, 2019) | 0.98 [0.84; 1.15] |
| Malawi – (NSO/Malawi, and ORC Macro, 2005) | 0.99 [0.92; 1.06] |
| Malawi – (NSO/Malawi, and ICF Macro, 2011) | 0.96 [0.88; 1.06] |
| Malawi – (NSO/Malawi, and ICF, 2017) | 1.00 [0.91; 1.09] |
| Namibia – (MoHSS/Namibia, and ICF International, 2014) | 0.98 [0.80; 1.19] |
| Nigeria – (NPC/Nigeria, and ICF Macro, 2009) | 0.99 [0.94; 1.05] |
| Nigeria – (NPC/Nigeria, and ICF International, 2014) | 0.97 [0.91; 1.03] |
| Nigeria – (NPC/Nigeria, and ICF, 2019) | 0.98 [0.90; 1.07] |
| Rwanda – (INSR/Rwanda, and ORC Macro, 2006) | 1.11 [0.99; 1.25] |
| Rwanda – (NISR/Rwanda, MOH/Rwanda, and ICF International, 2012) | 1.15 [1.04; 1.27] |
| Rwanda – (DHS, 2015) | 1.04 [0.89; 1.23] |
| Senegal – (ANSD/Sénégal, and ICF, 2018) | 0.94 [0.79; 1.11] |
| Sierra Leone – (SSL/Sierra Leone, and ICF International, 2014) | 1.03 [0.93; 1.13] |
| Sao Tome and Principe – (INE/São Tomé, and ICF Macro, 2010) | 0.98 [0.84; 1.15] |
| Chad – (INSEED/Tchad, MSP/Tchad, and ICF International, 2016) | 0.96 [0.85; 1.08] |
| Togo – (MPDAT/Togo, MS/Togo, and ICF International, 2015) | 1.00 [0.89; 1.12] |
| Tanzania – (NBS/Tanzania, and ICF Macro, 2011) | 0.95 [0.88; 1.03] |
| Tanzania – (MoHCDGEC/Tanzania, MoH/Zanzibar, NBS/Tanzania, OCGS/Zanzibar, and ICF, 2016) | 0.99 [0.92; 1.06] |
| Uganda – (UBOS/Uganda, and Macro International, 2007) | 1.18 [1.02; 1.35] |
| Uganda – (DHS, 2011) | 1.15 [1.00; 1.34] |
| Uganda – (UBOS/Uganda, and ICF, 2018) | 1.05 [0.98; 1.13] |
| South Africa – (NDH/South Africa, and ICF, 2019) | 0.99 [0.82; 1.20] |
| Zambia – (CSO/Zambia, MOH/Zambia, TDRC/Zambia, and University of Zambia, 2009) | 0.99 [0.91; 1.09] |
| Zambia – (CSO/Zambia, MOH/Zambia, University of Zambia THVL, DPS, TDRC/Zambia, and ICF International, 2015) | 1.07 [1.00; 1.14] |
| Zimbabwe – (CSO/Zimbabwe, and Macro International, 2007) | 1.11 [1.01; 1.22] |
| Zimbabwe – (ZIMSTAT/Zimbabwe, and ICF International, 2012) | 1.07 [0.98; 1.17] |
| Zimbabwe – (ZNSA/Zimbabwe, and ICF International, 2016) | 1.06 [0.96; 1.17] |
| Total | 1.02 [1.00; 1.04] |
| Heterogeneity: $\chi^2_{48} = 61.2$ ($P = .10$), $I^2 = 22\%$ | |
| **SuperRegion = Central Europe, Eastern Europe & Central Asia** | |
| Armenia – (DHS, 2016) | 0.73 [0.55; 0.96] |
| Azerbaijan – (SSC/Azerbaijan, and Macro International, 2008) | 0.90 [0.77; 1.04] |
| Kyrgyz Republic – (NSC, MOH/Kyrgyz Republic NSC, and ICF International, 2013) | 0.99 [0.88; 1.10] |
| Moldova – (NCPM/Moldova, and ORC Macro, 2006) | 0.82 [0.74; 0.90] |
| Tajikistan – (SA/Tajikistan, MOH/Tajikistan, and ICF International, 2013) | 0.97 [0.86; 1.09] |
| Tajikistan – (SA, MOH/Tajikistan, and ICF, 2018) | 0.97 [0.88; 1.07] |
| Ukraine – (UCSR, SSC/Ukraine, MOH/Ukraine, and Macro International, 2008) | 0.68 [0.54; 0.84] |
| Total | 0.88 [0.78; 1.00] |
| Heterogeneity: $\chi^2_6 = 18.91$ ($P = .004$), $I^2 = 68\%$ | |
| **SuperRegion = Latin America & Caribbean** | |
| Colombia – (DHS, 2005) | 0.79 [0.76; 0.83] |
| Colombia – (Profamilia/Colombia, 2005) | 0.77 [0.74; 0.80] |
| Colombia – (MSPSP, 2015) | 0.74 [0.71; 0.77] |
| Dominican Republic – (Achécar et al., 2003) | 0.78 [0.71; 0.85] |
| Dominican Republic – (CESDEM/Republica Dominicana and Macro International, 2008) | 0.93 [0.84; 1.03] |
| Dominican Republic – (CESDEM/Republica Dominicana, and ICF International, 2014) | 0.84 [0.75; 0.94] |
| Guatemala – (DHS, 2015) | 1.00 [0.88; 1.13] |
| Honduras – (SS/Honduras, INE/Honduras, and Macro International, 2006) | |
| Honduras – (SS/Honduras, INE/Honduras, and ICF International, 2013) | 1.02 [0.94; 1.11] |
| Haïti – (Cayemittes et al., 2001) | 0.84 [0.74; 0.96] |
| Haiti – (DHS, 2006) | 1.00 [0.86; 1.16] |
| Haïti – (Cayemittes et al., 2013) | 1.07 [0.97; 1.18] |
| Haïti – (IHE/Haïti, and ICF, 2018) | 1.06 [0.94; 1.21] |
| Total | 0.89 [0.81; 0.97] |
| Heterogeneity: $\chi^2_{11} = 136.39$ ($P < .001$), $I^2 = 92\%$ | |
| **SuperRegion = South Asia** | |
| India – (DHS, 2006) | 1.06 [1.03; 1.09] |
| Nepal – (MOHP/Nepal, New ERA/Nepal, and ICF International, 2012) | 1.03 [0.90; 1.18] |
| Nepal – (MOH/Nepal, New ERA/Nepal, and ICF, 2017) | 1.01 [0.87; 1.16] |
| Pakistan – (NIPS/Pakistan, and ICF, 2019) | 1.03 [0.90; 1.18] |
| Total | 1.06 [1.04; 1.08] |
| Heterogeneity: $\chi^2_3 = 0.66$ ($P = .88$), $I^2 = 0\%$ | |
| **SuperRegion = South–East Asia, East Asia & Oceania** | |
| Cambodia – (NIS/Cambodia, DGH/Cambodia, and ORC Macro, 2001) | 0.93 [0.80; 1.09] |
| Cambodia – (NIPH/Cambodia, NIS/Cambodia, and ORC Macro, 2006) | 1.06 [0.86; 1.30] |
| Cambodia – (NIS/Cambodia, DGH/Cambodia, and ICF International, 2015) | 0.99 [0.85; 1.15] |
| Maldives – (DHS, 2016) | 0.90 [0.72; 1.12] |
| Myanmar – (DHS, 2016) | 0.96 [0.82; 1.13] |
| Philippines – (NSO/Philippines, and ICF Macro, 2009) | 1.00 [0.90; 1.12] |
| Philippines – (PSA/Philippines, and ICF International, 2014) | 0.98 [0.87; 1.11] |
| Philippines – (PSA/Philippines, and ICF, 2018) | 1.01 [0.90; 1.12] |
| Papua New Guinea – (DHS, 2017) | 1.05 [0.96; 1.14] |
| Timor–leste – (NSD/Timor–Leste, MF/Timor–Leste, and ICF Macro, 2010) | 1.02 [0.90; 1.17] |
| Timor–leste – (GDS, MF/Timor Leste, and ICF, 2018) | 1.00 [0.91; 1.11] |
| Total | 1.00 [0.98; 1.03] |
| Heterogeneity: $\chi^2_{10} = 3.47$ ($P = .97$), $I^2 = 0\%$ | |
| Total | 0.99 [0.97; 1.01] |
| Heterogeneity: $\chi^2_{87} = 689.14$ ($P < .001$), $I^2 = 87\%$ | |
| Residual heterogeneity: $\chi^2_{82} = 221.67$ ($P < .001$), $I^2 = 63\%$ | |

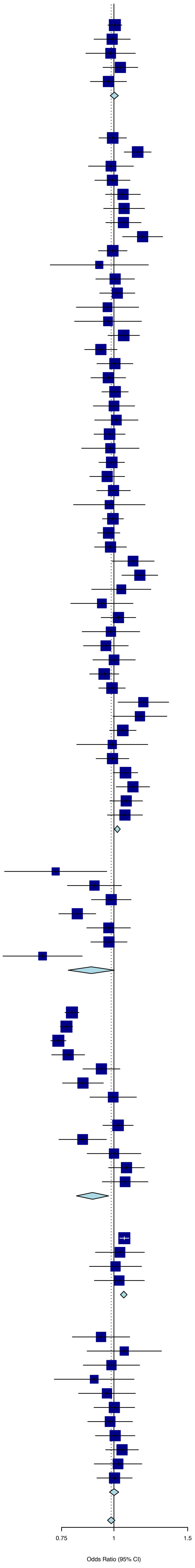

**Figure S10.** Forest plot of the random effect meta-analysis for lifetime intimate partner violence (IPV) when the reference partner is the current or most recent one, as compared to any current or previous ones.

| Source | OR (95% CI) |
|---|---|
| **SuperRegion = High Income** | |
| Argentina – (Ministerio de Justicia y Derechos Humanos Presidencia de la Nacion, 2017) | 0.12 [0.08; 0.20] |
| Argentina – (Ministerio de Justicia y Derechos Humanos Presidencia de la Nacion, 2017) | 0.15 [0.10; 0.24] |
| Australia – (Personal Safety Survey, 2005) | 0.12 [0.08; 0.18] |
| Total | 0.13 [0.09; 0.19] |
| Heterogeneity: $\chi^2_2 = 0.81$ ($P = .67$), $I^2 = 0\%$ | |
| **SuperRegion = Central Europe, Eastern Europe & Central Asia** | |
| Kyrgyz Republic – (NSC, MOH/Kyrgyz Republic NSC, and ICF International, 2013) | 0.94 [0.81; 1.09] |
| Tajikistan – (SA/Tajikistan, MOH/Tajikistan, and ICF International, 2013) | 0.95 [0.80; 1.12] |
| Total | 0.94 [0.90; 0.98] |
| Heterogeneity: $\chi^2_1 = 0$ ($P = .95$), $I^2 = 0\%$ | |
| **SuperRegion = South–East Asia, East Asia & Oceania** | |
| Cambodia – (NIS/Cambodia, DGH/Cambodia, and ICF International, 2015) | 0.91 [0.75; 1.11] |
| Maldives – (Ministry of Health Maldives, 2018) | 0.74 [0.59; 0.93] |
| Myanmar – (MoHS/Myanmar, and ICF, 2017) | 0.93 [0.75; 1.15] |
| Philippines – (PSA/Philippines, and ICF International, 2014) | 0.84 [0.73; 0.97] |
| Philippines – (PSA/Philippines, and ICF, 2018) | 0.80 [0.71; 0.90] |
| Papua New Guinea – (NSO/Papua New Guinea, and ICF, 2019) | 0.95 [0.82; 1.10] |
| Timor–Leste – (General Directorate of Statistics, Ministry of Finance/Timor Leste, and ICF, 2018) | 1.00 [0.85; 1.17] |
| Total | 0.88 [0.80; 0.96] |
| Heterogeneity: $\chi^2_6 = 8.95$ ($P = .18$), $I^2 = 33\%$ | |
| **SuperRegion = South Asia** | |
| Nepal – (MOH/Nepal, New ERA/Nepal, and ICF, 2017) | 0.96 [0.81; 1.14] |
| Pakistan – (NIPS/Pakistan, and ICF, 2019) | 0.96 [0.80; 1.14] |
| Total | 0.96 [0.91; 1.01] |
| Heterogeneity: $\chi^2_1 = 0$ ($P = .95$), $I^2 = 0\%$ | |
| **SuperRegion = Latin America & Caribbean** | |
| Dominican Republic – (CESDEM/Republica Dominicana and Macro International, 2008) | 1.00 [0.85; 1.18] |
| Dominican Republic – (CESDEM/Republica Dominicana, and ICF International, 2014) | 0.64 [0.56; 0.74] |
| Guatemala – (MSPAS, INE, and Segeplán, 2017) | 1.00 [0.87; 1.15] |
| Honduras – (SS/Honduras, INE/Honduras, and ICF International, 2013) | 0.72 [0.65; 0.79] |
| Haïti – (Cayemittes et al., 2013) | 0.79 [0.69; 0.91] |
| Haïti – (IHE/Haïti, and ICF, 2018) | 0.87 [0.74; 1.03] |
| Total | 0.82 [0.68; 0.99] |
| Heterogeneity: $\chi^2_5 = 32.84$ ($P < .001$), $I^2 = 85\%$ | |
| **SuperRegion = North Africa & Middle East** | |
| Afghanistan – (CSO/Afghanistan, MPH/Afghanistan, and ICF, 2017) | 1.00 [0.94; 1.06] |
| Egypt – (MHP/Egypt and ICF International, 2015) | 0.98 [0.87; 1.11] |
| Jordan – (DoS/Jordan, and ICF International, 2013) | 0.96 [0.85; 1.09] |
| Total | 0.99 [0.95; 1.04] |
| Heterogeneity: $\chi^2_2 = 0.36$ ($P = .84$), $I^2 = 0\%$ | |
| **SuperRegion = Sub–Saharan Africa** | |
| Angola – (INE/Angola, MINSA/Angola, and ICF, 2017) | 0.96 [0.87; 1.06] |
| Benin – (INSAE/Bénin, and ICF, 2019) | 0.92 [0.79; 1.09] |
| Côte d'Ivoire – (INS/Côte d'Ivoire, and ICF International, 2013) | 1.00 [0.86; 1.17] |
| Comoros – (DGSP/Comores, and ICF International, 2014) | 0.75 [0.53; 1.08] |
| Cabo verde – (ICIEG, INE/Cabo Verde, 2016) | 1.00 [0.72; 1.40] |
| Ethiopia – (Central Statistical Agency, 2017) | 0.92 [0.79; 1.06] |
| Gabon – (DGS/Gabon, and ICF International, 2013) | 0.82 [0.70; 0.94] |
| Gambia – (GBOS, and ICF International, 2014) | 0.89 [0.74; 1.08] |
| Kenya – (KNBS/Kenya, MOH/Kenya, NACC/Kenya, KMRI/Kenya, NCPD/Kenya, 2015) | 0.95 [0.82; 1.09] |
| Mali – (CPS/SSDSPF/Mali, INSTAT/Mali, INFO–STAT/Mali, and ICF International, 2014) | 0.96 [0.81; 1.14] |
| Moçambique – (MISAU/Moçambique, INE/Moçambique, and ICF International, 2013) | 1.00 [0.88; 1.13] |
| Moçambique – (INS, INE, PNCM/Moçambique and ICF, 2019) | 0.83 [0.68; 1.03] |
| Malawi – (NSO/Malawi, and ICF, 2017) | 0.85 [0.75; 0.97] |
| Namibia – (MoHSS/Namibia, and ICF International, 2014) | 0.91 [0.66; 1.26] |
| Nigeria – (NPC/Nigeria, and ICF International, 2014) | 0.92 [0.85; 0.99] |
| Nigeria – (NPC/Nigeria, and ICF, 2019) | 0.95 [0.84; 1.07] |
| Rwanda – (NISR/Rwanda, MFEP/Rwanda, MOH/Rwanda, and ICF International, 2016) | 0.89 [0.71; 1.11] |
| Senegal – (ANSD/Sénégal, and ICF, 2018) | 0.94 [0.75; 1.17] |
| Sierra Leone – (SSL/Sierra Leone, and ICF International, 2014) | 0.87 [0.76; 1.00] |
| Chad – (INSEED/Tchad, MSP/Tchad, and ICF International, 2016) | 0.94 [0.80; 1.11] |
| Togo – (MPDAT/Togo, MS/Togo, and ICF International, 2015) | 0.85 [0.73; 0.98] |
| Tanzania – (MoHCDGEC/Tanzania, MoH/Zanzibar, NBS/Tanzania, OCGS/Zanzibar, and ICF, 2016) | 0.83 [0.75; 0.92] |
| Uganda – (Ministry of Health/Uganda, and ICF International, 2012) | 0.82 [0.65; 1.02] |
| Uganda – (Uganda Bureau of Statistics, 2018) | 0.88 [0.79; 0.97] |
| South Africa – (NDH/South Africa, and ICF, 2019) | 0.65 [0.56; 0.75] |
| Zambia – (CSO/Zambia, MOH/Zambia, University of Zambia THVL, DPS, TDRC/Zambia, and ICF International, 2015) | 0.88 [0.80; 0.97] |
| Zimbabwe – (ZNSA/Zimbabwe, and ICF International, 2016) | 0.91 [0.80; 1.03] |
| Total | 0.89 [0.86; 0.92] |
| Heterogeneity: $\chi^2_{26} = 35.68$ ($P = .10$), $I^2 = 27\%$ | |
| Total | 0.84 [0.77; 0.93] |
| Heterogeneity: $\chi^2_{49} = 329.05$ ($P < .001$), $I^2 = 85\%$ | |
| Residual heterogeneity: $\chi^2_{43} = 78.63$ ($P < .001$), $I^2 = 45\%$ | |

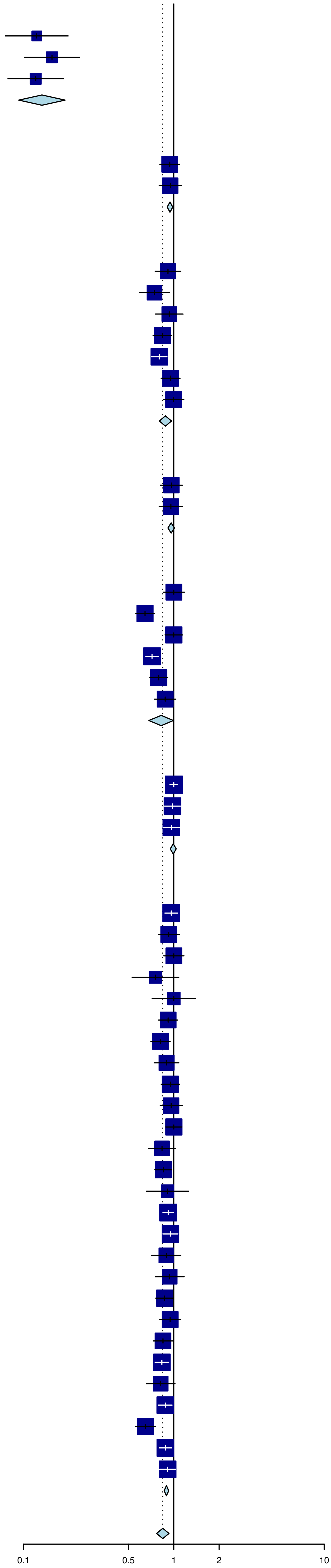

**Figure S11.** Forest plot of the random effect meta-analysis for past year intimate partner violence (IPV) when the reference partner is the current or most recent one, as compared to any current or previous ones.

| Source | OR (95% CI) |
|---|---|
| **SuperRegion = High Income** | |
| Argentina – (Ministerio de Justicia y Derechos Humanos Presidencia de la Nacion, 2017) | 1.24 [0.59; 2.60] |
| Argentina – (Ministerio de Justicia y Derechos Humanos Presidencia de la Nacion, 2017) | 0.88 [0.43; 1.78] |
| Australia – (Personal Safety Survey, 2013) | 0.55 [0.25; 1.20] |
| Canada – (Juristat, 2006) | 0.59 [0.43; 0.80] |
| Canada – (Juristat, 2011) | 0.67 [0.47; 0.96] |
| Canada – (Juristat, 2016) | 0.72 [0.51; 1.03] |
| Total | 0.68 [0.55; 0.85] |
| Heterogeneity: $\chi^2_5 = 4.27$ ($P = .51$), $I^2 = 0\%$ | |
| **SuperRegion = Central Europe, Eastern Europe & Central Asia** | |
| Kyrgyz Republic – (NSC, MOH/Kyrgyz Republic NSC, and ICF International, 2013) | 1.00 [0.84; 1.19] |
| Tajikistan – (SA, MOH/Tajikistan, and ICF, 2018) | 0.88 [0.75; 1.03] |
| Total | 0.93 [0.42; 2.10] |
| Heterogeneity: $\chi^2_1 = 1.1$ ($P = .29$), $I^2 = 9\%$ | |
| **SuperRegion = South–East Asia, East Asia & Oceania** | |
| Myanmar – (MoHS/Myanmar, and ICF, 2017) | 1.00 [0.78; 1.29] |
| Philippines – (PSA/Philippines, and ICF International, 2014) | 0.98 [0.80; 1.20] |
| Philippines – (PSA/Philippines, and ICF, 2018) | 0.98 [0.82; 1.18] |
| Papua New Guinea – (NSO/Papua New Guinea, and ICF, 2019) | 1.00 [0.87; 1.16] |
| Timor–Leste – (General Directorate of Statistics, Ministry of Finance/Timor Leste, and ICF, 2018) | 1.00 [0.85; 1.17] |
| Total | 0.99 [0.98; 1.01] |
| Heterogeneity: $\chi^2_4 = 0.04$ ($P > .99$), $I^2 = 0\%$ | |
| **SuperRegion = Latin America & Caribbean** | |
| Dominican Republic – (CESDEM/Republica Dominicana and Macro International, 2008) | 1.00 [0.83; 1.20] |
| Dominican Republic – (CESDEM/Republica Dominicana, and ICF International, 2014) | 0.97 [0.82; 1.14] |
| Honduras – (SS/Honduras, INE/Honduras, and ICF International, 2013) | 0.99 [0.87; 1.13] |
| Haïti – (Cayemittes et al., 2013) | 0.99 [0.85; 1.17] |
| Haïti – (IHE/Haïti, and ICF, 2018) | 0.99 [0.81; 1.22] |
| Total | 0.99 [0.97; 1.00] |
| Heterogeneity: $\chi^2_4 = 0.08$ ($P > .99$), $I^2 = 0\%$ | |
| **SuperRegion = North Africa & Middle East** | |
| Jordan – (DoS/Jordan, and ICF International, 2013) | 1.00 [0.86; 1.16] |
| Total | 1.00 [0.86; 1.16] |
| Heterogeneity: not applicable | |
| **SuperRegion = Sub–Saharan Africa** | |
| Angola – (INE/Angola, MINSA/Angola, and ICF, 2017) | 1.00 [0.89; 1.11] |
| Burundi – (MPBGP, MSPLS,– ISTEEBU/Burundi, and ICF, 2017) | 0.99 [0.88; 1.12] |
| Benin – (INSAE/Bénin, and ICF, 2019) | 0.60 [0.48; 0.74] |
| Côte d'Ivoire – (INS/Côte d'Ivoire, and ICF International, 2013) | 0.95 [0.81; 1.11] |
| Comoros – (DGSP/Comores, and ICF International, 2014) | 1.00 [0.65; 1.54] |
| Cabo verde – (ICIEG, INE/Cabo Verde, 2016) | 1.00 [0.71; 1.41] |
| Gabon – (DGS/Gabon, and ICF International, 2013) | 0.99 [0.84; 1.16] |
| Gambia – (GBOS, and ICF International, 2014) | 1.00 [0.74; 1.34] |
| Kenya – (CBS/Kenya, MOH/Kenya, and ORC Macro, 2004) | 0.98 [0.84; 1.15] |
| Kenya – (KNBS/Kenya, NACC/Kenya, NASCP/Kenya, MPHS/Kenya, and KMRI, 2010) | 0.90 [0.78; 1.04] |
| Kenya – (KNBS/Kenya, MOH/Kenya, NACC/Kenya, KMRI/Kenya, NCPD/Kenya, 2015) | 1.00 [0.85; 1.17] |
| Mali – (CPS/SSDSPF/Mali, INSTAT/Mali, INFO–STAT/Mali, and ICF International, 2014) | 0.99 [0.82; 1.18] |
| Malawi – (NSO/Malawi, and ICF, 2017) | 0.99 [0.86; 1.14] |
| Namibia – (MoHSS/Namibia, and ICF International, 2014) | 1.00 [0.70; 1.42] |
| Nigeria – (NPC/Nigeria, and ICF International, 2014) | 0.99 [0.90; 1.09] |
| Nigeria – (NPC/Nigeria, and ICF, 2019) | 0.99 [0.86; 1.14] |
| Rwanda – (NISR/Rwanda, MFEP/Rwanda, MOH/Rwanda, and ICF International, 2016) | 0.99 [0.76; 1.29] |
| Senegal – (ANSD/Sénégal, and ICF, 2018) | 1.00 [0.76; 1.32] |
| Sierra Leone – (SSL/Sierra Leone, and ICF International, 2014) | 0.99 [0.86; 1.16] |
| Chad – (INSEED/Tchad, MSP/Tchad, and ICF International, 2016) | 0.99 [0.82; 1.20] |
| Togo – (MPDAT/Togo, MS/Togo, and ICF International, 2015) | 1.00 [0.83; 1.21] |
| Tanzania – (MoHCDGEC/Tanzania, MoH/Zanzibar, NBS/Tanzania, OCGS/Zanzibar, and ICF, 2016) | 0.99 [0.89; 1.12] |
| Uganda – (UBOS/Uganda, and Macro International, 2007) | 0.91 [0.73; 1.14] |
| Uganda – (Ministry of Health/Uganda, and ICF International, 2012) | 1.07 [0.84; 1.34] |
| Uganda – (Uganda Bureau of Statistics, 2018) | 0.99 [0.88; 1.11] |
| Zambia – (CSO/Zambia, MOH/Zambia, TDRC/Zambia, and University of Zambia, 2009) | 1.04 [0.90; 1.20] |
| Zambia – (CSO/Zambia, MOH/Zambia, University of Zambia THVL, DPS, TDRC/Zambia, and ICF International, 2015) | 0.99 [0.89; 1.10] |
| Zimbabwe – (ZNSA/Zimbabwe, and ICF International, 2016) | 0.99 [0.86; 1.15] |
| Total | 0.98 [0.95; 1.01] |
| Heterogeneity: $\chi^2_{27} = 23.21$ ($P = .67$), $I^2 = 0\%$ | |
| Total | 0.97 [0.95; 1.00] |
| Heterogeneity: $\chi^2_{46} = 44.84$ ($P = .52$), $I^2 = 0\%$ | |
| Residual heterogeneity: $\chi^2_{41} = 28.70$ ($P = .93$), $I^2 = 0\%$ | |

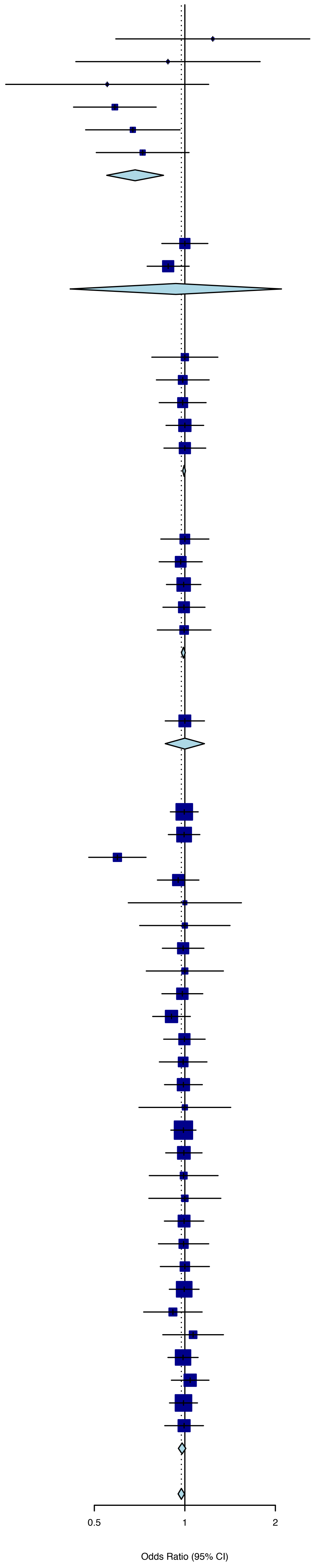

**Figure S12.** Plot of the data used to estimate, through random-effect logistic regression, the adjustment factor for lifetime intimate partner violence (IPV) in urban regions, as compared to a nationally representative sample.

| Source | OR (95% CI) |
|---|---|
| **SuperRegion = Central Europe, Eastern Europe & Central Asia** | |
| Albania – (Organisation for Security and Conflict in Europe, 2019) | 1.13 [0.92; 1.39] |
| Azerbaijan – (SSC/Azerbaijan, and Macro International, 2008) | 0.96 [0.82; 1.12] |
| Bosnia and Hezergovina – (Organisation for Security and Conflict in Europe, 2019) | 1.00 [0.78; 1.27] |
| Czechia – (...Buriánek et al., 2013) | 0.99 [0.80; 1.21] |
| Georgia – (UN Women/GEOSTAT, 2018) | 1.18 [0.90; 1.55] |
| Kazakhstan – (Ministry of National Economy of the Republic of Kazakhstan, 2017) | 0.99 [0.92; 1.07] |
| Kyrgyz Republic – (NSC, MOH/Kyrgyz Republic NSC, and ICF International, 2013) | 1.06 [0.93; 1.22] |
| Moldova – (UNW, UNDP, and UNFPA, 2011) | 0.72 [0.48; 1.07] |
| North Macedonia – (Organisation for Security and Conflict in Europe, 2019) | 1.11 [0.87; 1.42] |
| Montenegro – (Organisation for Security and Conflict in Europe, 2019) | 1.00 [0.79; 1.28] |
| Mongolia – (NSO, UNFPA, SDC, Australian Aid, 2018) | 1.01 [0.92; 1.10] |
| Kosovo – (Organisation for Security and Conflict in Europe, 2019) | 1.00 [0.76; 1.32] |
| Tajikistan – (SA/Tajikistan, MOH/Tajikistan, and ICF International, 2013) | 1.11 [0.94; 1.31] |
| Tajikistan – (SA, MOH/Tajikistan, and ICF, 2018) | 0.84 [0.72; 0.98] |
| Ukraine – (UCSR, SSC/Ukraine, MOH/Ukraine, and Macro International, 2008) | 0.94 [0.78; 1.13] |
| Ukraine – (GfK Ukraine, 2014) | 0.93 [0.73; 1.19] |
| **SuperRegion = South–East Asia, East Asia & Oceania** | |
| Fiji – (Fiji Women's Crisis Centre, 2013) | 0.77 [0.67; 0.87] |
| Cambodia – (NIS/Cambodia, DGH/Cambodia, and ORC Macro, 2001) | 0.94 [0.69; 1.27] |
| Cambodia – (NIPH/Cambodia, NIS/Cambodia, and ORC Macro, 2006) | 1.07 [0.76; 1.52] |
| Cambodia – (NIS/Cambodia, DGH/Cambodia, and ICF International, 2015) | 0.70 [0.61; 0.80] |
| Cambodia – (Ministry of Women's Affairs, 2015) | 0.69 [0.61; 0.79] |
| Maldives – (Ministry of Health Maldives, 2018) | 0.90 [0.73; 1.11] |
| Marshall Islands – (DHS/Marshall Islands, 2008) | 1.00 [0.79; 1.25] |
| Marshall Islands – (Ministry of Internal Affairs, 2014) | 0.99 [0.81; 1.22] |
| Myanmar – (MoHS/Myanmar, and ICF, 2017) | 0.86 [0.69; 1.07] |
| Philippines – (NSO/Philippines, and ICF Macro, 2009) | 0.97 [0.87; 1.07] |
| Philippines – (PSA/Philippines, and ICF International, 2014) | 1.02 [0.91; 1.14] |
| Philippines – (PSA/Philippines, and ICF, 2018) | 0.99 [0.90; 1.09] |
| Papua New Guinea – (NSO/Papua New Guinea, and ICF, 2019) | 1.35 [1.10; 1.67] |
| Timor–leste – (NSD/Timor–Leste, MF/Timor–Leste, and ICF Macro, 2010) | 1.58 [1.29; 1.94] |
| Timor–Leste – (Asia Foundation, 2016) | 1.15 [0.97; 1.36] |
| Timor–Leste – (General Directorate of Statistics, Ministry of Finance/Timor Leste, and ICF, 2018) | 0.65 [0.55; 0.77] |
| Tonga – (Ma'a Fafine mo e Famili, 2012) | 1.09 [0.72; 1.65] |
| **SuperRegion = South Asia** | |
| Bangladesh – (NIPORT/Bangladesh, Mitra, Associates/Bangladesh, and Macro International, 2009) | 0.80 [0.70; 0.92] |
| Bhutan – (National Commission for Women and Children, 2019) | 0.99 [0.77; 1.27] |
| India – (IIPS/India, and Macro International, 2007) | 0.74 [0.71; 0.76] |
| India – (IIPS/India, and ICF, 2017) | 0.76 [0.73; 0.78] |
| Nepal – (MOHP/Nepal, New ERA/Nepal, and ICF International, 2012) | 0.87 [0.73; 1.04] |
| Nepal – (MOH/Nepal, New ERA/Nepal, and ICF, 2017) | 0.95 [0.84; 1.08] |
| Pakistan – (NIPS/Pakistan, and ICF, 2019) | 0.83 [0.71; 0.97] |
| **SuperRegion = Latin America & Caribbean** | |
| Colombia – (Profamilia/Colombia, 2000) | 1.07 [1.00; 1.15] |
| Colombia – (Profamilia/Colombia, 2005) | 1.03 [0.99; 1.07] |
| Colombia – (Profamilia/Colombia, 2005) | 1.05 [1.01; 1.08] |
| Dominican Republic – (Achécar et al., 2003) | 1.07 [0.98; 1.18] |
| Dominican Republic – (CESDEM/Republica Dominicana and Macro International, 2008) | 0.98 [0.89; 1.07] |
| Dominican Republic – (CESDEM/Republica Dominicana, and ICF International, 2014) | 1.04 [0.94; 1.15] |
| Guatemala – (MSPAS, 2011) | 1.12 [1.04; 1.20] |
| Guatemala – (MSPAS, INE, and Segeplán, 2017) | 1.17 [1.04; 1.32] |
| Guyana – (Contreras–Urbina et al., 2019) | 0.87 [0.65; 1.16] |
| Honduras – (SS/Honduras, INE/Honduras, and ICF International, 2013) | 1.13 [1.05; 1.22] |
| Haïti – (Cayemittes et al., 2001) | 1.02 [0.86; 1.22] |
| Haïti – (Cayemittes et al., 2007) | 0.94 [0.71; 1.25] |
| Haïti – (Cayemittes et al., 2013) | 1.14 [1.02; 1.27] |
| Haïti – (IHE/Haïti, and ICF, 2018) | 1.23 [1.08; 1.41] |
| Jamaica – (National Family Planning Board, 2010) | 1.03 [0.92; 1.14] |
| Jamaica – (Williams et al., 2018) | 1.01 [0.80; 1.28] |
| Nicaragua – (INIDE MINSA, 2008) | 1.17 [1.09; 1.25] |
| Nicaragua – (INIDE MINSA, 2014) | 1.24 [1.15; 1.33] |
| Panama – (Instituto Conmemorativo Gorgas de Estudios de la Salud, 2011) | 1.06 [0.88; 1.28] |
| Panama – (Instituto Conmemorativo Gorgas de Estudios de la Salud, 2018) | 0.91 [0.75; 1.11] |
| Peru – (INEI/Perú, 2012) | 1.04 [0.98; 1.10] |
| Peru – (INEI/Perú, 2013) | 1.03 [0.98; 1.09] |
| Peru – (INEI/Perú, 2014) | 1.03 [0.97; 1.09] |
| Peru – (INEI/Perú, 2015) | 1.02 [0.97; 1.08] |
| Peru – (INEI/Perú, 2016) | 1.03 [0.98; 1.07] |
| Peru – (INEI/Perú, 2017) | 1.04 [0.99; 1.09] |
| Paraguay – (Centro paraguayo de estudios de población, 2009) | 1.17 [1.04; 1.32] |
| El salvador – (Asociación Demográfica Salvadoreña – ADS/El Salvador, 2008) | 1.17 [1.07; 1.28] |
| Trinidad and Tobago – (Pemberton and Joseph, 2018) | 1.00 [0.81; 1.23] |
| **SuperRegion = North Africa & Middle East** | |
| Afghanistan – (CSO/Afghanistan, MPH/Afghanistan, and ICF, 2017) | 0.66 [0.62; 0.71] |
| Egypt – (MHP/Egypt and ICF International, 2015) | 0.90 [0.80; 1.00] |
| Egypt – (Duvuury et al., 2015) | 0.75 [0.71; 0.80] |
| Jordan – (DoS/Jordan, and Macro International, 2008) | 1.05 [0.93; 1.17] |
| Jordan – (DoS/Jordan, and ICF International, 2013) | 1.04 [0.96; 1.13] |
| Jordan – (DoS/Jordan, and ICF, 2019) | 1.02 [0.93; 1.11] |
| Turkey – (Ministry of Family and Social Policies, Hacettepe University and NEE Institute of Population Studies, 2009) | 0.94 [0.88; 0.99] |
| Turkey – (Ministry of Family and Social Policies, Hacettepe University and NEE Institute of Population Studies, 2015) | 0.98 [0.90; 1.06] |
| **SuperRegion = Sub–Saharan Africa** | |
| Angola – (INE/Angola, MINSA/Angola, and ICF, 2017) | 1.06 [0.99; 1.14] |
| Burundi – (MPBGP, MSPLS,– ISTEEBU/Burundi, and ICF, 2017) | 0.54 [0.46; 0.64] |
| Benin – (INSAE/Bénin, and ICF, 2019) | 0.90 [0.78; 1.03] |
| Burkina faso – (Institut National de la Statistique et de la Démographie, 2012) | 1.36 [1.19; 1.56] |
| Central African Republic – (Institut Centrafricain des Statistiques, et des Etudes Economiques et Sociales, 2009) | 1.40 [1.30; 1.51] |
| Côte d'Ivoire – (INS/Côte d'Ivoire, and ICF International, 2013) | 1.19 [1.05; 1.34] |
| Cameroon – (INS/Cameroun, and ORC Macro, 2005) | 1.11 [0.96; 1.28] |
| Cameroon – (INS/Cameroun, and ICF International, 2012) | 0.96 [0.89; 1.03] |
| Cabo verde – (Instituto Nacional de Estatística, 2008) | 1.18 [0.87; 1.60] |
| Cabo verde – (ICIEG, INE/Cabo Verde, 2016) | 1.11 [0.89; 1.39] |
| Ethiopia – (Central Statistical Agency, 2017) | 0.69 [0.57; 0.83] |
| Gabon – (DGS/Gabon, and ICF International, 2013) | 0.97 [0.88; 1.07] |
| Ghana – (GSS, GHS, and Macro International, 2009) | 1.05 [0.85; 1.29] |
| Gambia – (GBOS, and ICF International, 2014) | 0.84 [0.72; 0.98] |
| Equatorial Guinea – (PIP/Guinea Ecuatorial, and ICF International, 2012) | 0.91 [0.70; 1.17] |
| Kenya – (CBS/Kenya, MOH/Kenya, and ORC Macro, 2004) | 0.75 [0.65; 0.88] |
| Kenya – (KNBS/Kenya, NACC/Kenya, NASCP/Kenya, MPHS/Kenya, and KMRI, 2010) | 0.77 [0.67; 0.89] |
| Kenya – (KNBS/Kenya, MOH/Kenya, NACC/Kenya, KMRI/Kenya, NCPD/Kenya, 2015) | 0.89 [0.79; 1.00] |
| Liberia – (LISGIS/Liberia, MOHSW/Liberia, NACP/Liberia, and Macro International, 2008) | 1.16 [1.02; 1.32] |
| Mali – (Samaké et al., 2007) | 0.87 [0.78; 0.97] |
| Mali – (CPS/SSDSPF/Mali, INSTAT/Mali, INFO–STAT/Mali, and ICF International, 2014) | 0.84 [0.69; 1.01] |
| Mali – (INSTAT/Mali, CPSSSD, and ICF, 2019) | 1.05 [0.89; 1.24] |
| Moçambique – (MISAU/Moçambique, INE/Moçambique, and ICF International, 2013) | 1.23 [1.10; 1.37] |
| Moçambique – (INS, INE, PNCM/Moçambique and ICF, 2019) | 1.43 [1.19; 1.72] |
| Malawi – (NSO/Malawi, and ORC Macro, 2005) | 0.98 [0.85; 1.12] |
| Malawi – (NSO/Malawi, and ICF Macro, 2011) | 1.34 [1.16; 1.55] |
| Malawi – (NSO/Malawi, and ICF, 2017) | 0.92 [0.79; 1.08] |
| Namibia – (MoHSS/Namibia, and ICF International, 2014) | 0.98 [0.77; 1.25] |
| Nigeria – (NPC/Nigeria, and ICF Macro, 2009) | 0.96 [0.89; 1.04] |
| Nigeria – (NPC/Nigeria, and ICF International, 2014) | 1.11 [1.04; 1.19] |
| Nigeria – (NPC/Nigeria, and ICF, 2019) | 0.95 [0.87; 1.05] |
| Rwanda – (INSR/Rwanda, and ORC Macro, 2006) | 0.90 [0.70; 1.15] |
| Rwanda – (NISR/Rwanda, MFEP/Rwanda, MOH/Rwanda, and ICF International, 2016) | 0.78 [0.60; 1.02] |
| Senegal – (ANSD/Sénégal, and ICF, 2018) | 1.05 [0.88; 1.27] |
| Sierra Leone – (SSL/Sierra Leone, and ICF International, 2014) | 1.05 [0.92; 1.19] |
| Sao Tome and Principe – (INE/São Tomé, and ICF Macro, 2010) | 1.10 [0.92; 1.31] |
| Chad – (INSEED/Tchad, MSP/Tchad, and ICF International, 2016) | 0.99 [0.83; 1.18] |
| Togo – (MPDAT/Togo, MS/Togo, and ICF International, 2015) | 0.88 [0.78; 1.00] |
| Tanzania – (NBS/Tanzania, and ICF Macro, 2011) | 0.93 [0.83; 1.05] |
| Tanzania – (MoHCDGEC/Tanzania, MoH/Zanzibar, NBS/Tanzania, OCGS/Zanzibar, and ICF, 2016) | 0.87 [0.79; 0.96] |
| Uganda – (UBOS/Uganda, and Macro International, 2007) | 0.56 [0.42; 0.73] |
| Uganda – (Ministry of Health/Uganda, and ICF International, 2012) | 0.77 [0.59; 0.99] |
| Uganda – (Uganda Bureau of Statistics, 2018) | 0.69 [0.62; 0.77] |
| South Africa – (NDH/South Africa, and ICF, 2019) | 0.98 [0.88; 1.08] |
| Zambia – (CSO/Zambia, MOH/Zambia, TDRC/Zambia, and University of Zambia, 2009) | 1.26 [1.12; 1.42] |
| Zambia – (CSO/Zambia, MOH/Zambia, University of Zambia THVL, DPS, TDRC/Zambia, and ICF International, 2015) | 1.00 [0.92; 1.08] |
| Zimbabwe – (CSO/Zimbabwe, and Macro International, 2007) | 0.79 [0.70; 0.89] |
| Zimbabwe – (ZIMSTAT/Zimbabwe, and ICF International, 2012) | 0.93 [0.83; 1.04] |
| Zimbabwe – (ZNSA/Zimbabwe, and ICF International, 2016) | 0.96 [0.87; 1.07] |

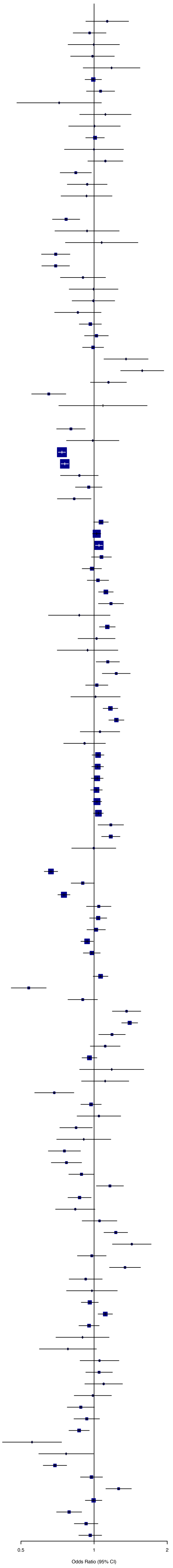


Heterogeneity: $\chi^2_{125} = 1467.34$ ($P < .001$), $I^2 = 91\%$

**Figure S13.** Plot of the data used to estimate, through random-effect logistic regression, the adjustment factor for past year intimate partner violence (IPV) in urban regions, as compared to a nationally representative sample.

| Source | OR (95% CI) |
|---|---|
| SuperRegion = Central Europe, Eastern Europe & Central Asia | |
| Azerbaijan – (SSC/Azerbaijan, and Macro International, 2008) | 0.94 [0.71; 1.25] |
| Czechia – (...Buriánek et al., 2013) | 0.75 [0.25; 2.25] |
| Georgia – (UN Women/GEOSTAT, 2018) | 0.75 [0.21; 2.66] |
| Kyrgyz Republic – (NSC, MOH/Kyrgyz Republic NSC, and ICF International, 2013) | 0.90 [0.69; 1.15] |
| Moldova – (NCPM/Moldova, and ORC Macro, 2006) | 0.75 [0.57; 0.99] |
| Moldova – (UNW, UNDP, and UNFPA, 2011) | 0.69 [0.25; 1.91] |
| Mongolia – (NSO, UNFPA, SDC, Australian Aid, 2018) | 1.00 [0.83; 1.22] |
| Tajikistan – (SA/Tajikistan, MOH/Tajikistan, and ICF International, 2013) | 1.09 [0.81; 1.47] |
| Tajikistan – (SA, MOH/Tajikistan, and ICF, 2018) | 0.82 [0.63; 1.07] |
| SuperRegion = South–East Asia, East Asia & Oceania | |
| Fiji – (Fiji Women's Crisis Centre, 2013) | 0.91 [0.72; 1.16] |
| Cambodia – (NIPH/Cambodia, NIS/Cambodia, and ORC Macro, 2006) | 0.72 [0.34; 1.53] |
| Cambodia – (Ministry of Women's Affairs, 2015) | 0.68 [0.46; 1.00] |
| Maldives – (Ministry of Health Maldives, 2018) | 0.97 [0.60; 1.57] |
| Marshall Islands – (Ministry of Internal Affairs, 2014) | 1.03 [0.68; 1.55] |
| Myanmar – (MoHS/Myanmar, and ICF, 2017) | 0.78 [0.51; 1.19] |
| Philippines – (PSA/Philippines, and ICF International, 2014) | 0.96 [0.75; 1.23] |
| Philippines – (PSA/Philippines, and ICF, 2018) | 0.92 [0.73; 1.16] |
| Papua New Guinea – (NSO/Papua New Guinea, and ICF, 2019) | 1.42 [1.03; 1.95] |
| Timor–leste – (NSD/Timor–Leste, MF/Timor–Leste, and ICF Macro, 2010) | 1.19 [0.85; 1.67] |
| Timor–Leste – (Asia Foundation, 2016) | 1.08 [0.81; 1.45] |
| Timor–Leste – (General Directorate of Statistics, Ministry of Finance/Timor Leste, and ICF, 2018) | 0.69 [0.53; 0.90] |
| Tonga – (Ma'a Fafine mo e Famili, 2012) | 1.41 [0.65; 3.06] |
| SuperRegion = South Asia | |
| Bhutan – (National Commission for Women and Children, 2019) | 0.95 [0.64; 1.41] |
| India – (IIPS/India, and Macro International, 2007) | 0.74 [0.69; 0.78] |
| India – (IIPS/India, and ICF, 2017) | 0.77 [0.73; 0.82] |
| Nepal – (MOHP/Nepal, New ERA/Nepal, and ICF International, 2012) | 1.12 [0.80; 1.58] |
| Nepal – (MOH/Nepal, New ERA/Nepal, and ICF, 2017) | 0.90 [0.68; 1.18] |
| Pakistan – (NIPS/Pakistan, and ICF, 2019) | 0.72 [0.52; 1.00] |
| SuperRegion = Latin America & Caribbean | |
| Dominican Republic – (CESDEM/Republica Dominicana and Macro International, 2008) | 0.97 [0.82; 1.15] |
| Dominican Republic – (CESDEM/Republica Dominicana, and ICF International, 2014) | 1.09 [0.91; 1.30] |
| Guatemala – (MSPAS, 2011) | 1.09 [0.93; 1.28] |
| Guatemala – (MSPAS, INE, and Segeplán, 2017) | 0.98 [0.75; 1.26] |
| Guyana – (Contreras–Urbina et al., 2019) | 0.57 [0.24; 1.35] |
| Honduras – (SS/Honduras, INE/Honduras, and Macro International, 2006) | 0.99 [0.85; 1.15] |
| Honduras – (SS/Honduras, INE/Honduras, and ICF International, 2013) | 1.07 [0.91; 1.25] |
| Haïti – (Cayemittes et al., 2001) | 0.82 [0.60; 1.13] |
| Haïti – (Cayemittes et al., 2007) | 1.01 [0.75; 1.36] |
| Haïti – (Cayemittes et al., 2013) | 1.11 [0.91; 1.36] |
| Haïti – (IHE/Haïti, and ICF, 2018) | 1.18 [0.91; 1.52] |
| Jamaica – (National Family Planning Board, 2010) | 1.10 [0.86; 1.40] |
| Jamaica – (Williams et al., 2018) | 1.07 [0.56; 2.03] |
| Nicaragua – (INIDE MINSA, 2008) | 1.14 [0.96; 1.35] |
| Nicaragua – (INIDE MINSA, 2014) | 1.19 [1.00; 1.42] |
| Peru – (INEI/Perú, 2012) | 1.04 [0.92; 1.18] |
| Peru – (INEI/Perú, 2013) | 1.01 [0.89; 1.15] |
| Peru – (INEI/Perú, 2014) | 1.04 [0.91; 1.18] |
| Peru – (INEI/Perú, 2015) | 1.00 [0.88; 1.13] |
| Peru – (INEI/Perú, 2016) | 1.02 [0.92; 1.13] |
| Peru – (INEI/Perú, 2017) | 1.03 [0.92; 1.15] |
| Peru – (INEI/Perú, 2018) | 0.99 [0.71; 1.37] |
| Paraguay – (Centro paraguayo de estudios de población, 2009) | 1.03 [0.79; 1.34] |
| El salvador – (Asociación Demográfica Salvadoreña – ADS/El Salvador, 2008) | 1.06 [0.84; 1.33] |
| Trinidad and Tobago – (Pemberton and Joseph, 2018) | 0.87 [0.44; 1.72] |
| SuperRegion = North Africa & Middle East | |
| Afghanistan – (CSO/Afghanistan, MPH/Afghanistan, and ICF, 2017) | 0.70 [0.63; 0.77] |
| Egypt – (MHP/Egypt and ICF International, 2015) | 0.89 [0.71; 1.11] |
| Egypt – (Duvuury et al., 2015) | 0.83 [0.73; 0.94] |
| Jordan – (DoS/Jordan, and Macro International, 2008) | 0.98 [0.79; 1.23] |
| Jordan – (DoS/Jordan, and ICF International, 2013) | 1.06 [0.91; 1.24] |
| Jordan – (DoS/Jordan, and ICF, 2019) | 1.02 [0.87; 1.19] |
| Turkey – (Ministry of Family and Social Policies, Hacettepe University and NEE Institute of Population Studies, 2009) | 0.98 [0.86; 1.12] |
| Turkey – (Ministry of Family and Social Policies, Hacettepe University and NEE Institute of Population Studies, 2015) | 1.03 [0.85; 1.25] |
| SuperRegion = Sub–Saharan Africa | |
| Angola – (INE/Angola, MINSA/Angola, and ICF, 2017) | 1.05 [0.93; 1.18] |
| Burundi – (MPBGP, MSPLS,– ISTEEBU/Burundi, and ICF, 2017) | 0.46 [0.33; 0.64] |
| Benin – (INSAE/Bénin, and ICF, 2019) | 0.90 [0.69; 1.18] |
| Central African Republic – (Institut Centrafricain des Statistiques, et des Etudes Economiques et Sociales, 2009) | 1.37 [1.21; 1.56] |
| Côte d'Ivoire – (INS/Côte d'Ivoire, and ICF International, 2013) | 1.23 [1.01; 1.51] |
| Cameroon – (INS/Cameroun, and ORC Macro, 2005) | 1.10 [0.86; 1.41] |
| Democratic Republic of the Congo – (MP/Congo, and Macro International, 2008) | 0.99 [0.79; 1.25] |
| Cabo verde – (ICIEG, INE/Cabo Verde, 2016) | 1.12 [0.78; 1.62] |
| Ethiopia – (Central Statistical Agency, 2017) | 0.55 [0.39; 0.79] |
| Gabon – (DGS/Gabon, and ICF International, 2013) | 0.96 [0.82; 1.14] |
| Ghana – (GSS, GHS, and Macro International, 2009) | 1.08 [0.76; 1.52] |
| Gambia – (GBOS, and ICF International, 2014) | 0.94 [0.65; 1.36] |
| Kenya – (CBS/Kenya, MOH/Kenya, and ORC Macro, 2004) | 0.68 [0.51; 0.90] |
| Kenya – (KNBS/Kenya, NACC/Kenya, NASCP/Kenya, MPHS/Kenya, and KMRI, 2010) | 0.87 [0.69; 1.11] |
| Kenya – (KNBS/Kenya, MOH/Kenya, NACC/Kenya, KMRI/Kenya, NCPD/Kenya, 2015) | 0.98 [0.79; 1.21] |
| Liberia – (LISGIS/Liberia, MOHSW/Liberia, NACP/Liberia, and Macro International, 2008) | 1.14 [0.93; 1.41] |
| Mali – (Samaké et al., 2007) | 0.88 [0.74; 1.05] |
| Mali – (CPS/SSDSPF/Mali, INSTAT/Mali, INFO–STAT/Mali, and ICF International, 2014) | 0.81 [0.59; 1.12] |
| Mali – (INSTAT/Mali, CPSSSD, and ICF, 2019) | 0.85 [0.61; 1.17] |
| Moçambique – (MISAU/Moçambique, INE/Moçambique, and ICF International, 2013) | 1.18 [0.98; 1.42] |
| Moçambique – (INS, INE, PNCM/Moçambique and ICF, 2019) | 1.43 [1.05; 1.95] |
| Malawi – (NSO/Malawi, and ORC Macro, 2005) | 0.77 [0.60; 1.01] |
| Namibia – (MoHSS/Namibia, and ICF International, 2014) | 0.95 [0.63; 1.44] |
| Nigeria – (NPC/Nigeria, and ICF, 2019) | 0.86 [0.71; 1.03] |
| Rwanda – (INSR/Rwanda, and ORC Macro, 2006) | 1.01 [0.66; 1.54] |
| Rwanda – (NISR/Rwanda, MOH/Rwanda, and ICF International, 2012) | 0.68 [0.48; 0.95] |
| Rwanda – (NISR/Rwanda, MFEP/Rwanda, MOH/Rwanda, and ICF International, 2016) | 0.88 [0.53; 1.45] |
| Senegal – (ANSD/Sénégal, and ICF, 2018) | 1.16 [0.82; 1.64] |
| Sierra Leone – (SSL/Sierra Leone, and ICF International, 2014) | 1.20 [0.96; 1.48] |
| Sao Tome and Principe – (INE/São Tomé, and ICF Macro, 2010) | 1.11 [0.83; 1.48] |
| Chad – (INSEED/Tchad, MSP/Tchad, and ICF International, 2016) | 1.04 [0.75; 1.45] |
| Togo – (MPDAT/Togo, MS/Togo, and ICF International, 2015) | 0.92 [0.71; 1.18] |
| Tanzania – (NBS/Tanzania, and ICF Macro, 2011) | 0.89 [0.73; 1.09] |
| Tanzania – (MoHCDGEC/Tanzania, MoH/Zanzibar, NBS/Tanzania, OCGS/Zanzibar, and ICF, 2016) | 0.90 [0.76; 1.06] |
| Uganda – (UBOS/Uganda, and Macro International, 2007) | 0.57 [0.35; 0.91] |
| Uganda – (Ministry of Health/Uganda, and ICF International, 2012) | 0.74 [0.47; 1.18] |
| Uganda – (Uganda Bureau of Statistics, 2018) | 0.68 [0.55; 0.83] |
| Zambia – (CSO/Zambia, MOH/Zambia, TDRC/Zambia, and University of Zambia, 2009) | 1.32 [1.09; 1.59] |
| Zambia – (CSO/Zambia, MOH/Zambia, University of Zambia THVL, DPS, TDRC/Zambia, and ICF International, 2015) | 0.95 [0.83; 1.10] |
| Zimbabwe – (CSO/Zimbabwe, and Macro International, 2007) | 0.82 [0.67; 1.01] |
| Zimbabwe – (ZNSA/Zimbabwe, and ICF International, 2016) | 1.00 [0.82; 1.23] |

Heterogeneity: $\chi^2_{101} = 355.27$ ($P < .001$), $I^2 = 72\%$

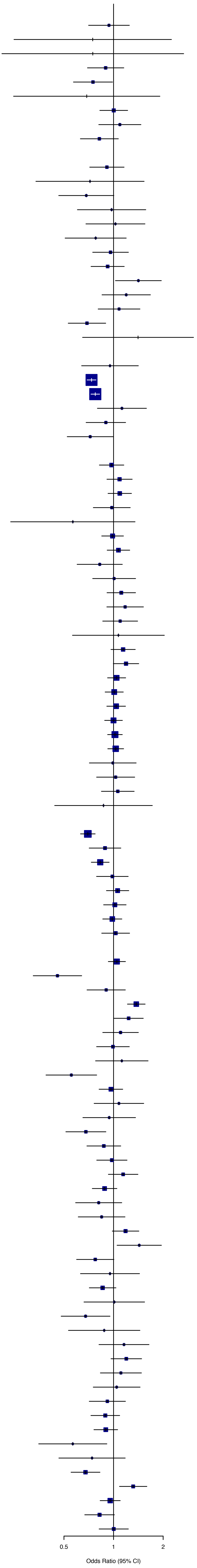

**Figure S14.** Plot of the data used to estimate, through random-effect logistic regression, the adjustment factor for lifetime intimate partner violence (IPV) in rural regions, as compared to a nationally representative sample.

| Source | OR (95% CI) |
| --- | --- |
| SuperRegion = Central Europe, Eastern Europe & Central Asia | |
| Albania – (Organisation for Security and Conflict in Europe, 2019) | 0.94 [0.75; 1.18] |
| Azerbaijan – (SSC/Azerbaijan, and Macro International, 2008) | 1.05 [0.89; 1.24] |
| Bosnia and Hezergovina – (Organisation for Security and Conflict in Europe, 2019) | 1.00 [0.80; 1.26] |
| Czechia – (...Buriánek et al., 2013) | 1.04 [0.78; 1.40] |
| Georgia – (UN Women/GEOSTAT, 2018) | 0.81 [0.59; 1.10] |
| Kazakhstan – (Ministry of National Economy of the Republic of Kazakhstan, 2017) | 1.01 [0.93; 1.11] |
| Kyrgyz Republic – (NSC, MOH/Kyrgyz Republic NSC, and ICF International, 2013) | 0.97 [0.87; 1.08] |
| Moldova – (UNW, UNDP, and UNFPA, 2011) | 1.30 [0.89; 1.89] |
| North Macedonia – (Organisation for Security and Conflict in Europe, 2019) | 0.68 [0.48; 0.95] |
| Montenegro – (Organisation for Security and Conflict in Europe, 2019) | 1.07 [0.78; 1.47] |
| Mongolia – (NSO, UNFPA, SDC, Australian Aid, 2018) | 0.98 [0.90; 1.07] |
| Kosovo – (Organisation for Security and Conflict in Europe, 2019) | 0.90 [0.69; 1.17] |
| Tajikistan – (SA/Tajikistan, MOH/Tajikistan, and ICF International, 2013) | 0.96 [0.86; 1.08] |
| Tajikistan – (SA, MOH/Tajikistan, and ICF, 2018) | 1.05 [0.96; 1.16] |
| Ukraine – (UCSR, SSC/Ukraine, MOH/Ukraine, and Macro International, 2008) | 1.14 [0.90; 1.44] |
| Ukraine – (GfK Ukraine, 2014) | 1.15 [0.86; 1.55] |
| SuperRegion = South–East Asia, East Asia & Oceania | |
| Fiji – (Fiji Women's Crisis Centre, 2013) | 1.27 [1.12; 1.45] |
| Cambodia – (NIS/Cambodia, DGH/Cambodia, and ORC Macro, 2001) | 1.01 [0.86; 1.19] |
| Cambodia – (NIPH/Cambodia, NIS/Cambodia, and ORC Macro, 2006) | 0.98 [0.81; 1.18] |
| Cambodia – (NIS/Cambodia, DGH/Cambodia, and ICF International, 2015) | 1.06 [0.93; 1.21] |
| Cambodia – (Ministry of Women's Affairs, 2015) | 1.10 [0.97; 1.24] |
| Maldives – (Ministry of Health Maldives, 2018) | 1.06 [0.89; 1.25] |
| Marshall Islands – (DHS/Marshall Islands, 2008) | 1.01 [0.73; 1.41] |
| Marshall Islands – (Ministry of Internal Affairs, 2014) | 1.03 [0.79; 1.34] |
| Myanmar – (MoHS/Myanmar, and ICF, 2017) | 1.05 [0.91; 1.22] |
| Philippines – (NSO/Philippines, and ICF Macro, 2009) | 1.04 [0.93; 1.16] |
| Philippines – (PSA/Philippines, and ICF International, 2014) | 0.98 [0.87; 1.09] |
| Philippines – (PSA/Philippines, and ICF, 2018) | 1.01 [0.92; 1.11] |
| Papua New Guinea – (NSO/Papua New Guinea, and ICF, 2019) | 0.96 [0.87; 1.06] |
| Timor–leste – (NSD/Timor–Leste, MF/Timor–Leste, and ICF Macro, 2010) | 0.85 [0.74; 0.99] |
| Timor–Leste – (Asia Foundation, 2016) | 0.95 [0.80; 1.12] |
| Timor–Leste – (General Directorate of Statistics, Ministry of Finance/Timor Leste, and ICF, 2018) | 1.15 [1.04; 1.28] |
| Tonga – (Ma'a Fafine mo e Famili, 2012) | 0.97 [0.73; 1.30] |
| SuperRegion = South Asia | |
| Bangladesh – (NIPORT/Bangladesh, Mitra, Associates/Bangladesh, and Macro International, 2009) | 1.07 [0.98; 1.17] |
| Bhutan – (National Commission for Women and Children, 2019) | 1.00 [0.82; 1.22] |
| India – (IIPS/India, and Macro International, 2007) | 1.13 [1.11; 1.16] |
| India – (IIPS/India, and ICF, 2017) | 1.15 [1.12; 1.18] |
| Nepal – (MOHP/Nepal, New ERA/Nepal, and ICF International, 2012) | 1.05 [0.93; 1.17] |
| Nepal – (MOH/Nepal, New ERA/Nepal, and ICF, 2017) | 1.07 [0.93; 1.23] |
| Pakistan – (NIPS/Pakistan, and ICF, 2019) | 1.11 [0.97; 1.26] |
| SuperRegion = Latin America & Caribbean | |
| Colombia – (Profamilia/Colombia, 2000) | 0.82 [0.74; 0.91] |
| Colombia – (Profamilia/Colombia, 2005) | 0.93 [0.88; 0.99] |
| Colombia – (Profamilia/Colombia, 2005) | 0.85 [0.81; 0.90] |
| Dominican Republic – (Achécar et al., 2003) | 0.86 [0.76; 0.98] |
| Dominican Republic – (CESDEM/Republica Dominicana and Macro International, 2008) | 1.06 [0.93; 1.20] |
| Dominican Republic – (CESDEM/Republica Dominicana, and ICF International, 2014) | 0.90 [0.77; 1.05] |
| Guatemala – (MSPAS, 2011) | 0.91 [0.85; 0.97] |
| Guatemala – (MSPAS, INE, and Segeplán, 2017) | 0.88 [0.79; 0.99] |
| Guyana – (Contreras–Urbina et al., 2019) | 1.00 [0.84; 1.19] |
| Honduras – (SS/Honduras, INE/Honduras, and ICF International, 2013) | 0.87 [0.80; 0.94] |
| Haïti – (Cayemittes et al., 2001) | 0.98 [0.84; 1.14] |
| Haïti – (Cayemittes et al., 2007) | 1.04 [0.92; 1.19] |
| Haïti – (Cayemittes et al., 2013) | 0.89 [0.80; 1.00] |
| Haïti – (IHE/Haïti, and ICF, 2018) | 0.84 [0.74; 0.96] |
| Jamaica – (National Family Planning Board, 2010) | 0.97 [0.88; 1.07] |
| Jamaica – (Williams et al., 2018) | 0.98 [0.76; 1.27] |
| Nicaragua – (INIDE MINSA, 2008) | 0.81 [0.75; 0.87] |
| Nicaragua – (INIDE MINSA, 2014) | 0.71 [0.66; 0.77] |
| Panama – (Instituto Conmemorativo Gorgas de Estudios de la Salud, 2011) | 0.82 [0.67; 1.00] |
| Panama – (Instituto Conmemorativo Gorgas de Estudios de la Salud, 2018) | 1.37 [1.15; 1.64] |
| Peru – (INEI/Perú, 2012) | 0.91 [0.85; 0.99] |
| Peru – (INEI/Perú, 2013) | 0.92 [0.85; 0.99] |
| Peru – (INEI/Perú, 2014) | 0.92 [0.85; 1.00] |
| Peru – (INEI/Perú, 2015) | 0.94 [0.87; 1.02] |
| Peru – (INEI/Perú, 2016) | 0.92 [0.86; 0.98] |
| Peru – (INEI/Perú, 2017) | 0.88 [0.82; 0.94] |
| Paraguay – (Centro paraguayo de estudios de población, 2009) | 0.75 [0.65; 0.86] |
| El salvador – (Asociación Demográfica Salvadoreña – ADS/El Salvador, 2008) | 0.80 [0.73; 0.88] |
| Trinidad and Tobago – (Pemberton and Joseph, 2018) | 1.01 [0.76; 1.33] |
| SuperRegion = North Africa & Middle East | |
| Afghanistan – (CSO/Afghanistan, MPH/Afghanistan, and ICF, 2017) | 1.12 [1.07; 1.17] |
| Egypt – (MHP/Egypt and ICF International, 2015) | 1.05 [0.97; 1.15] |
| Egypt – (Duvuury et al., 2015) | 1.24 [1.17; 1.30] |
| Jordan – (DoS/Jordan, and Macro International, 2008) | 0.98 [0.79; 1.23] |
| Jordan – (DoS/Jordan, and ICF International, 2013) | 0.82 [0.71; 0.95] |
| Jordan – (DoS/Jordan, and ICF, 2019) | 0.81 [0.66; 1.01] |
| Turkey – (Ministry of Family and Social Policies, Hacettepe University and NEE Institute of Population Studies, 2009) | 1.21 [1.11; 1.32] |
| Turkey – (Ministry of Family and Social Policies, Hacettepe University and NEE Institute of Population Studies, 2015) | 1.07 [0.96; 1.18] |
| SuperRegion = Sub–Saharan Africa | |
| Angola – (INE/Angola, MINSA/Angola, and ICF, 2017) | 0.90 [0.83; 0.98] |
| Burundi – (MPBGP, MSPLS,– ISTEEBU/Burundi, and ICF, 2017) | 1.07 [1.00; 1.15] |
| Benin – (INSAE/Bénin, and ICF, 2019) | 1.07 [0.96; 1.21] |
| Burkina faso – (Institut National de la Statistique et de la Démographie, 2012) | 0.90 [0.82; 1.00] |
| Central African Republic – (Institut Centrafricain des Statistiques, et des Etudes Economiques et Sociales, 2009) | 0.77 [0.72; 0.83] |
| Côte d'Ivoire – (INS/Côte d'Ivoire, and ICF International, 2013) | 0.88 [0.78; 0.99] |
| Cameroon – (INS/Cameroun, and ORC Macro, 2005) | 0.90 [0.78; 1.04] |
| Cameroon – (INS/Cameroun, and ICF International, 2012) | 1.05 [0.94; 1.17] |
| Cabo verde – (Instituto Nacional de Estatística, 2008) | 0.78 [0.54; 1.12] |
| Cabo verde – (ICIEG, INE/Cabo Verde, 2016) | 0.71 [0.49; 1.02] |
| Ethiopia – (Central Statistical Agency, 2017) | 1.08 [0.98; 1.19] |
| Gabon – (DGS/Gabon, and ICF International, 2013) | 1.21 [1.00; 1.47] |
| Ghana – (GSS, GHS, and Macro International, 2009) | 0.96 [0.79; 1.16] |
| Gambia – (GBOS, and ICF International, 2014) | 1.17 [1.01; 1.35] |
| Equatorial Guinea – (PIP/Guinea Ecuatorial, and ICF International, 2012) | 1.08 [0.85; 1.38] |
| Kenya – (CBS/Kenya, MOH/Kenya, and ORC Macro, 2004) | 1.09 [0.99; 1.20] |
| Kenya – (KNBS/Kenya, NACC/Kenya, NASCP/Kenya, MPHS/Kenya, and KMRI, 2010) | 1.08 [0.99; 1.19] |
| Kenya – (KNBS/Kenya, MOH/Kenya, NACC/Kenya, KMRI/Kenya, NCPD/Kenya, 2015) | 1.08 [0.98; 1.20] |
| Liberia – (LISGIS/Liberia, MOHSW/Liberia, NACP/Liberia, and Macro International, 2008) | 0.92 [0.83; 1.03] |
| Mali – (Samaké et al., 2007) | 1.06 [0.98; 1.15] |
| Mali – (CPS/SSDSPF/Mali, INSTAT/Mali, INFO–STAT/Mali, and ICF International, 2014) | 1.05 [0.94; 1.17] |
| Mali – (INSTAT/Mali, CPSSSD, and ICF, 2019) | 0.99 [0.89; 1.10] |
| Moçambique – (MISAU/Moçambique, INE/Moçambique, and ICF International, 2013) | 0.91 [0.83; 0.99] |
| Moçambique – (INS, INE, PNCM/Moçambique and ICF, 2019) | 0.83 [0.71; 0.97] |
| Malawi – (NSO/Malawi, and ORC Macro, 2005) | 1.01 [0.93; 1.08] |
| Malawi – (NSO/Malawi, and ICF Macro, 2011) | 0.94 [0.86; 1.02] |
| Malawi – (NSO/Malawi, and ICF, 2017) | 1.01 [0.93; 1.10] |
| Namibia – (MoHSS/Namibia, and ICF International, 2014) | 1.03 [0.79; 1.35] |
| Nigeria – (NPC/Nigeria, and ICF Macro, 2009) | 1.02 [0.96; 1.08] |
| Nigeria – (NPC/Nigeria, and ICF International, 2014) | 0.93 [0.87; 0.98] |
| Nigeria – (NPC/Nigeria, and ICF, 2019) | 1.04 [0.96; 1.14] |
| Rwanda – (INSR/Rwanda, and ORC Macro, 2006) | 1.01 [0.89; 1.15] |
| Rwanda – (NISR/Rwanda, MFEP/Rwanda, MOH/Rwanda, and ICF International, 2016) | 1.05 [0.90; 1.22] |
| Senegal – (ANSD/Sénégal, and ICF, 2018) | 0.97 [0.82; 1.14] |
| Sierra Leone – (SSL/Sierra Leone, and ICF International, 2014) | 0.97 [0.89; 1.07] |
| Sao Tome and Principe – (INE/São Tomé, and ICF Macro, 2010) | 0.89 [0.73; 1.08] |
| Chad – (INSEED/Tchad, MSP/Tchad, and ICF International, 2016) | 1.01 [0.90; 1.12] |
| Togo – (MPDAT/Togo, MS/Togo, and ICF International, 2015) | 1.09 [0.98; 1.21] |
| Tanzania – (NBS/Tanzania, and ICF Macro, 2011) | 1.02 [0.94; 1.11] |
| Tanzania – (MoHCDGEC/Tanzania, MoH/Zanzibar, NBS/Tanzania, OCGS/Zanzibar, and ICF, 2016) | 1.07 [1.00; 1.15] |
| Uganda – (UBOS/Uganda, and Macro International, 2007) | 1.11 [0.95; 1.28] |
| Uganda – (Ministry of Health/Uganda, and ICF International, 2012) | 1.05 [0.91; 1.22] |
| Uganda – (Uganda Bureau of Statistics, 2018) | 1.12 [1.04; 1.20] |
| South Africa – (NDH/South Africa, and ICF, 2019) | 1.04 [0.92; 1.18] |
| Zambia – (CSO/Zambia, MOH/Zambia, TDRC/Zambia, and University of Zambia, 2009) | 0.87 [0.79; 0.96] |
| Zambia – (CSO/Zambia, MOH/Zambia, University of Zambia THVL, DPS, TDRC/Zambia, and ICF International, 2015) | 1.00 [0.94; 1.08] |
| Zimbabwe – (CSO/Zimbabwe, and Macro International, 2007) | 1.13 [1.03; 1.24] |
| Zimbabwe – (ZIMSTAT/Zimbabwe, and ICF International, 2012) | 1.04 [0.95; 1.14] |
| Zimbabwe – (ZNSA/Zimbabwe, and ICF International, 2016) | 1.02 [0.94; 1.12] |

Heterogeneity: $\chi^2_{125} = 826.19$ ($P < .001$), $I^2 = 85\%$

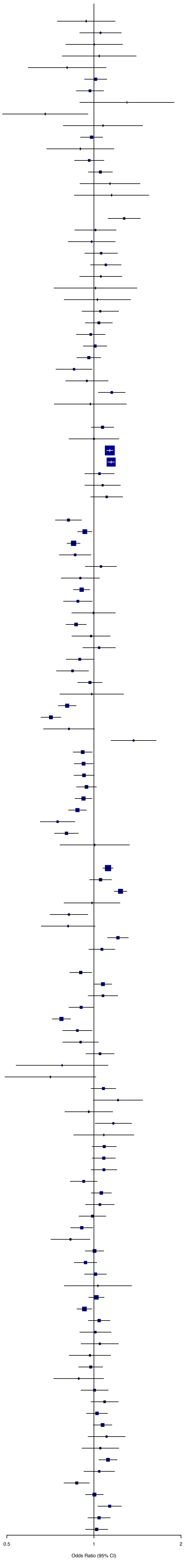

**Figure S15.** Plot of the data used to estimate, through random-effect logistic regression, the adjustment factor for past year intimate partner violence (IPV) in rural regions, as compared to a nationally representative sample.

| Source | OR (95% CI) |
|---|---|
| **SuperRegion = Central Europe, Eastern Europe & Central Asia** | |
| Azerbaijan – (SSC/Azerbaijan, and Macro International, 2008) | 1.08 [0.80; 1.46] |
| Czechia – (...Buriánek et al., 2013) | 1.29 [0.34; 4.83] |
| Georgia – (UN Women/GEOSTAT, 2018) | 1.51 [0.56; 4.04] |
| Kyrgyz Republic – (NSC, MOH/Kyrgyz Republic NSC, and ICF International, 2013) | 1.07 [0.88; 1.30] |
| Moldova – (NCPM/Moldova, and ORC Macro, 2006) | 1.18 [0.95; 1.46] |
| Moldova – (UNW, UNDP, and UNFPA, 2011) | 1.27 [0.51; 3.17] |
| Mongolia – (NSO, UNFPA, SDC, Australian Aid, 2018) | 0.99 [0.82; 1.20] |
| Tajikistan – (SA/Tajikistan, MOH/Tajikistan, and ICF International, 2013) | 0.97 [0.79; 1.19] |
| Tajikistan – (SA, MOH/Tajikistan, and ICF, 2018) | 1.07 [0.90; 1.26] |
| **SuperRegion = South–East Asia, East Asia & Oceania** | |
| Fiji – (Fiji Women's Crisis Centre, 2013) | 1.09 [0.87; 1.35] |
| Cambodia – (NIPH/Cambodia, NIS/Cambodia, and ORC Macro, 2006) | 1.07 [0.75; 1.52] |
| Cambodia – (Ministry of Women's Affairs, 2015) | 1.10 [0.85; 1.42] |
| Maldives – (Ministry of Health Maldives, 2018) | 1.02 [0.69; 1.50] |
| Marshall Islands – (Ministry of Internal Affairs, 2014) | 1.00 [0.58; 1.72] |
| Myanmar – (MoHS/Myanmar, and ICF, 2017) | 1.08 [0.83; 1.42] |
| Philippines – (PSA/Philippines, and ICF International, 2014) | 1.03 [0.81; 1.32] |
| Philippines – (PSA/Philippines, and ICF, 2018) | 1.06 [0.86; 1.31] |
| Papua New Guinea – (NSO/Papua New Guinea, and ICF, 2019) | 0.96 [0.83; 1.11] |
| Timor–leste – (NSD/Timor–Leste, MF/Timor–Leste, and ICF Macro, 2010) | 0.95 [0.75; 1.19] |
| Timor–Leste – (Asia Foundation, 2016) | 0.97 [0.77; 1.23] |
| Timor–Leste – (General Directorate of Statistics, Ministry of Finance/Timor Leste, and ICF, 2018) | 1.13 [0.95; 1.35] |
| Tonga – (Ma'a Fafine mo e Famili, 2012) | 0.88 [0.49; 1.57] |
| **SuperRegion = South Asia** | |
| Bhutan – (National Commission for Women and Children, 2019) | 1.05 [0.70; 1.58] |
| India – (IIPS/India, and Macro International, 2007) | 1.13 [1.08; 1.18] |
| India – (IIPS/India, and ICF, 2017) | 1.13 [1.08; 1.18] |
| Nepal – (MOHP/Nepal, New ERA/Nepal, and ICF International, 2012) | 0.97 [0.76; 1.23] |
| Nepal – (MOH/Nepal, New ERA/Nepal, and ICF, 2017) | 1.16 [0.86; 1.55] |
| Pakistan – (NIPS/Pakistan, and ICF, 2019) | 1.17 [0.92; 1.48] |
| **SuperRegion = Latin America & Caribbean** | |
| Dominican Republic – (CESDEM/Republica Dominicana and Macro International, 2008) | 1.06 [0.84; 1.33] |
| Dominican Republic – (CESDEM/Republica Dominicana, and ICF International, 2014) | 0.88 [0.67; 1.15] |
| Guatemala – (MSPAS, 2011) | 0.92 [0.79; 1.08] |
| Guatemala – (MSPAS, INE, and Segeplán, 2017) | 1.03 [0.82; 1.29] |
| Guyana – (Contreras–Urbina et al., 2019) | 1.11 [0.73; 1.68] |
| Honduras – (SS/Honduras, INE/Honduras, and Macro International, 2006) | 1.00 [0.85; 1.17] |
| Honduras – (SS/Honduras, INE/Honduras, and ICF International, 2013) | 0.95 [0.80; 1.12] |
| Haïti – (Cayemittes et al., 2001) | 1.13 [0.88; 1.46] |
| Haïti – (Cayemittes et al., 2007) | 0.99 [0.75; 1.32] |
| Haïti – (Cayemittes et al., 2013) | 0.92 [0.75; 1.11] |
| Haïti – (IHE/Haïti, and ICF, 2018) | 0.87 [0.68; 1.11] |
| Jamaica – (National Family Planning Board, 2010) | 0.85 [0.67; 1.07] |
| Jamaica – (Williams et al., 2018) | 0.91 [0.44; 1.87] |
| Nicaragua – (INIDE MINSA, 2008) | 0.83 [0.69; 0.99] |
| Nicaragua – (INIDE MINSA, 2014) | 0.75 [0.61; 0.91] |
| Peru – (INEI/Perú, 2012) | 0.90 [0.76; 1.07] |
| Peru – (INEI/Perú, 2013) | 0.96 [0.81; 1.14] |
| Peru – (INEI/Perú, 2014) | 0.91 [0.75; 1.10] |
| Peru – (INEI/Perú, 2015) | 1.01 [0.85; 1.20] |
| Peru – (INEI/Perú, 2016) | 0.92 [0.79; 1.08] |
| Peru – (INEI/Perú, 2017) | 0.92 [0.77; 1.09] |
| Peru – (INEI/Perú, 2018) | 1.02 [0.73; 1.40] |
| Paraguay – (Centro paraguayo de estudios de población, 2009) | 0.94 [0.71; 1.26] |
| El salvador – (Asociación Demográfica Salvadoreña – ADS/El Salvador, 2008) | 0.94 [0.74; 1.19] |
| Trinidad and Tobago – (Pemberton and Joseph, 2018) | 1.46 [0.68; 3.17] |
| **SuperRegion = North Africa & Middle East** | |
| Afghanistan – (CSO/Afghanistan, MPH/Afghanistan, and ICF, 2017) | 1.10 [1.03; 1.17] |
| Egypt – (MHP/Egypt and ICF International, 2015) | 1.07 [0.90; 1.27] |
| Egypt – (Duvuury et al., 2015) | 1.14 [1.03; 1.27] |
| Jordan – (DoS/Jordan, and Macro International, 2008) | 1.10 [0.73; 1.65] |
| Jordan – (DoS/Jordan, and ICF International, 2013) | 0.75 [0.55; 1.01] |
| Jordan – (DoS/Jordan, and ICF, 2019) | 0.84 [0.57; 1.23] |
| Turkey – (Ministry of Family and Social Policies, Hacettepe University and NEE Institute of Population Studies, 2009) | 1.03 [0.86; 1.25] |
| Turkey – (Ministry of Family and Social Policies, Hacettepe University and NEE Institute of Population Studies, 2015) | 0.91 [0.70; 1.18] |
| **SuperRegion = Sub–Saharan Africa** | |
| Angola – (INE/Angola, MINSA/Angola, and ICF, 2017) | 0.92 [0.79; 1.06] |
| Burundi – (MPBGP, MSPLS,– ISTEEBU/Burundi, and ICF, 2017) | 1.09 [0.96; 1.23] |
| Benin – (INSAE/Bénin, and ICF, 2019) | 1.07 [0.85; 1.33] |
| Central African Republic – (Institut Centrafricain des Statistiques, et des Etudes Economiques et Sociales, 2009) | 0.78 [0.69; 0.88] |
| Côte d'Ivoire – (INS/Côte d'Ivoire, and ICF International, 2013) | 0.85 [0.70; 1.03] |
| Cameroon – (INS/Cameroun, and ORC Macro, 2005) | 0.90 [0.70; 1.17] |
| Democratic Republic of the Congo – (MP/Congo, and Macro International, 2008) | 1.01 [0.82; 1.23] |
| Cabo verde – (ICIEG, INE/Cabo Verde, 2016) | 0.68 [0.37; 1.24] |
| Ethiopia – (Central Statistical Agency, 2017) | 1.11 [0.94; 1.32] |
| Gabon – (DGS/Gabon, and ICF International, 2013) | 1.24 [0.90; 1.71] |
| Ghana – (GSS, GHS, and Macro International, 2009) | 0.96 [0.69; 1.32] |
| Gambia – (GBOS, and ICF International, 2014) | 1.06 [0.74; 1.51] |
| Kenya – (CBS/Kenya, MOH/Kenya, and ORC Macro, 2004) | 1.11 [0.94; 1.30] |
| Kenya – (KNBS/Kenya, NACC/Kenya, NASCP/Kenya, MPHS/Kenya, and KMRI, 2010) | 1.04 [0.89; 1.21] |
| Kenya – (KNBS/Kenya, MOH/Kenya, NACC/Kenya, KMRI/Kenya, NCPD/Kenya, 2015) | 1.02 [0.85; 1.22] |
| Liberia – (LISGIS/Liberia, MOHSW/Liberia, NACP/Liberia, and Macro International, 2008) | 0.93 [0.78; 1.11] |
| Mali – (Samaké et al., 2007) | 1.06 [0.93; 1.20] |
| Mali – (CPS/SSDSPF/Mali, INSTAT/Mali, INFO–STAT/Mali, and ICF International, 2014) | 1.06 [0.87; 1.28] |
| Mali – (INSTAT/Mali, CPSSSD, and ICF, 2019) | 1.05 [0.86; 1.28] |
| Moçambique – (MISAU/Moçambique, INE/Moçambique, and ICF International, 2013) | 0.92 [0.80; 1.07] |
| Moçambique – (INS, INE, PNCM/Moçambique and ICF, 2019) | 0.83 [0.63; 1.08] |
| Malawi – (NSO/Malawi, and ORC Macro, 2005) | 1.04 [0.91; 1.18] |
| Namibia – (MoHSS/Namibia, and ICF International, 2014) | 1.07 [0.68; 1.68] |
| Nigeria – (NPC/Nigeria, and ICF, 2019) | 1.10 [0.94; 1.29] |
| Rwanda – (INSR/Rwanda, and ORC Macro, 2006) | 1.00 [0.81; 1.24] |
| Rwanda – (NISR/Rwanda, MOH/Rwanda, and ICF International, 2012) | 1.06 [0.90; 1.25] |
| Rwanda – (NISR/Rwanda, MFEP/Rwanda, MOH/Rwanda, and ICF International, 2016) | 1.03 [0.78; 1.35] |
| Senegal – (ANSD/Sénégal, and ICF, 2018) | 0.90 [0.65; 1.24] |
| Sierra Leone – (SSL/Sierra Leone, and ICF International, 2014) | 0.92 [0.78; 1.09] |
| Sao Tome and Principe – (INE/São Tomé, and ICF Macro, 2010) | 0.87 [0.63; 1.20] |
| Chad – (INSEED/Tchad, MSP/Tchad, and ICF International, 2016) | 0.98 [0.80; 1.21] |
| Togo – (MPDAT/Togo, MS/Togo, and ICF International, 2015) | 1.07 [0.86; 1.32] |
| Tanzania – (NBS/Tanzania, and ICF Macro, 2011) | 1.04 [0.91; 1.19] |
| Tanzania – (MoHCDGEC/Tanzania, MoH/Zanzibar, NBS/Tanzania, OCGS/Zanzibar, and ICF, 2016) | 1.05 [0.93; 1.20] |
| Uganda – (UBOS/Uganda, and Macro International, 2007) | 1.09 [0.87; 1.37] |
| Uganda – (Ministry of Health/Uganda, and ICF International, 2012) | 1.16 [0.91; 1.49] |
| Uganda – (Uganda Bureau of Statistics, 2018) | 1.11 [0.98; 1.26] |
| Zambia – (CSO/Zambia, MOH/Zambia, TDRC/Zambia, and University of Zambia, 2009) | 0.85 [0.72; 1.00] |
| Zambia – (CSO/Zambia, MOH/Zambia, University of Zambia THVL, DPS, TDRC/Zambia, and ICF International, 2015) | 1.03 [0.91; 1.17] |
| Zimbabwe – (CSO/Zimbabwe, and Macro International, 2007) | 1.11 [0.94; 1.30] |
| Zimbabwe – (ZNSA/Zimbabwe, and ICF International, 2016) | 1.01 [0.85; 1.19] |

Heterogeneity: $\chi^2_{101} = 145.79$ ($P = .002$), $I^2 = 31\%$

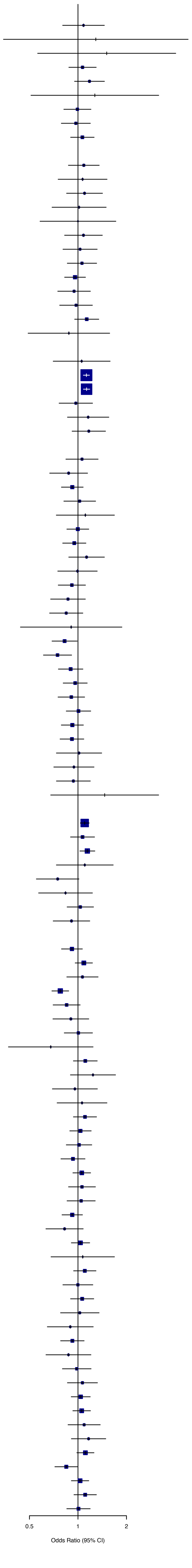

**Figure S16.** Posterior predictive checks for the lifetime intimate partner violence (IPV) model.

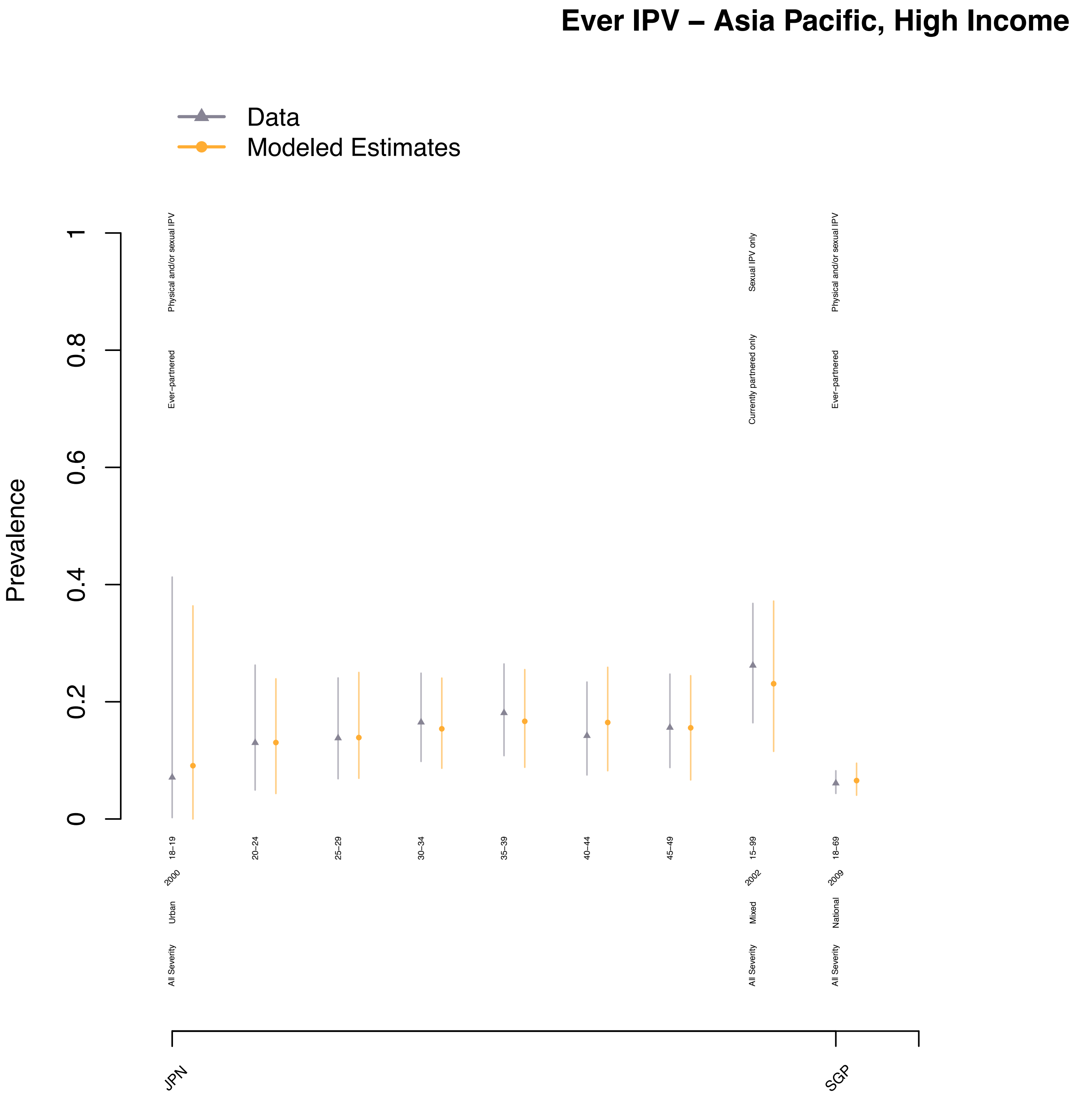

Ever IPV – Asia Pacific, High Income
Data
Modeled Estimates
Prevalence
0
0.2
0.4
0.6
0.8
1
Physical and/or sexual IPV
Ever−partnered
Sexual IPV only
Currently partnered only
Physical and/or sexual IPV
Ever−partnered
18−19
20−24
25−29
30−34
35−39
40−44
45−49
15−99
18−69
2000
2002
2009
Urban
Mixed
National
All Severity
All Severity
All Severity
JPN
SGP

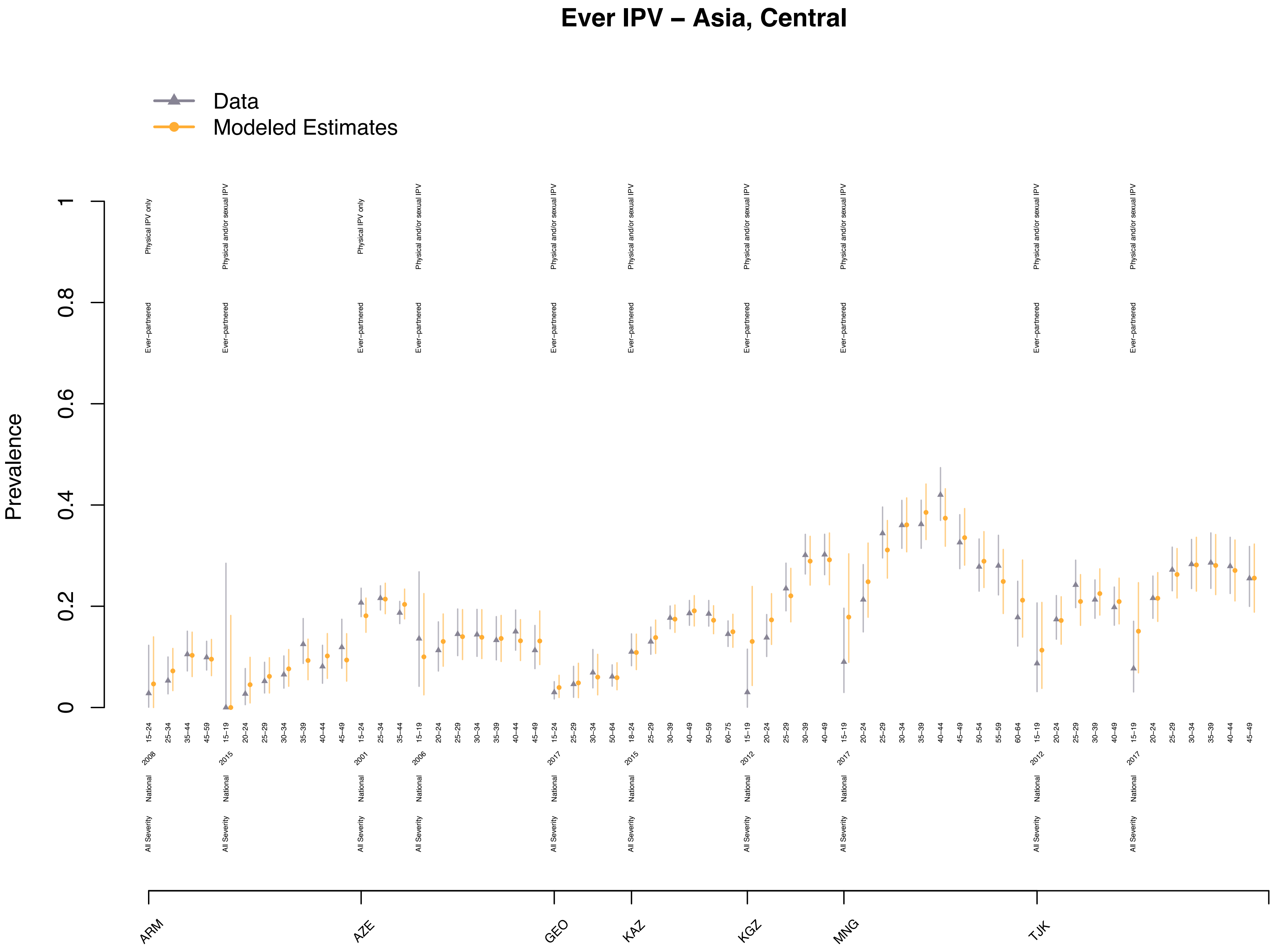
Ever IPV – Asia, Central
Data
Modeled Estimates
Prevalence
0
0.2
0.4
0.6
0.8
1
Physical IPV only
Physical and/or sexual IPV
Ever−partnered
National
All Severity
ARM
AZE
GEO
KAZ
KGZ
MNG
TJK
2008
2015
2001
2006
2017
2015
2012
2017
2012
2017

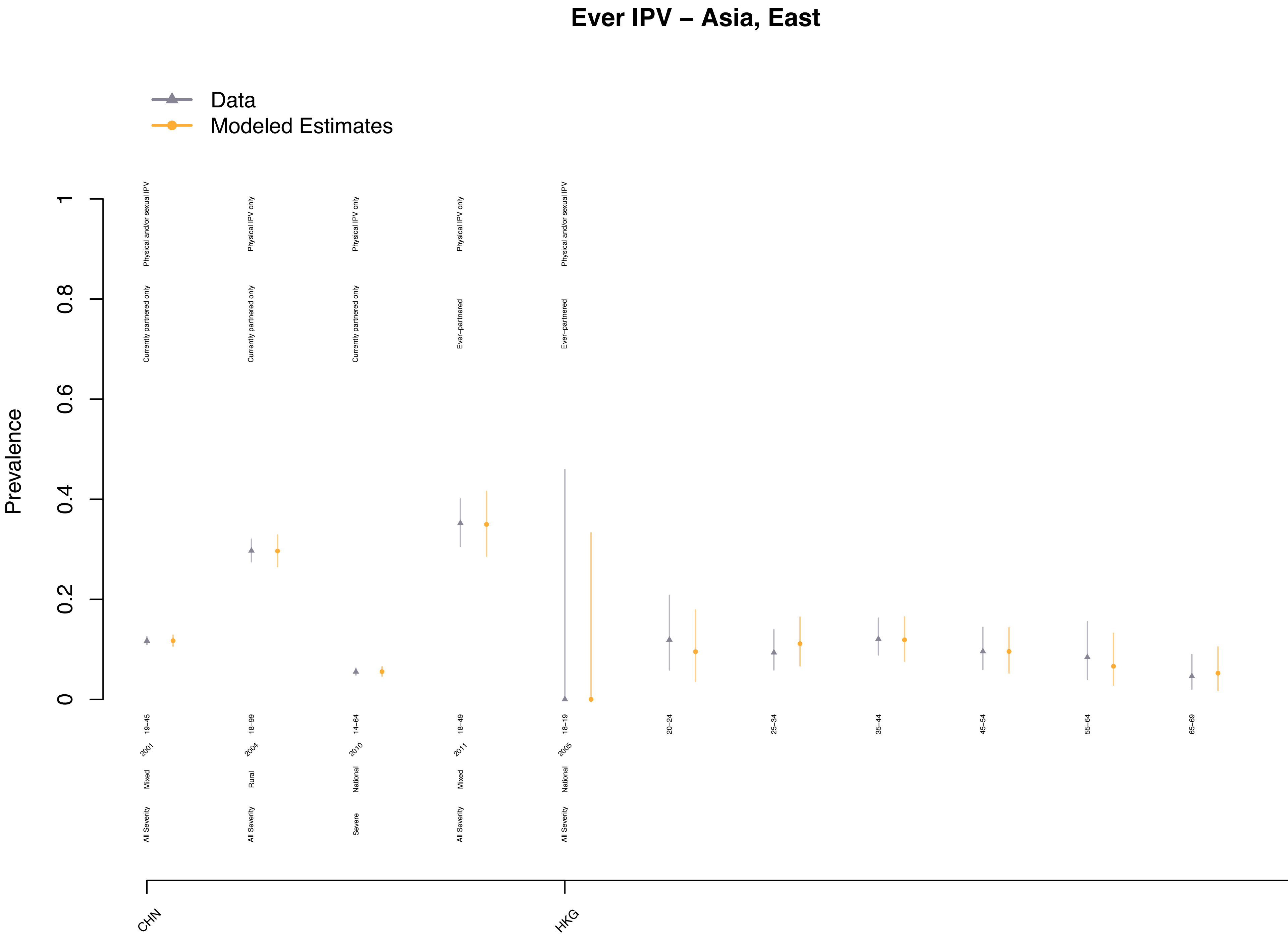
Ever IPV – Asia, East
Data
Modeled Estimates
Prevalence
0
0.2
0.4
0.6
0.8
1
Physical and/or sexual IPV
Currently partnered only
Physical IPV only
Currently partnered only
Physical IPV only
Currently partnered only
Physical IPV only
Ever−partnered
Physical and/or sexual IPV
Ever−partnered
19−45
2001
Mixed
All Severity
18−99
2004
Rural
All Severity
14−64
2010
National
Severe
18−49
2011
Mixed
All Severity
18−19
2005
National
All Severity
20−24
25−34
35−44
45−54
55−64
65−69
CHN
HKG

### Ever IPV – Asia, South

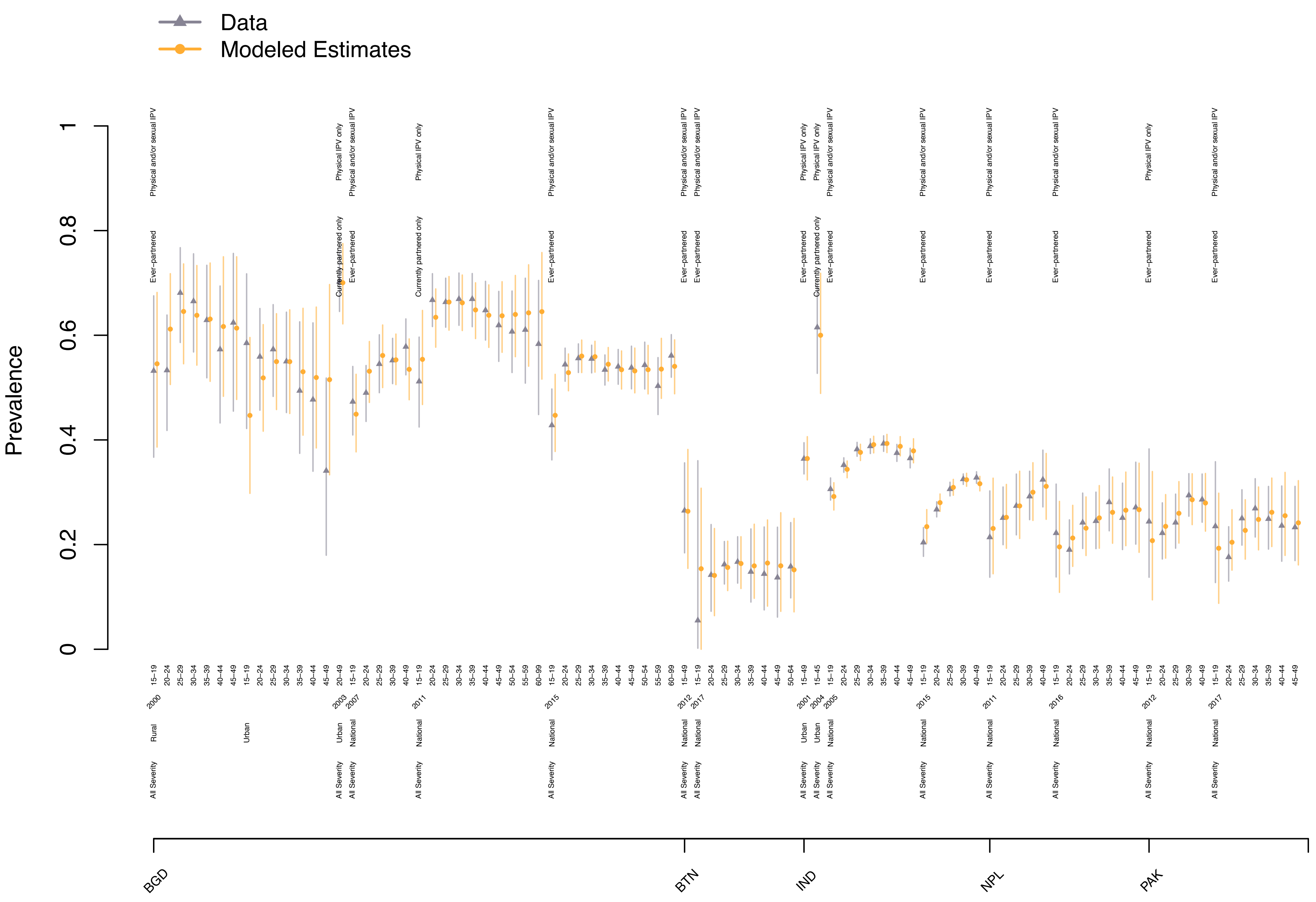

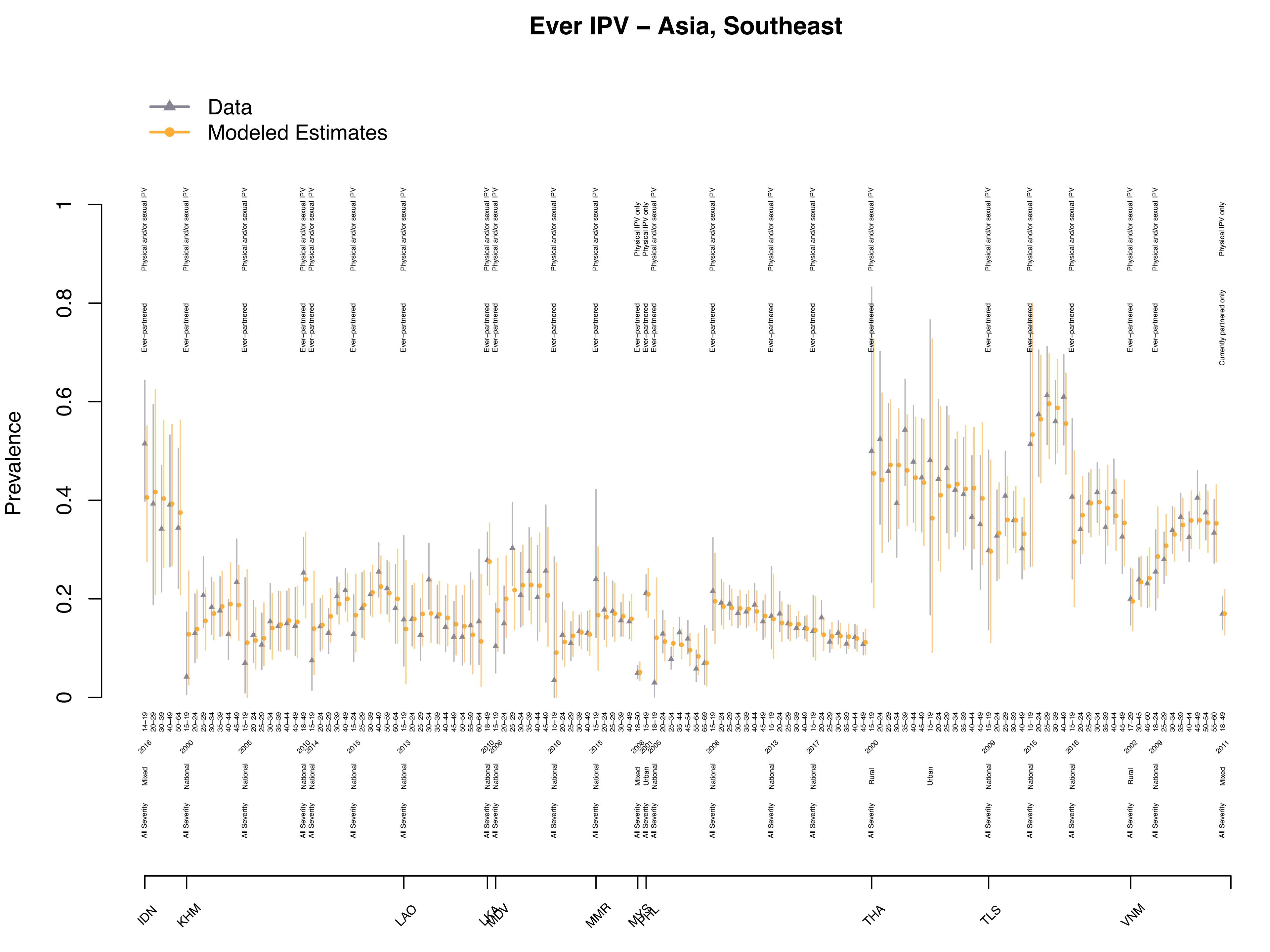
Ever IPV – Asia, Southeast
Data
Modeled Estimates
Prevalence
IDN
KHM
LAO
LKA
MDV
MMR
MYS
PHL
THA
TLS
VNM

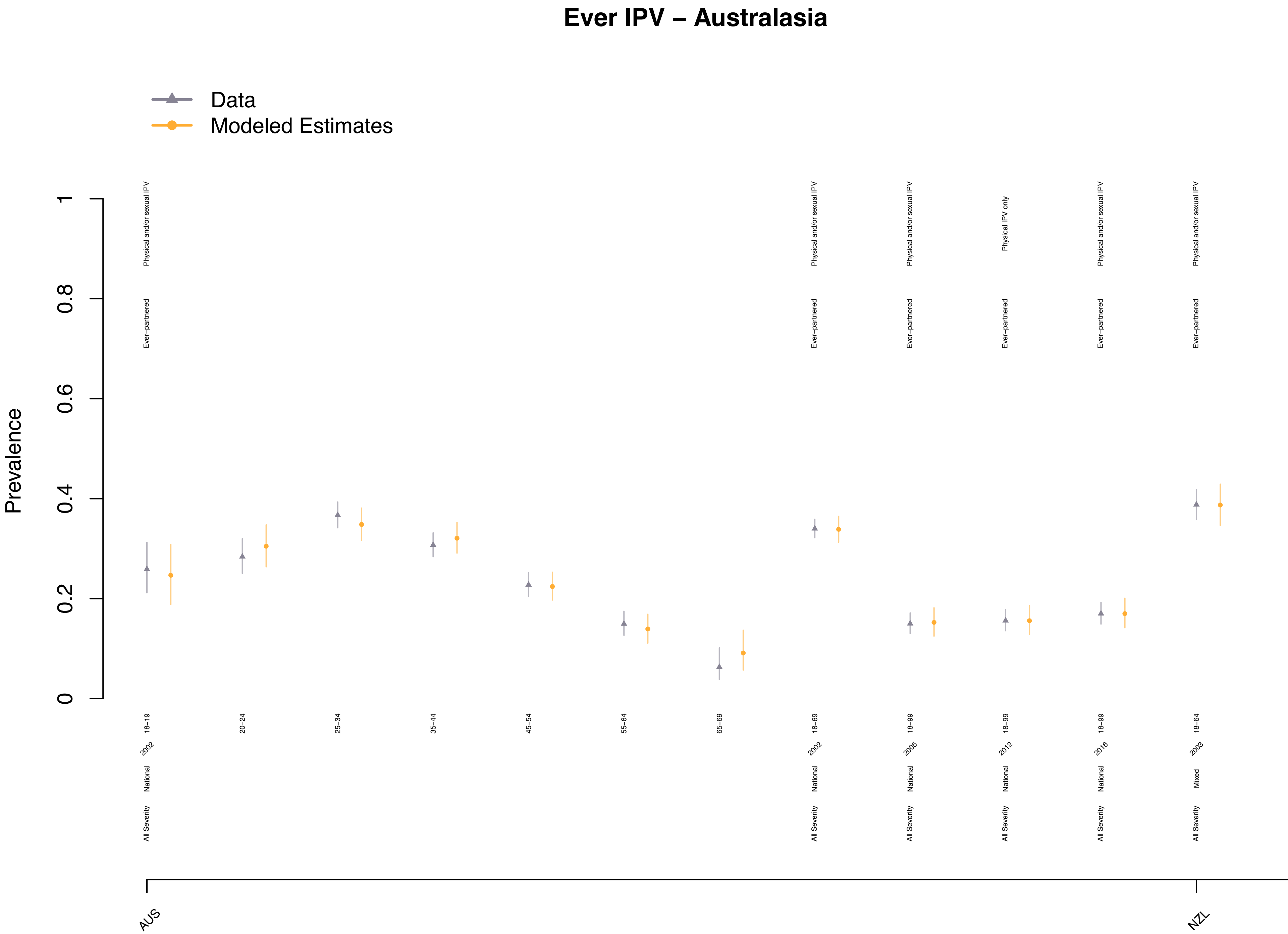

Ever IPV – Australasia
Data
Modeled Estimates
Prevalence
0
0.2
0.4
0.6
0.8
1
Physical and/or sexual IPV
Ever−partnered
Physical IPV only
18−19
20−24
25−34
35−44
45−54
55−64
65−69
18−69
18−99
18−64
2002
2005
2012
2016
2003
National
Mixed
All Severity
AUS
NZL

#### Ever IPV – Caribbean

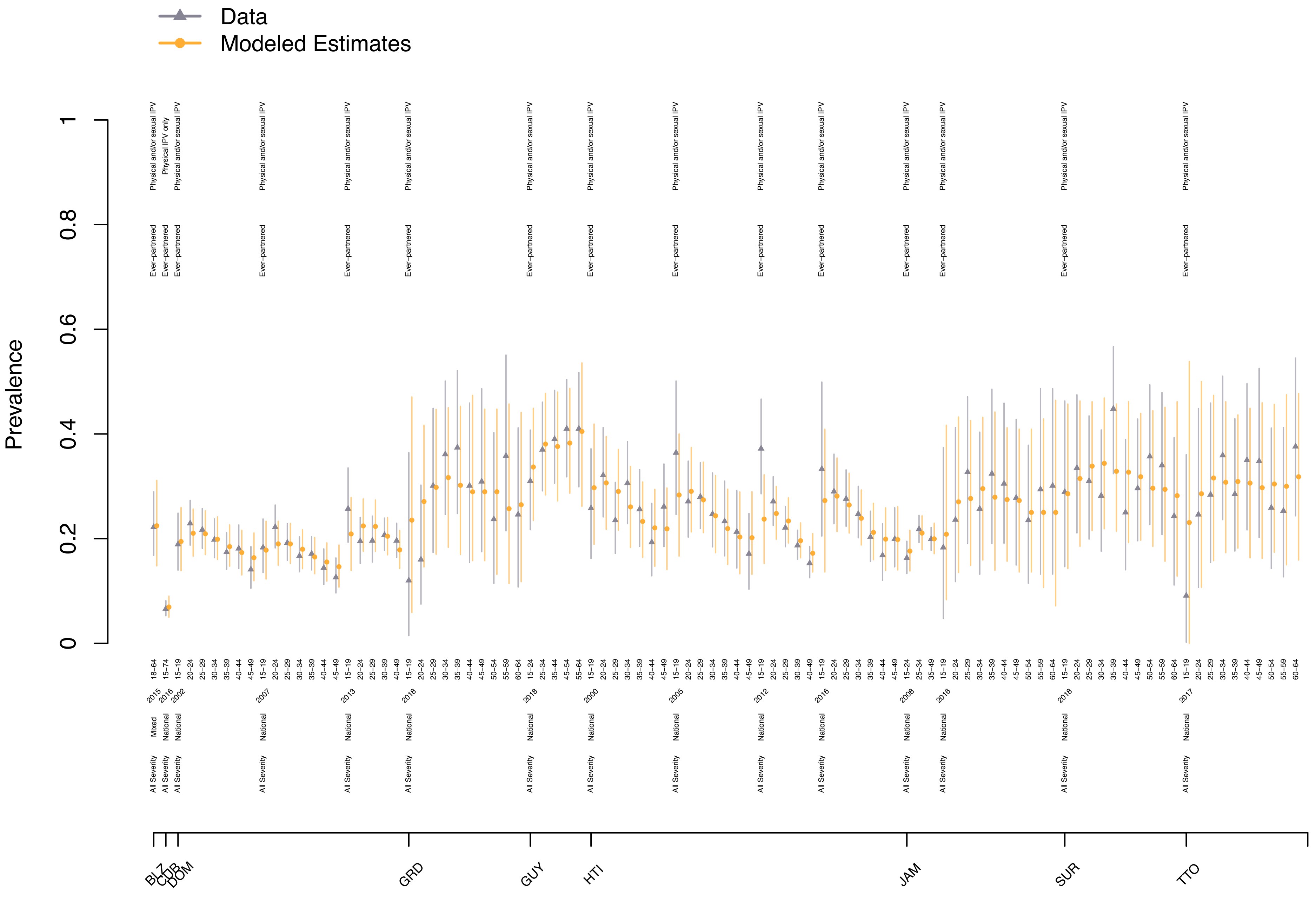

### Ever IPV – Europe, Central

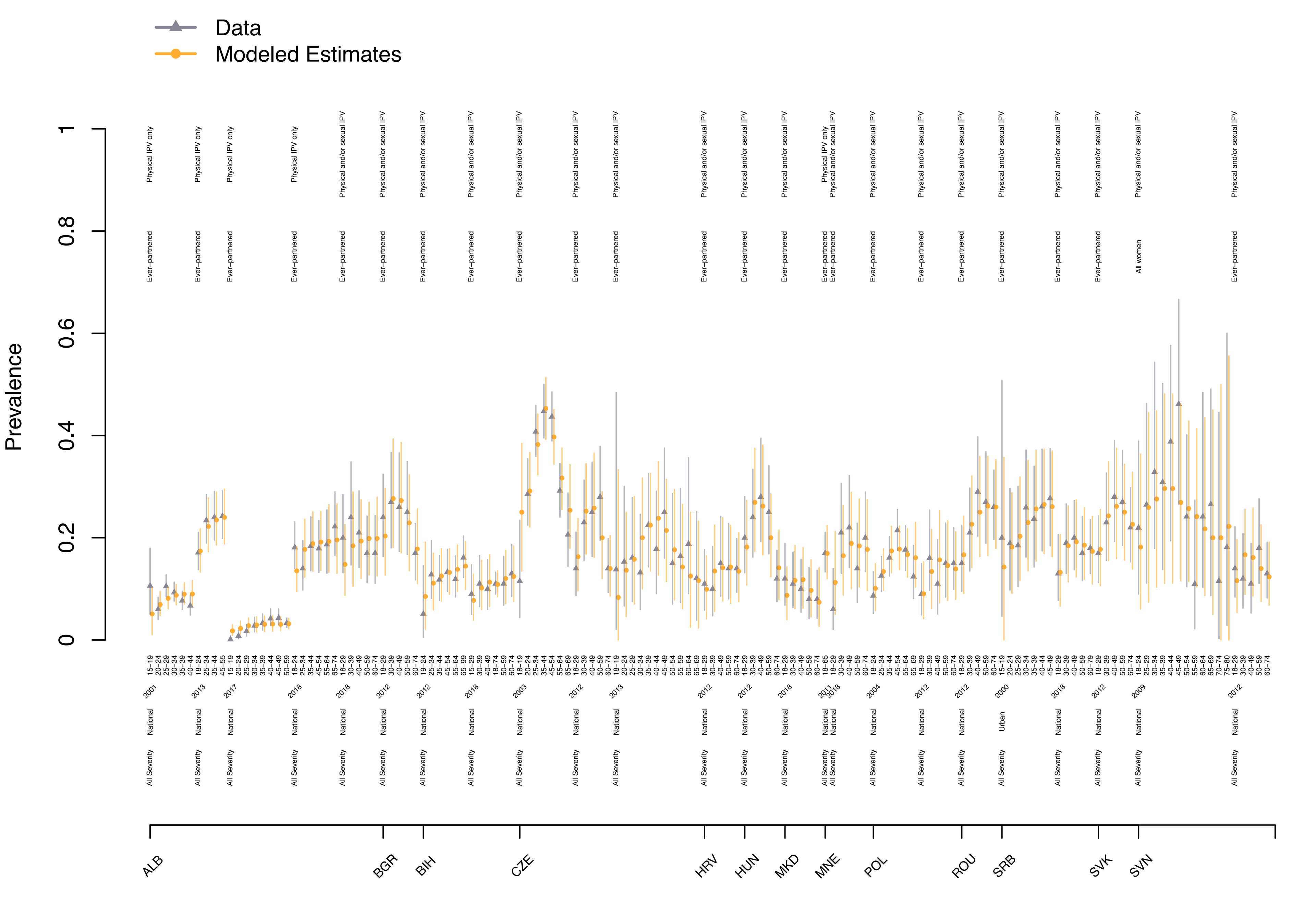

### Ever IPV – Europe, Eastern

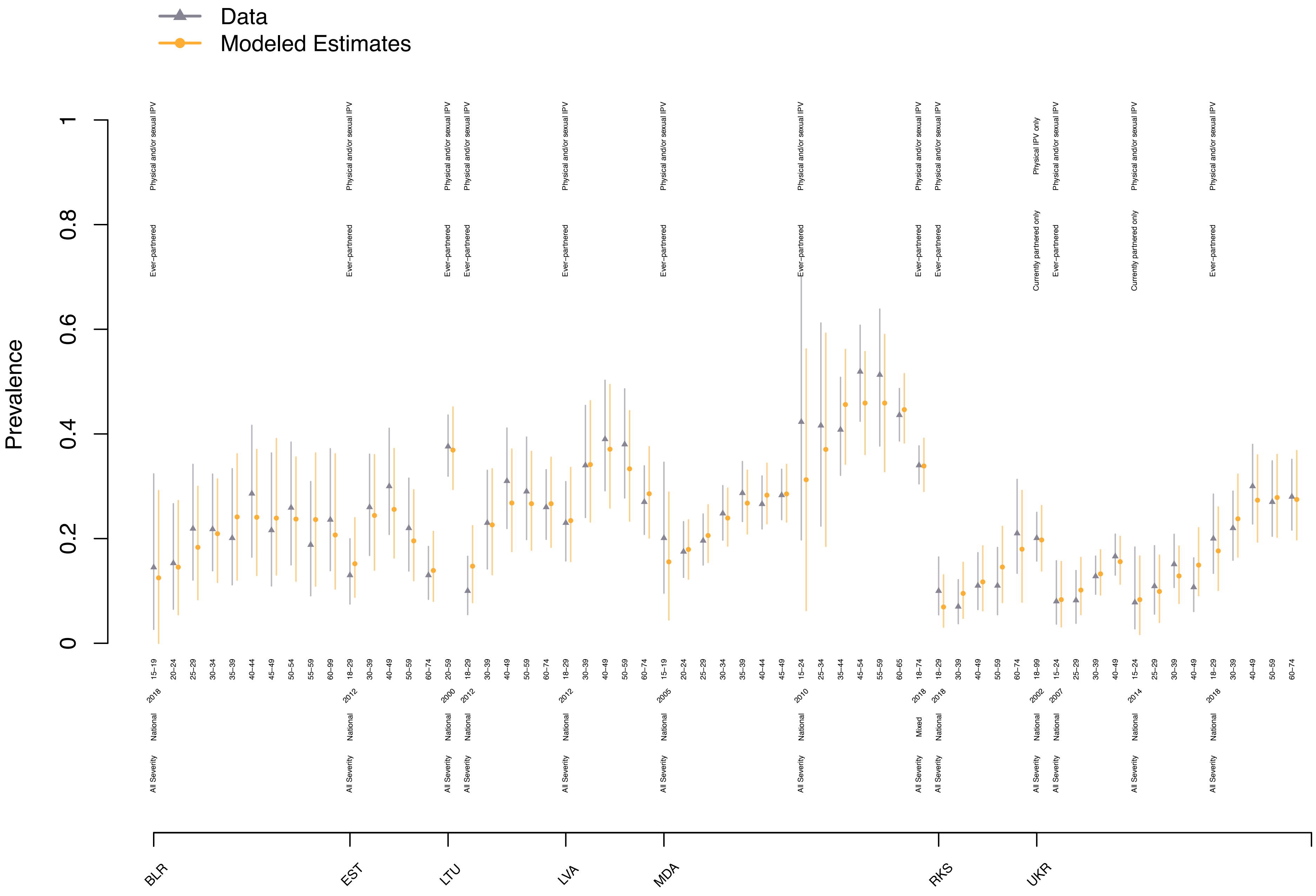

### Ever IPV – Europe, Western

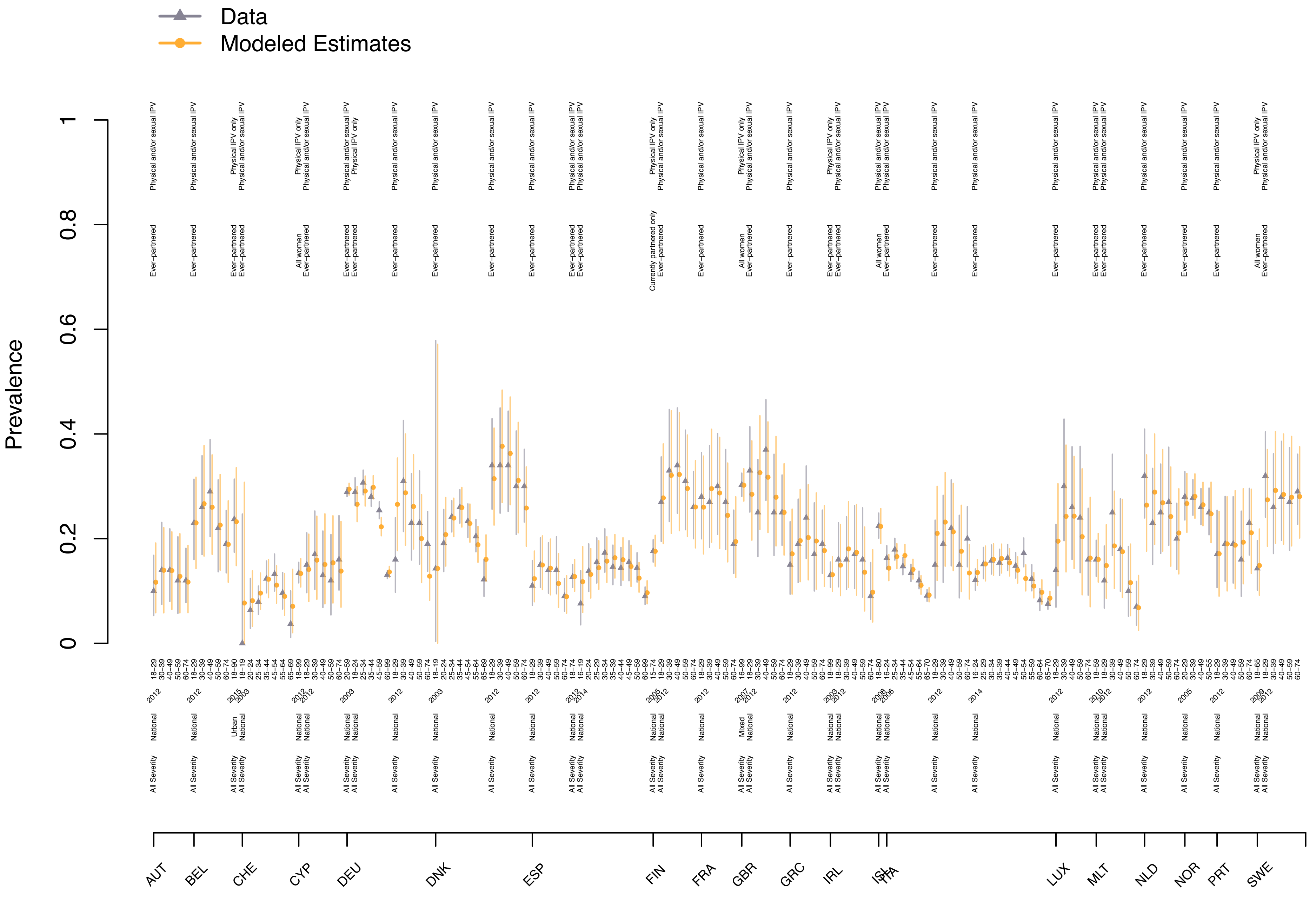

### Ever IPV – Latin America, Andean

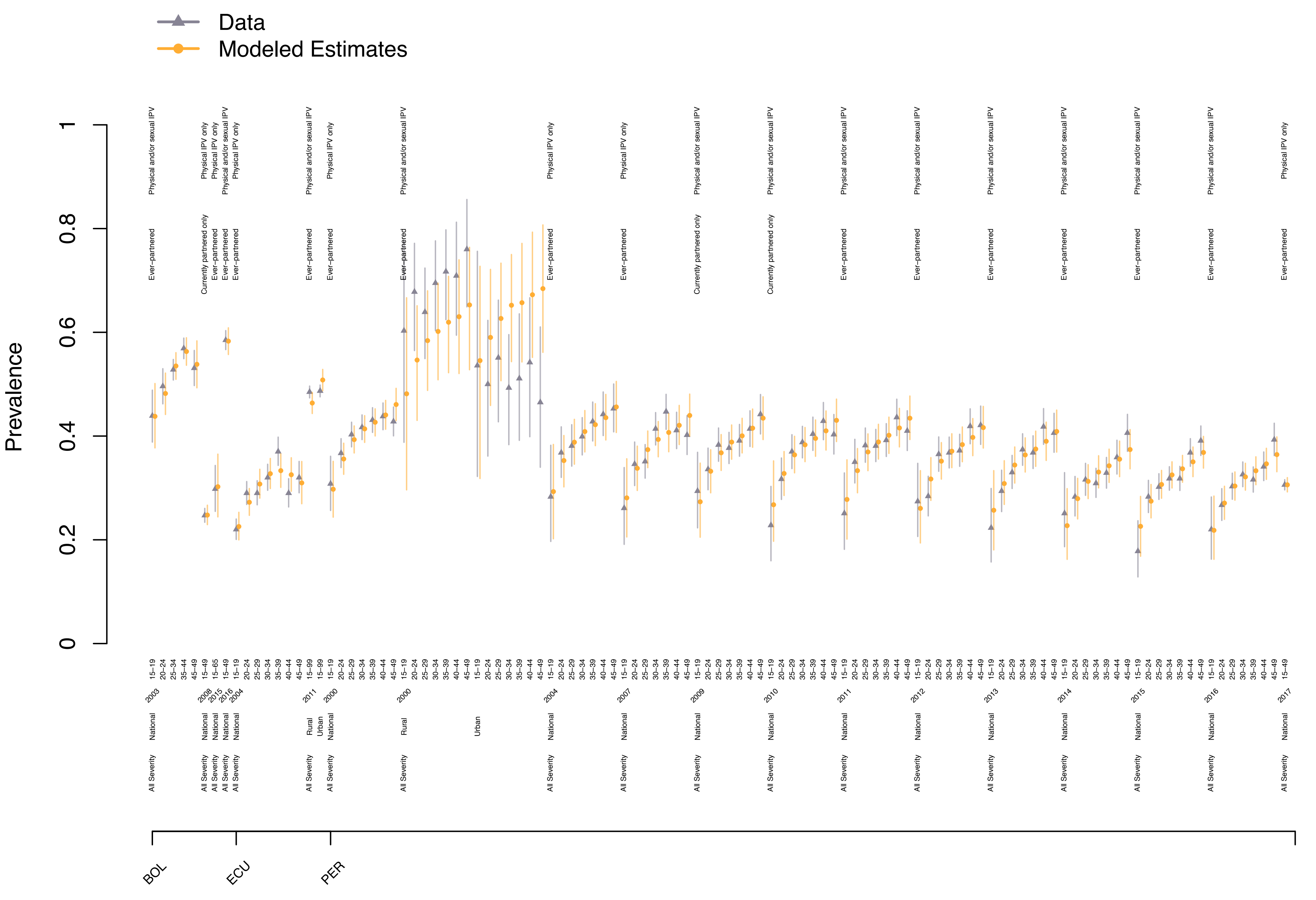

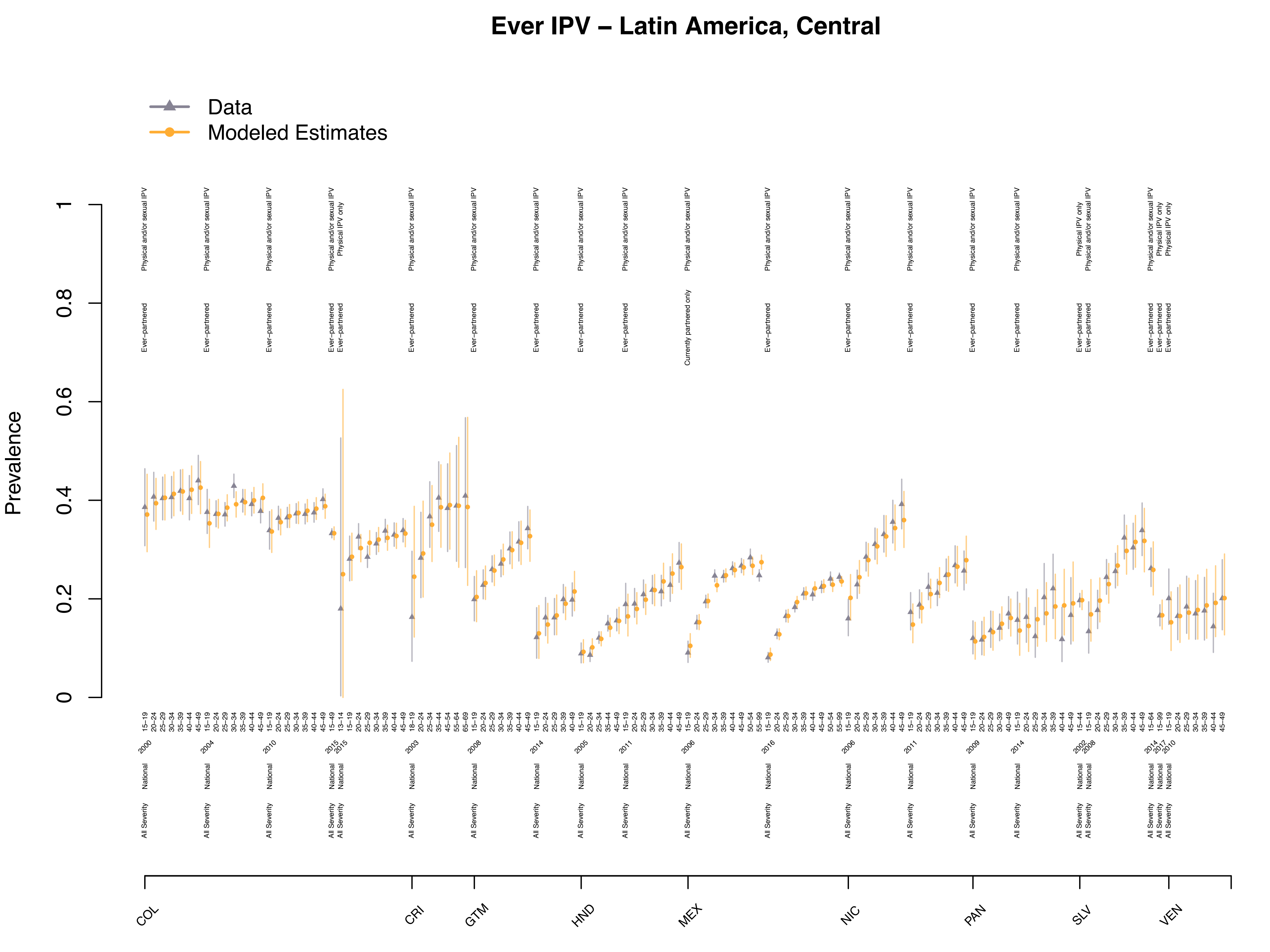
Ever IPV – Latin America, Central
Data
Modeled Estimates
Prevalence
COL
CRI
GTM
HND
MEX
NIC
PAN
SLV
VEN

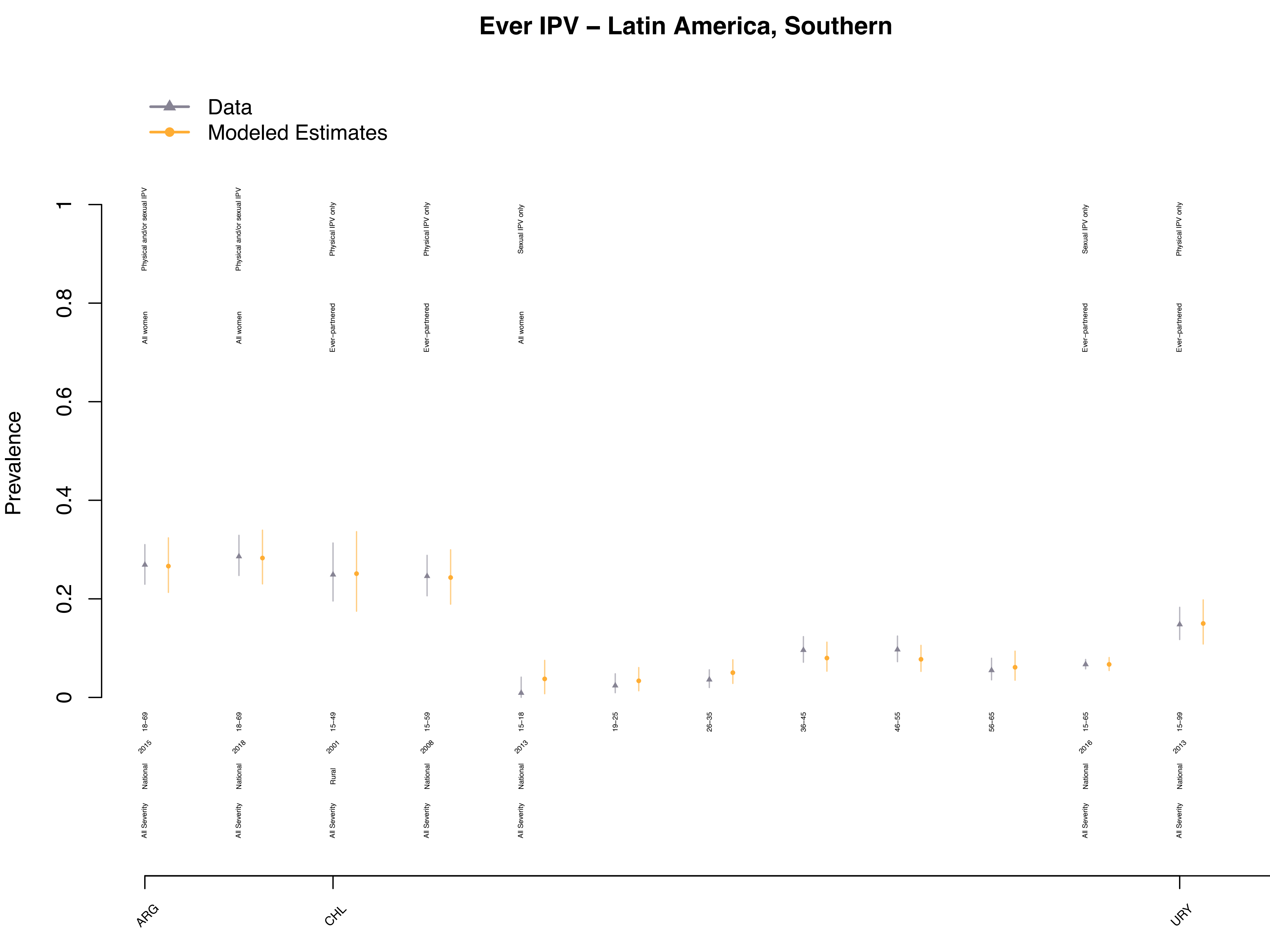

Ever IPV – Latin America, Southern
Data
Modeled Estimates
Prevalence
0
0.2
0.4
0.6
0.8
1
Physical and/or sexual IPV
All women
18–69
2015
National
All Severity
Physical and/or sexual IPV
All women
18–69
2018
National
All Severity
Physical IPV only
Ever–partnered
15–49
2001
Rural
All Severity
Physical IPV only
Ever–partnered
15–59
2008
National
All Severity
Sexual IPV only
All women
15–18
2013
National
All Severity
19–25
26–35
36–45
46–55
56–65
Sexual IPV only
Ever–partnered
15–65
2016
National
All Severity
Physical IPV only
Ever–partnered
15–99
2013
National
All Severity
ARG
CHL
URY

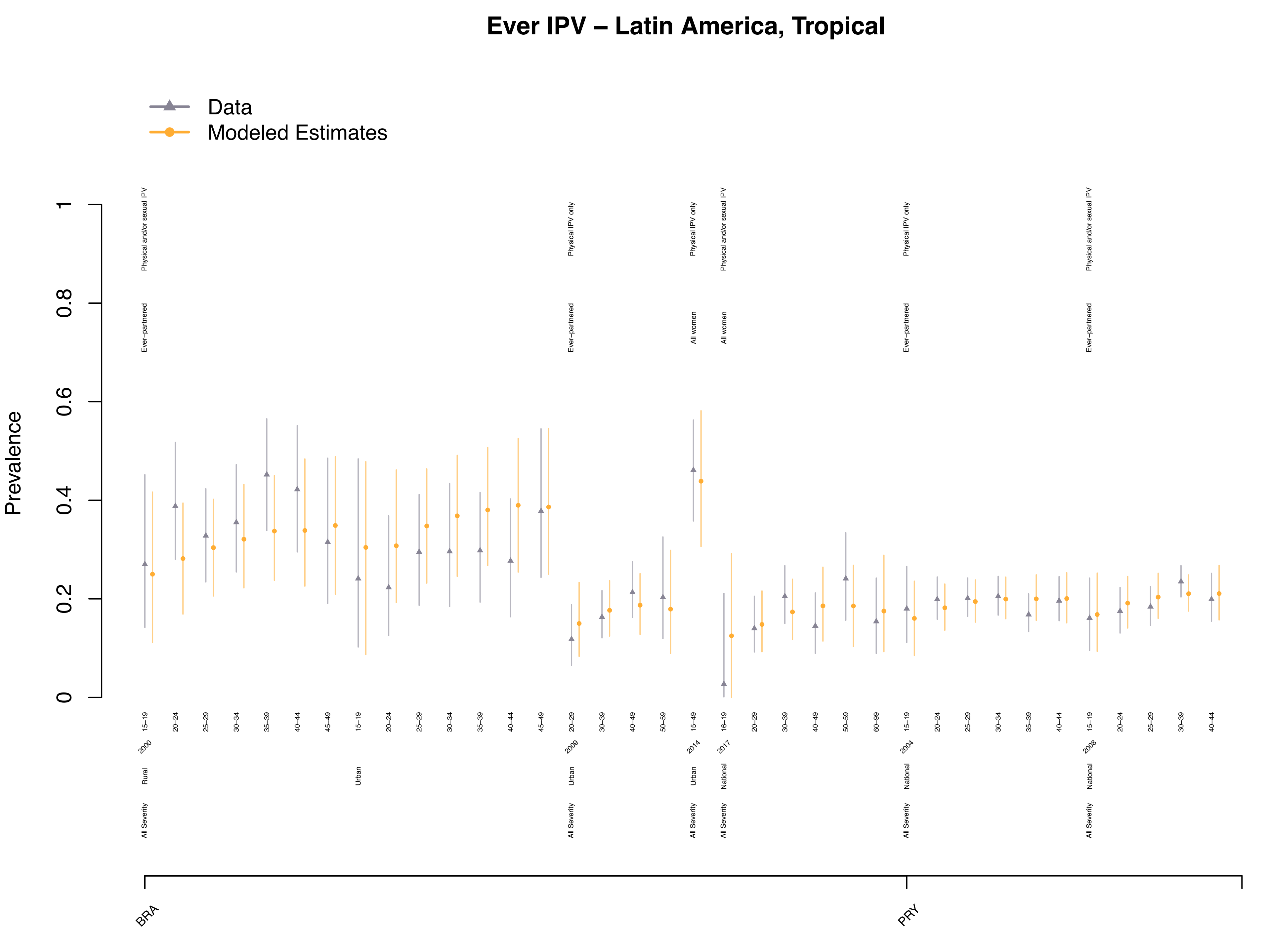

Ever IPV – Latin America, Tropical
Data
Modeled Estimates
Prevalence
BRA
PRY

### Ever IPV – North Africa/Middle East

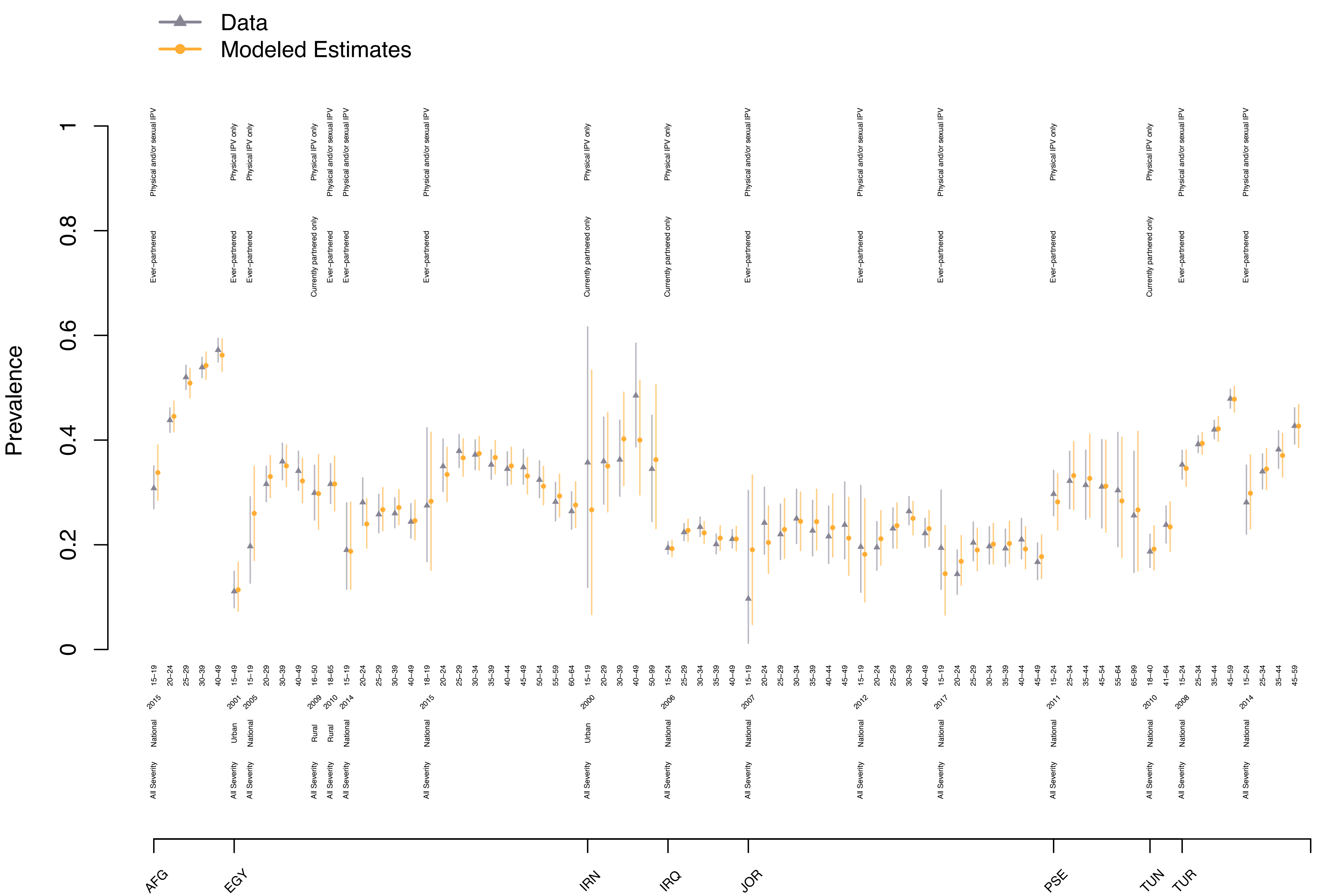

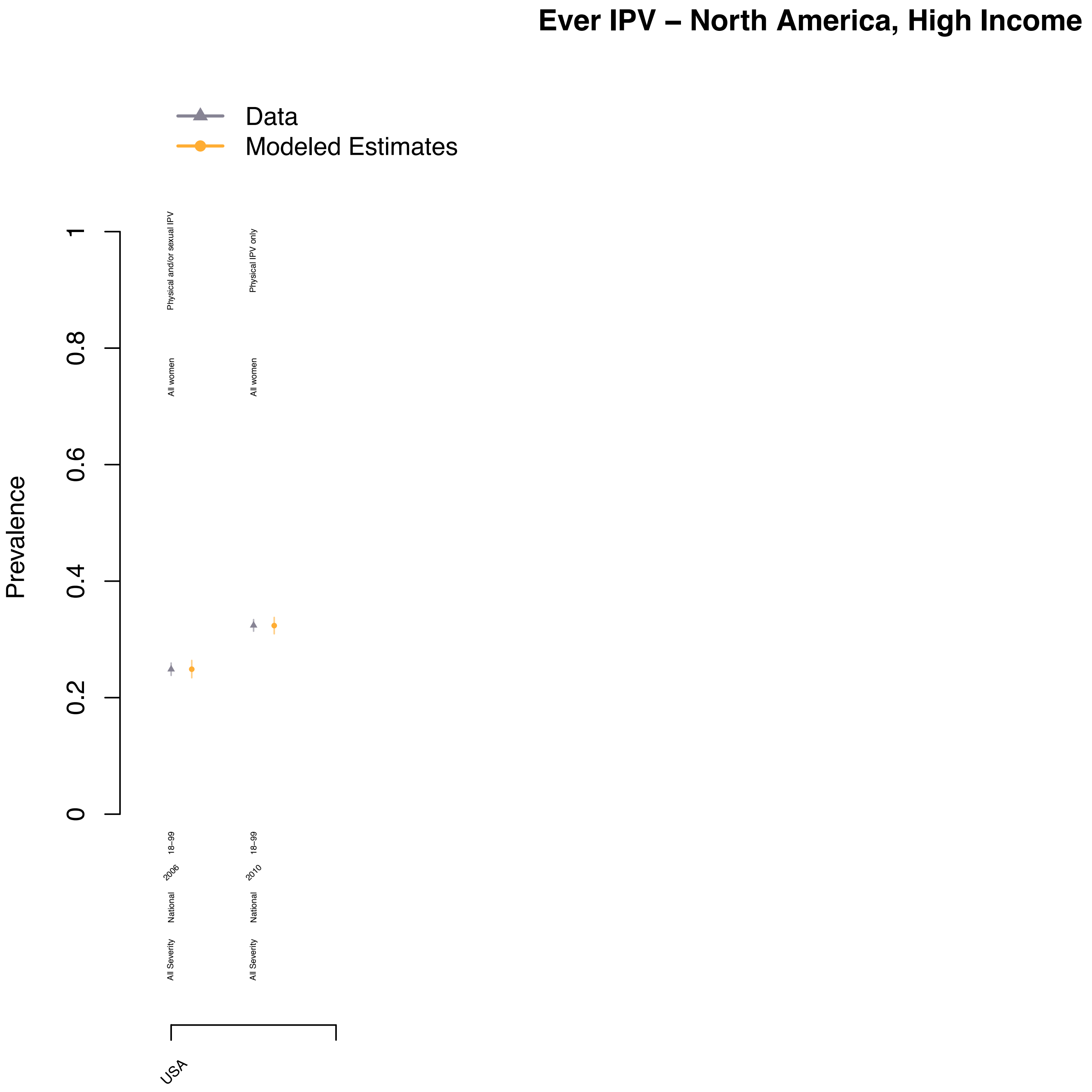
Ever IPV – North America, High Income
Data
Modeled Estimates
Prevalence
0
0.2
0.4
0.6
0.8
1
Physical and/or sexual IPV
All women
Physical IPV only
All women
18–99
2006
National
All Severity
18–99
2010
National
All Severity
USA

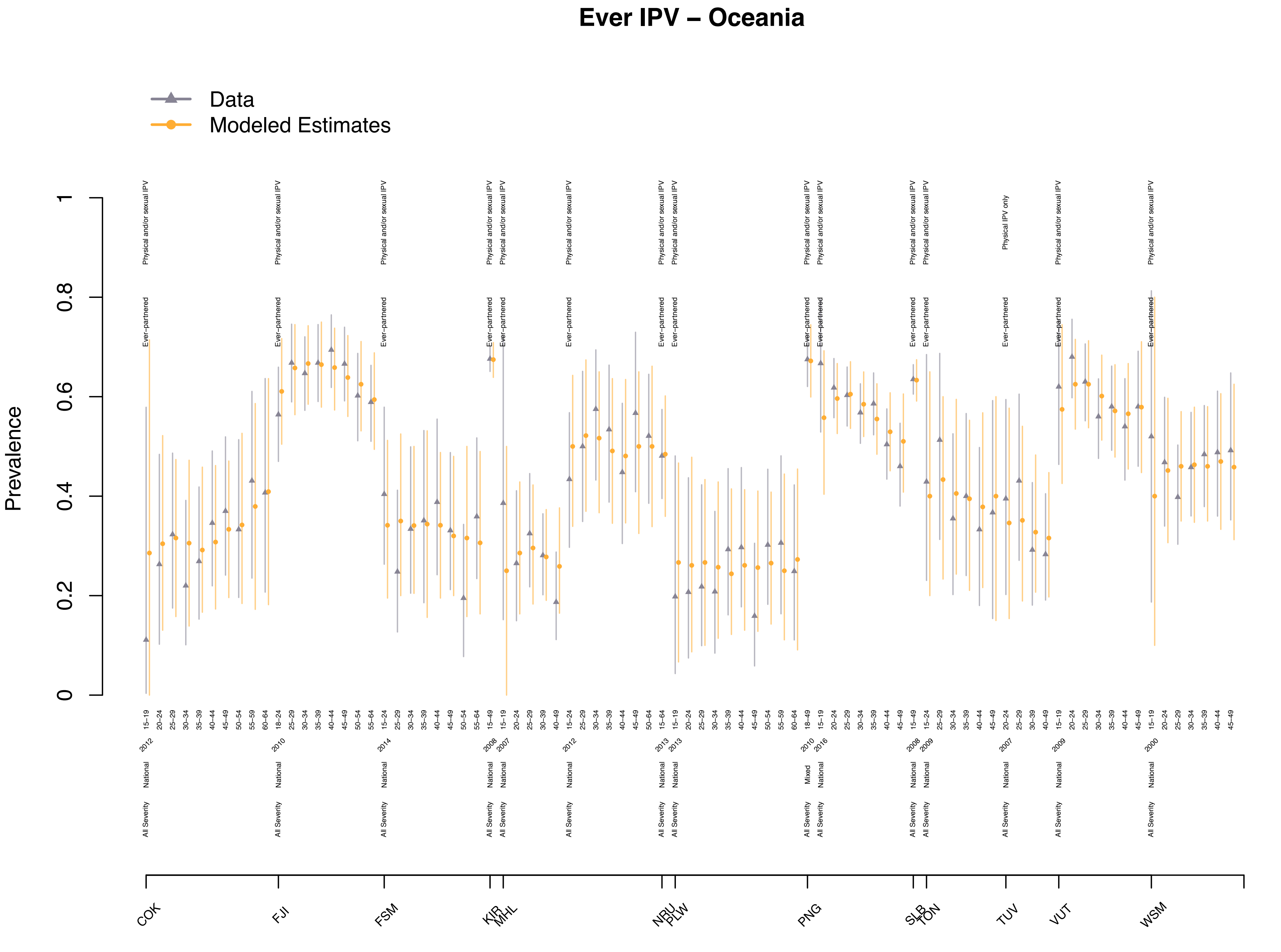

Ever IPV – Oceania
Data
Modeled Estimates
Prevalence
COK
FJI
FSM
KIR
MHL
NRU
PLW
PNG
SLB
TON
TUV
VUT
WSM

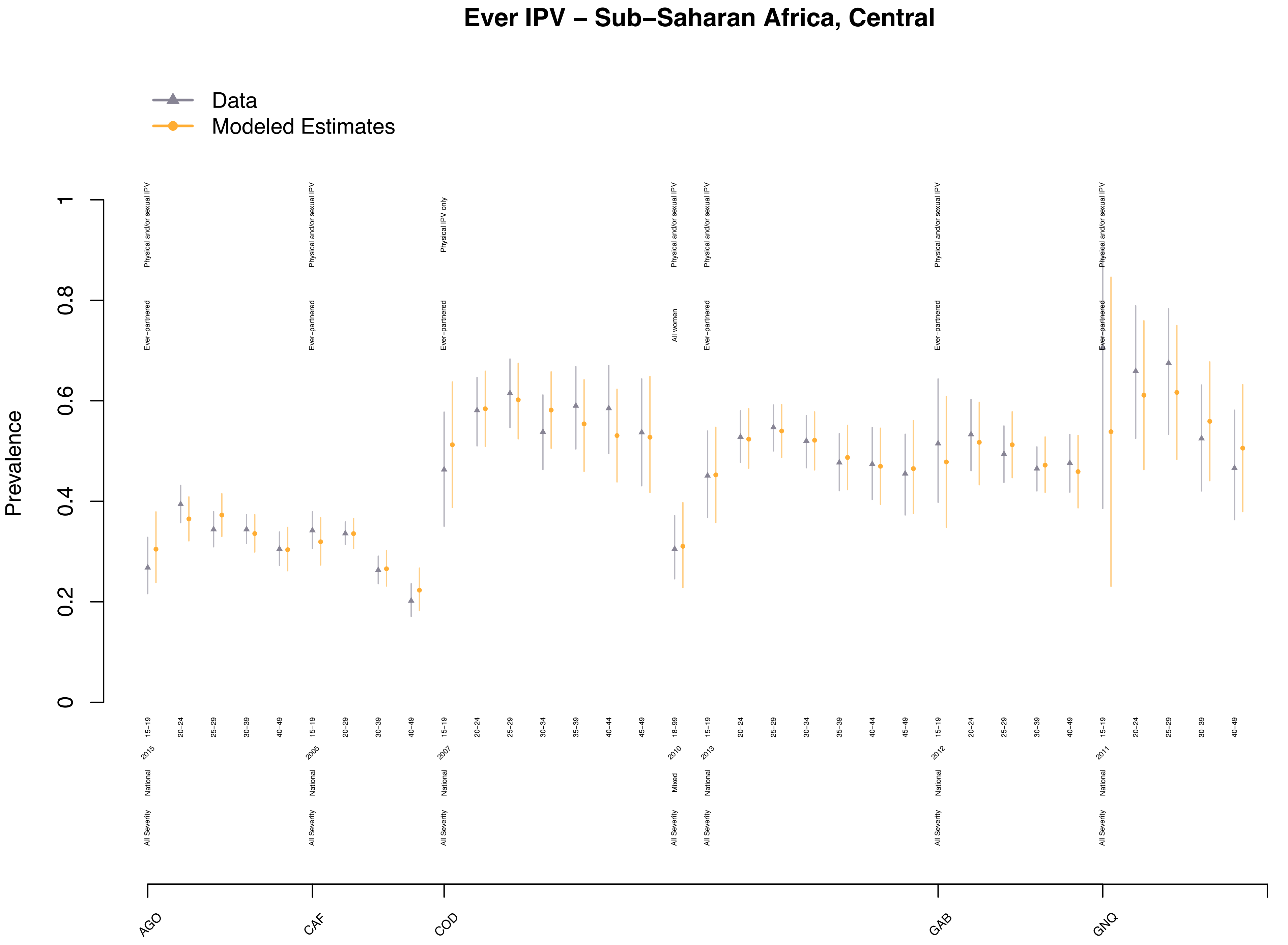

Ever IPV – Sub–Saharan Africa, Central
Data
Modeled Estimates
Prevalence
0
0.2
0.4
0.6
0.8
1
Physical and/or sexual IPV
Physical IPV only
Ever–partnered
All women
15–19
20–24
25–29
30–39
40–49
30–34
35–39
40–44
45–49
18–99
20–29
2015
2005
2007
2010
2013
2012
2011
National
Mixed
All Severity
AGO
CAF
COD
GAB
GNQ

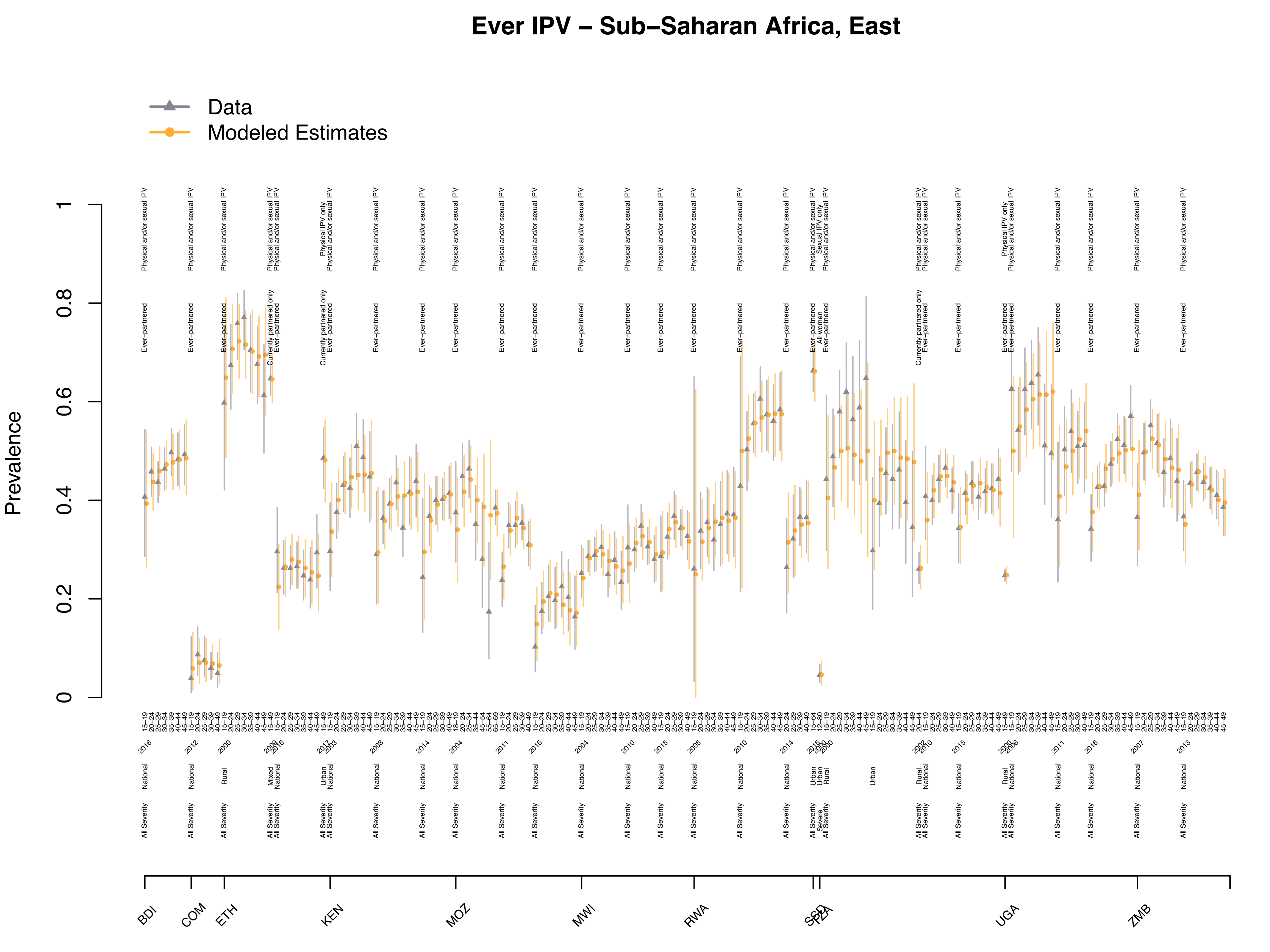

Ever IPV – Sub–Saharan Africa, East
Data
Modeled Estimates
Prevalence
BDI
COM
ETH
KEN
MOZ
MWI
RWA
SSD
TZA
UGA
ZMB

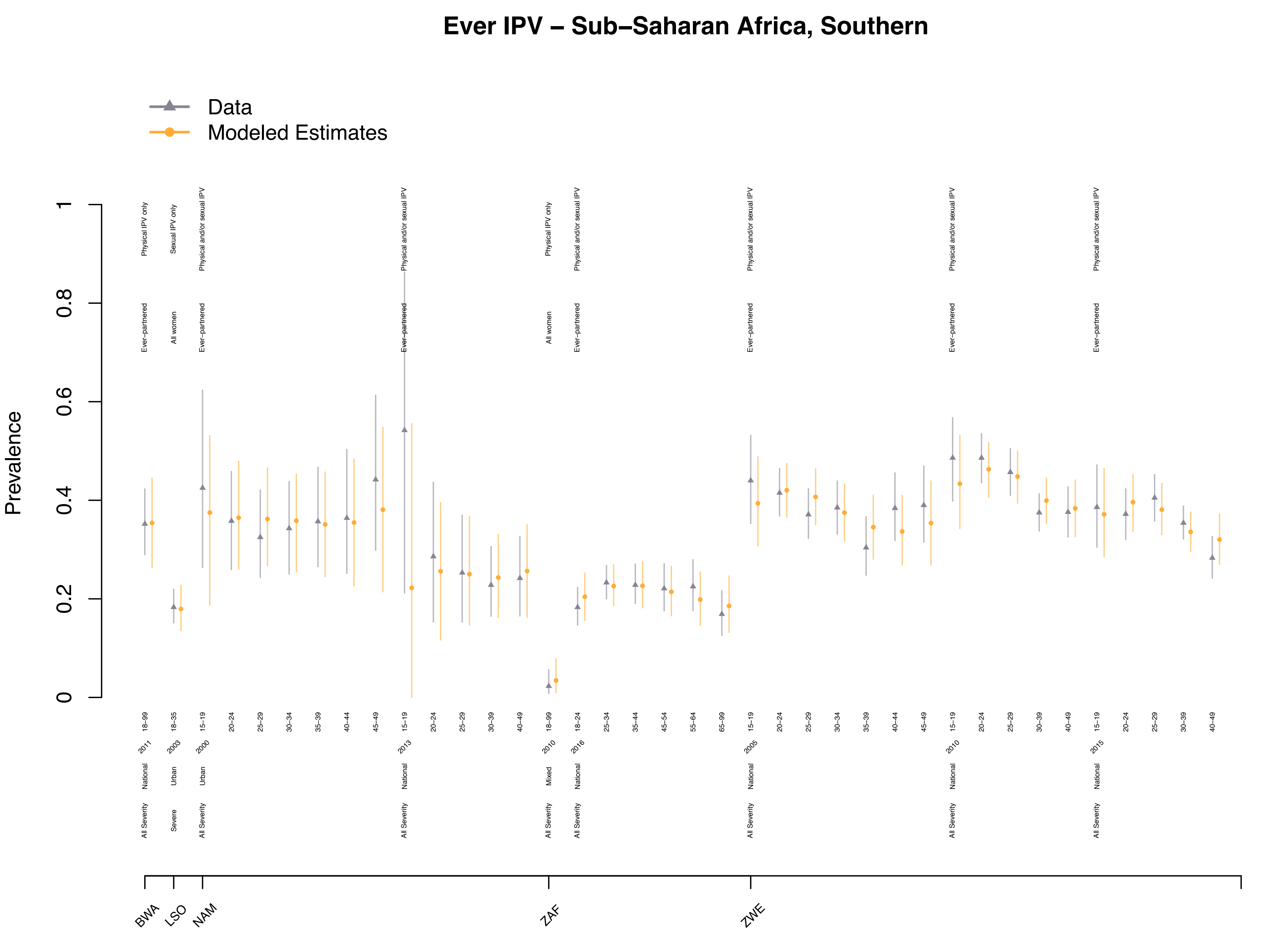
Ever IPV – Sub–Saharan Africa, Southern
Data
Modeled Estimates
Prevalence
0
0.2
0.4
0.6
0.8
1
Physical IPV only
Sexual IPV only
Physical and/or sexual IPV
Ever–partnered
All women
All Severity
Severe
National
Urban
Mixed
2011
2003
2000
2013
2010
2016
2005
2015
18–99
18–35
15–19
20–24
25–29
30–34
35–39
40–44
45–49
30–39
40–49
18–24
25–34
35–44
45–54
55–64
65–99
BWA
LSO
NAM
ZAF
ZWE

### Ever IPV – Sub–Saharan Africa, West

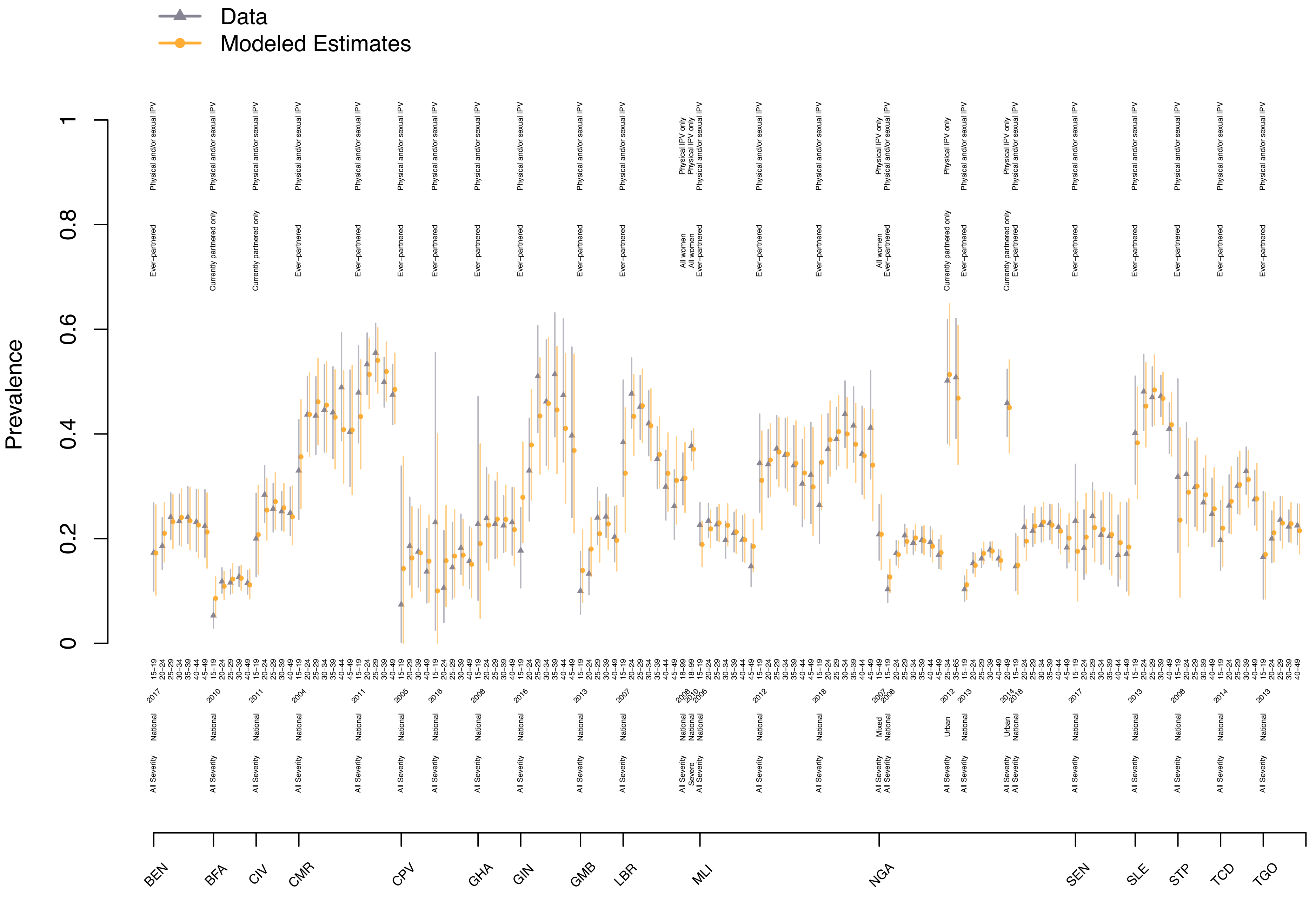

**Figure S17.** Posterior predictive checks for the past year intimate partner violence (IPV) model.

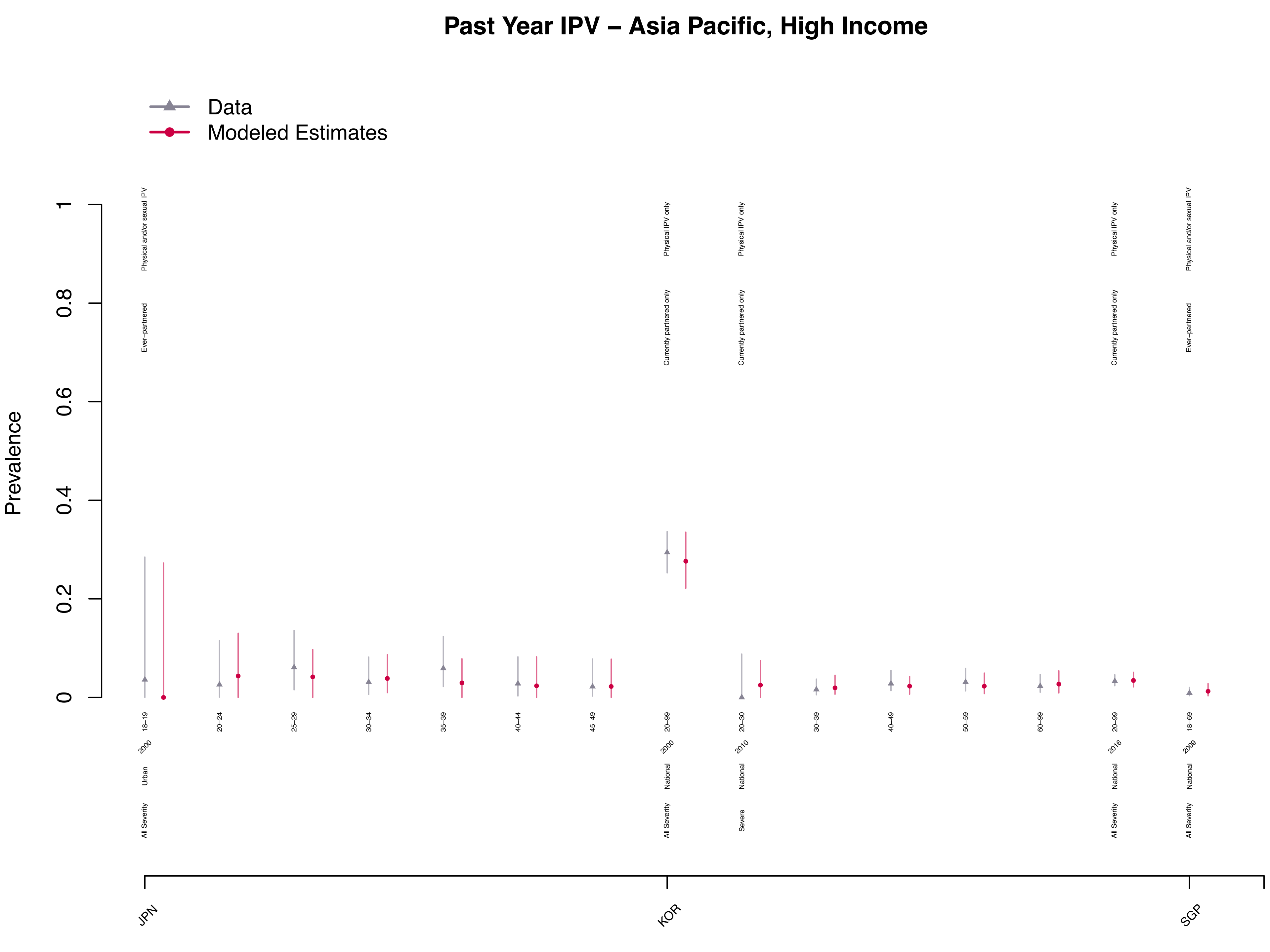

Past Year IPV – Asia Pacific, High Income
Data
Modeled Estimates
Prevalence
0
0.2
0.4
0.6
0.8
1
Physical and/or sexual IPV
Ever−partnered
Physical IPV only
Currently partnered only
18−19
20−24
25−29
30−34
35−39
40−44
45−49
20−99
20−30
30−39
40−49
50−59
60−99
18−69
2000
2010
2016
2009
Urban
National
All Severity
Severe
JPN
KOR
SGP

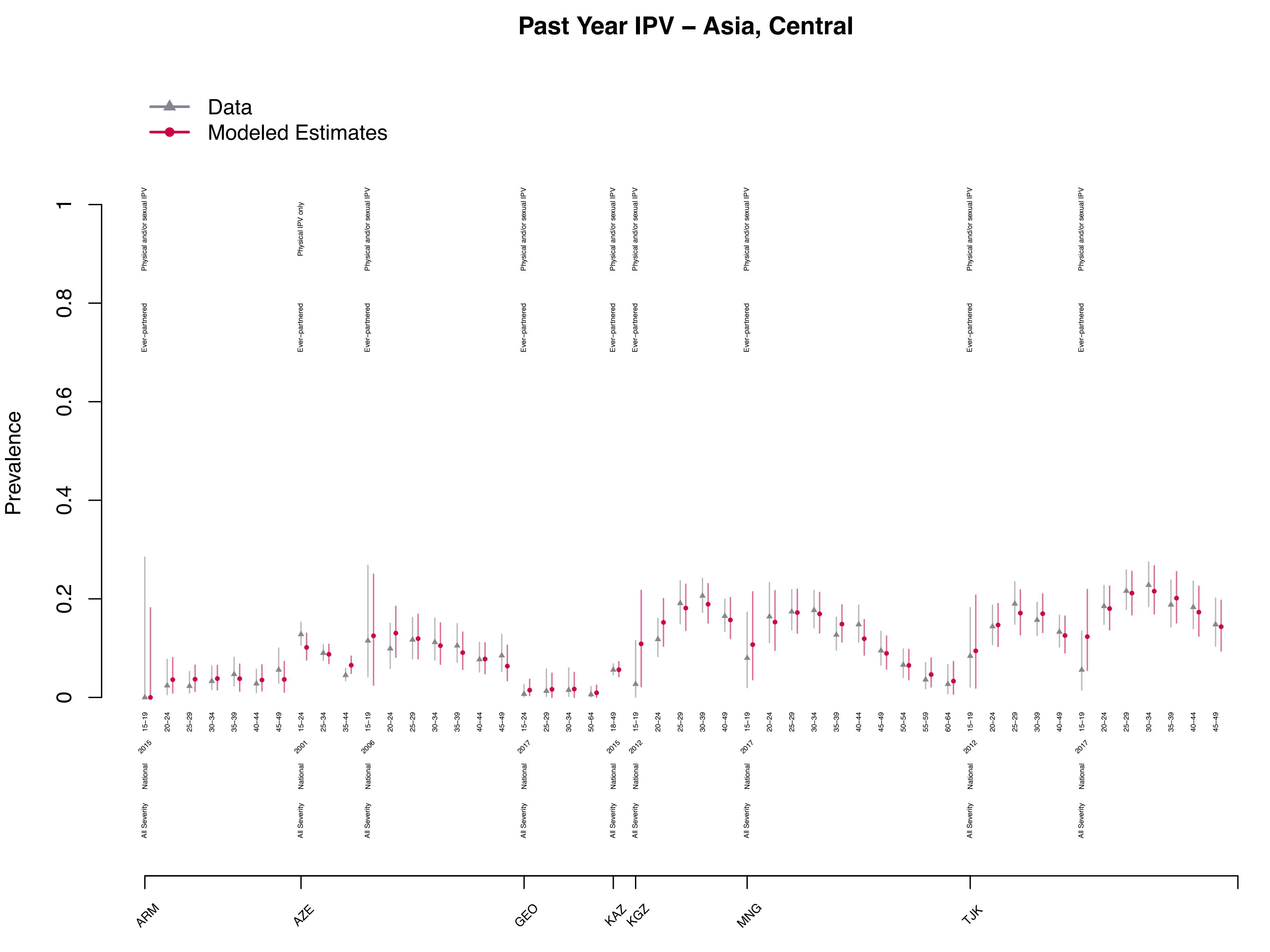
Past Year IPV – Asia, Central
Data
Modeled Estimates
Prevalence
0
0.2
0.4
0.6
0.8
1
Physical and/or sexual IPV
Physical IPV only
Ever−partnered
National
All Severity
ARM
AZE
GEO
KAZ
KGZ
MNG
TJK

### Past Year IPV – Asia, East

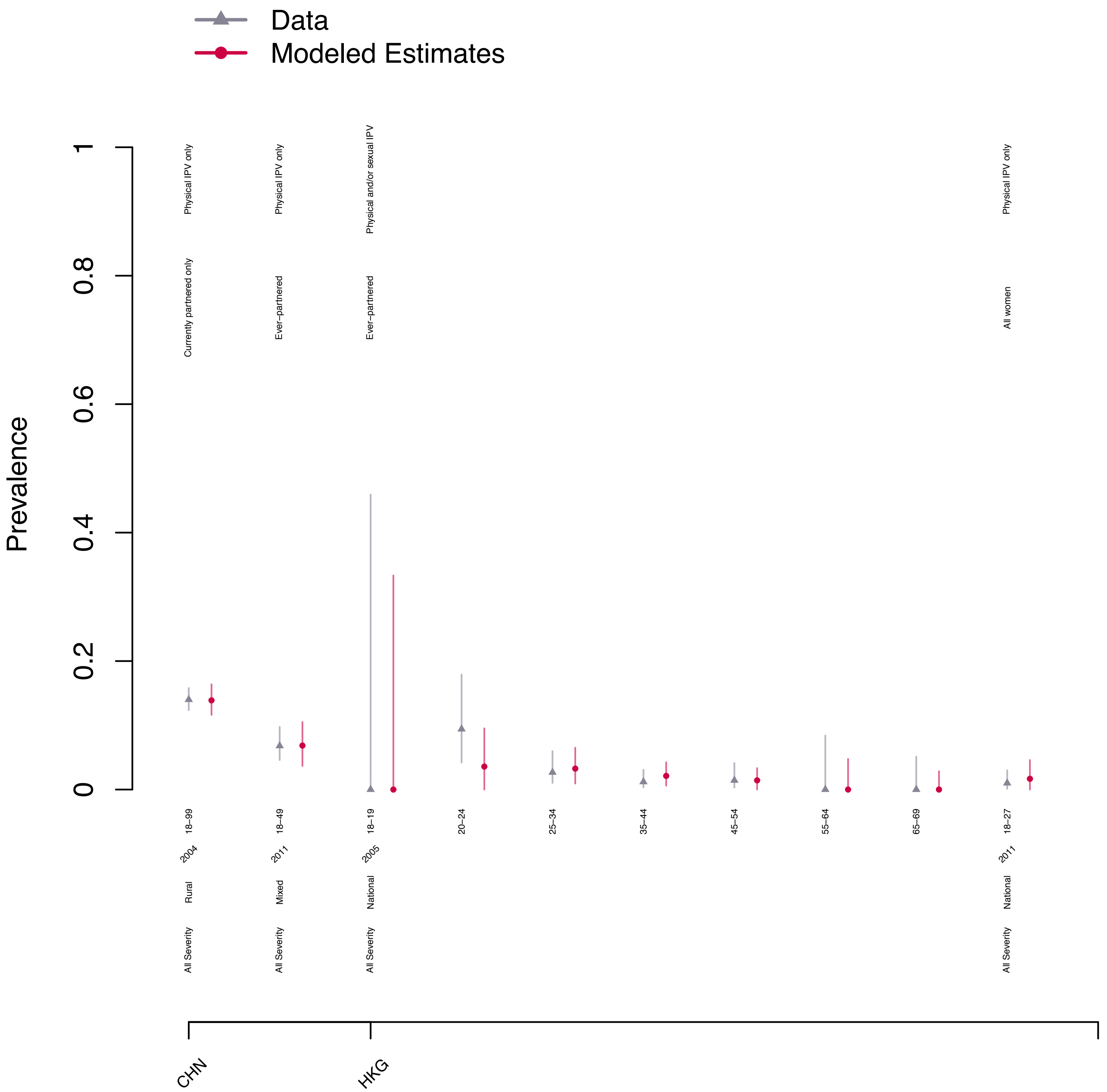

### Past Year IPV – Asia, South

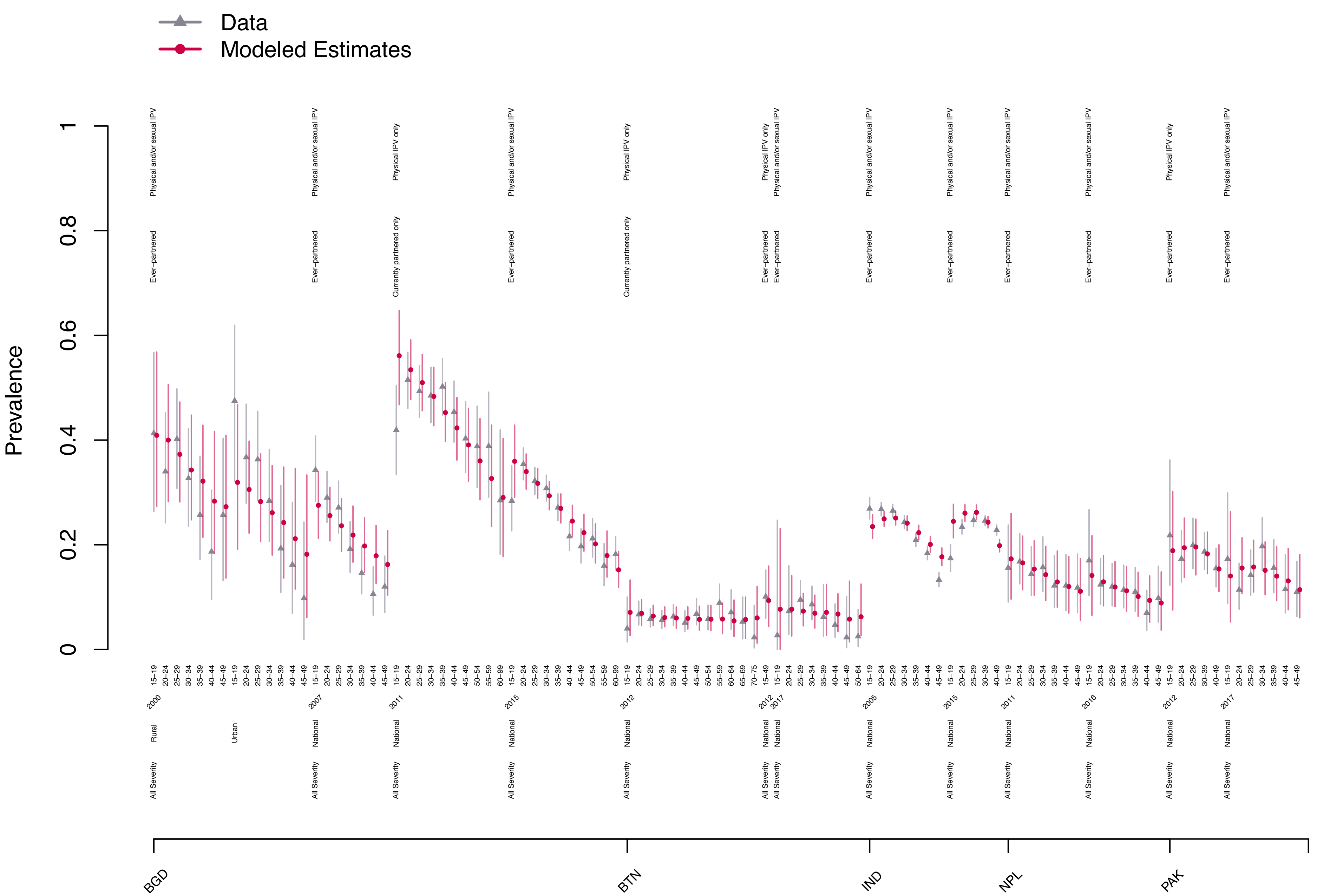

### Past Year IPV – Asia, Southeast

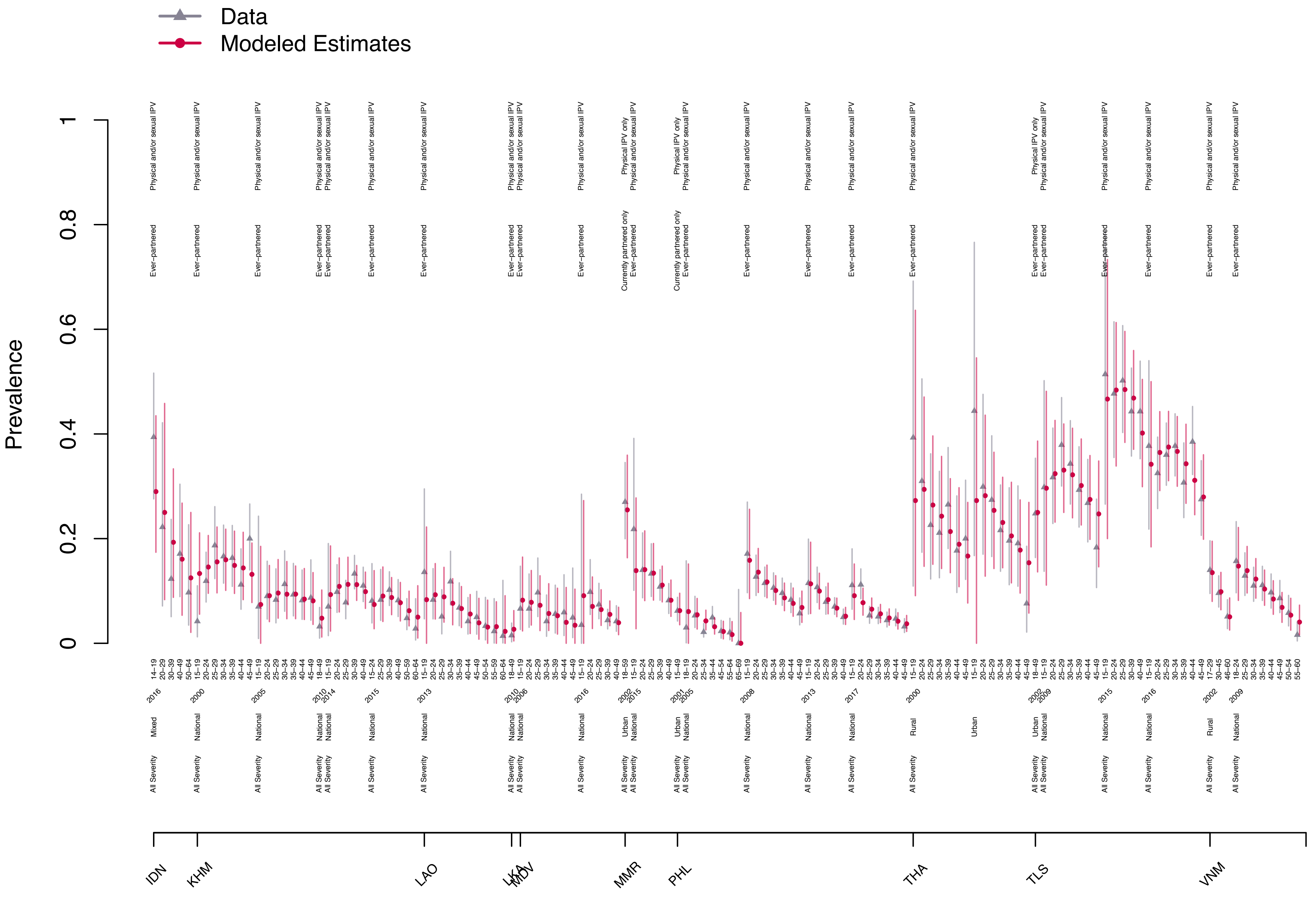

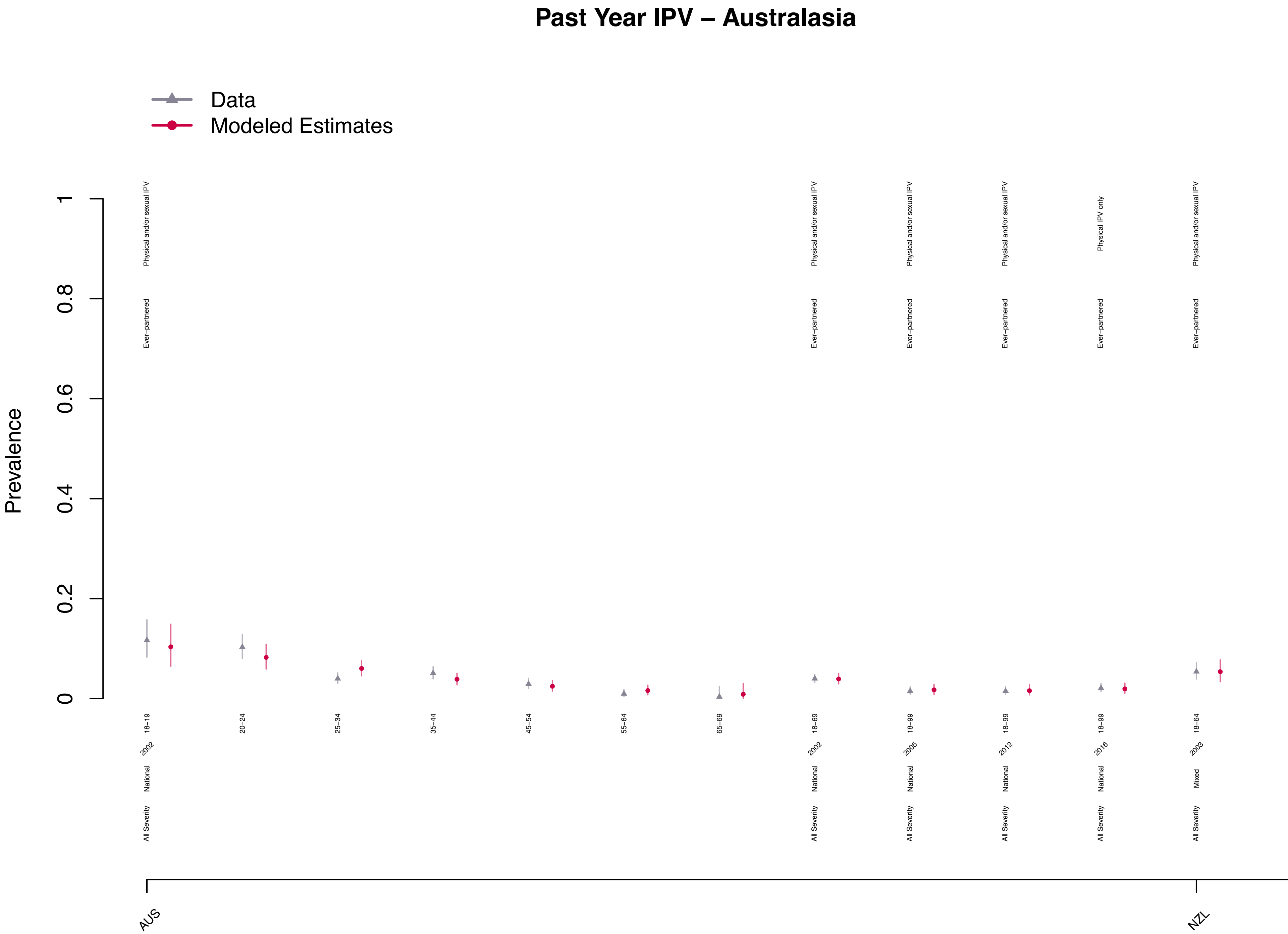

Past Year IPV – Australasia
Data
Modeled Estimates
Prevalence
0
0.2
0.4
0.6
0.8
1
Physical and/or sexual IPV
Physical IPV only
Ever–partnered
18–19
20–24
25–34
35–44
45–54
55–64
65–69
18–69
18–99
18–64
2002
2005
2012
2016
2003
National
Mixed
All Severity
AUS
NZL

#### Past Year IPV – Caribbean

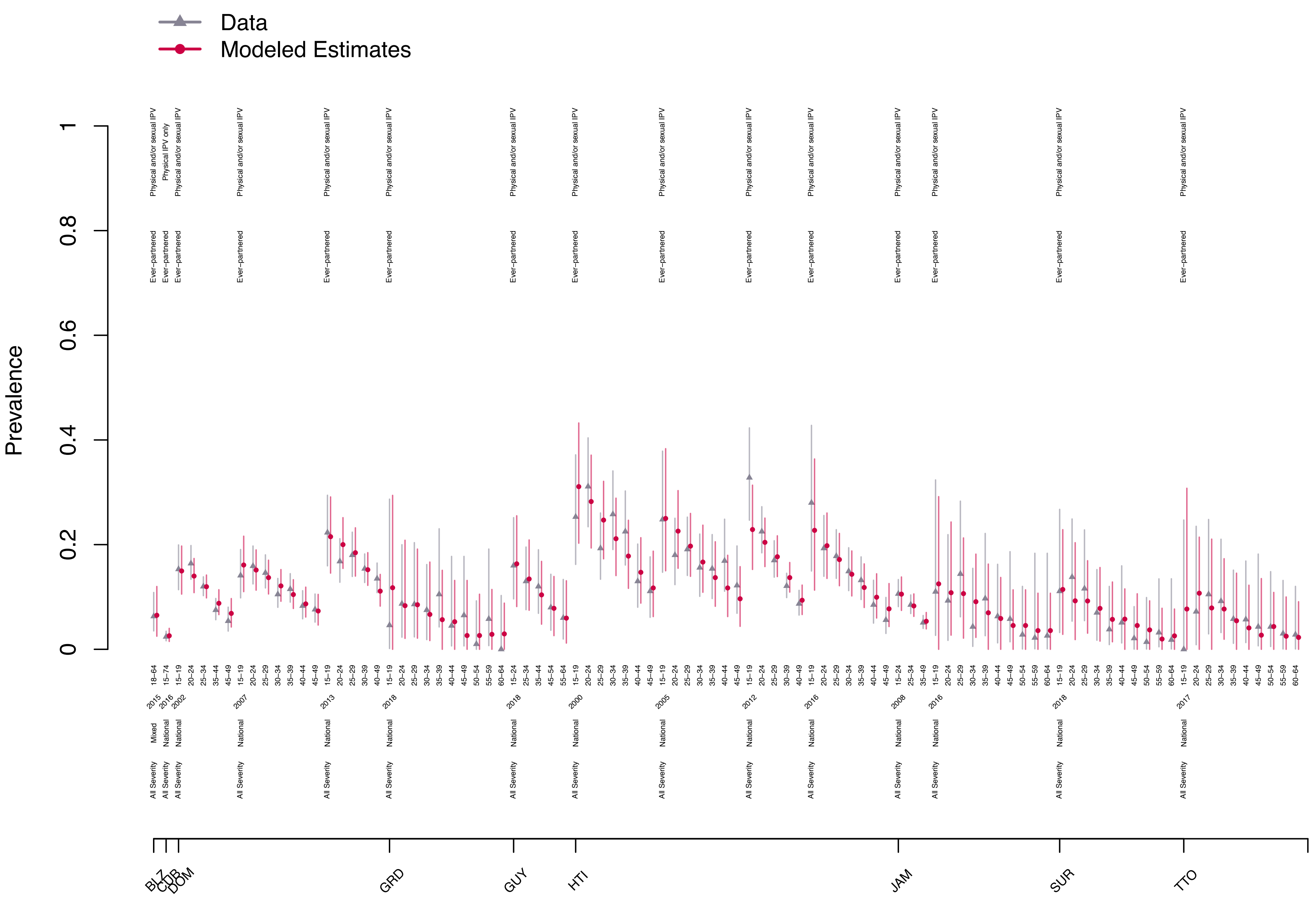

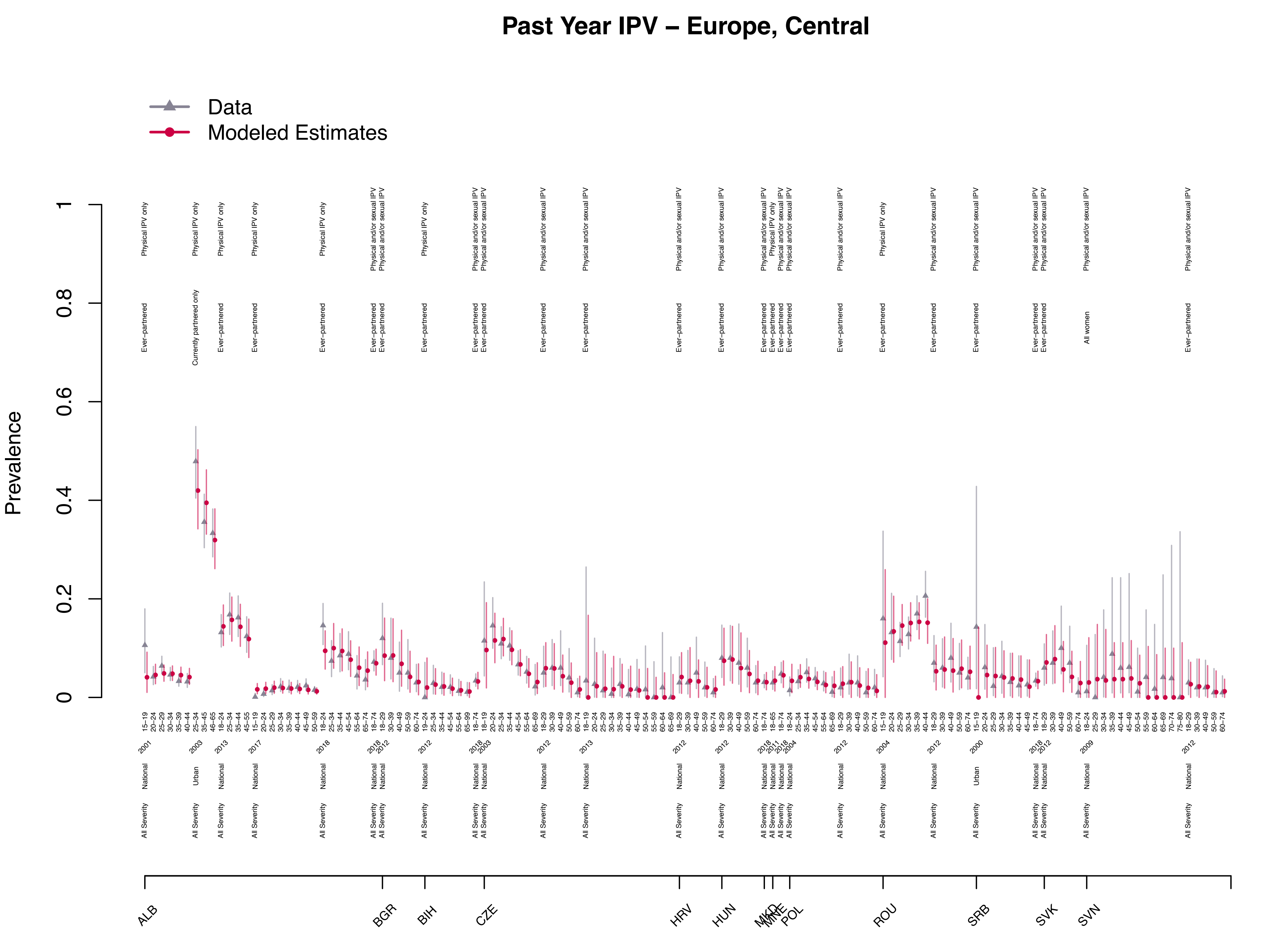
Past Year IPV – Europe, Central
Data
Modeled Estimates
Prevalence
ALB
BGR
BIH
CZE
HRV
HUN
MKD
MNE
POL
ROU
SRB
SVK
SVN

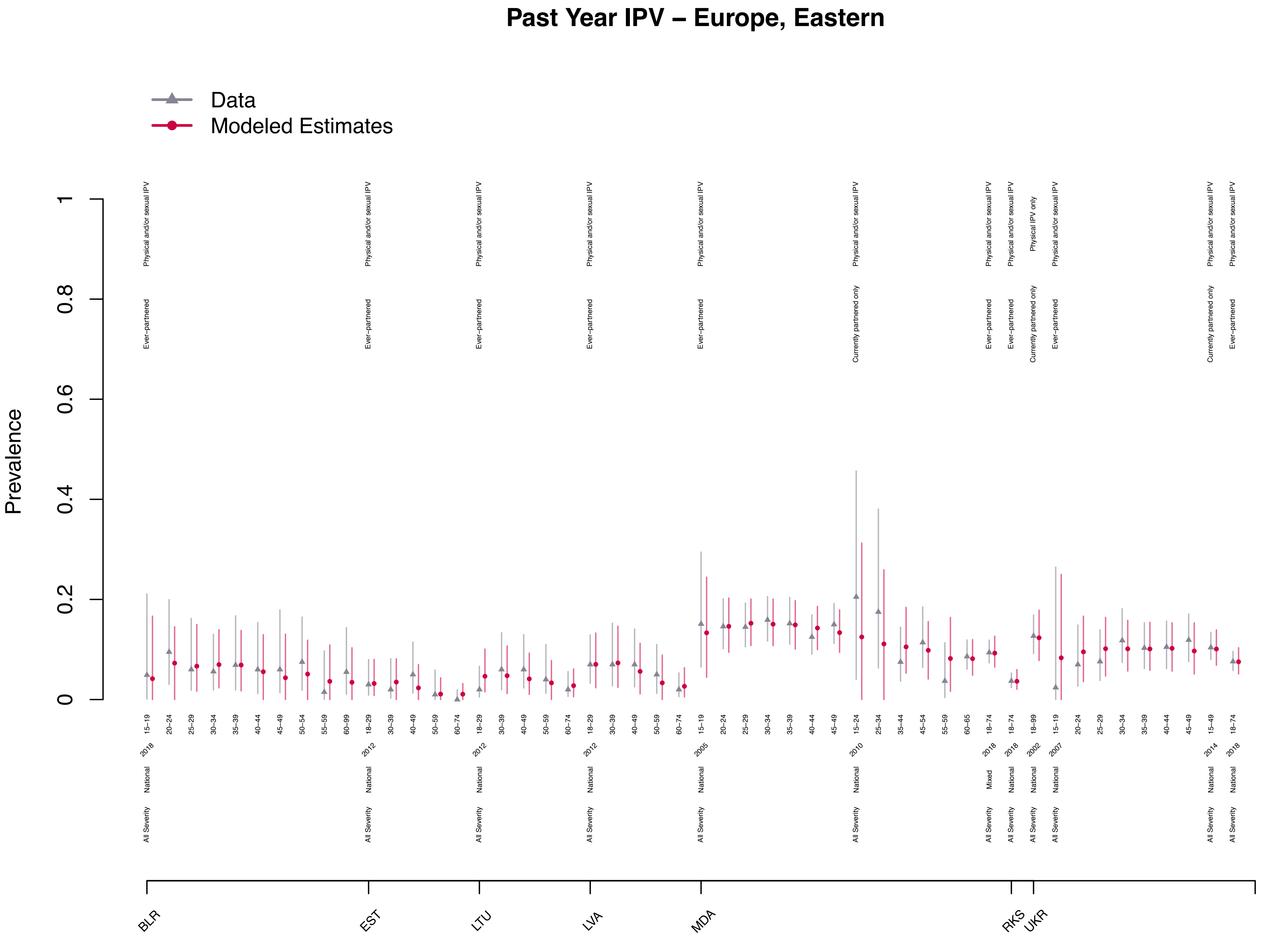

Past Year IPV – Europe, Eastern
Data
Modeled Estimates
Prevalence
0
0.2
0.4
0.6
0.8
1
BLR
EST
LTU
LVA
MDA
RKS
UKR

### Past Year IPV – Europe, Western

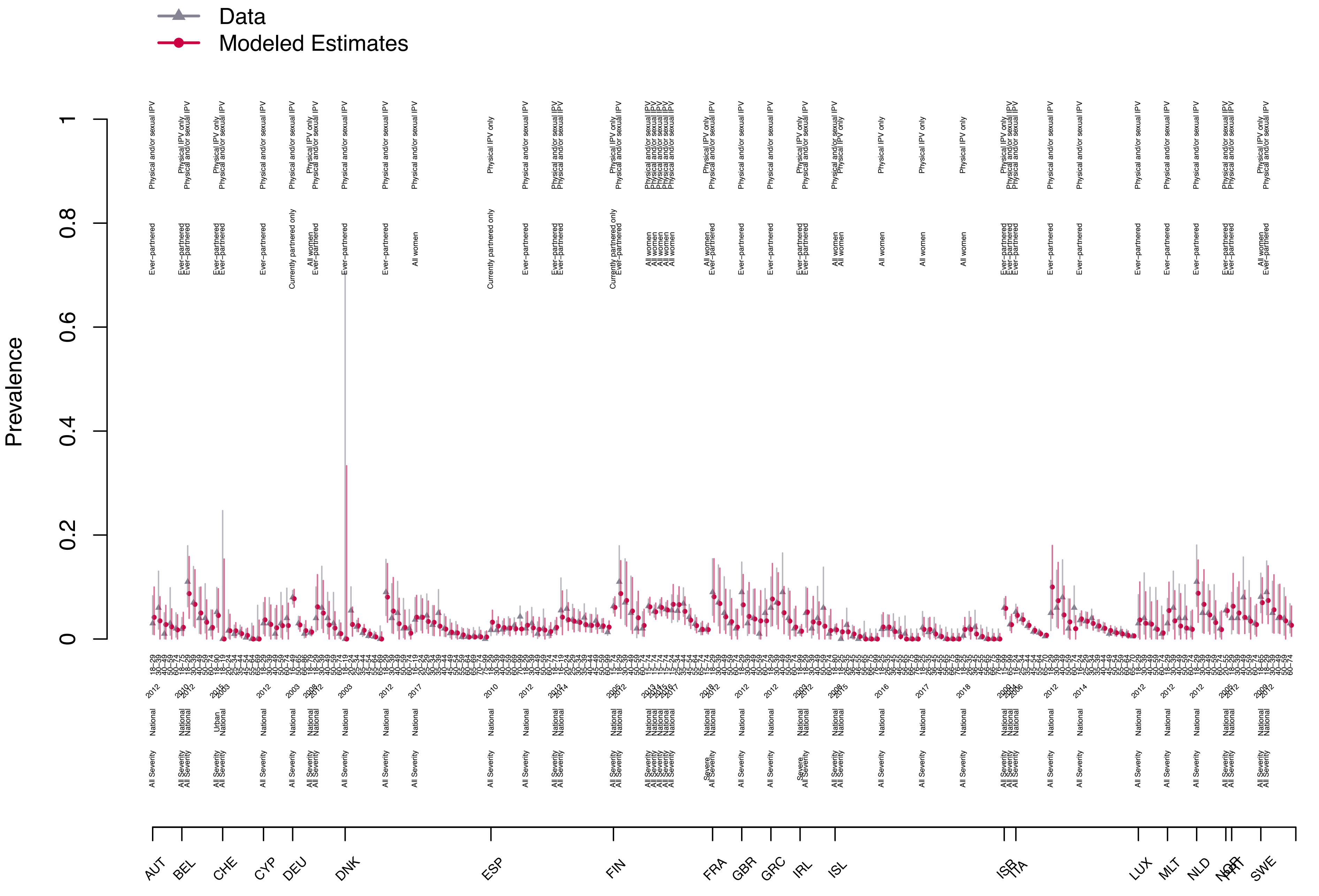

### Past Year IPV – Latin America, Andean

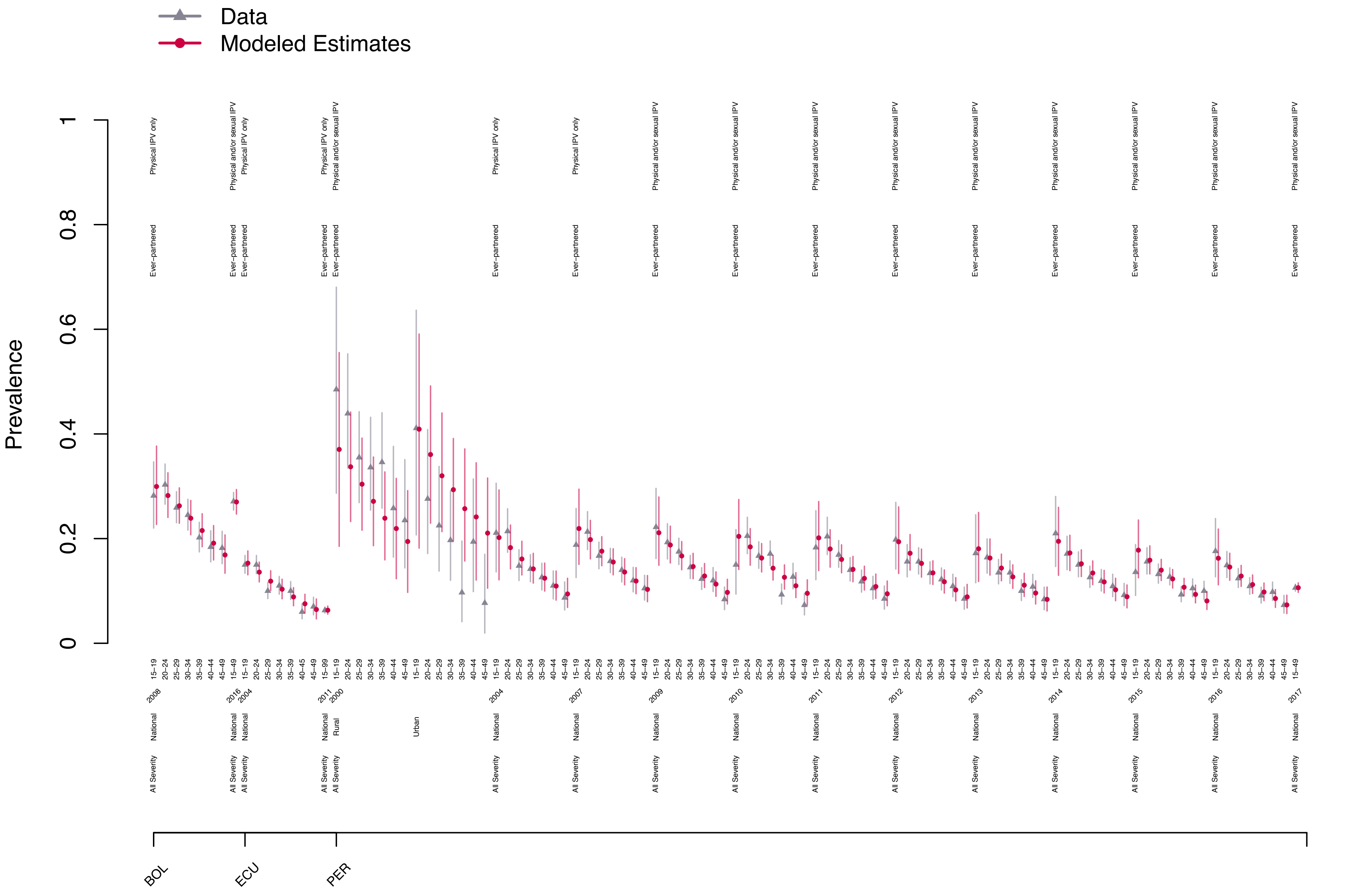

### Past Year IPV – Latin America, Central

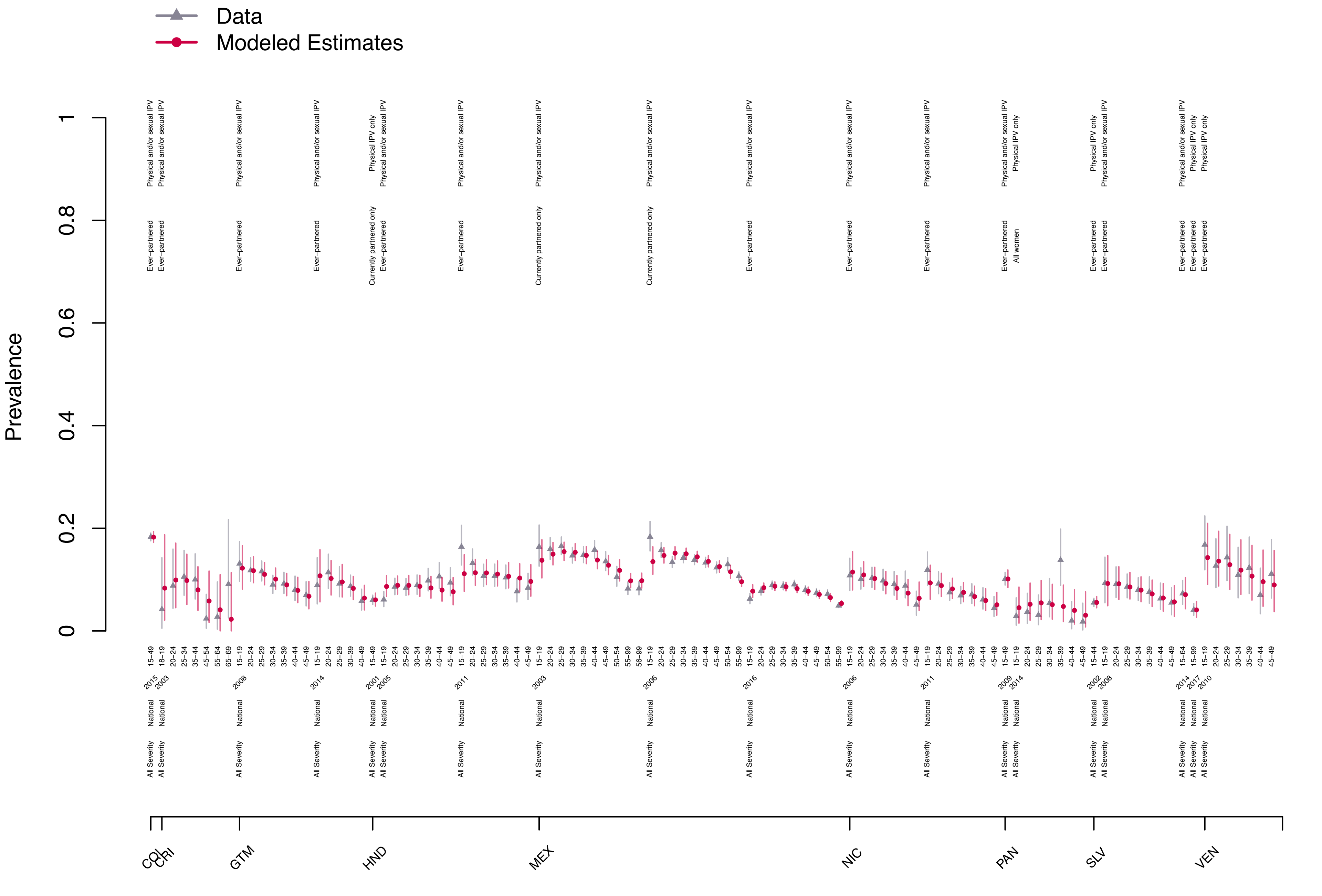

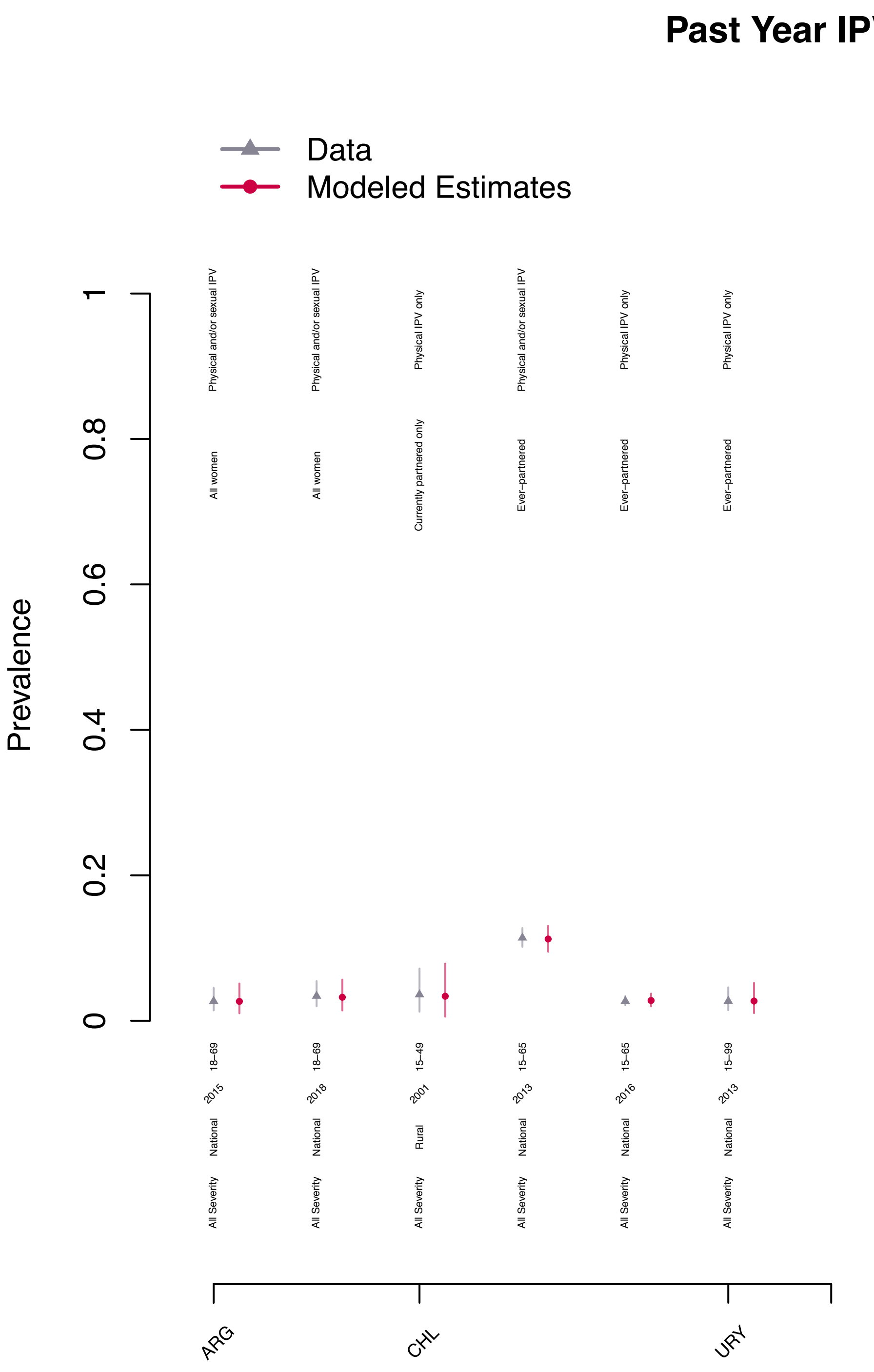

Past Year IPV – Latin America, Southern
Data
Modeled Estimates
Prevalence
0
0.2
0.4
0.6
0.8
1
Physical and/or sexual IPV
All women
18–69
2015
National
All Severity
Physical and/or sexual IPV
All women
18–69
2018
National
All Severity
Physical IPV only
Currently partnered only
15–49
2001
Rural
All Severity
Physical and/or sexual IPV
Ever–partnered
15–65
2013
National
All Severity
Physical IPV only
Ever–partnered
15–65
2016
National
All Severity
Physical IPV only
Ever–partnered
15–99
2013
National
All Severity
ARG
CHL
URY

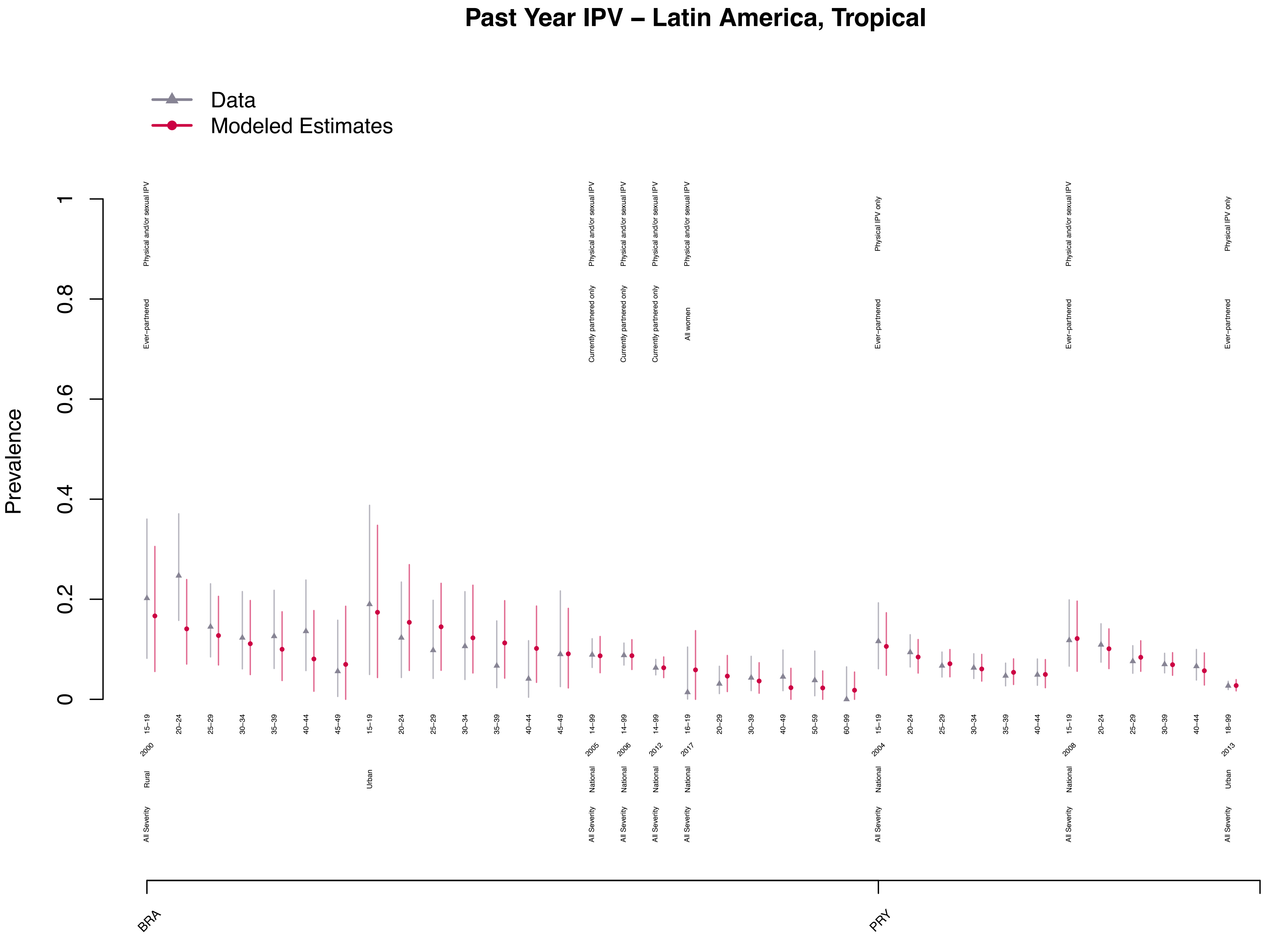
Past Year IPV – Latin America, Tropical
Data
Modeled Estimates
Prevalence
0
0.2
0.4
0.6
0.8
1
Physical and/or sexual IPV
Ever−partnered
Currently partnered only
All women
Physical IPV only
15−19
20−24
25−29
30−34
35−39
40−44
45−49
14−99
16−19
20−29
30−39
40−49
50−59
60−99
18−99
2000
2005
2006
2012
2017
2004
2008
2013
Rural
Urban
National
All Severity
BRA
PRY

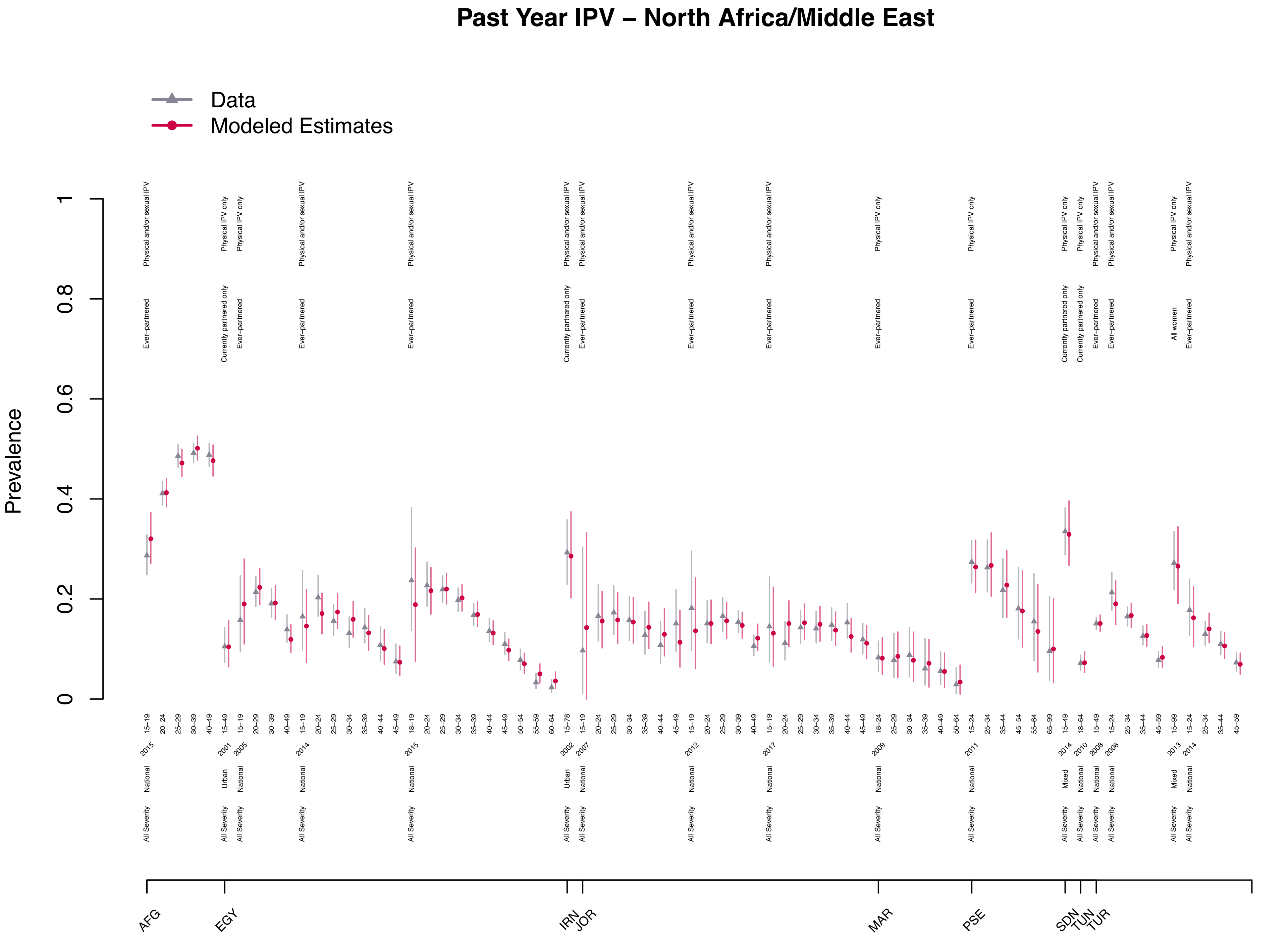

Past Year IPV – North Africa/Middle East
Data
Modeled Estimates
Prevalence
0
0.2
0.4
0.6
0.8
1
AFG
EGY
IRN
JOR
MAR
PSE
SDN
TUN
TUR

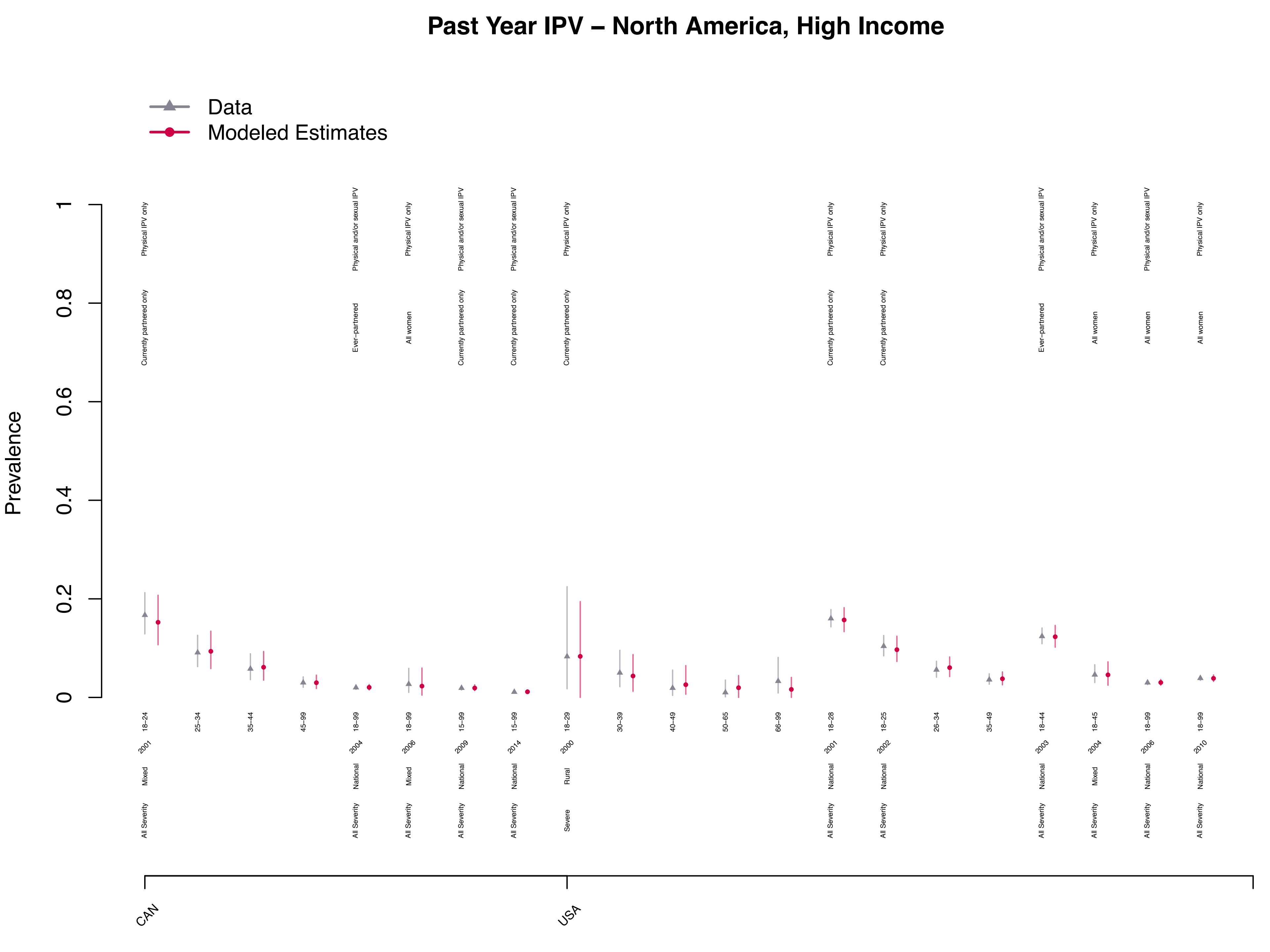

Past Year IPV – North America, High Income
Data
Modeled Estimates
Prevalence
0
0.2
0.4
0.6
0.8
1
Physical IPV only
Physical and/or sexual IPV
Currently partnered only
Ever−partnered
All women
18−24
25−34
35−44
45−99
18−99
15−99
18−29
30−39
40−49
50−65
66−99
18−28
18−25
26−34
35−49
18−44
18−45
2001
2004
2006
2009
2014
2000
2002
2003
2010
Mixed
National
Rural
All Severity
Severe
CAN
USA

### Past Year IPV – Oceania

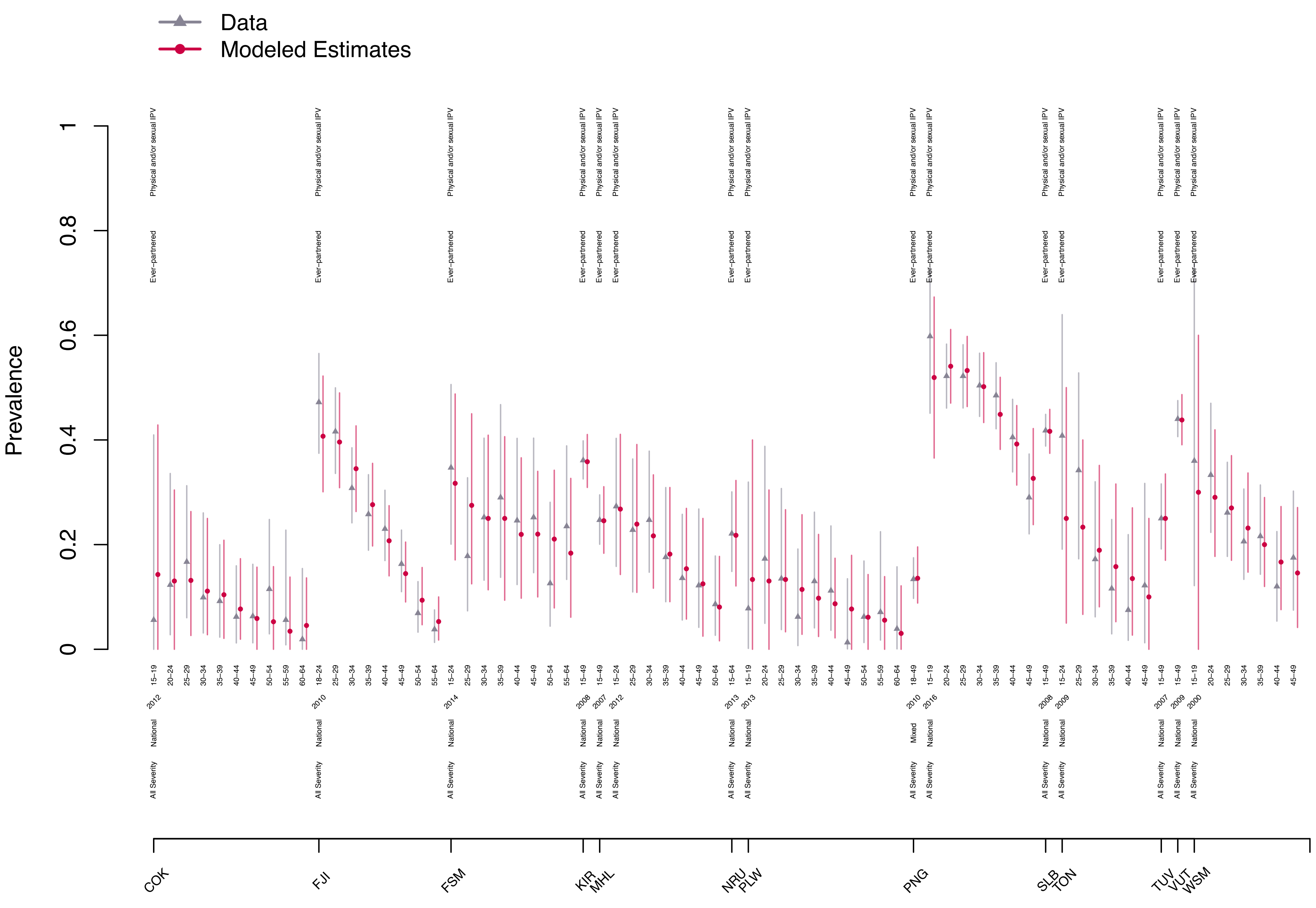

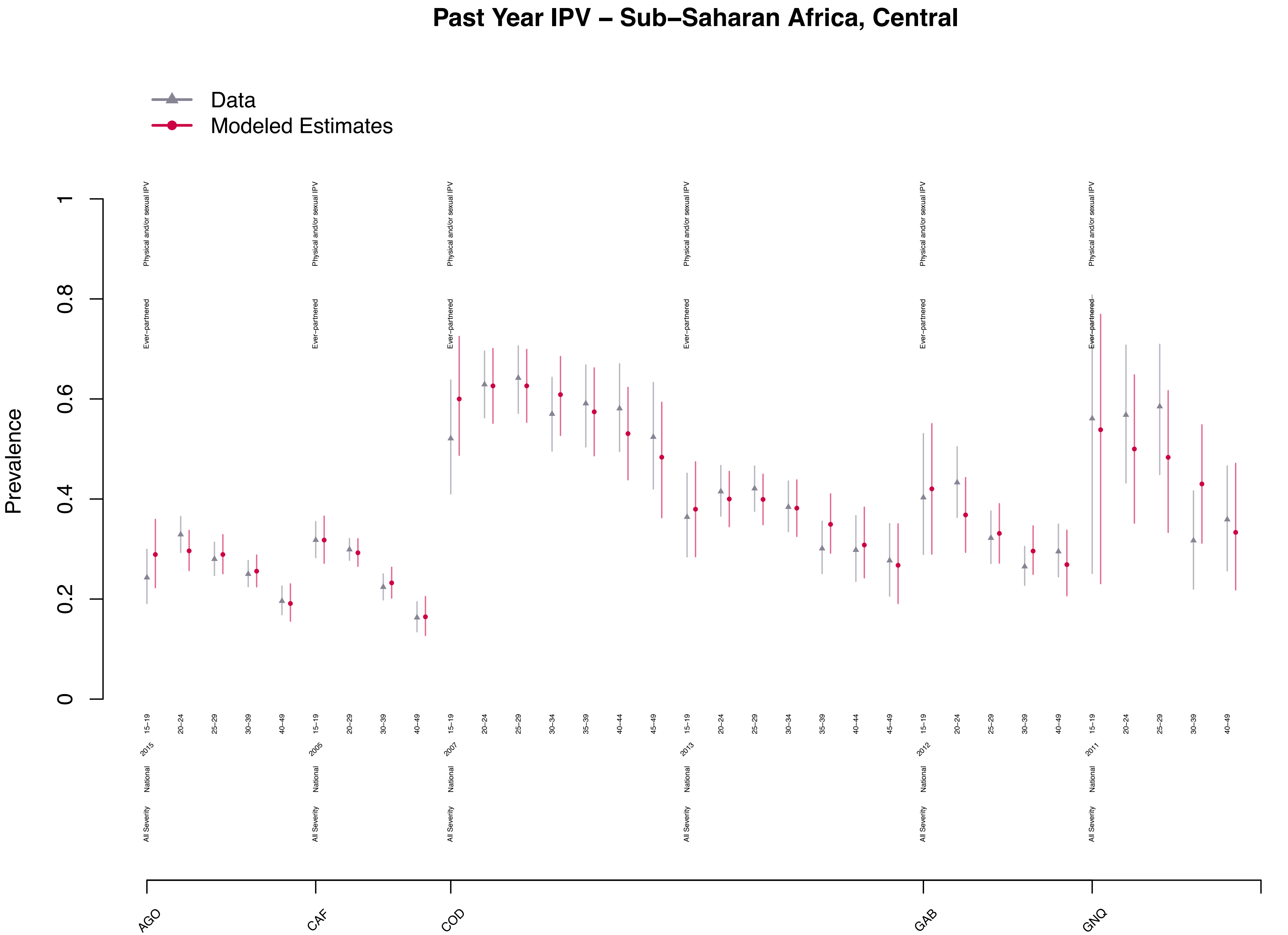

Past Year IPV – Sub–Saharan Africa, Central
Data
Modeled Estimates
Prevalence
0
0.2
0.4
0.6
0.8
1
Physical and/or sexual IPV
Ever–partnered
National
All Severity
AGO
2015
CAF
2005
COD
2007
2013
GAB
2012
GNQ
2011
15–19
20–24
25–29
30–34
35–39
40–44
45–49
30–39
40–49
20–29

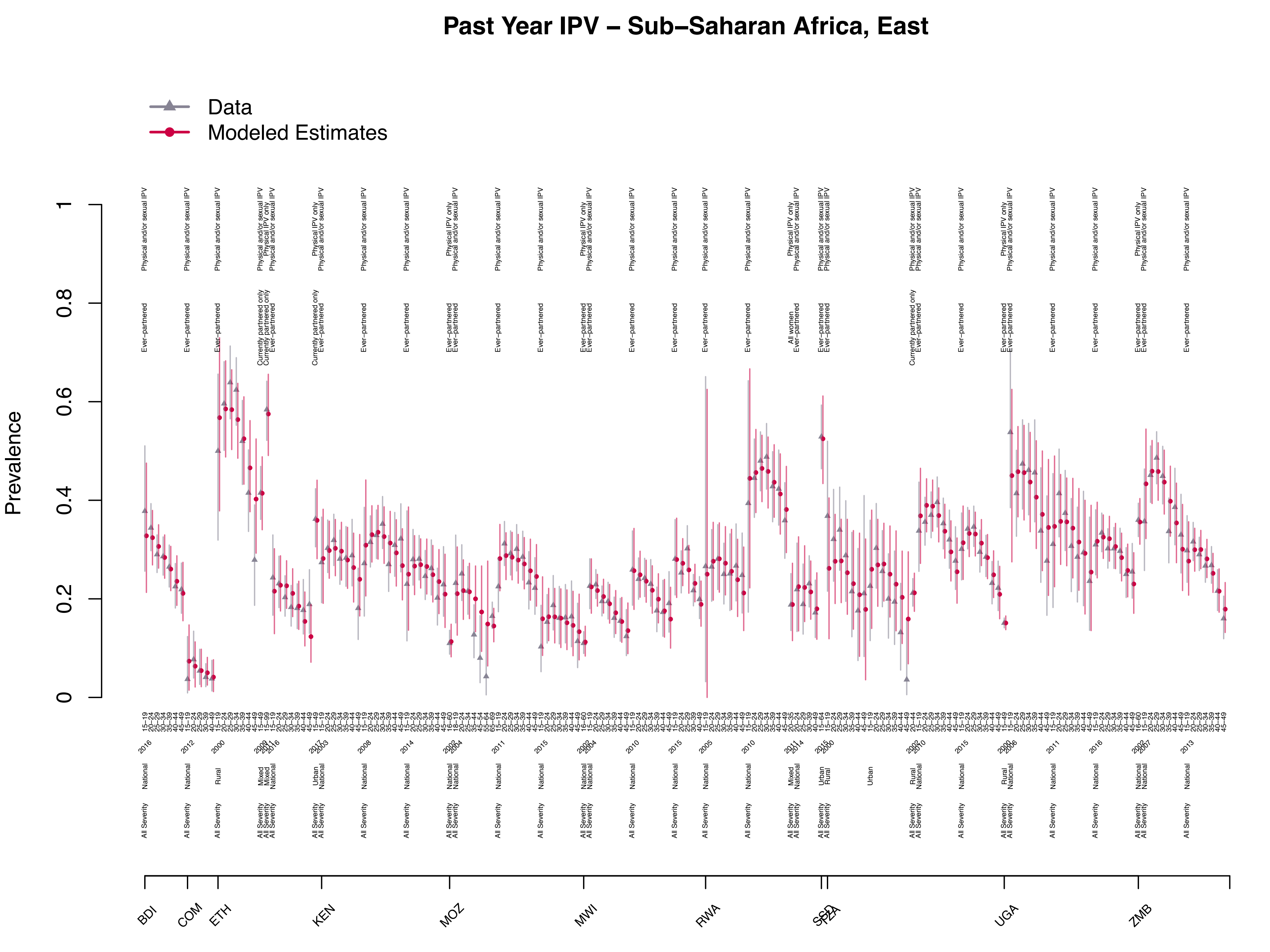
Past Year IPV – Sub–Saharan Africa, East
Data
Modeled Estimates
Prevalence
BDI
COM
ETH
KEN
MOZ
MWI
RWA
UGA
ZMB

#### Past Year IPV – Sub–Saharan Africa, Southern

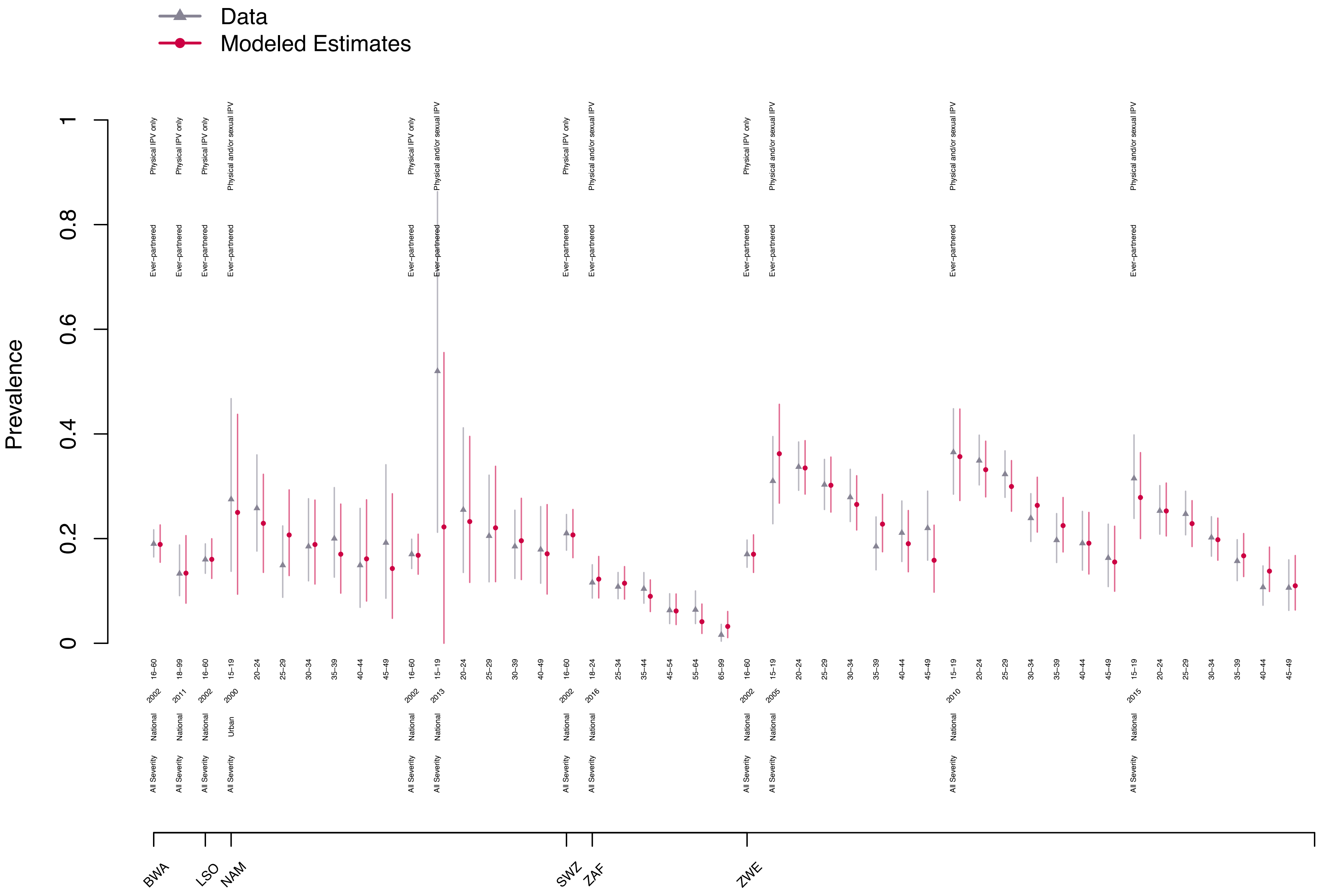

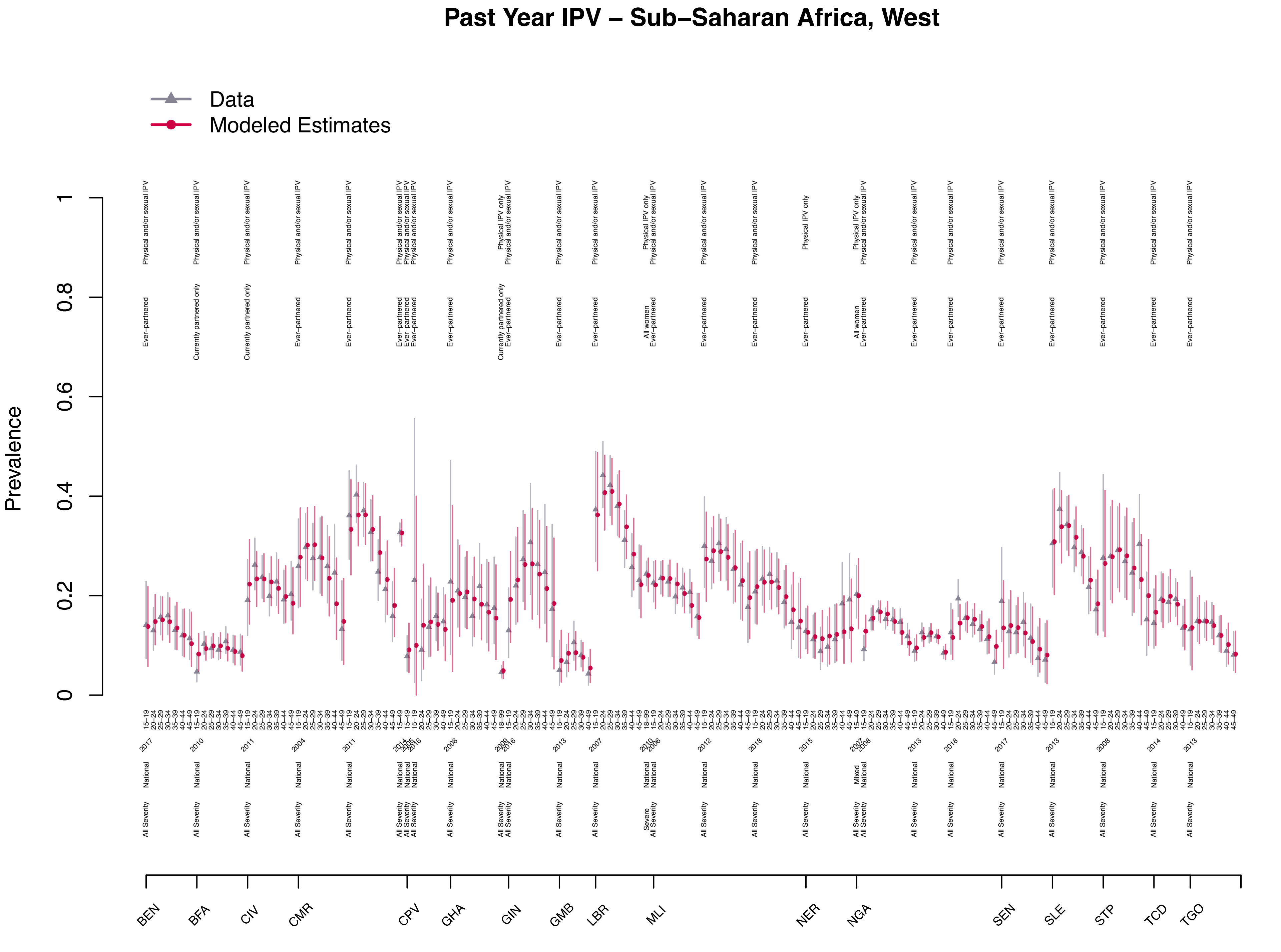

Past Year IPV – Sub–Saharan Africa, West
Data
Modeled Estimates
Prevalence
BEN
BFA
CIV
CMR
CPV
GHA
GIN
GMB
LBR
MLI
NER
NGA
SEN
SLE
STP
TCD
TGO
